# Barriers to engagement by people with active tuberculosis in the care cascade in India: a systematic review of two decades of quantitative research

**DOI:** 10.1101/2023.05.12.23288882

**Authors:** Tulip A. Jhaveri, Disha Jhaveri, Amith Galivanche, Dominic Voehler, Maya Lubeck-Schricker, Mei Chung, Pruthu Thekkur, Vineet Chadha, Ruvandhi Nathavitharana, Ajay M.V. Kumar, Hemant Deepak Shewade, Katherine Powers, Kenneth H. Mayer, Jessica E. Haberer, Paul Bain, Madhukar Pai, Srinath Satyanarayana, Ramnath Subbaraman

**Affiliations:** Division of Geographic Medicine and Infectious Diseases, Tufts Medical Center, Boston, USA; Division of Infectious Diseases, University of Mississippi Medical Center, Jackson, USA; Department of Public Health and Community Medicine and Center for Global Public Health, Tufts University School of Medicine, Boston, USA; Harvard T.H. Chan School of Public Health, Boston, USA; Friedman School of Nutrition Science and Policy, Tufts University, Boston, USA; Center for Operational Research, International Union Against Tuberculosis and Lung Disease (The Union), Paris, France; South-East Asia Office, International Union Against Tuberculosis and Lung Disease (The Union), New Delhi, India; National Tuberculosis Institute, Bengaluru, India; Division of Infectious Diseases, Beth Israel Deaconess Medical Center and Harvard Medical School, Boston, USA; Department of Community Medicine, Yenepoya Medical College, Yenepoya (deemed to be university), Mangalore, India; Division of Health Systems Research, ICMR-National Institute of Epidemiology, Chennai, India; The Fenway Institute, Boston, USA; Center for Global Health, Massachusetts General Hospital and Harvard Medical School, Boston, USA; Countway Library of Medicine, Boston, USA; Department of Epidemiology, Biostatistics and Occupational Health and McGill International TB Centre, McGill University, Montreal, Canada

## Abstract

**Background:** India has the highest burden of tuberculosis (TB), accounting for more than one-quarter of people with active TB and nearly one-third of TB deaths globally. Most people contracting active TB in India do not successfully navigate all stages of the care cascade to receive treatment and achieve TB recurrence-free survival. Understanding reasons for losses across the care cascade is critical to improve outcomes. In this paper, guided by a PECO (population/exposure/comparison/outcome) framework, we describe quantitative findings of a systematic review aimed at identifying factors contributing to unfavorable outcomes experienced by people with TB at each care cascade gap in India.

**Methods and findings:** We defined care cascade gaps as comprising people with confirmed or presumptive TB who did not: start the TB diagnostic workup (Gap 1), complete the diagnostic workup (Gap 2), start treatment (Gap 3), achieve treatment success (Gap 4), or achieve TB recurrence-free survival (Gap 5). Three systematic searches were conducted to identify 147 unique articles published from 2000 to 2021 that evaluated factors associated with unfavorable outcomes for each gap (reported as odds, relative risk, or hazard ratios) and, among people experiencing unfavorable outcomes, reasons reported for these outcomes (reported as proportions). Findings were organized into patient-, family-, society-, or health system-related factors, using a social-ecological framework.

Some factors were common and associated with unfavorable outcomes across multiple care cascade stages. These included male sex, older age (variably defined across studies), a broad array of poverty-related factors, lower symptom severity or duration, undernutrition, alcohol use, smoking, and distrust of (or dissatisfaction with) local government health services. People who had been previously treated for TB were more likely to seek care and engage in the TB diagnostic workup (Gaps 1 and 2) but were also more likely to suffer pretreatment loss to follow-up (Gap 3) and unfavorable outcomes during TB treatment (Gap 4), especially those who had been lost to follow-up during their prior treatment episode.

For individual care cascade gaps, multiple studies highlighted the importance of lack of TB knowledge and structural barriers to care (e.g., transport or financial challenges in reaching clinics) in contributing to lack of care-seeking for TB symptoms (Gap 1, 15 studies or analyses); lack of access to diagnostics (e.g., chest X-ray), non-identification of eligible patients for testing, and failure of providers to communicate concern for TB to patients in contributing to non-completion of the diagnostic workup (Gap 2, 20 studies or analyses); TB stigma, poor recording of patient contact information by providers, and early death due to diagnostic delays in contributing to pretreatment loss to follow-up (Gap 3, 25 studies); and medication adverse effects, TB stigma, and lack of TB knowledge in contributing to unfavorable treatment outcomes (Gap 4, 104 studies). Medication nonadherence contributed to unfavorable treatment outcomes (Gap 4) and post-treatment TB recurrence (Gap 5, 15 studies).

**Conclusions:** This extensive systematic review illuminates common patterns of risk that shape outcomes for people with TB in India, while also highlighting gaps in knowledge, particularly with regard to TB care for children or in the private sector, that can help to guide future research. These findings may help inform targeting of additional support services to people with TB who are at higher risk of poor outcomes and inform development of multi-component interventions to close gaps in India’s TB care cascade.

**Author Summary:** *Why was this study done?:*

- India has the highest burden of tuberculosis (TB), accounting for more than one-quarter of people with active TB and nearly one-third of TB deaths globally.
- Of the nearly 3 million people contracting active TB every year in India, most do not successfully traverse all stages of the care needed to receive TB treatment and achieve an optimal long-term outcome, and serial losses of people with TB across these stages is often referred to as the “cascade of care.”
- Understanding risk factors among people with active or presumptive TB that contribute to losses from care, and why these losses occur, is crucial to inform the development of targeted interventions to prevent them from experiencing unfavorable outcomes.

*What did the researchers do and find?:*

- To understand reasons why people with TB are lost from care in India, we conducted three systematic searches of the medical literature to identify 147 unique and relevant articles published from 2000 to 2021.
- We extracted information from these studies on risk factors for unfavorable outcomes at each care cascade gap, as well as reasons reported by people with TB who experienced unfavorable outcomes and were surveyed by researchers.
- Some barriers to care or characteristics of people with TB contributed to losses at multiple stages of the care cascade, including male sex, older age, poverty-related factors, history of previous TB treatment, lower symptom severity or duration, undernutrition, alcohol use, smoking, and distrust of (or dissatisfaction with) local government health services.
- Other barriers contributed more substantially to individual care cascade gaps: lack of TB knowledge and structural barriers to care (e.g., transport barriers to clinics) contributed to lack of care-seeking for TB symptoms (Gap 1); poor accessibility of diagnostic testing, non-identification of eligible patients for testing, and failure of providers to communicate concern for TB to patients contributed to non-completion of the TB diagnostic workup (Gap 2); early death due to delays in diagnosis, TB stigma, and poor recording of patient contact information by healthcare providers contributed to losses of diagnosed patients before they started treatment (Gap 3); medication adverse effects, TB stigma, and lack of TB knowledge contributed to unfavorable TB treatment outcomes (Gap 4); and medication nonadherence contributed to unfavorable treatment outcomes and post-treatment TB recurrence (Gaps 4 and 5).

*What do these findings mean?:*

- The reasons for losses of people with TB across the care cascade are complex, vary by care cascade gap, and involve a mix of patient- and health system-related barriers.
- Given the complexity of the barriers contributing to unfavorable outcomes in India’s TB care cascade, future implementation interventions should consider involving multiple components that target different challenges faced by patients and the health system.
- In addition, India’s TB program and those in other high incidence settings should target additional services to people with TB who are at higher risk of experiencing poor outcomes.

## Introduction

With an estimated annual incidence of nearly 3 million people contracting active tuberculosis (TB) in 2021, India has the highest TB burden, accounting for more than one-quarter of people with active TB and nearly one-third of TB deaths globally [1]. Prior to the COVID-19 pandemic, India already accounted for the largest number of “missing” people with TB, individuals who have not been reported to TB programs and who therefore may not have received effective care [2]. During the COVID-19 pandemic, case notifications to India’s National TB Elimination Programme (NTEP) dropped by more than 40% in 2020 as compared to 2019, suggesting further decline in people reaching care, though there was partial recovery in notifications in 2021 [1,3]. COVID-19 may have also adversely impacted TB clinical outcomes [4].

Losses of people with a disease condition across sequential stages of care required to achieve a favorable health outcome is referred to as the care cascade (or continuum) [5]. Recent TB care cascade analyses for India and other countries have provided insights into shortcomings in quality of care contributing to unfavorable outcomes for people with TB, especially since TB is almost always curable [6–10]. For example, although the TB community has historically focused on improving outcomes during treatment, in India’s NTEP, there are larger losses from care during diagnostic workup and linkage to treatment [6]. Many people with TB in India also experience TB recurrence in the year after treatment completion [6]. Based on these insights, India’s National Strategic Plan for TB (2017-2025) emphasizes the importance of measuring and reducing losses of people across the care cascade to achieve the 2030 World Health Organization End TB targets, which align with the Sustainable Development Goal [11–13].

While prior TB care cascade analyses quantified gaps in care, few studies have evaluated *who* is falling out of the care cascade and *why* people are lost from care [14]. Improving outcomes requires understanding of patient-, family-, society-, and health system-related factors contributing to losses at each care cascade stage to inform intervention development. In India, many studies have described barriers to care for people with TB; however, most only evaluated a single care cascade gap in one geographic location. By aggregating findings across studies, systematic reviews can identify common risk factors that contribute to unfavorable outcomes across different care cascade gaps and geographic contexts.

India’s NTEP expanded the directly observed therapy short-course (DOTS) strategy in a phased manner starting from 1997 to achieve coverage of most of the population with public TB services by the early 2000s, making it the world’s largest TB program [15]. In this paper, we describe quantitative findings of a systematic review assessing the past two decades of literature aimed at identifying factors contributing to unfavorable outcomes across the TB care cascade in India. The NTEP has also mandated reporting of people undergoing TB treatment in the private sector since 2012 and provides support through public-private initiatives [16,17]. As such, while factors shaping TB outcomes undoubtedly vary across India’s large and diverse population, identifying common challenges may inform local and national implementation strategies, since care delivery across the country is informed by uniform guidelines (for the public sector [11,18]) and recommended standards (for the private sector [19]). By identifying reasons for poor outcomes [14], this review aims to inform intervention development across care cascade stages to improve the lives of people with TB and advance disease elimination in the world’s largest TB epidemic.

## Methods

### TB care cascade framework

This review expands upon an earlier systematic review that estimated losses across India’s TB care cascade, from a starting point of people contracting active TB in the population (i.e., annual incidence) to the end outcome of one-year post-treatment recurrence-free survival [6] (Table 1). The framework guiding this review is also informed by subsequent guidelines [5] and other national-level TB care cascade analyses from South Africa, Zambia, and Madagascar [7–9].

**Table 1.** Definitions for each TB care cascade gap and specific dimensions investigated in this systematic review. [5]

| Care cascade gap | General definition | Specific populations and outcomes assessed |
| --- | --- | --- |
| Gap 1 | People with confirmed TB (i.e., active TB not previously diagnosed before the community survey) or presumptive TB in the community who do not reach care at facilities and start the TB diagnostic workup | We included studies evaluating people with confirmed or presumptive TB in the community who had or had not sought any care for their symptoms. Studies of people who reached care without initiation of a TB diagnostic workup were not included. |
| Gap 2 | People with presumptive TB who start but do not complete the appropriate diagnostic workup | Given changes in diagnostic algorithms over time, we report findings by non-completion of each diagnostic modality, including non-pursual of workup by patients despite referral, non-completion of sputum microscopy evaluation, non-completion of chest X-ray, and non-completion of NAAT (e.g., Xpert MTB/RIF, Truenat). |
| Gap 3 | People who are diagnosed with TB but do not start or get registered in treatment | We disaggregate findings by the following sub-populations: drug-susceptible TB patients, drug-resistant TB patients, and pediatric TB patients. |
| Gap 4 | People who start or get registered in treatment but do not achieve treatment success | We disaggregate findings by the following sub-populations: new TB patients, previously treated TB patients, rifampin- or multidrug-resistant TB patients, multiple populations of TB patients, people with HIV being treated for active TB, and pediatric TB patients. Outcomes of interest included standard NTEP- and WHO-defined unfavorable outcomes of death, treatment failure, loss to follow-up, or non-evaluation (i.e., not reported, transferred out, or treatment regimen modified), alone or in combination. Medication nonadherence was included as an unfavorable outcome, as it is a mediator of other unfavorable treatment outcomes [20]. |
| Gap 5 | People who achieve TB treatment success, but experience disease recurrence or death evaluated for the 12 months after treatment completion, at minimum | We disaggregated studies evaluating post-treatment TB recurrence (with or without death) and studies evaluating post-treatment mortality alone. Studies sometimes evaluated recurrence or death only in the post-treatment phase or included post-treatment outcomes as part of a composite outcome, along with unfavorable on-treatment outcomes. Findings using both approaches were included in Gap 5. |

NAAT, nucleic acid amplification testing; TB, tuberculosis; WHO, World Health Organization.

As this comprehensive review spans 5 care cascade gaps, the protocol was submitted to PROSPERO in October 2019 and registered in April 2020 under three IDs: CRD42020159337 (for Gap 1), CRD42020159361 (for Gaps 2, 3 and 4), and CRD42020159355 (for Gap 5). A detailed protocol for each care cascade gap is provided in the S1—S5 Appendices. In the rest of this section, we discuss aspects of our methodological approach that were common across all gaps.

### PECO framework

Using a PECO (population, exposures, comparisons, outcomes) framework [21], the *population* comprised people with active or presumptive TB in India, regardless of whether they sought or received care in the public or private sector. We extracted data on a wide array of *exposures* (e.g., sociodemographic, clinical, psychosocial, structural, and health system factors) that may have contributed to poor outcomes at each care cascade gap. The *comparison* group depended on the specific exposure. For example, for male sex, female sex was the comparison (reference) group, but for age, the age range used as a reference group varied by study. We report quantitative findings in the current review; qualitative findings will be reported in a future manuscript.

Exposures represented findings from two study designs or analytical approaches. The first set of findings—referred to hereafter as “factors associated with unfavorable outcomes”—derived from cohort, case-control, and cross-sectional studies comparing exposures between people who completed a specific care cascade stage versus those who did not. These studies usually reported findings for exposures as unadjusted or adjusted odds ratios, relative risk (prevalence) ratios, or hazard ratios, in relation to the outcome (i.e., care cascade gap).

The second set of findings—referred to hereafter as “reasons reported by people with TB for unfavorable outcomes”—were extracted from studies that conducted structured interviews with people who experienced unfavorable outcomes for a given care cascade gap. Interviews sought to understand why people with TB symptoms in the community had not sought care (for Gap 1), why people diagnosed with TB had not started treatment (for Gap 3), or why people had not achieved treatment success (for Gap 4). Studies described the number and proportion of people reporting specific reasons for unfavorable outcomes, among all people who were interviewed (as the denominator). Similar findings were available for patients who had not completed the diagnostic workup (Gap 2); however, for this gap, studies also reported reasons based on health system data (e.g., failure of sputum transport to testing sites). Because interview-based studies only collected data from people who experienced unfavorable outcomes, there was no comparison or reference group.

*Outcomes* of interest were guided by the broad definition of each care cascade gap, within which nuanced definitions were needed that were relevant to study designs and subpopulations (Table 1). For Gap 1, we included studies evaluating people with confirmed or presumptive TB in the community who had or had not sought care when the survey was conducted. We defined “presumptive TB” as comprising individuals with symptoms suggestive of TB (e.g., cough >2 weeks, fevers, night sweats), who serve as a proxy for understanding the behavior of people with undiagnosed active TB in the population. A limitation is that our approach to Gap 1 only provides insights into factors shaping the behavior of people with TB symptoms *before* they seek care. Many barriers contributing to losses in Gap 1 occur *after* people seek care, due to failure of healthcare providers to test for TB or delays in starting treatment [22–26]. While recognizing limitations of our approach to Gap 1, we decided not to include studies of delays in TB care because these are comprised of biased samples of people who ultimately started treatment (i.e., did not get lost from care), and these studies have been covered in a previous systematic review [26]. Studies assessing the failure of providers to test people with presumptive TB were excluded because they are framed from the perspective of providers (rather than people with TB), as they have involved assessments of provider knowledge (using surveys [22]) or behavior (using standardized patients [24,25]).

For Gap 2, a challenge is that TB diagnostic algorithms evolved over 2 decades, especially with growing use of nucleic acid amplification tests (NAAT; e.g., Xpert MTB/RIF, Truenat). Despite this evolution, most algorithms culminated in completion of a specific diagnostic modality for each type of TB. For example, sputum microscopy had to be completed to diagnose smear-positive TB; chest X-ray had to be completed to diagnose smear-negative (or NAAT-negative) TB; and NAAT or mycobacterial culture had to be completed to diagnose rifampin-resistant (RR) or multidrug-resistant TB (MDR TB). As such, we disaggregated Gap 2 findings by non-completion of specific diagnostic modalities, regardless of the algorithm used.

For Gaps 3 and 4, it was possible to disaggregate findings by patient subpopulations (Table 1). Gap 4 outcomes comprised NTEP [27] and World Health Organization (WHO) [28] case definitions for the following treatment outcomes: loss to follow-up, treatment failed, died, not evaluated (e.g., not reported, transferred out, treatment modified [often considered a form of treatment failure]), or a composite of more than one of these. In addition, when studies reported loss to follow-up of less than two months (i.e., the programmatic definition of loss to follow-up) or medication nonadherence using other measures, we also analyzed these as unfavorable outcomes representing medication nonadherence, given nonadherence is a mediator of other unfavorable outcomes [20,29].

For Gap 5, we disaggregated studies that evaluated post-treatment recurrence (with or without mortality) and those that evaluated post-treatment mortality alone. Studies evaluated post-treatment outcomes starting either from treatment initiation or treatment completion—with the former approach reporting post-treatment outcomes as part of a composite outcome that included unfavorable on-treatment outcomes. We included findings from both approaches in Gap 5, as post-treatment follow-up in these studies was longer than on-treatment follow-up.

### Search strategy and study selection

Given heterogeneity in studies included in various care cascade gaps, we conducted three sets of searches. The first set identified articles for Gap 1; the second set identified articles for Gaps 2, 3, and 4 (as these 3 gaps similarly looked at loss to follow-up from care); and the third set identified articles for Gap 5 (Table A in each of the S1—S5 Appendices).

Because Gap 1 focused on care-seeking by people with confirmed or presumptive TB, these searches involved the following umbrella terms and related variants: “tuberculosis,” “tuberculosis symptoms,” “healthcare-seeking behavior,” and “India.” As Gaps 2, 3, and 4 involve loss to follow-up of patients, searches for these gaps comprised the following umbrella terms and related variants: “tuberculosis,” “loss to follow-up,” and “India.” As Gap 5 focused on post-treatment TB recurrence, searches comprised the following umbrella terms and related variants: “tuberculosis,” “recurrence,” and “India.”

We searched PubMed, Embase, and Web of Science for articles starting from 2000, when India’s modern TB program—known previously as the Revised National TB Control Programme and since 2017 as the NTEP—achieved national coverage [15]. Given the large number of articles and extensive data extraction process, trained medical librarians conducted an initial search and two refresher searches for each gap spanning January 1, 2000 to October 1, 2015; October 2, 2015 to October 1, 2019; and October 2, 2019 to May 17, 2021. Notably, the January 2000 to October 2015 searches had been conducted as part of a previous study quantifying gaps in India’s TB care cascade [6]. We screened all articles identified in searches for the prior systematic review for inclusion in the current review; however, whereas we extracted findings on *outcomes* for the prior review, we extracted findings on *exposures* for the current review. In addition to the database searches, studies were also identified by looking at references lists of the included primary studies and outreach to experts.

Facilitated by Covidence software (Veritas Health Innovation, Australia), identification of articles for eligibility at the title and abstract and full text stages was performed independently by at least two reviewers (among TJ, DJ, AG, DV, MLS, and KP). Disagreements were resolved by discussion among reviewers or, if necessary, through consultation of a third reviewer (RS). PRISMA flow diagrams describing the study identification process for each gap, including refresher searches, are provided in the S1—S5 Appendices.

Inclusion and exclusion criteria varied by care cascade gap and are described in detail in the S1—S5 Appendices; however, some general criteria applied across all gaps. Included studies had to have either compared people with TB who did or did not complete a specific care cascade stage (i.e., factors associated with unfavorable outcomes) or have surveyed of people who experienced unfavorable outcomes to identify reasons for those outcomes (i.e., reasons reported by people for unfavorable outcomes). Studies that only described the proportion of people experiencing unfavorable outcomes, without analyzing reasons for these outcomes, were excluded from the review. Finally, because the current paper’s focus is on reasons for unfavorable outcomes in “real world” public or private sector care, we excluded findings from clinical trials, which often use external resources to retain patients.

### Quality assessment

In our prior systematic review, we developed quality criteria relevant to studies focused on evaluating poor outcomes in India’s TB care cascade [6], because of variability in the study designs included in that review and the lack of standardized guidelines for assessing the quality of these observational studies. Although we extracted exposures rather than outcomes in the current review, we used similar quality criteria because they are appropriate for describing overall methodological rigor and risk of bias (Table B in each of the S1-S5 Appendices). Studies were not excluded based on quality unless they involved convenience sampling, as we felt such sampling approaches had substantial risk of being non-representative.

Quality criteria vary by gap (S1-S5 Appendices), but we discuss nuances of these criteria here. For Gap 1, studies involved cross-sectional population-based screening to identify individuals with presumptive TB (i.e., TB symptoms) or confirmed active TB. Bias may be introduced if only a small sample of the population gets screened, so we included the proportion of the population screened as a quality criterion (S1 Appendix, Table B). Among individuals identified as having presumptive or confirmed TB, bias might be introduced if only a small proportion were surveyed about care-seeking behavior (i.e., high non-response rate). As such, the proportion of individuals surveyed was included as a quality criterion.

Gaps 2 and 3 evaluate stages before treatment initiation, when determining outcomes can be challenging. For example, for Gap 2, patients who must obtain a chest X-ray for a diagnosis of sputum smear-negative or NAAT-negative TB may receive this imaging at a different clinic from where diagnostic workup was initiated. Similarly, for Gap 3, patients may start treatment at a clinic that is different from where the diagnosis was made. For this reason, Gap 2 and 3 outcomes are more accurately determined through patient tracking by a dedicated research team, rather than by relying on data collection from medical records alone; therefore, studies using the former approach were rated as being higher in quality. Studies assessing outcomes within a shorter time frame (e.g., within 1 to 2 months) for Gaps 2 and 3 were also rated as being higher in quality than studies assessing outcomes later, because tracking patients becomes harder with time due to patient mobility and recall bias (S2 Appendix, Table B and S3 Appendix, Table B).

Gap 4 and 5 studies followed standard cohort designs, so for both gaps we rated studies with prospective data collection as higher in quality than studies with retrospective data collection. For Gap 5, identification of people with post-treatment TB recurrence may be influenced by whether surveillance was active (e.g., prospective screening) or passive (e.g., waiting for people to return to care if they develop symptoms), with the former approach rated as higher in quality. In addition, identification of TB recurrence depends on the diagnostic modality used. We rated studies involving repeated post-treatment screening with microbiological tests (e.g., mycobacterial culture or NAAT) as being higher in quality than studies relying on clinical diagnosis alone for identifying TB recurrence (S5 Appendix, Table B).

### Data extraction

Two reviewers (among TJ, DJ, AG, DV, and MLS) independently extracted data from each study into a structured Excel spreadsheet. Disagreements were resolved by discussion or, if necessary, by consulting a third reviewer (RS). We extracted data on each study’s design, location, setting (i.e., urban versus rural), sample size, and other descriptors for specific care cascade gaps (e.g., type of TB for Gaps 3 and 4). For studies reporting factors associated with unfavorable outcomes, we extracted unadjusted and adjusted effect estimates (odds ratios, relative risk ratios, and hazard ratios) and 95% confidence intervals (CIs) for all exposures. Some studies only compared the proportion of individuals with a given exposure who did or did not experience a given outcome, using Chi-squared or Fisher’s exact tests to assess statistical significance. When possible, we used these raw data to estimate unadjusted odds ratios with 95%CIs to facilitate comparison with findings of other studies. For studies reporting reasons for unfavorable outcomes, we extracted proportions (numerator and denominator) for every reported reason with 95%CIs. When not reported, we estimated 95%CIs, assuming an infinite population size. Given the large volume of findings, the senior author (RS) verified all extracted findings against the original papers after creation of the final tables and Forest plots.

Some studies presented the association of exposures with favorable (rather than unfavorable) outcomes. This was particularly common for Gap 1, with many studies reporting the odds of having sought care for TB symptoms. For consistency, we “flipped” these effect estimates to present the association with experiencing unfavorable (rather than favorable) outcomes.

For some variables, we also changed the reference group for consistency of reporting across studies. For example, because most studies compared men to the reference group of women, we “flipped” effect estimates for studies presenting men as the reference group. This allows readers more easily identify common trends (or discordant findings) across studies. Similarly, although the age category used as a reference group varied across studies, we consistently presented the youngest age category as the reference group.

We extracted unadjusted effect estimates to understand relevant baseline factors that may predict unfavorable outcomes. We also extracted adjusted effect estimates to elucidate potential causal influence. We reported all unadjusted and adjusted effect estimates, regardless of statistical significance, organized by study in Table D in each of the S1—S5 Appendices. In the main manuscript and Forest plots, we restricted ourselves to presenting statistically significant adjusted effect estimates from multivariable analyses, as these may represent more meaningful associations from higher-quality analyses.

### Analytical framework for organizing and visualizing findings

Losses across care cascade gaps occur due to a diverse array of barriers. Informed by the multi-level social-ecological framework [30] and a previous review of non-initiation of treatment among people with HIV [31], we created a framework that organizes findings into “patient-, family, or society-related factors” and “health system-related factors,” with subcategories to facilitate reporting of findings (Table 2). While these categories may be simplistic—because many barriers represent an interaction between patient circumstances and health system limitations—this organizing framework may help identify where to focus interventions.

**Table 2.**
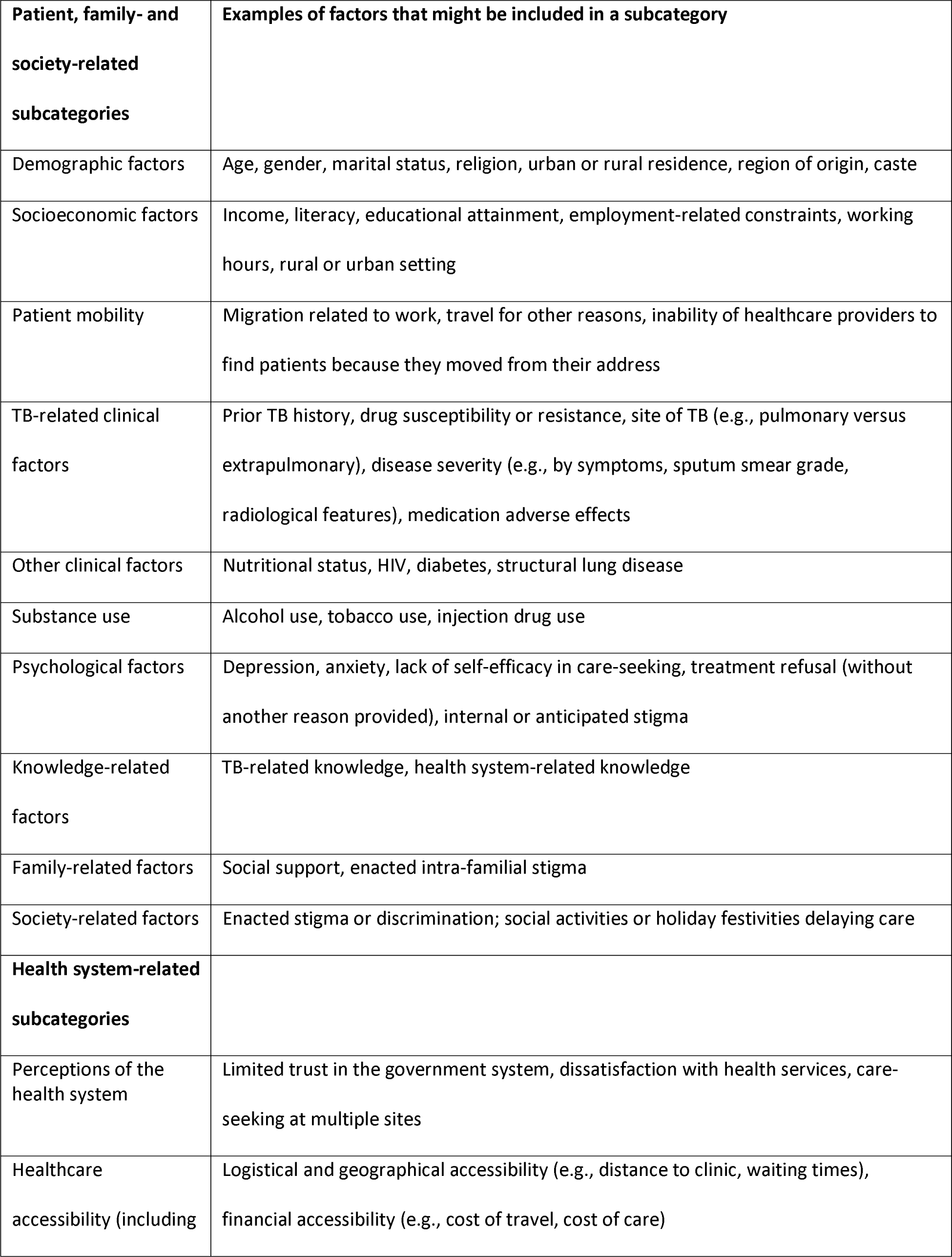
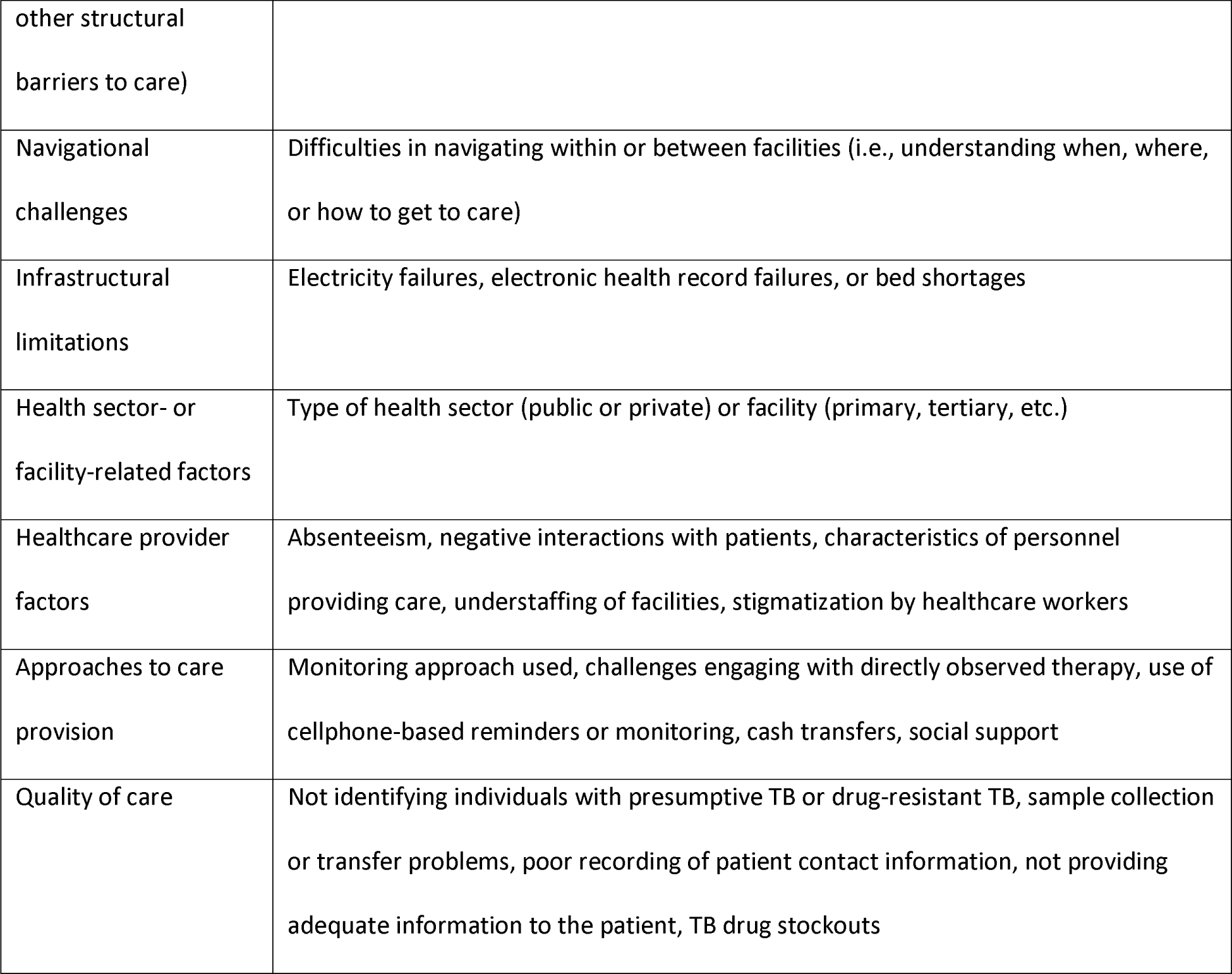
Framework for organizing factors contributing to unfavorable care cascade outcomes.

To help readers visualize common trends or discordant finding across studies, we generated Forest plots of statistically significant adjusted effect estimates from multivariable analyses using Stata version 16.1 (College Station, USA). Meta-analyses were not performed, because we extracted findings from diverse exposures; therefore, we felt pooling the data was inappropriate. We also generated Forest plots for reasons reported by people with TB for unfavorable outcomes, for which all reported findings were proportions.

During the visualization process, we had to manage the heterogeneity of reported effect estimates (i.e., odds ratios, relative risk ratios, hazard ratios) and outcomes for some care cascade gaps (e.g., death, loss to follow-up, treatment failure, and medication nonadherence for Gap 4). Making separate Forest plots for each type of effect estimate and outcome would not only have been prohibitive, but it also would have made it harder for readers to visualize common trends. To guide intervention development, understanding the exact type of unfavorable outcome associated with an exposure may be less important than understanding how common this association is across different settings. For these reasons, we combined different types of effect estimates and outcomes in the same Forest plots for different gaps, but provide this information in footnotes to each figure.

Although terminology varied by study and over time, we occasionally modified language used in the original studies (as long as it did not change the meaning) when reporting findings in Forest Plots to ensure use of person-centered language, as suggested by the Stop TB Partnership [32]. We use “patients” to describe people with TB only after their engagement with health services. Older terminology that referred both to a treatment approach and a patient subpopulation (e.g., category 2 treatment) has also been changed to reflect contemporary terminology (e.g., previously treated patient).

## Results

### Gap 1—Barriers contributing to people with confirmed or presumptive TB in the community not having sought care for TB symptoms

#### Characteristics and quality of the included studies

Across three searches spanning January 1, 2000, to May 17, 2021, we screened titles and abstracts of 4,977 unique reports and identified 307 reports for full text review, of which 12 met inclusion criteria (Fig A in S1 Appendix). 1 additional study meeting inclusion criteria was identified by reviewing references of other articles [33]. Three datasets—1 of which was linked to 1 of the 12 studies meeting inclusion criteria—were identified by outreach to experts. At our request, the authors conducted secondary analyses of those datasets to evaluate factors associated with not having sought care for TB symptoms [34–36].

As such, 15 articles or analyses were included in the Gap 1 review. One study reported data from two geographic locations [37], so we present characteristics across 16 studies or locations. Of these, 13 presented findings on factors associated with not having sought care and 9 presented findings on reasons for not seeking care reported by people with presumptive TB (Table C in S1 Appendix). Studies were conducted in 7 of India’s 28 states and 8 union territories, including Uttar Pradesh and Madhya Pradesh, two of India’s high-population and low-income states by gross domestic product (GDP) per capita, and 3 studies were conducted in multiple states. 5 studies were conducted in rural areas, 8 in urban areas, and 3 in both (Table C in S1 Appendix).

All studies involved a high-quality sampling strategy (i.e., random or comprehensive sampling). 1 study was low quality with regard to sample size [38]. 10 studies did not report the proportion of people screened for presumptive TB during population-based data collection (low quality). 1 study did not report the proportion of individuals with presumptive TB who completed an interview [32] (low quality), while, in another study, only 68% of individuals with presumptive TB were interviewed [39] (medium quality) (Table C in S1 Appendix).

#### Factors associated with not having sought care for TB symptoms

We identified commonalities in findings across studies analyzing factors associated with not having sought care for TB symptoms (Fig 1). With regard to gender, 1 study found men had higher adjusted odds of not seeking care than women in 2 different Indian states [37]. Although another study found women had higher adjusted odds of not seeking care, this association was evident only after controlling for smoking and alcohol use, which were reported exclusively by men and considerably increased the odds of not seeking care [59]. Lower socioeconomic status—whether measured using household income, educational attainment, or occupation (e.g., daily wage laborer)—was associated with higher adjusted odds of not seeking care. People with TB symptoms living in locations with lower income or weaker health systems (e.g., rural locations and North or East India) had higher adjusted odds of not seeking care.

**Fig 1.**
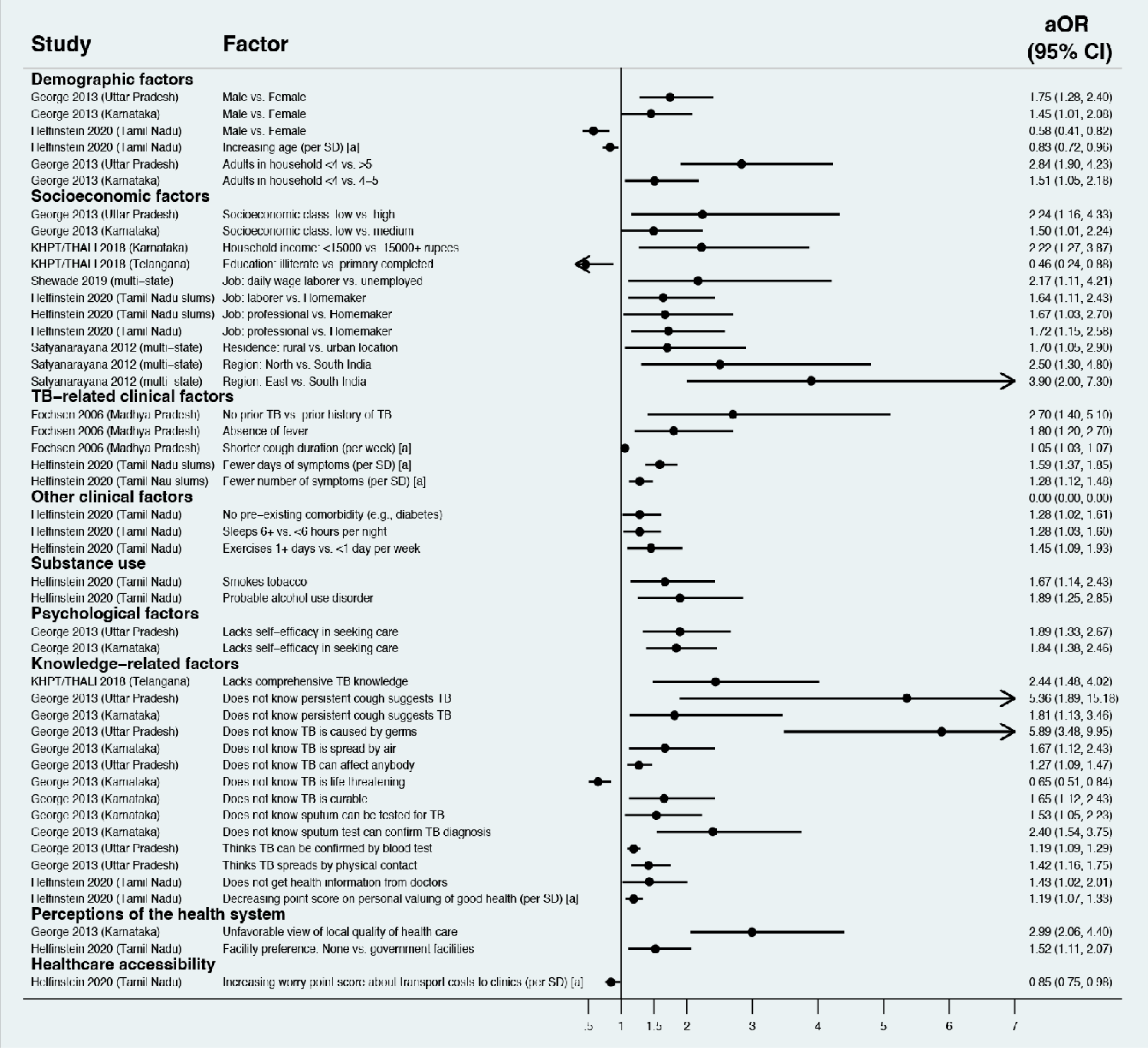
Factors associated with individuals in the population not having sought care for tuberculosis-associated symptoms (Gap 1). All studies used multivariable logistic regression with findings reported as adjusted odds ratios. Only statistically significant findings are presented. Estimates greater than 1 represent increased adjusted odds of not seeking care; estimates less than 1 represent decreased adjusted odds of not seeking care. Variables labelled [a] represent continuous variables included in regression analyses; effect estimates should be interpreted per one level change in the unit given in parentheses. CI, confidence interval; KHPT/THALI, Karnataka Health Promotion Trust/Tuberculosis Health Action Learning Initiative; aOR, adjusted odds ratio; SD, standard deviation; TB, tuberculosis.

TB clinical and knowledge-related factors also influenced care-seeking behavior. Not having had TB previously and lower symptom severity—e.g., absence of fever, fewer symptoms, and shorter duration—were associated with higher adjusted odds of not seeking care. Lacking knowledge of symptoms, mode of transmission, and the curative nature of therapy were associated with higher adjusted odds of not seeking care. Not getting information from doctors, having an unfavorable view of the quality of local health facilities, and lack of preference for government facilities were also associated with higher odds of not seeking care.

In general, studies that only performed unadjusted analyses had similar findings, including associations between low educational attainment, unemployment, lower symptom severity, and lower TB knowledge with higher risk of not seeking care (Table D in S1 Appendix) [33,60,61].

#### Reasons reported by individuals for not seeking care

We also identified commonalities in studies that interviewed people with TB symptoms to identify reasons they had not pursued care (Fig 2). Socioeconomic constraints—including lack of money and work limitations—were frequently reported, with a few studies finding about half of individuals experienced these barriers [37,61,62]. Lack of symptom severity or resolving symptoms, were also major reasons for not seeking care, with a few studies finding that half or more of individuals experienced these barriers [33,38,62,63]. Healthcare-related barriers were common, with one-fifth or more of patients in several studies reporting that dissatisfaction with local health facilities [37], perceived indifference of local healthcare providers [61], or structural barriers to accessing clinics (e.g., distance [38,61]) prevented them from seeking care.

**Fig 2.**
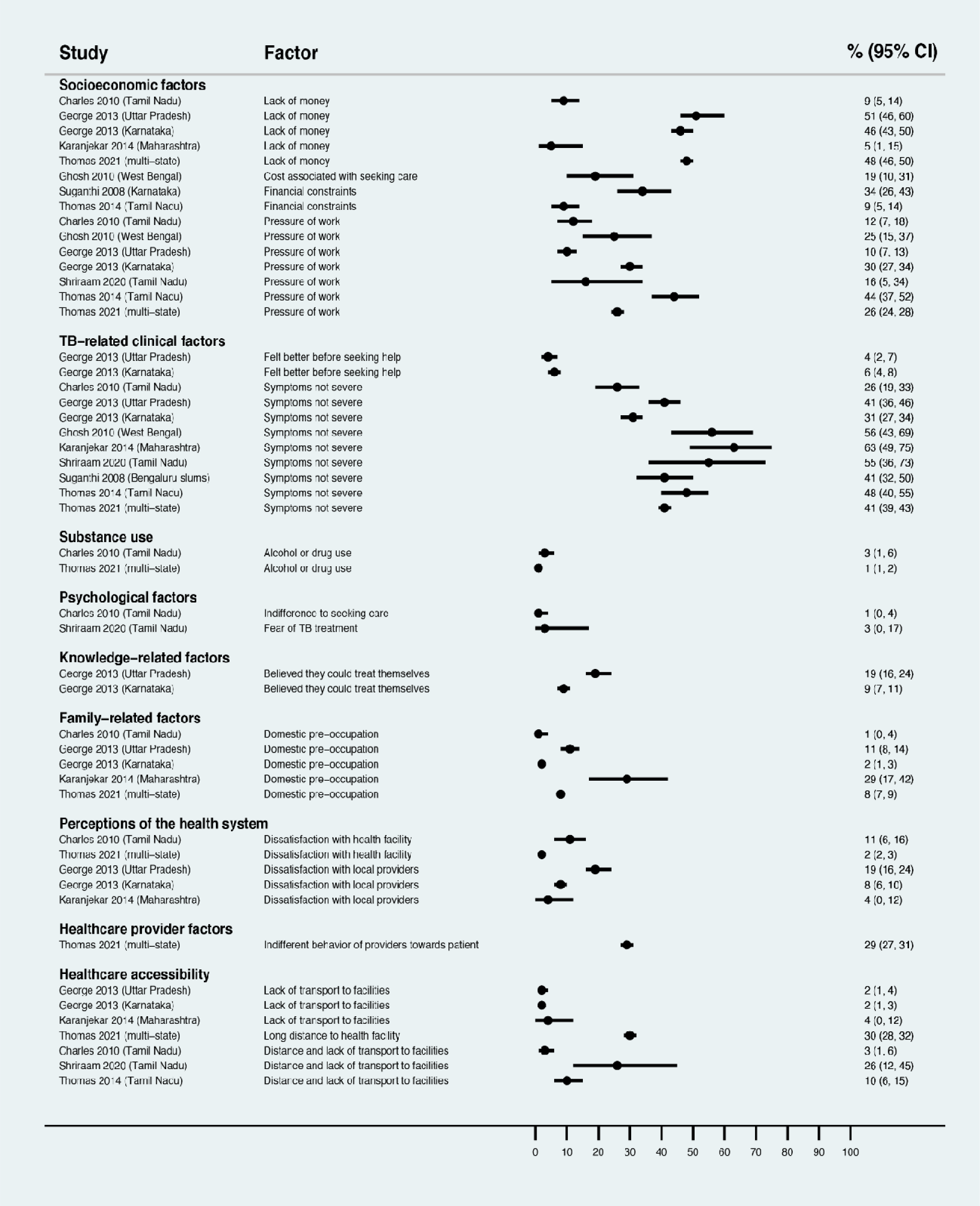
Reasons that individuals in the population had not sought care for tuberculosis-associated symptoms (Gap 1). Estimates represent the percentage of individuals interviewed who reported a given reason for not seeking care. The denominator for all percentages only includes individuals who had not sought care for TB symptoms in population-based surveys. CI, confidence interval; TB, tuberculosis.

### Gap 2—Barriers to completing the appropriate diagnostic workup for presumptive TB

#### Characteristics and quality of the included studies

Across searches spanning January 1, 2000, to May 17, 2021, we screened titles and abstracts of 3,244 unique reports and identified 302 for full text review, of which 20 met inclusion criteria (Fig A in S2 Appendix). Of these, 15 studies presented findings on factors associated with not completing steps of the diagnostic workup, of which 3 reported on non-pursual of diagnostic workup despite referral [40–42]; 4 reported on non-completion of sputum microscopy evaluation [43–46]; 2 reported on chest X-ray non-completion [47,48]; and 6 reported on non-completion of NAAT, line probe assay, or mycobacterial culture among people at higher risk for drug-resistant TB [47,49–53]. 10 studies described reasons for not completing the diagnostic workup, of which 2 reported on non-completion of sputum microscopy evaluation from patient interviews [44,54], 2 reported on chest X-ray non-completion from patient interviews [55,56], and 6 reported on NAAT non-completion from evaluation of health records [49–52,57,58] (Table C in S2 Appendix).

19 of the studies were conducted in 11 of India’s states and union territories (Table C in S2 Appendix). In addition, 1 study reported findings from multiple states. 4 of the studies reported findings from Bihar, Madhya Pradesh, and Chhattisgarh, which are some of India’s poorest states [41,45,51,58]. 7 studies were conducted in urban areas, 6 in rural areas, and 7 in both.

All studies involved high-quality sampling of patients (i.e., random or comprehensive patient sampling). 2 studies were low quality for sample size [53,56]. 8 studies did not report the time frame of research fieldwork, while 3 studies collected data or followed-up patients >3 months after presentation (low quality). 2 studies collected data or followed-up patients 1 to 3 months after presentation (medium quality). 3 studies relied on self-report by the government TB program to determine outcomes (medium to low quality) (Table C in S2 Appendix).

#### Factors associated with non-pursual of the TB diagnostic workup despite referral

Findings varied regarding factors associated with non-pursual of the diagnostic workup despite referral (Fig 3). The impact of gender was mixed. Greater symptom severity—having hemoptysis or symptoms other than cough—was associated with lower adjusted risk of non-pursual of workup, as was having a prior TB treatment history. In contrast, patients with a family history of TB had higher adjusted risk of non-pursual of workup. Patients with alcohol use or missing information regarding age or HIV status in medical records also had higher adjusted risk of non-pursual of workup.

**Fig 3.**
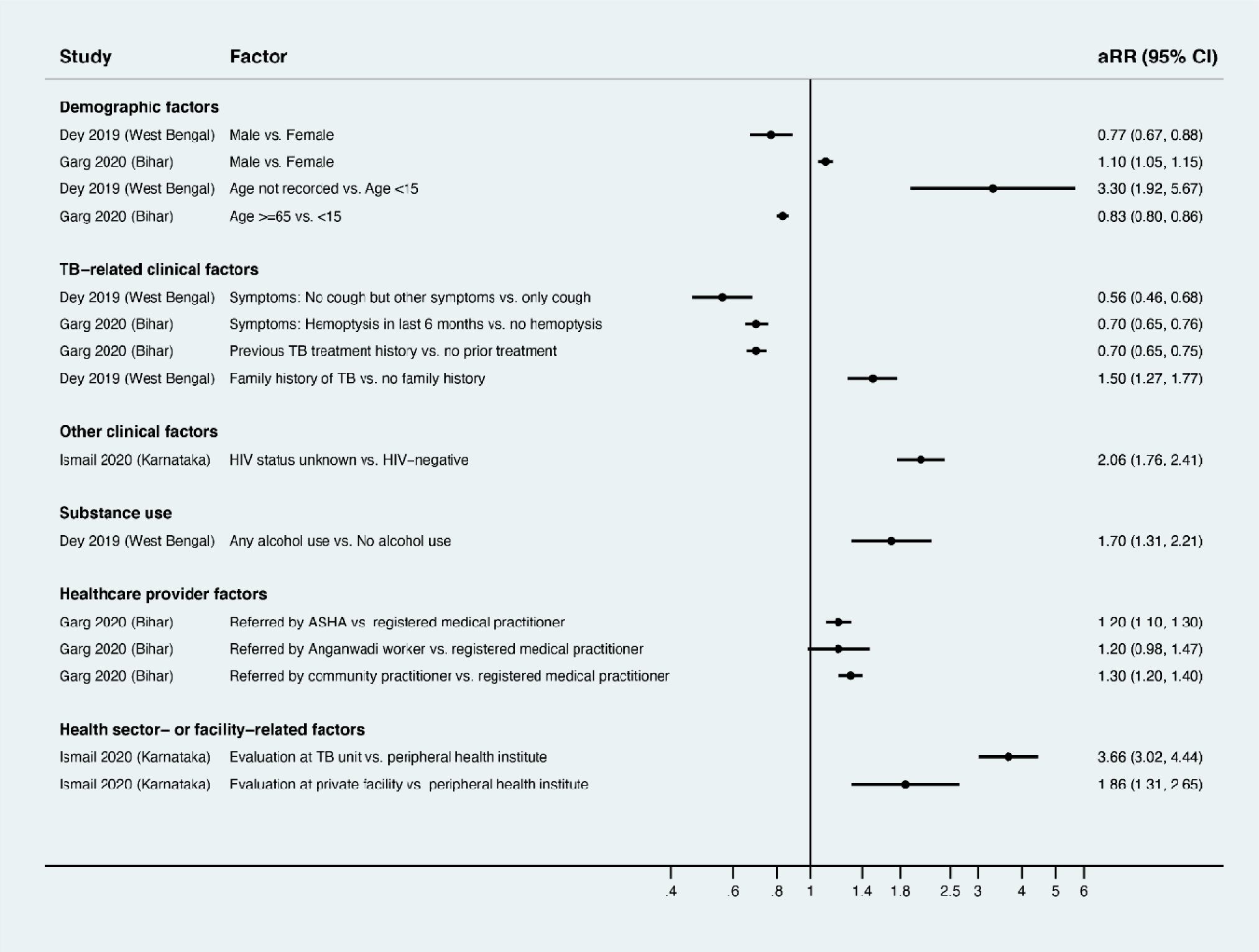
Factors associated with non-pursual of TB diagnostic workup after referral among individuals with presumptive TB (Gap 2). All studies used multivariable relative risk regression, with findings reported as adjusted risk ratios. Only statistically significant findings are presented. Estimates greater than 1 represent increased adjusted risk of non-pursual of diagnostic workup; estimates less than 1 represent decreased adjusted risk of non-pursual of diagnostic workup. ASHA, accredited social health activist; CI, confidence interval; aRR, adjusted relative risk; TB, tuberculosis.

Patients referred by non-physicians—accredited social health activists (ASHAs; government-supported community health workers) or Anganwadi workers (government-supported child health workers)—had higher adjusted risk of non-pursual of diagnostic workup, as compared to patients referred by registered medical practitioners, who often have MBBS degrees (i.e., physicians). Patients referred by healthcare providers at TB units (i.e., government TB centers) or private sector facilities also had higher adjusted risk of non-pursual of diagnostic workup, as compared with patients referred from government peripheral health institutes (i.e., primary health centers).

#### Factors associated with non-completion of sputum microscopy evaluation

1 study conducted a multivariable logistic regression analysis evaluating factors associated with non-completion of sputum microscopy evaluation [44]. This study found older age (>50 years), shorter symptom duration (<=15 days), and lack of support to accompany patients to clinic were associated with higher adjusted odds of non-completion of sputum microscopy evaluation (Fig 4). Patients who were not informed by providers about concern for TB—or who did not remember being informed—had considerably higher adjusted odds of non-completion of sputum microscopy evaluation. In studies that only conducted unadjusted analyses, male sex [43] and living a greater distance from a designated microscopy center [46] were associated with non-completion of sputum microscopy evaluation (Table D in S2 Appendix) [49–53]. 1 study also found the odds of non-completion was higher with use of sputum microscopy alone as compared to a later period when Xpert MTB/RIF was rolled out for initial sputum testing [45].

**Fig 4.**
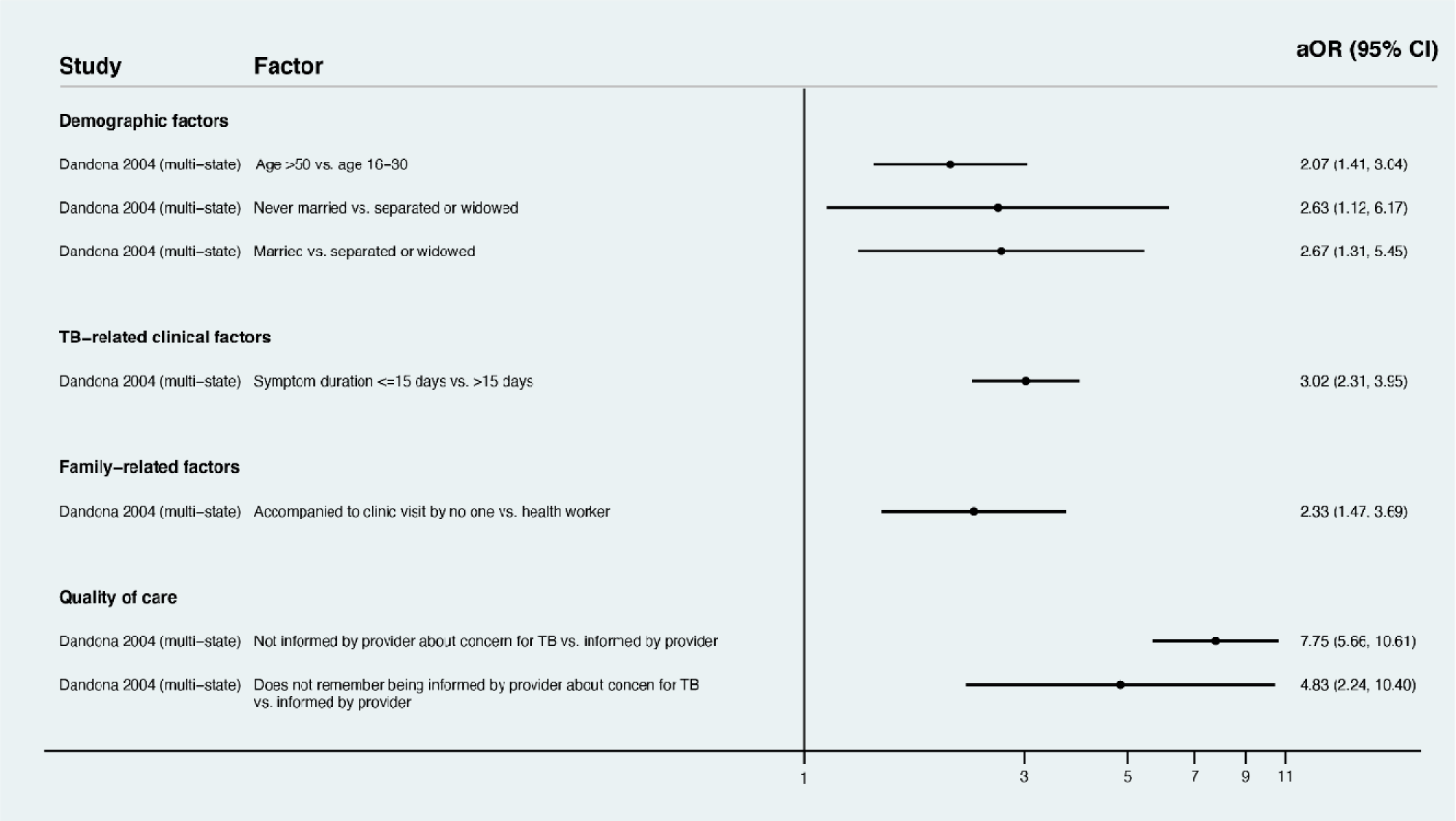
Factors associated with non-completion of sputum microscopy evaluation in a multi-site study in India. [44] **(Gap 2)**. The study used multivariable logistic regression, with findings reported as adjusted odds ratios. Only statistically significant findings are presented. Estimates greater than 1 represent increased adjusted odds of non-completion of sputum microscopy evaluation; estimates less than 1 represent decreased adjusted odds of non-completion of sputum microscopy evaluation. CI, confidence interval; OR, adjusted odds ratio; TB, tuberculosis.

#### Reasons reported by patients for non-completion of sputum microscopy evaluation

Across 2 studies, the most common reasons reported for non-completion of sputum microscopy evaluation were work-related barriers; improvement in symptoms; fear, stigma, or lack of motivation in relation to the concern for TB; and health system-related barriers, especially negative interactions with, or lack of availability of, healthcare providers (Fig 5).

**Fig 5.**
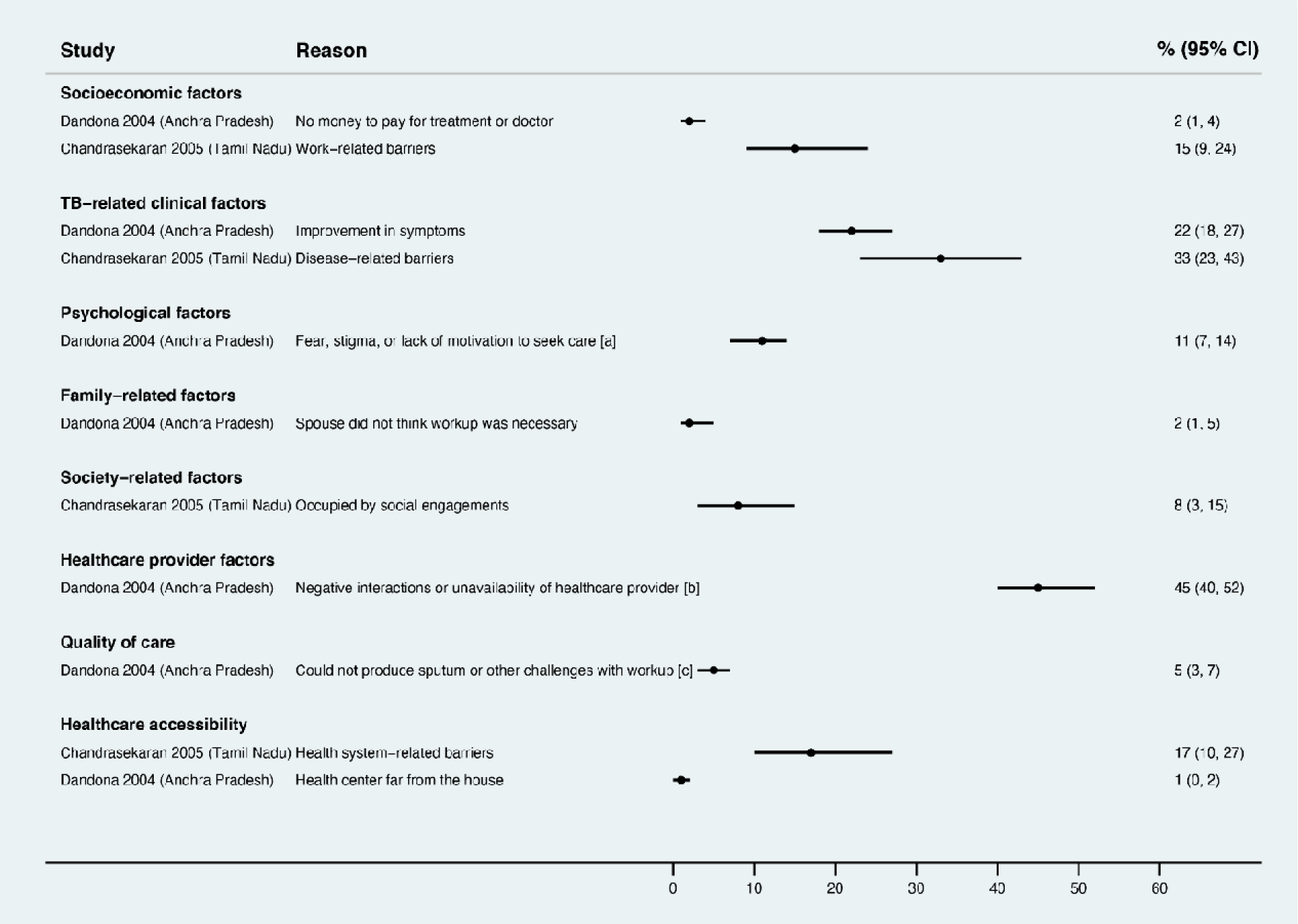
Reasons reported by individuals with presumptive TB for non-completion of sputum microscopy evaluation (Gap 2). Estimates represent the percent of individuals interviewed who reported a given reason for non-completion of sputum microscopy evaluation. [a] “Fear, stigma, or lack of motivation to seek care” summarizes the following reasons: no time, too busy, scared of TB, did not think further workup was necessary, could not go due to ill health, did not go because knew of other patients who were not cured by government care; [b] “Negative interactions or unavailability of healthcare providers” summarizes: provider not aware three sputum samples were needed, lab personnel did not behave well towards the patient, had to wait too long at the center, lab personnel or doctor was not available; [c] “Could not produce sputum or other challenges with workup” summarizes: could not produce enough sputum, referred for x-ray. CI, confidence interval.

#### Factors associated with chest X-ray non-completion

Across 2 studies, socioeconomic factors were strongly associated with higher adjusted risk of chest X-ray non-completion—specifically not being able to afford X-rays in the private sector and being below the poverty line (Fig 6). Structural barriers were also strongly associated with higher adjusted risk of chest X-ray non-completion—specifically being >30 kilometers away from a public facility with chest X-ray or initial evaluation at a district hospital (versus a smaller sub-district hospital). Patients who had not been informed a chest X-ray was needed for their diagnostic workup had higher adjusted odds of non-completion.

**Fig 6.**
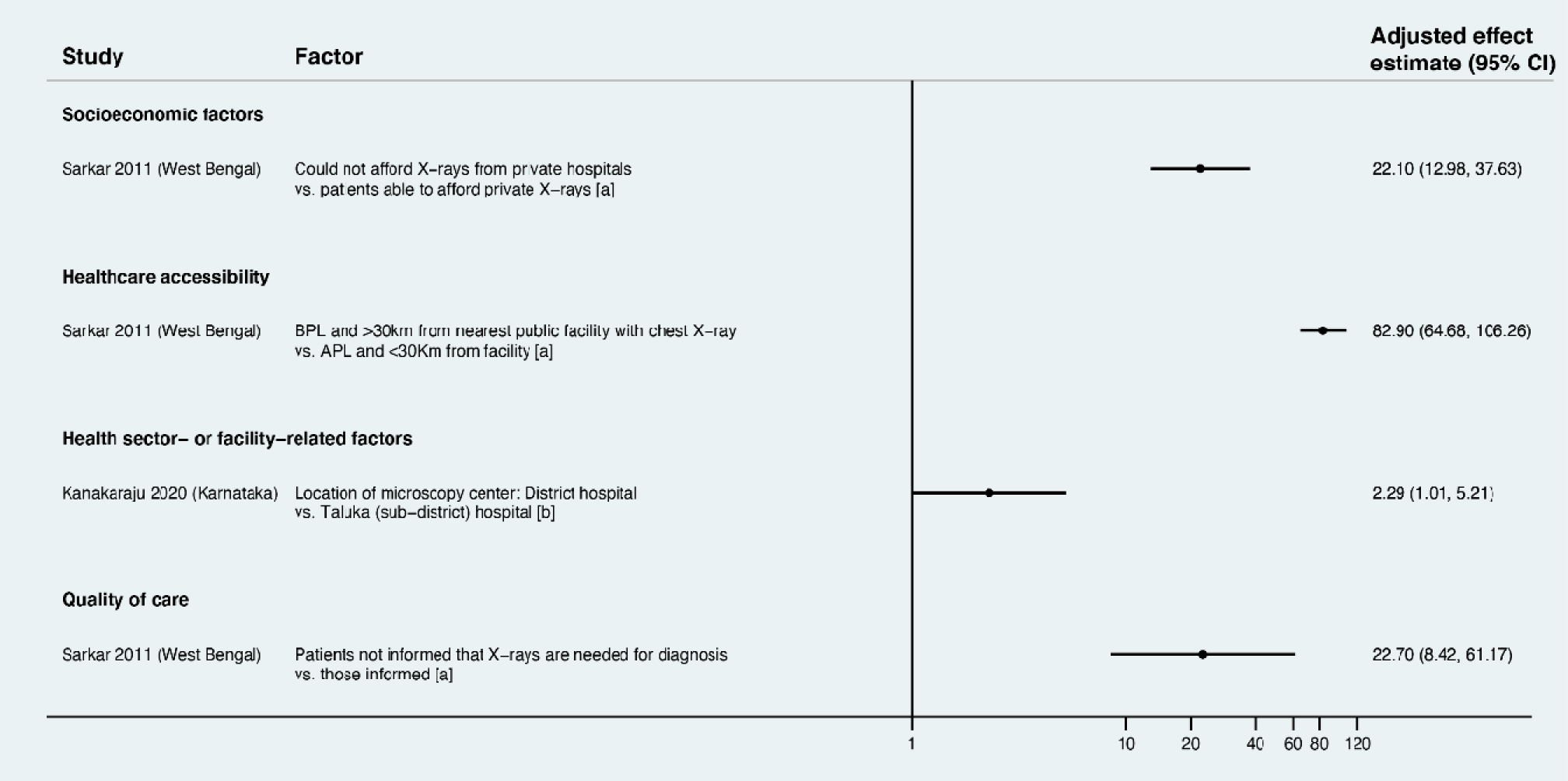
Factors associated with non-completion of chest X-ray as part of the TB diagnostic workup (Gap 2). Only statistically significant findings are presented. Estimates greater than 1 represent increased adjusted odds or risk of chest X-ray non-completion; estimates less than 1 represent decreased adjusted odds or risk of chest X-ray non-completion. [a] Effect estimate is an adjusted odds ratio; [b] effect estimate is an adjusted relative risk ratio. APL, above poverty line; BPL, below poverty line; CI, confidence interval; TB, tuberculosis.

#### Reasons reported by patients for chest X-ray non-completion

Common reasons for chest X-ray non-completion included work-related constraints, improvement of symptoms, consultation of other providers (i.e., care seeking at multiple sites), and not having been informed by providers about the need for further workup (Fig 7).

**Fig 7.**
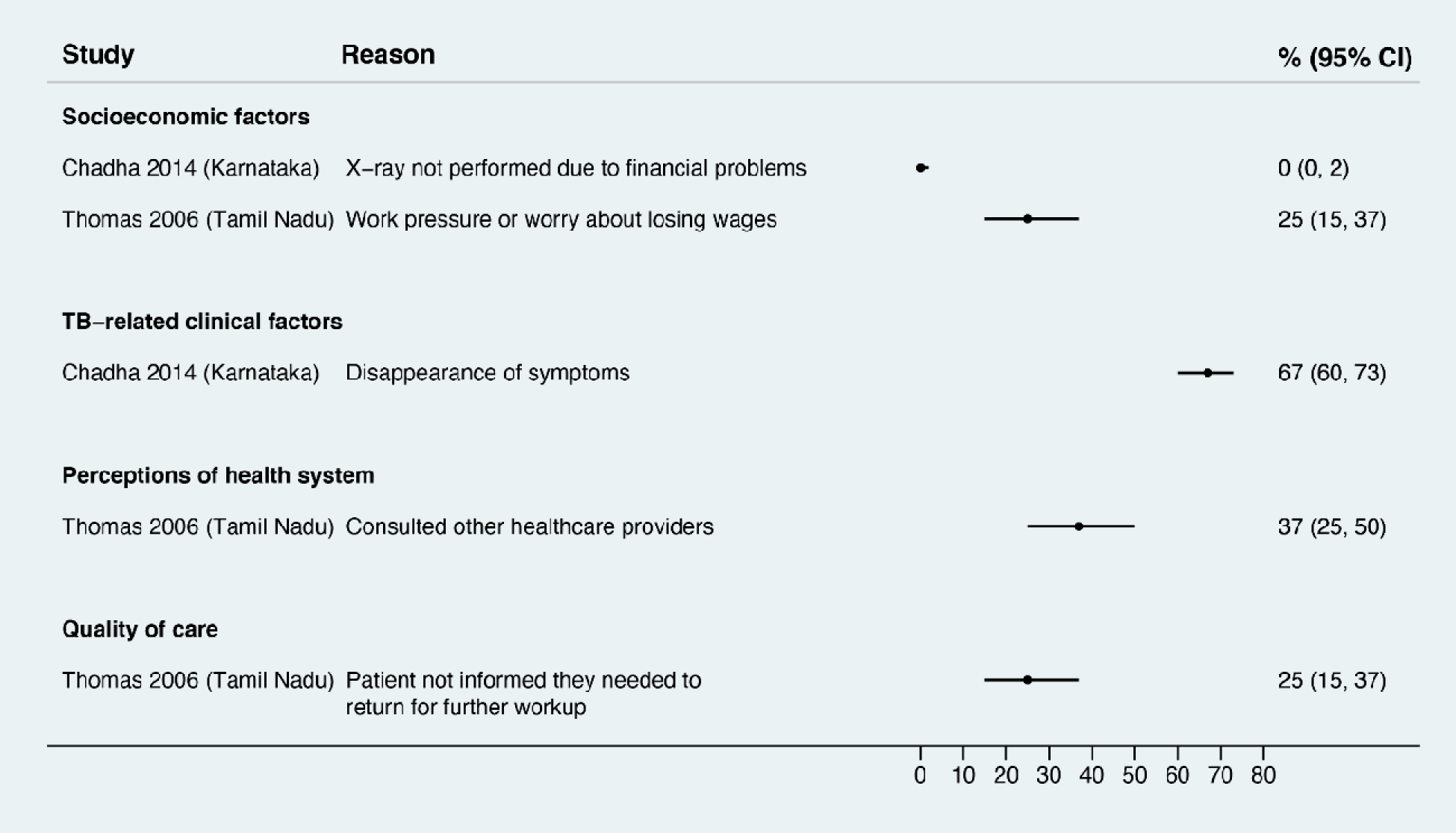
Reasons reported by patients for non-completion of chest X-ray (Gap 2). Estimates represent the percentage of patients who reported a given reason for non-completion of chest X-ray. CI, confidence interval; TB, tuberculosis.

#### Factors associated with non-completion of NAAT, line probe assay, or mycobacterial culture

1 study conducted a multivariable relative risk regression analysis evaluating factors associated with NAAT non-completion for patients at high risk for drug-resistant TB (Fig 8). Patients who were older (>=65 years old), had extrapulmonary disease, had any indication for NAAT other than treatment failure, and who were evaluated at medical colleges had higher adjusted risk of not undergoing NAAT [51]. In studies that only conducted unadjusted analyses, individuals with sputum negative or extrapulmonary TB, whose indication for testing was having HIV, and who were evaluated at a medical college had higher risk of NAAT non-completion (Table D in S2 Appendix) [47,49–53]. One study also found that drug susceptibility testing (DST) non-completion was higher before the rollout of line probe assay in an unadjusted analysis [53].

**Fig 8.**
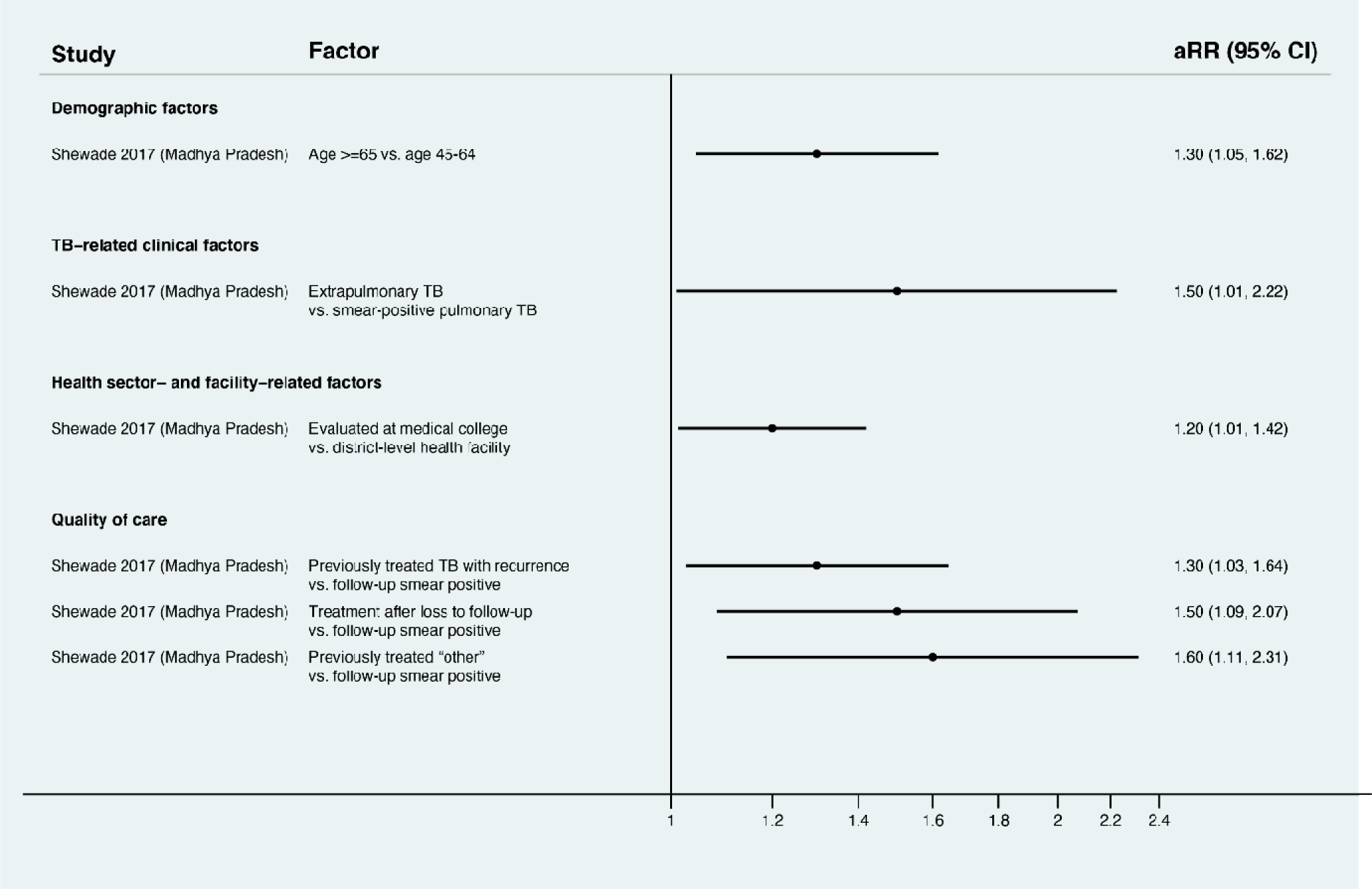
Factors associated with non-completion of NAAT testing in a study conducted in Bhopal, Madhya Pradesh (Gap 2). The study used multivariable relative risk regression, with findings reported as adjusted risk ratios. Only statistically significant findings are presented. Estimates greater than 1 represent increased adjusted risk of NAAT non-completion; estimates less than 1 represent decreased adjusted risk of NAAT non-completion. NAAT, nucleic acid amplification test; CI, confidence interval; aRR, adjusted relative risk.

#### Reasons identified in health system records for non-completion of NAAT, line probe assay, or mycobacterial culture

A major reason patients did not undergo NAAT (in 5 studies) or mycobacterial culture (in 1 study) was that healthcare providers missed identifying (or “line listing”) patients with DR TB risk factors who merited additional testing, with the proportion of eligible patients who were missed ranging from 8% to 54% [49–52,57,58] (Fig 9). Another common reason for non-completion of NAAT (in 4 studies) or mycobacterial culture (in 1 study) was loss of sputum samples during transfer from health centers to laboratories for testing, with the proportion of eligible patients whose samples were lost ranging from 3% to 32% [49–52,57].

**Fig 9.**
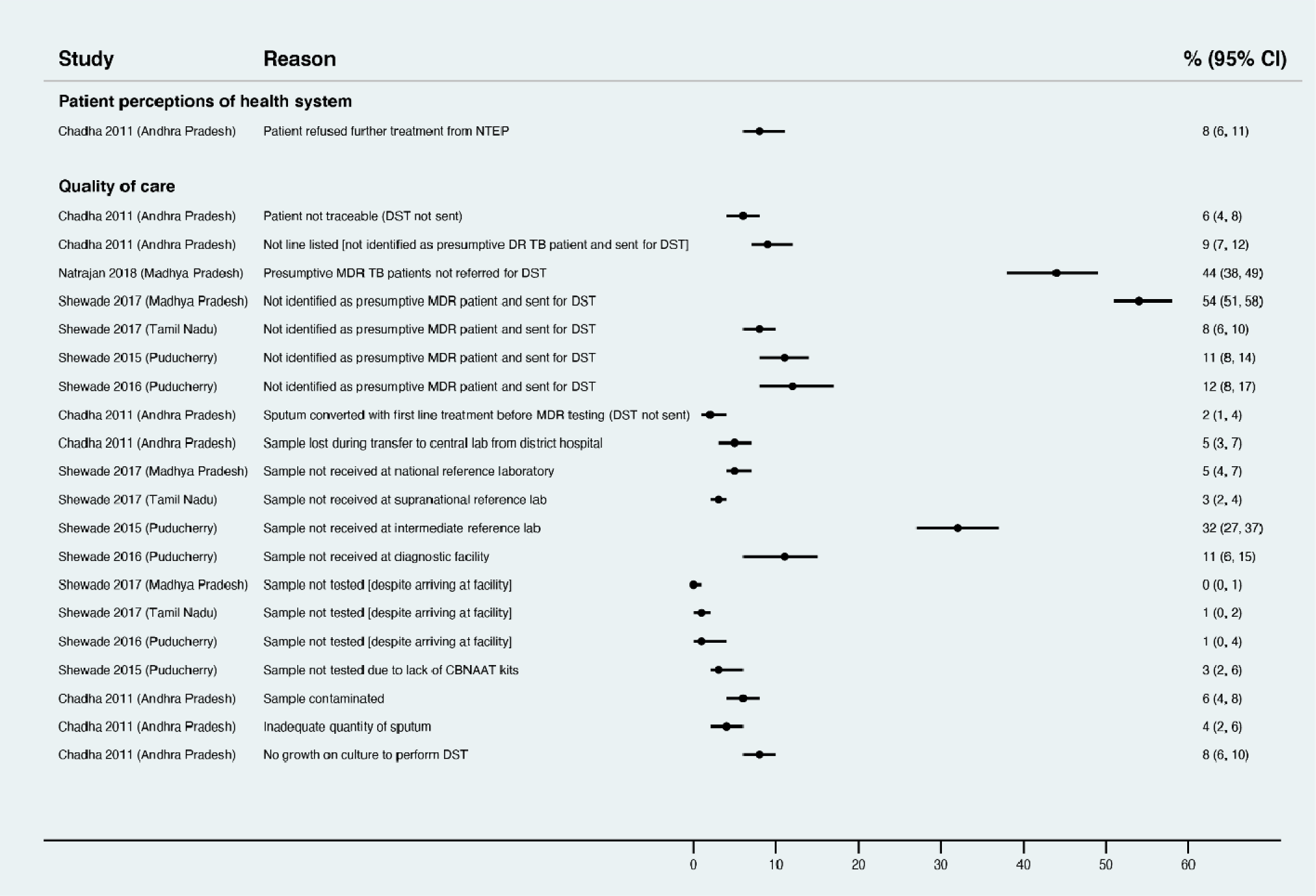
Reasons in health system records for non-completion of NAAT or mycobacterial culture (Gap 2). Estimates represent the percent of all patients who were eligible for drug susceptibility testing who experienced a given reason for non-completion of NAAT or mycobacterial culture. All studies evaluated NAAT non-completion as the outcome except for Chadha et al. 2011, which evaluated non-completion of mycobacterial culture. CI, confidence interval; NAAT, nucleic acid amplification testing.

### Gap 3—Barriers to starting on, or registering in, treatment for individuals diagnosed with active TB

#### Characteristics and quality of the included studies

Across searches for studies spanning January 1, 2000, to May 17, 2021, we screened titles and abstracts of 3,244 unique reports and identified 302 reports for full text review (Fig A in S3 Appendix). Of these, 25 studies met the inclusion criteria of evaluating patients diagnosed with active TB who did not start on, or get registered in, treatment, which we also refer to as pretreatment loss to follow-up (PTLFU). Note that the definitions varied across studies, with most (though not all [64]) studies considering PTLFU as including patients who left the public sector for private sector TB care. Among included studies, 18 reported factors associated with PTLFU, and 11 reported reasons for PTLFU (Table C in S3 Appendix). Studies were conducted in 11 of India’s states and union territories. In addition, 4 studies collected data from more than one state [44,65–67], and 1 study reported findings from a nationally-representative sample of households [66]. Nine studies were conducted in urban areas, 6 in rural areas, and 10 in both.

All studies involved a high-quality (i.e., random or comprehensive) sampling strategy. 8 studies were low quality with regard to sample size [53,58,68–73]. 13 studies did not report time frame of research fieldwork while 3 studies assessed outcomes >3 months after presentation (low quality). 3 studies assessed outcomes 1-3 months after presentation (medium quality). 7 studies relied on self-reported outcomes by the government TB program, without verification through patient tracking by the research team (low quality) (Table C in S3 Appendix).

18 studies evaluated PTLFU in adults with confirmed or presumed drug-susceptible TB. 6 studies evaluated adults with drug-resistant TB, while 1 study evaluated children with drug-susceptible or drug-resistant TB (Table C in S3 Appendix). We present findings separately for adults with drug-susceptible TB, adults with drug-resistant TB, and children with TB.

#### Factors associated with PTLFU in adults with drug-susceptible TB

We identified commonalities in patient-, family-, and society-related factors associated with PTLFU for drug-susceptible TB patients across adjusted analyses (Fig 10) and unadjusted analyses (Table D in S3 Appendix). In 1 adjusted [64] and 2 unadjusted analyses [43,74], older age—classified as greater than 44, 50, or 64 years depending on the study—was associated with higher PTLFU risk. Lower socioeconomic status—assessed using wealth or educational attainment in 1 adjusted analysis [66] and illiteracy in another unadjusted analysis [72]—was associated with higher PTLFU risk. Patients who traveled more than 10 kilometers in 1 unadjusted analysis [46]—or who lived in rural areas but sought medical evaluation in cities in 1 adjusted and 1 unadjusted analysis [64,72]—had higher PTLFU risk. 2 studies found patients with a previous TB history had higher adjusted PTLFU risk [42,64].

**Fig 10.**
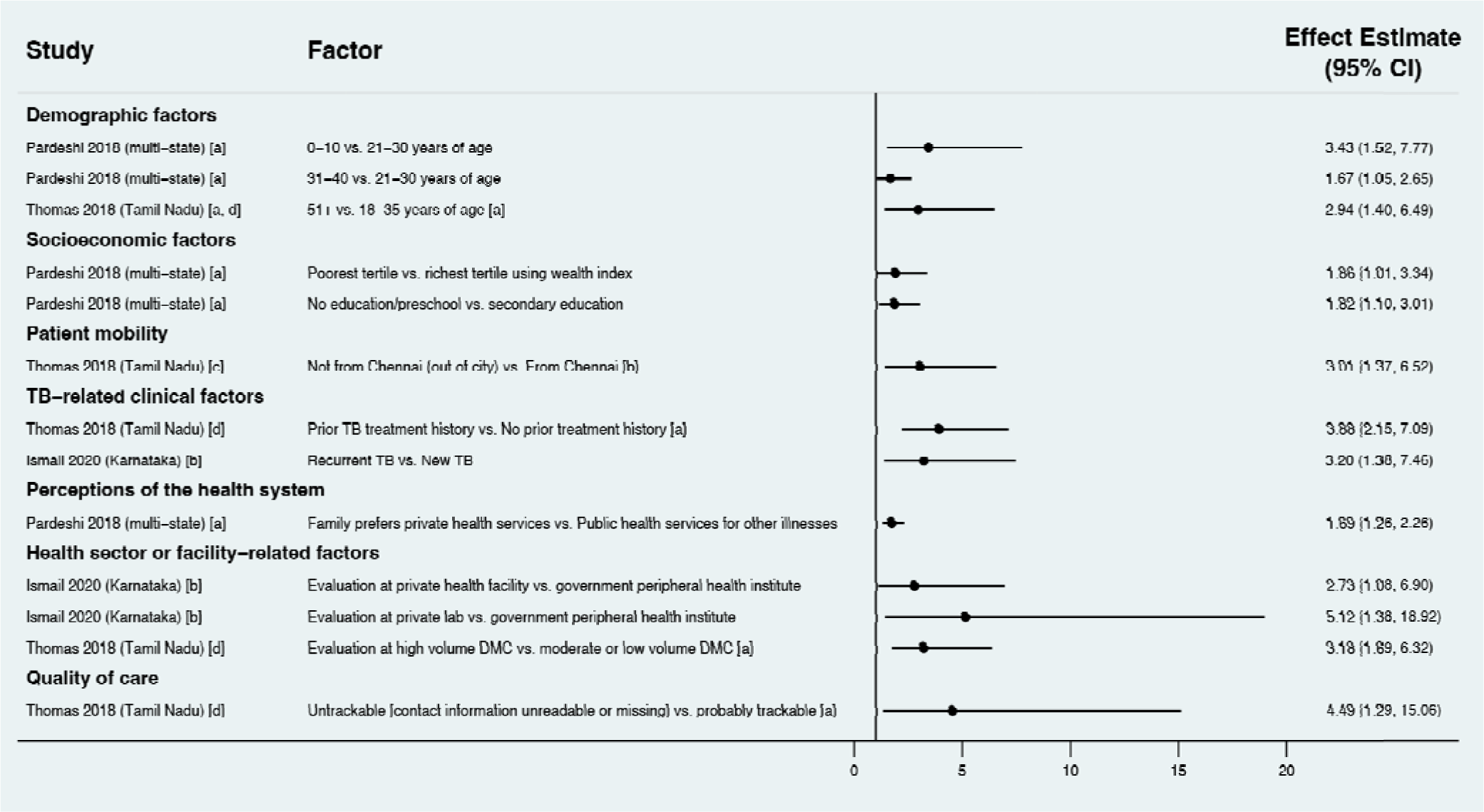
Factors associated with pretreatment loss to follow-up after diagnosis among adult patients with drug-susceptible TB (Gap 3). All studies reported findings as adjusted odds ratios, except for Ismail et al., which reported findings as adjusted relative risk ratios. Only statistically significant findings are presented. Estimates greater than 1 represent increased adjusted odds or risk of PTLFU; estimates less than 1 represent decreased adjusted odds or risk of PTLFU. [a] analysis in which the outcome was non-registration in treatment; [b] analysis in which the outcome was not starting treatment; [c] outcome was non-initiation of TB treatment; [d] outcome was non-registration in the TB program. CI, confidence interval; DMC, designated microscopy center; TB, tuberculosis.

Health system-related factors were also associated with PTLFU. In 1 study, patients whose families preferred private sector (rather than public sector) health services had higher adjusted odds of PTLFU [66]. Similarly, patients who were initially evaluated at private sector labs or facilities (as compared to public primary health centers) had higher adjusted PTLFU risk [42]. Patients evaluated at high-volume diagnostic facilities (e.g., tertiary care hospitals) had higher adjusted odds of PTLFU [64]. In 2 unadjusted analyses, patients diagnosed by community active case finding strategies—as compared with those diagnosed by health facility-based case finding (i.e., routine care seeking)—had higher odds of PTLFU [74,75]. Patients who were not informed of their TB diagnosis—because they did not return to the health facility to pick up test results or because a family member informed them—had higher odds of PTLFU in 1 unadjusted analysis [44]. Patients whose contact information in the diagnostic facility register was missing or unreadable (making them untrackable by providers) had higher adjusted odds of PTLFU in 1 study [64].

#### Reasons for PTLFU in adults with drug-susceptible TB

Patient-, family, or society-related reasons for PTLFU among drug-susceptible TB patients included financial and job constraints preventing patients from returning to health facilities to start therapy [68,76] (Fig 11). Patient mobility was a barrier across multiple studies [67,68,77], with nearly one-third of patients in 1 study experiencing PTLFU due to temporary job-related migration [68]. Psychological reasons were common, with 5% to 25% of patients across multiple studies citing concerns about TB stigma, disbelief in their diagnosis, or treatment refusal as contributing to PTLFU [44,67,72,76,77]. Death before treatment initiation affected 4% to 40% of patients across multiple studies [64,67,72,74,77].

**Fig 11.**
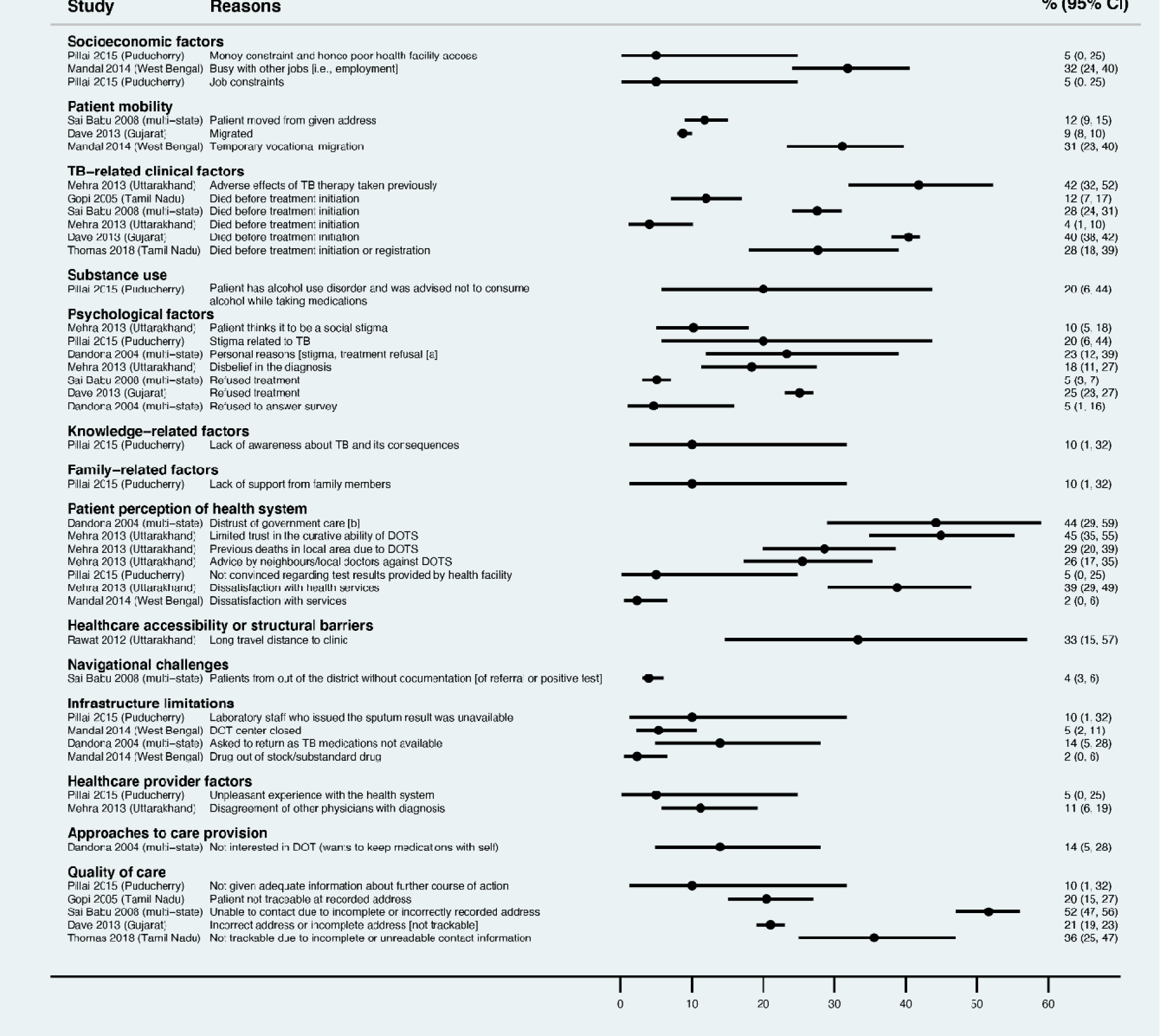
Reasons for pretreatment loss to follow-up among adults with drug-susceptible TB (Gap 3). Estimates represent the percentage of individuals interviewed who reported a given reason for not starting on, or registering in, TB treatment. [a] summarized the following: “no time or busy, afraid that someone would come to know of disease, was very sick, and did not know about TB treatment;” [b] summarized the following responses: “did not want treatment at a government center, did not have belief in government doctors, no confidence in the doctor, and unable to meet the doctor.” CI, confidence interval; DOT, directly observed therapy; TB, tuberculosis.

With regard to health system-related factors, across multiple studies that evaluated public sector TB care, 2% to 35% of patients reported negative perceptions of government health services as contributing to PTLFU [44,68,72,76]. Infrastructural limitations—including HCP absenteeism, closure of directly observed therapy (DOT) centers, and stockouts of TB drugs—also contributed to PTLFU across three studies [44,68,76]. Quality of care was also a barrier. In particular, four studies found poor recording of patient contact information resulted in HCPs being unable to find patients to tell them their diagnostic results and encourage them to start treatment, a finding reported by 21% to 52% of patients [64,67,74,77].

#### Factors associated with PTLFU in adults with drug-resistant TB

Of studies evaluating PTLFU among patients with drug-resistant TB, only 1 [78] performed multivariable regression analysis (Table D in S3 Appendix). This study found that patients whose DST indication was having a positive follow-up sputum microscopy result during their previous treatment for drug-susceptible TB had an adjusted relative PTLFU risk that was 6.0 (95%CI 2.3—15.2) compared to patients whose DST indication was presentation with recurrent TB. In addition, patients with drug-resistant pulmonary TB whose sputum microscopy result was missing—suggesting that they had not completed clinical evaluation—had an adjusted relative risk of PTLFU that was 17.1 (95%CI 7.7—39.3) compared to patients who had a positive sputum microscopy result. In 1 unadjusted analysis [53], the odds of PTLFU for drug-resistant TB patients was 4.69 (95%CI 3.15—6.97) higher preceding rollout of line probe assay for DST, compared to the post-rollout time period.

#### Reasons for PTLFU in adults with drug-resistant TB

Two studies also evaluated reasons for PTLFU among patients diagnosed with drug-resistant TB through audits of health system records [57,58] and interviews with patients (Fig 12). As with studies of drug-susceptible TB, inability to track patients due to poor recording of contact information, death before treatment initiation, and refusal of treatment were common reasons for PTLFU.

**Fig 12.**
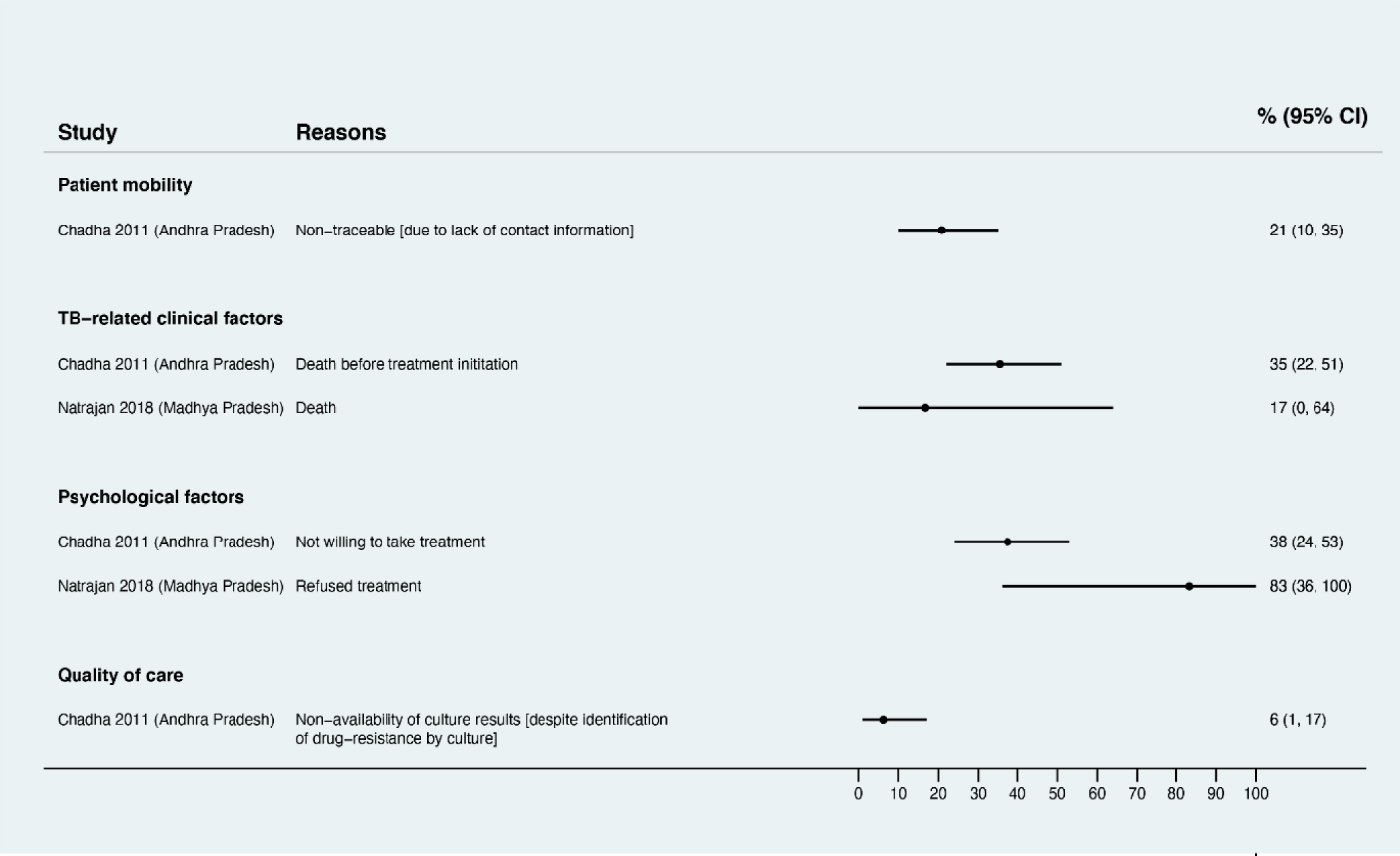
Reasons for pretreatment loss to follow-up among adults with drug-resistant TB (Gap 3). Estimates represent the percentage of individuals interviewed who had a given reason identified for not starting on, or registering in, treatment for drug-resistant TB. CI, confidence interval; TB, tuberculosis.

#### Factors associated with PTLFU in children with drug-susceptible or drug-resistant TB

1 study evaluated factors associated with PTLFU in children with drug-susceptible or drug-resistant TB. For children with either drug-susceptible or drug-resistant TB, those 0 to 4 years had significantly higher odds of PTLFU, as compared to those 10 to 14 years of age (Table D in the S3 Appendix).

### Gap 4—Barriers to achieving treatment success in individuals with TB who start treatment

#### Characteristics and quality of the included studies

Across searches spanning January 1, 2000, to May 19, 2021, we screened titles and abstracts of 3,244 unique reports and identified 302 reports that underwent full text review, of which 104 met Gap 4 inclusion criteria (Fig A in S4 Appendix).

As such, 104 articles were included in the Gap 4 analysis. Of these, 97 presented findings on factors associated with unfavorable treatment outcomes, and 14 presented findings on reasons for unfavorable treatment outcomes (Table C in S4 Appendix). Studies were conducted in 24 of India’s states and union territories. In addition, 11 studies collected data from multiple states [34,44,65,79–86], including 1 study comprising a nationally-representative sample from India’s 2006 TB register [83]. 24 studies were conducted in rural areas, 44 in urban areas, and 29 in both. Except for 5 studies [87–91] where TB was treated in the private sector, all other studies evaluated patients in the public sector (Table C in S4 Appendix).

All included studies involved high-quality (i.e., random or comprehensive) sampling. 20 were low quality with regard to sample size. 47 assessed exposures and outcomes retrospectively from medical records without data collection from TB patients (low quality) (Table C in S4 Appendix).

Given the large number of studies identified, we present findings separately for patient subpopulations: those with new confirmed or presumed drug-susceptible TB (n=25), those with a previous TB treatment history being treated for confirmed or presumed drug-susceptible TB (n=17), those with RR or MDR TB (n=16), those with HIV being treated for TB (n=4), and children with TB (n=3). We also separately present findings for studies involving multiple patient subpopulations—i.e., more than one of the aforementioned subpopulations without disaggregation (n=46) (Table C in S4 Appendix).

#### Factors associated with unfavorable outcomes during TB treatment among new drug-susceptible TB patients

We identified common factors associated with unfavorable treatment outcomes across adjusted analyses (Fig 13) and unadjusted analyses (Table D1 in S4 Appendix) for new drug-susceptible TB patients. With regard to patient-, family-, or society-related factors, male sex was associated with increased risk for unfavorable outcomes in 3 unadjusted analyses [43,92,93]. In 1 adjusted [92] and 3 unadjusted analyses, older age—classified as greater than 44 or 60 years depending on the study—was associated with higher risk of unfavorable outcomes. In 1 adjusted analysis, separated or divorced men had higher risk of suboptimal outcomes than men with married, widowed, or single relationship status [82].

**Fig 13.**
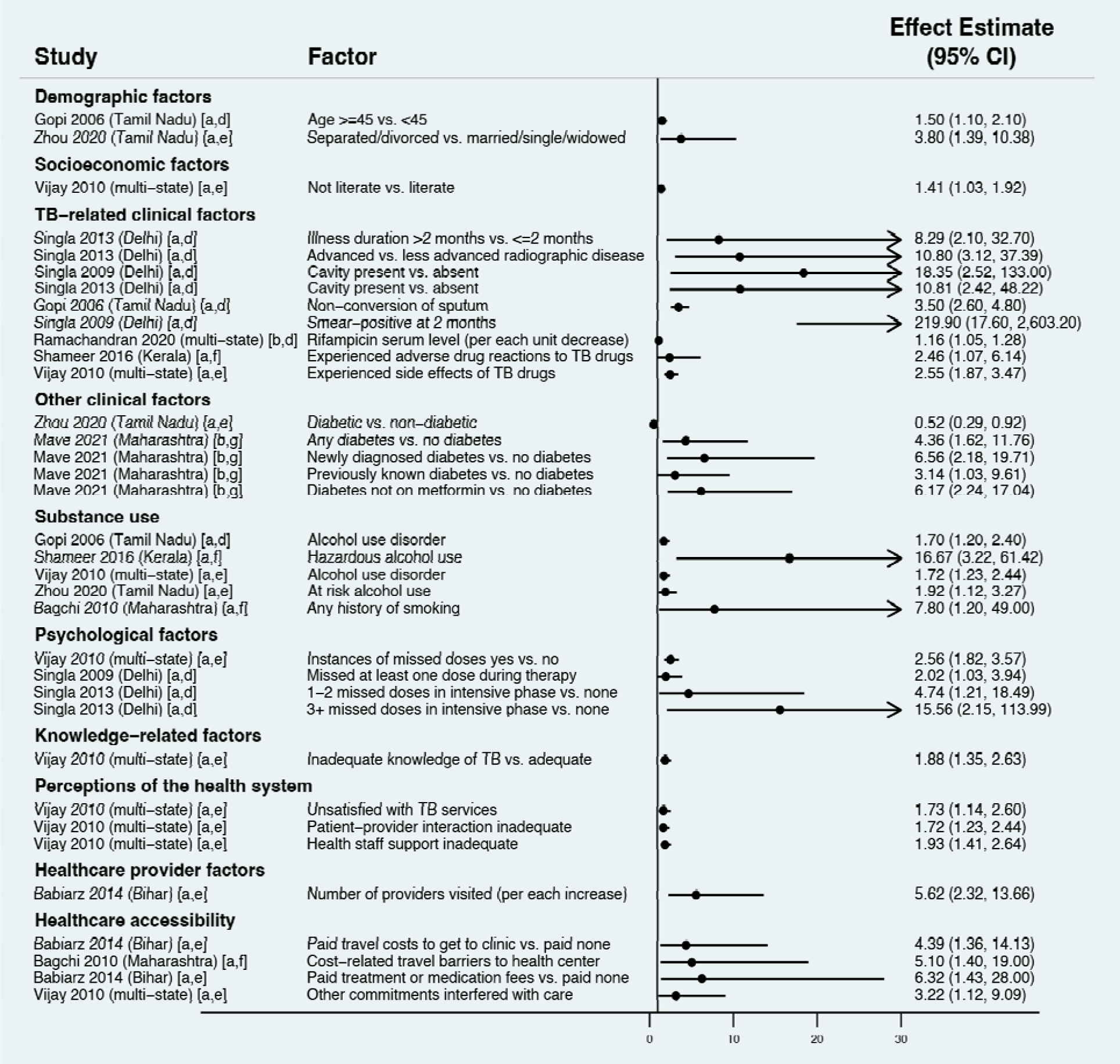
Factors associated with unfavorable outcomes during TB treatment among new confirmed or presumed drug-susceptible TB patients (Gap 4). Estimates greater than 1 represent increased adjusted odds of unfavorable outcomes; estimates less than 1 represent decreased adjusted odds of unfavorable outcomes CI, confidence interval; OR, odds ratio; TB, tuberculosis. [a] Effect estimate is an adjusted odds ratio; [b] effect estimate is an adjusted incidence rate ratio; [c] outcome is any unfavorable treatment outcome; [d] outcome is treatment failure; [e] outcome is loss to follow-up; [f] outcome is medication nonadherence; [g] outcome is death.

In general, lower socioeconomic status was associated with unfavorable outcomes. People who were illiterate or only had primary education were more likely to have unfavorable outcomes in 1 adjusted analysis [81] and 3 unadjusted analyses [92,93,96], as were individuals in the lower socioeconomic strata in 1 unadjusted analysis [93]. Employed individuals and female sex workers were more likely to have unfavorable outcomes in 1 unadjusted analysis [93]. In the same study [93], migrants were at increased risk of unfavorable outcomes, which may reflect challenges related to job roles or patient mobility.

Clinical factors were associated with unfavorable outcomes. Some factors were associated with greater symptom duration or diagnostic delay. In adjusted and unadjusted analyses [97,98], illness >2 months was associated with increased risk of unfavorable outcomes, as was cough >=4 weeks and lag >6 weeks between symptom onset and treatment initiation in unadjusted analyses [92,99]. Other factors indicated advanced TB. Advanced radiological disease (1 adjusted analysis [97] and 1 unadjusted analysis [98]), cavitary disease (2 adjusted analyses [97,98]), and higher sputum smear grade (3 unadjusted analyses [92,97,98]) were associated with unfavorable outcomes. In 2 adjusted and 2 unadjusted analyses, suboptimal treatment response (i.e., lack of sputum conversion) was associated with unfavorable outcomes [92,97,98,100]. Medications-related issues—subtherapeutic rifampin level in 1 adjusted analysis [79] and adverse drug reactions in 2 adjusted analyses [81,101]—were associated with unfavorable outcomes.

With regard to comorbid conditions, diabetes, particularly untreated, was associated with death in 4 adjusted analyses from 1 study [96] and 1 unadjusted analysis [102]; however, an adjusted analysis in another study showed diabetes was protective [82]. Being underweight [82] or having HIV [103] were associated with high unadjusted risk of unfavorable outcomes. Alcohol use (current or past) was associated with unfavorable outcomes in 4 adjusted [81,82,92,101] and 2 unadjusted analyses [80,93]. Patients with current or past smoking had higher risk of unfavorable outcomes in 1 adjusted [104] and 5 unadjusted analyses [81,82,92,93,101]. In 3 adjusted analyses [81,97,98], medication nonadherence was associated with unfavorable outcomes. In 1 adjusted [81] and 1 unadjusted analysis [93], inadequate TB knowledge was associated with increased risk of unfavorable outcomes.

With regard to family- and society-related barriers, not living with one’s family [93], having another family member with active TB [101], and higher perceived TB stigma [101] were associated with increased unadjusted risk of unfavorable outcomes.

Health system-related factors were also associated with unfavorable outcomes. In 1 study [81], dissatisfaction with TB services, poor patient-provider interactions, and inadequate health staff support were associated with higher adjusted odds of loss to follow-up. In unadjusted analyses, other health system factors associated with loss to follow-up included the clinic not providing DOT monitoring and lack of patient address verification by the TB program [81]. Increasing number of providers visited before diagnosis was associated with unfavorable outcomes in 1 adjusted analysis, which is indicative of diagnostic delay due to poor quality of care [105].

Healthcare accessibility challenges also contributed to unfavorable outcomes. In 3 unadjusted analyses [99,101,106], distance to the nearest TB center—classified as more than 1, 2, or 20 kilometers—was associated with unfavorable outcomes. In unadjusted analyses in 1 study, patient perceptions that travel to the health center was a problem, needing to use travel modes besides walking, and concerns about the transportation, distance, or time to reach the health center were associated with unfavorable outcomes [104]. Cost of travel to clinics [104,105] and treatment costs [105] were associated with unfavorable outcomes in other adjusted analyses.

#### Reasons reported by new drug-susceptible TB patients for loss to follow-up or medication nonadherence during treatment

Several studies evaluated reasons for loss to follow-up or medication nonadherence among drug-susceptible TB patients (Fig 14). Patient-, family-, or society-related factors included barriers related to work [80,93,99] and patient mobility [80,93]. Side-effects of TB therapy or early symptom resolution, before completing treatment, were also reported as reasons for loss to follow-up or medication nonadherence across multiple studies [80,93,99]. Nearly one-third of patients in one study stopped therapy due to TB medication side effects [99]. Health system-related factors also contributed to loss to follow-up or medication nonadherence, including having to travel a long distance to the health center and uncooperative health center staff [99].

**Fig 14.**
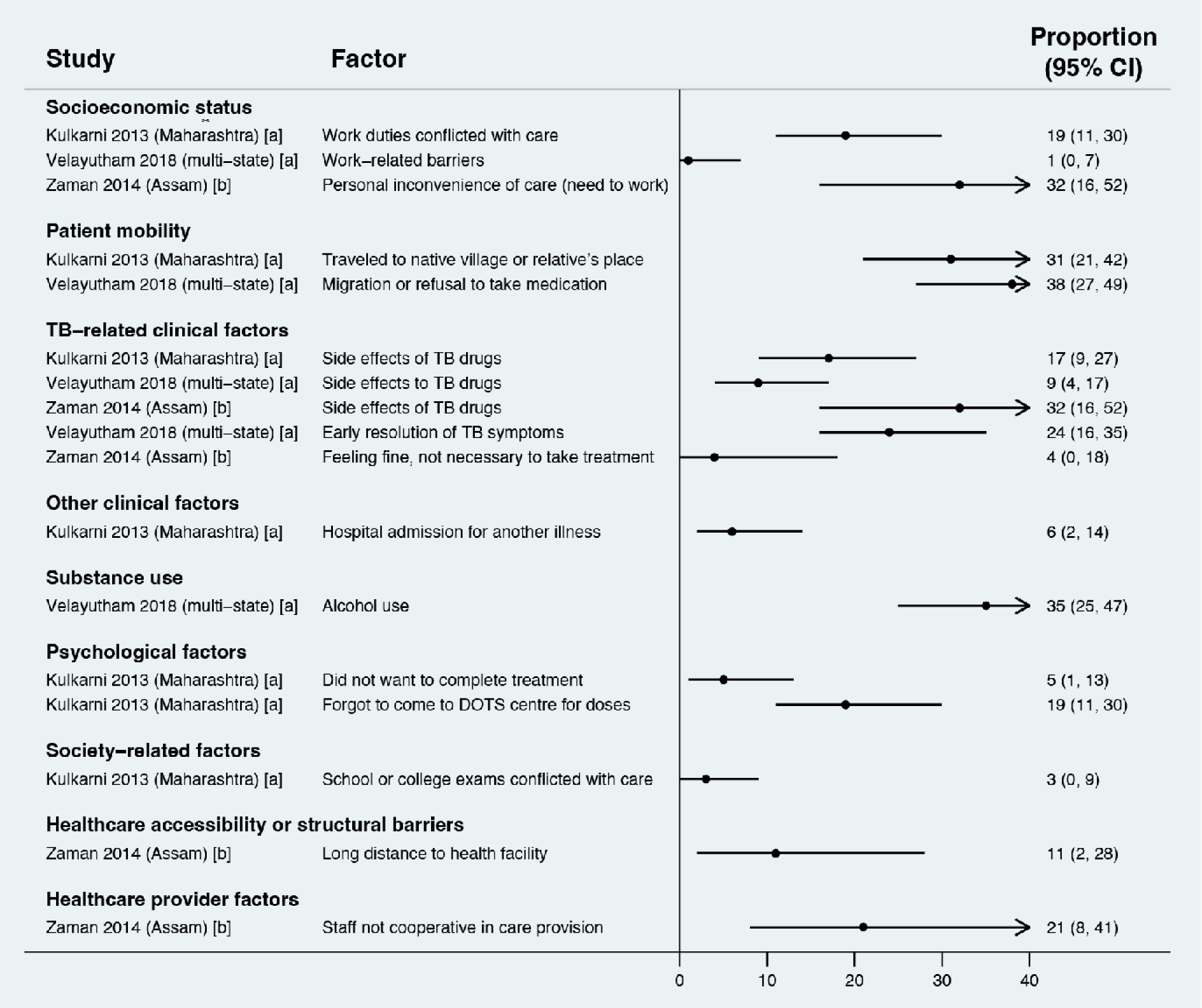
Reasons reported by new confirmed or presumed drug-susceptible TB patients for experiencing unfavorable outcomes during TB treatment in quantitative surveys (Gap 4). Estimates represent the percentage of individuals interviewed who reported a given reason for experiencing unfavorable outcomes. [a] Study reported reasons for loss to follow-up; [b] study reported reasons for medication nonadherence or loss to follow-up. CI, confidence interval; TB, tuberculosis.

#### Factors associated with unfavorable treatment outcomes among previously treated patients with confirmed or presumed drug-susceptible TB

We identified common findings regarding patient-, family-, or society-related factors associated with unfavorable outcomes across adjusted analyses (Fig 15) and unadjusted analyses (Table D2 in S4 Appendix) among previously treated TB patients. In 2 adjusted analysis [83,107] and 2 unadjusted analyses [108,109], male sex was associated with unfavorable outcomes. Illiteracy [105,110] and fewer years of education [105] were also associated with unfavorable outcomes. In contrast, being employed (versus unemployed), which is usually a marker of better socioeconomic status, was associated with unfavorable outcomes in 1 adjusted analysis [110].

**Fig 15.**
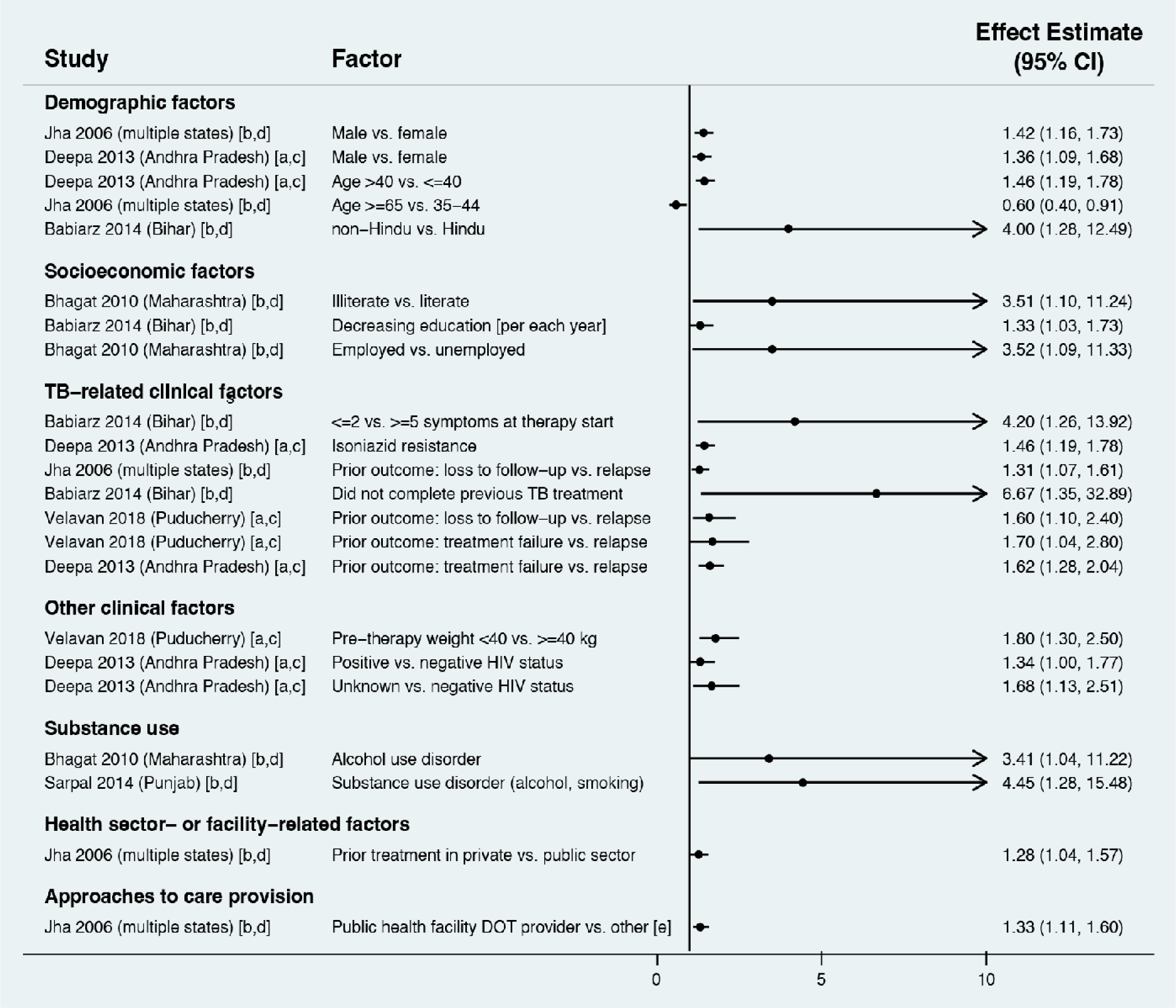
Factors associated with unfavorable outcomes during TB treatment among confirmed or presumed drug-susceptible TB patients with a prior treatment history (Gap 4). Estimates greater than 1 represent increased adjusted odds of unfavorable outcomes; estimates less than 1 represent decreased adjusted odds of unfavorable outcomes CI, confidence interval; OR, odds ratio; TB, tuberculosis. [a] Effect estimate is an adjusted relative risk ratio; [b] effect estimate is an adjusted odds ratio; [c] outcome is any unfavorable treatment outcome; [d] outcome is loss to follow-up; [e] other DOT providers included community providers, medical providers, private practitioners, or non-governmental organizations.

The outcome of a patient’s prior treatment episode was an important predictor of subsequent TB treatment outcomes. As compared to patients who completed their prior treatment—and were therefore considered to have disease relapse—patients who were lost to follow-up during their previous treatment were more likely to have unfavorable outcomes in 3 adjusted [83,105,111] and 4 unadjusted analyses [108,112–114]. Similarly, as compared to patients with disease relapse, those who experienced prior treatment failure had higher risk of unfavorable outcomes in 2 adjusted [107,111] and 3 unadjusted analyses [108,112,113]. Previously treated “others”—a category describing patients with sputum negative or extrapulmonary TB—had lower risk of unfavorable outcomes in 2 unadjusted analyses [109,111]. Drug resistance also influenced subsequent outcomes. In 1 adjusted analysis, isoniazid monoresistance [107] was associated with unfavorable outcomes, as was resistance to isoniazid, ethambutol, and/or streptomycin in 1 unadjusted analysis [115].

Comorbid conditions, including baseline weight <40kg (vs. >=40kg) [111] and alcohol and other substance use disorders [108,110], were associated with unfavorable outcomes. Positive or unknown HIV status in 1 adjusted [107] and 1 unadjusted analysis [109], and not receiving antiretroviral therapy (among people with HIV) in 1 unadjusted analysis [107], were associated with unfavorable outcomes.

Few health system-related factors were assessed in studies of previously treated TB patients. Prior treatment in the private sector was associated with unfavorable outcomes in 1 adjusted [83] and 1 unadjusted analysis [114]. In 1 adjusted analysis, patients who underwent public health facility-based treatment observation—as compared with care provision by community providers, local medical providers, private practitioners, or non-governmental organizations—had higher risk of unfavorable outcomes [83].

#### Reasons reported by previously treated TB patients for loss to follow-up during treatment

Patient-, family, or society-related reasons for loss to follow-up reported by previously treated patients included socioeconomic barriers, such as loss of wages or work [108]; barriers due to patient mobility such as migration outside the clinic’s geographical area [98]; substance use-related barriers such as alcohol use [108]; and society-related barriers such as TB stigma [108] (Fig 16). TB clinical factors such as treatment side effects, long symptom duration, lack of symptom improvement despite treatment, and early symptom improvement (with resulting loss of motivation to continue treatment) contributed to loss to follow-up [98,108]. Notably, in 1 study, nearly half of patients who were lost to follow-up had discontinued treatment due to medication side effects [108]. Lack of faith in the government’s DOTS model was another common reason for loss to follow-up in 1 study [108].

**Fig 16.**
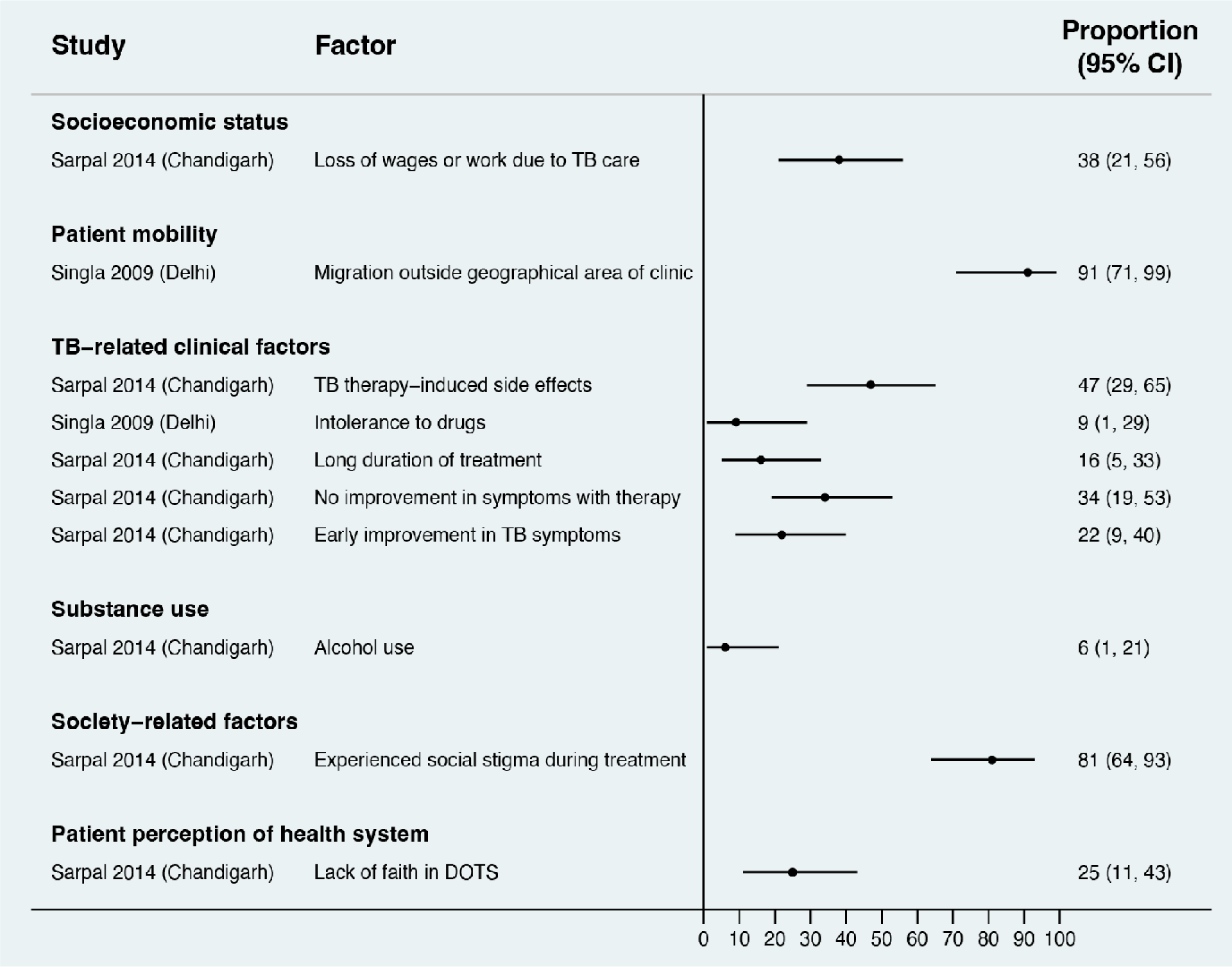
Reasons reported by patients with a prior TB treatment history for experiencing unfavorable outcomes during TB treatment in quantitative surveys (Gap 4). Estimates represent the percentage of individuals interviewed who reported a given reason for experiencing unfavorable outcomes. CI, confidence interval; TB, tuberculosis.

#### Factors associated with unfavorable treatment outcomes among RR or MDR TB patients

We identified common factors associated with unfavorable treatment outcomes across adjusted analyses (Fig 17) and unadjusted analyses (Table D3 in S4 Appendix) for patients with RR or MDR TB. With regard to patient-, family-, or society-related factors, male sex was associated with unfavorable outcomes in 6 adjusted [84,116–118] and 2 unadjusted analyses [119,120]. Older age—classified as more than 35 or 45 years or per each year increase as a continuous variable—was associated with unfavorable outcomes in 4 adjusted analyses [88,116,118,121].

**Fig 17.**
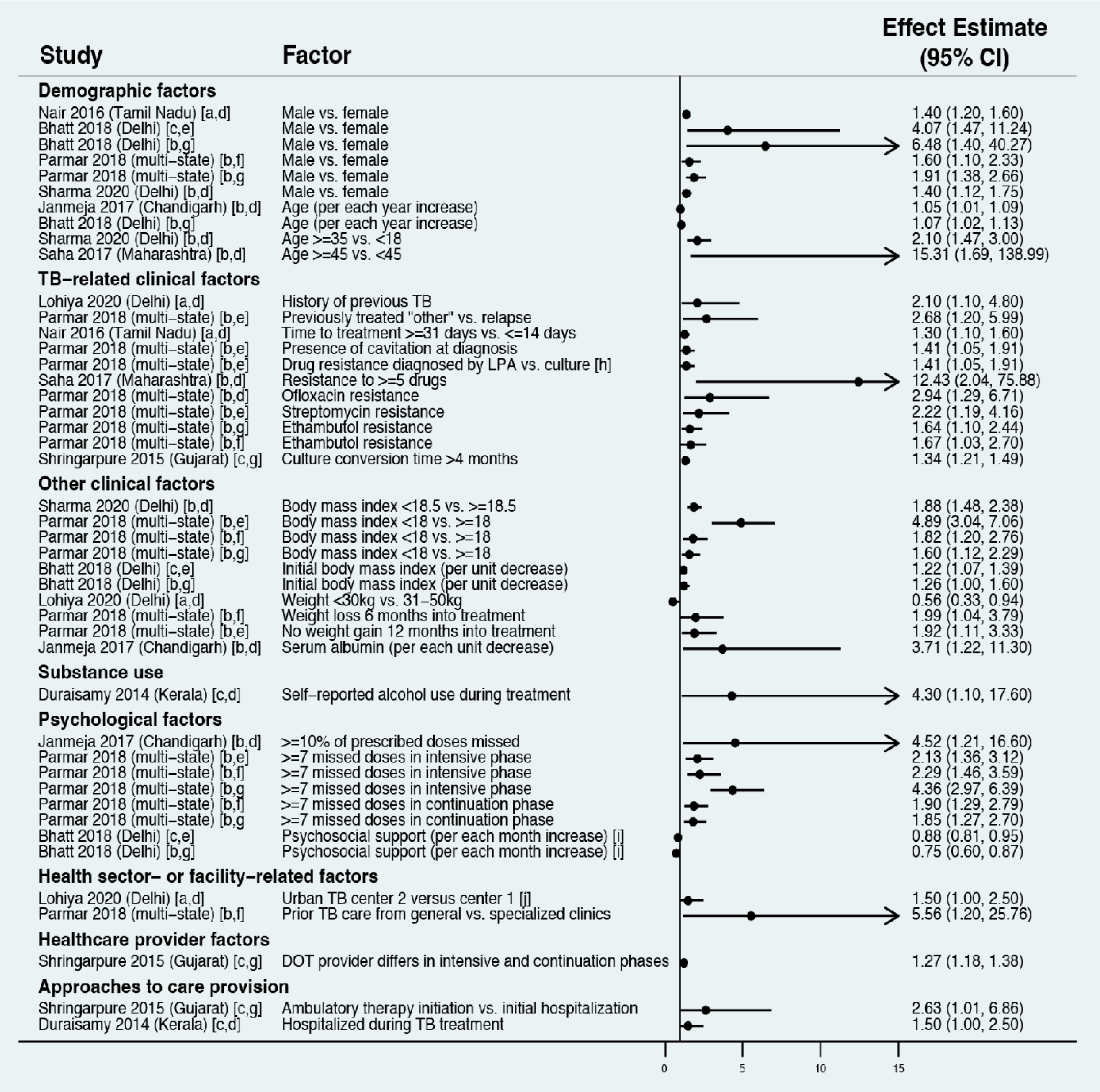
Factors associated with unfavorable outcomes during treatment among RR or MDR TB patients (Gap 4). Estimates greater than 1 represent increased adjusted odds of unfavorable outcomes; estimates less than 1 represent decreased adjusted odds of unfavorable outcomes. [a] Effect estimate is an adjusted relative risk ratio; [b] effect estimate is an adjusted odds ratio; [c] effect estimate is an adjusted hazard ratio; [d] outcome is any unfavorable treatment outcome; [e] outcome is death; [f] outcome is treatment failure; [g] outcome is loss to follow-up; [h] higher mortality among patients diagnosed by line probe assay might reflect survivor bias among patients diagnosed by culture, as many patients diagnosed by culture may have died before starting drug-resistant TB therapy; [g] the reason given by the study authors for poorer outcomes at one of the drug-resistant TB centers was that it took care of more patients who lived in rural areas, rather than within the city of Delhi. CI, confidence interval; OR, odds ratio; MDR, multidrug-resistant; RR, rifampin-resistant; TB, tuberculosis.

Clinical factors influenced RR or MDR TB outcomes. Factors suggesting advanced disease or risk of drug resistance at diagnosis were associated with unfavorable outcomes, including previous TB history in 1 adjusted analysis [122], increasing number of prior TB treatment episodes in 2 unadjusted analyses [84,121], longer time to treatment initiation in 1 adjusted analysis [117], lung cavitation on chest radiography in 1 adjusted analysis [84], and advanced X-ray findings in 1 unadjusted analysis [123]. Diagnosis of DR TB via line probe assay, as compared to culture, was associated with higher risk of unfavorable treatment outcomes in 1 adjusted analysis [84] and 1 unadjusted analysis [123]; however, study authors hypothesized this finding may represent survivor bias due to the longer time to diagnosis and treatment initiation with culture. In adjusted analyses, resistance to >=5 drugs [88] and individual resistance to ofloxacin, streptomycin, or ethambutol [84] were associated with unfavorable treatment outcomes. Lack of treatment response was also associated with unfavorable outcomes, as represented by lack of radiographic improvement in 1 unadjusted analysis [119] and longer time to culture conversion in 1 adjusted [124] and 1 unadjusted analysis [119].

Comorbid conditions also increased risk of unfavorable outcomes. Undernutrition was associated with unfavorable outcomes using various measures, including body mass index <18 or 18.5 in 2 adjusted [84,116] and 1 unadjusted analysis [121], decreasing body mass index (continuous variable in) 1 adjusted analysis [118], weight <30 kilograms in 1 adjusted analysis [122], weight loss or lack of weight gain during treatment in 1 adjusted [84] and 1 unadjusted analysis [120], and decreasing serum albumin (continuous variable) in 1 adjusted analysis [121].

Psychosocial, behavioral, and substance use barriers also contributed to unfavorable outcomes. Substance use—alcohol use in 1 adjusted [125] and 1 unadjusted analysis [119] and smoking in 1 unadjusted analysis [119]—was associated with unfavorable outcomes. Medication nonadherence—defined as missing >=10% of all doses [121] or >=7 doses in the intensive or continuation treatment phase [84]—was associated with unfavorable outcomes in adjusted analyses. In 1 study, longer exposure to a psychosocial support package—involving motivational counseling, patient-provider group meetings, nutritional supplementation, and cash transfer—was associated with lower adjusted risk of unfavorable outcomes [118].

Health system-related factors were also associated with unfavorable outcomes. In 1 study, patients previously treated for TB through general government health services had higher adjusted risk of treatment failure when compared to those previously treated through special health services for central government employees (e.g., employees of India’s railway system) [84]. Patients with a different DOT provider in the intensive and continuation treatment phases had a higher adjusted risk of unfavorable outcomes when compared to those with the same DOT provider in both phases [124]. Ambulatory treatment initiation without initial hospitalization was associated with unfavorable treatment outcomes in 1 study [124]. In contrast, another study found hospitalization during treatment was associated with higher adjusted odds of unfavorable outcomes [125].

#### Reasons reported by RR or MDR TB patients for loss to follow-up or medication nonadherence during treatment

Patient-, family-, or society-related reasons for loss to follow-up or medication nonadherence among patients with RR or MDR TB included having a busy schedule (implying work-related barriers), substance use, and lack of family support [126] (Fig 18). Loss to follow-up due to migration to another location was reported by 93% of patients interviewed in 1 study [123]. Clinical barriers such as medication side effects, complexity (or composition) of the drug regimen, the daunting long duration treatment, lack of symptom improvement with treatment, and early relief of symptoms (resulting in loss of motivation to continue therapy) were seen across studies [123,126,127]. In 1 study, nearly three-quarters of patients who discontinued treatment did so due to medication adverse effects [126]. No health system-related barriers were reported, although this may be limited by the interview approach used in the included studies.

**Fig 18.**
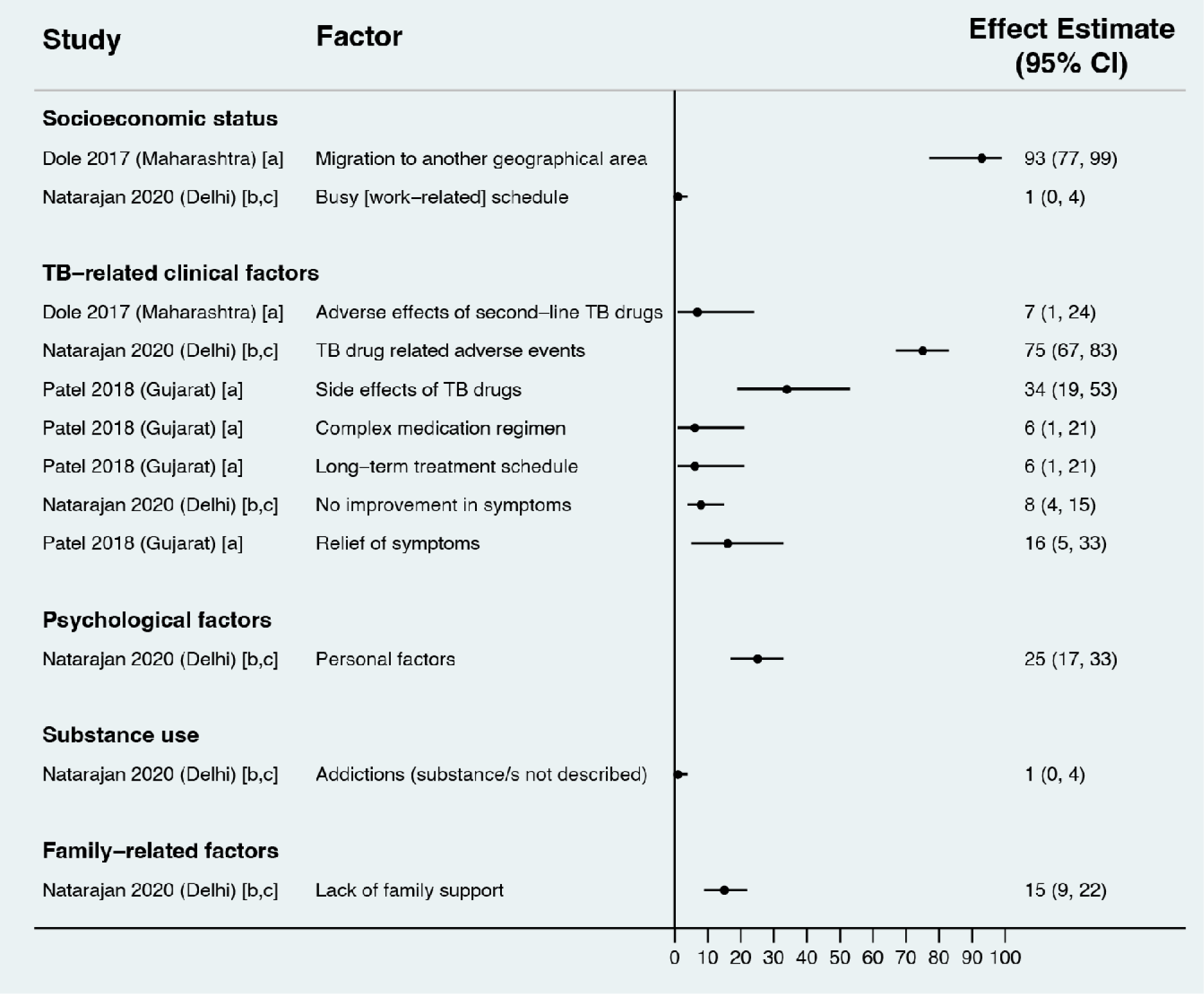
Reasons reported by rifampin-resistant or multidrug-resistant (MDR) TB patients for experiencing unfavorable outcomes during treatment in quantitative surveys (Gap 4). Estimates represent the percentage of individuals interviewed who reported a given reason for experiencing unfavorable outcomes. CI, confidence interval.

#### Factors associated with unfavorable TB treatment outcomes in studies including multiple populations of TB patients

Given numerous studies for this subgroup, we only describe findings from adjusted analyses; findings from unadjusted analyses are reported in Table D4 in S4 Appendix. With regard to patient-, family-, or society-related factors, male sex was associated with unfavorable outcomes in 8 adjusted analyses [43,44,86,128–132] (Fig 19). In 7 adjusted analyses, older age was associated with unfavorable outcomes across a range of age thresholds [86,89,91,129,130,133,134]. Living in a joint (vs nuclear) family [128] and being ever married (versus never having been married) [44] were associated with higher adjusted odds of unfavorable outcomes.

**Fig 19.**
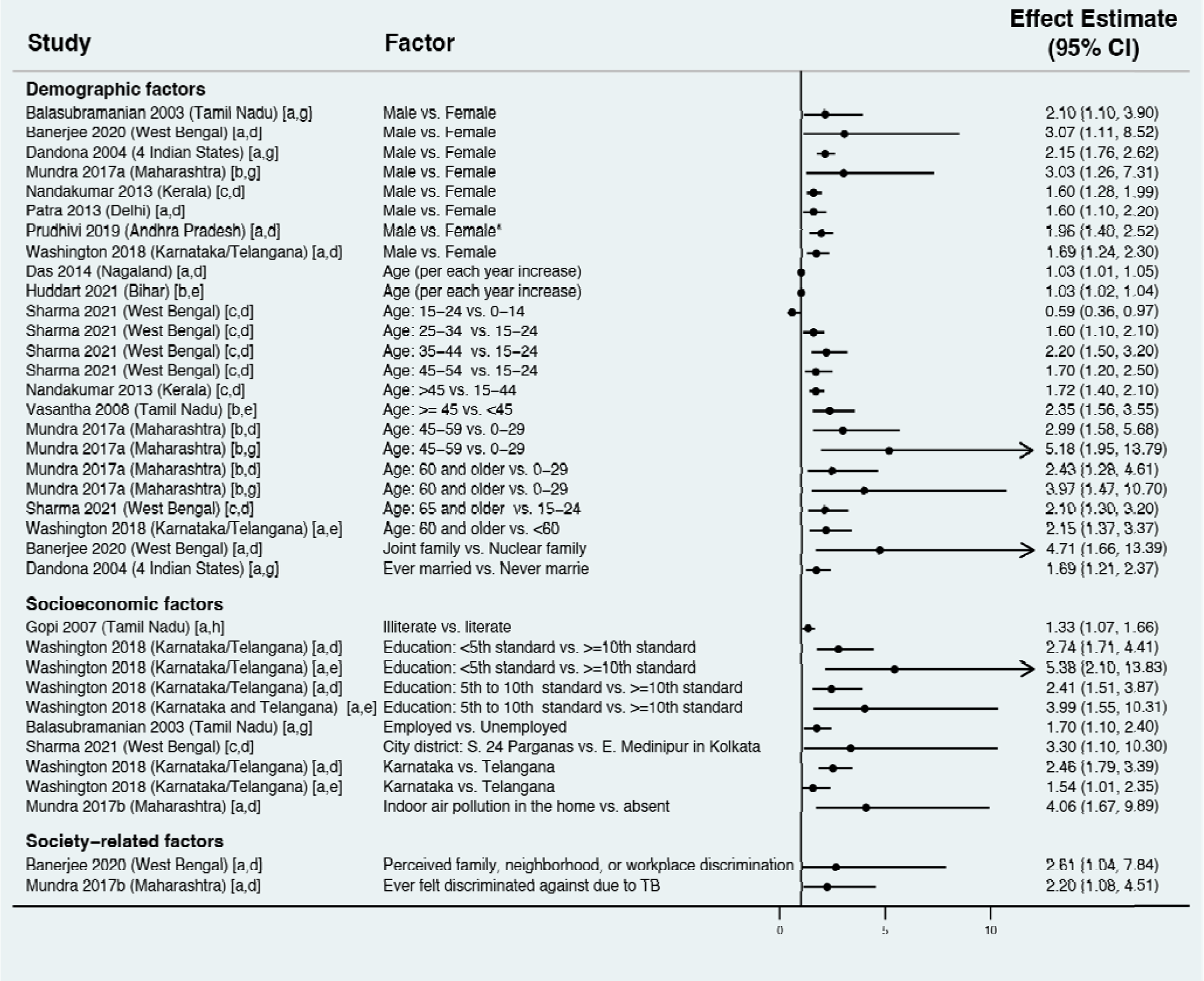
Patient-, family-, and society-related factors associated with unfavorable outcomes during TB treatment in studies including multiple populations of TB patients (Gap 4). Estimates greater than 1 represent increased adjusted risk of unfavorable outcomes; estimates less than 1 represent decreased adjusted risk of unfavorable outcomes CI, confidence interval; OR, odds ratio; TB, tuberculosis. [a] Effect estimate is an adjusted odds ratio; [b] effect estimate is an adjusted hazard ratio; [c] effect estimate is an adjusted relative risk ratio; [d] outcome is any unfavorable treatment outcome; [e] outcome is death; [f] outcome is treatment failure; [g] outcome is loss to follow-up; [h] outcome is medication nonadherence.

Socioeconomic barriers were also associated with unfavorable outcomes. Lower education was associated with unfavorable outcomes, whether measured as illiteracy [135] or educational attainment less than the 10^th^ standard [86]. Being employed (versus unemployed), usually a marker of higher socioeconomic status [43], was associated with higher adjusted risk of unfavorable outcomes, consistent with findings from a study of previously treated patients [110]. Indoor air pollution, which is a marker of lower socioeconomic status and an environmental TB risk factor, was associated with higher adjusted odds of unfavorable outcomes [129]. With regard to society-related factors, in 2 adjusted analyses, perceived discrimination due to TB was associated with unfavorable outcomes [128,129].

Clinical factors associated with increased adjusted odds of unfavorable outcomes included sputum smear positive pulmonary TB (versus extrapulmonary or smear negative pulmonary TB) in 5 studies [129,131,132,136,137], previous TB treatment history (versus new patients) in 9 studies [43,75,86,105,128,130,132,134,137], and drug resistance in 1 study [86] (Fig 20). Undernutrition—defined as weight <35kg [134] or baseline weight less than the median for the cohort [86]—was associated with increased adjusted risk of unfavorable outcomes. People with HIV, or with unknown HIV status, had higher adjusted risk of unfavorable outcomes, as compared to people without HIV in 3 studies [86,130,132]. Unknown diabetes status (versus not having diabetes) was also associated with unfavorable outcomes in 1 adjusted analysis [130]. Alcohol use was associated with unfavorable outcomes in 5 adjusted analyses [43,86,104,132,134], as was any history of smoking in 1 adjusted analysis [132].

**Fig 20.**
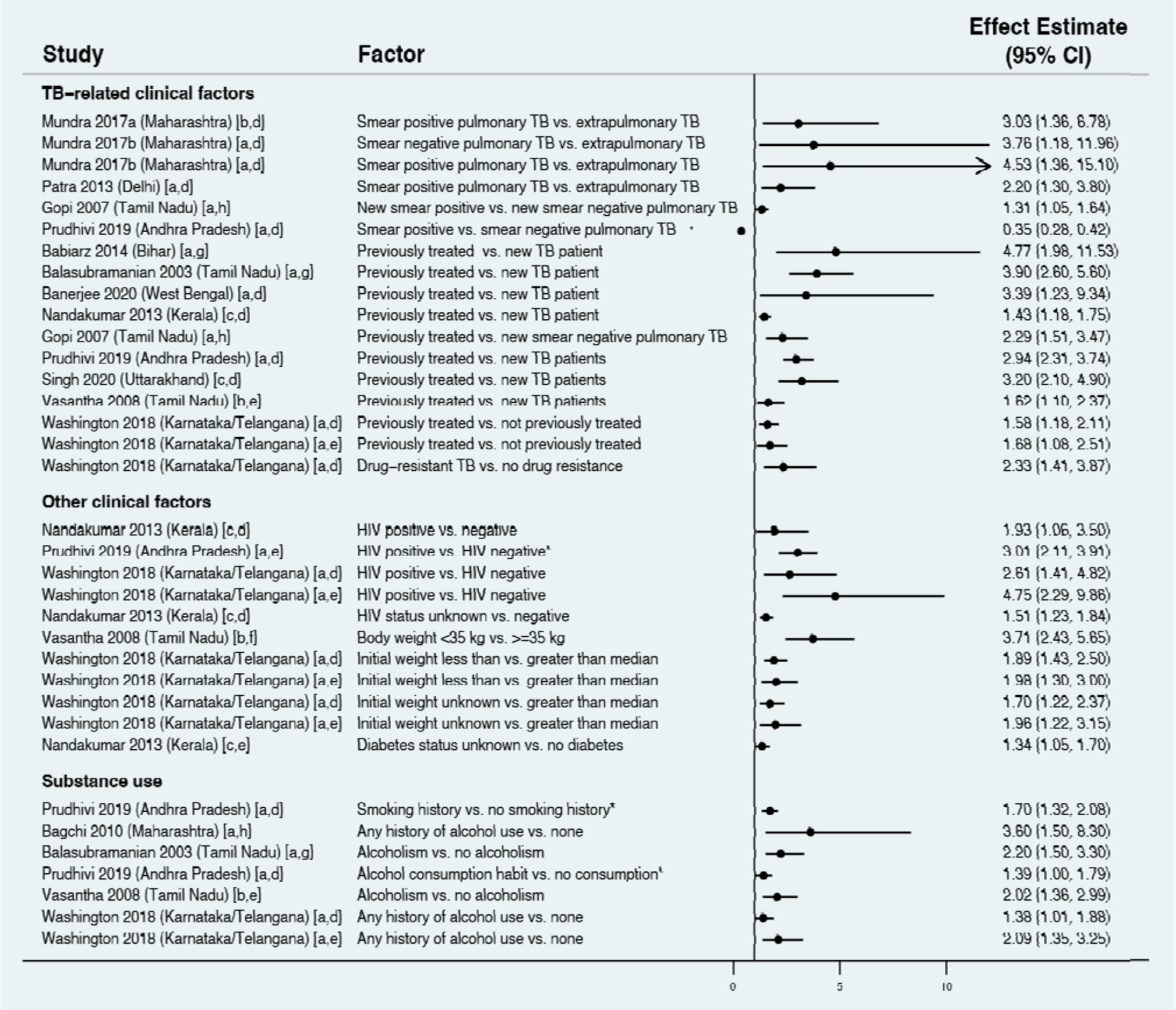
Clinical factors associated with unfavorable outcomes during TB treatment in studies including multiple populations of TB patients (Gap 4). Estimates greater than 1 represent increased adjusted odds of unfavorable outcomes; estimates less than 1 represent decreased adjusted odds of unfavorable outcomes CI, confidence interval; OR, odds ratio; TB, tuberculosis. [a] Effect estimate is an adjusted odds ratio; [b] effect estimate is an adjusted hazard ratio; [c] effect estimate is an adjusted relative risk ratio; [d] outcome is any unfavorable treatment outcome; [e] outcome is death; [f] outcome is treatment failure; [g] outcome is loss to follow-up; [h] outcome is medication nonadherence.

With regard to health system-related factors, negative patient user experience—specifically, feeling lower levels of satisfaction with TB services or DOTS providers—was associated with unfavorable outcomes in 2 adjusted analyses [44,129] (Fig 21). Barrier to healthcare access also contributed to unfavorable outcomes [137], including having to spend on medications or travel to health facilities [105]. Approaches to care provision influenced outcomes. In 1 adjusted analysis, patients diagnosed by active case finding (versus passive case finding) had a higher risk of unfavorable outcomes [75]. In 2 adjusted analyses, patients with a government health facility DOT provider had better outcomes when compared to supervision by an Anganwadi worker, non-governmental organization, or self-administered therapy [44,137]. Suboptimal quality of care also contributed to unfavorable outcomes, including patients not being told that TB is curable or treatment duration [44] and missed treatment due to lack of drug availability [104].

**Fig 21.**
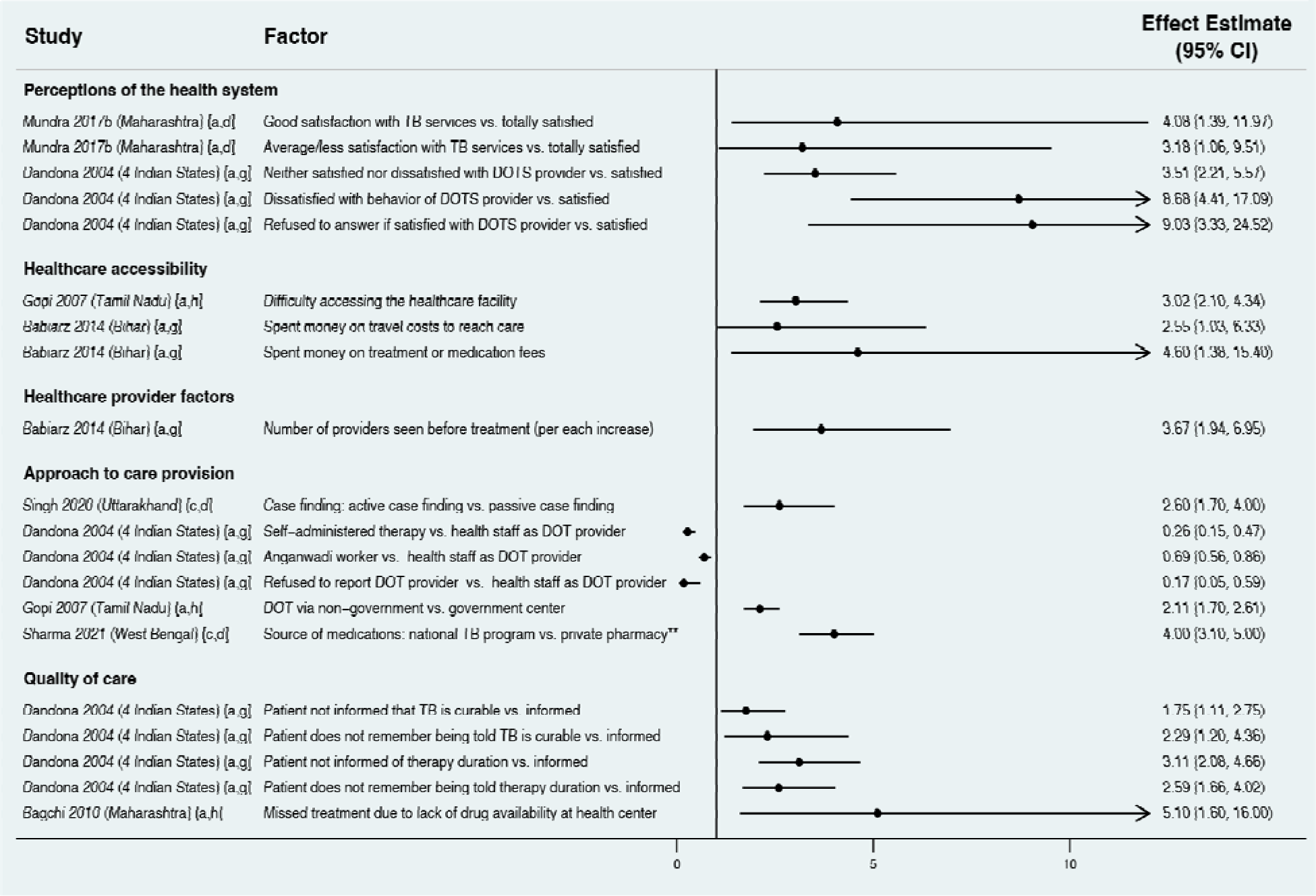
Health system-related factors associated with unfavorable treatment outcomes in studies including multiple populations of TB patients (Gap 4). Estimates greater than 1 represent increased adjusted risk of unfavorable outcomes; estimates less than 1 represent decreased adjusted risk of unfavorable outcomes CI, confidence interval; OR, odds ratio; TB, tuberculosis. [a] Effect estimate is an adjusted odds ratio; [b] effect estimate is an adjusted hazard ratio; [c] effect estimate is an adjusted relative risk ratio; [d] outcome is any unfavorable treatment outcome; [e] outcome is death; [f] outcome is treatment failure; [g] outcome is loss to follow-up; [h] outcome is medication nonadherence.

#### Reasons for loss to follow-up or medication nonadherence during TB treatment in studies including multiple populations of TB patients

With regard to patient-related reasons, socioeconomic barriers contributing to loss to follow-up or medication nonadherence included illiteracy [138] and work-related barriers in 3 studies [139–141] (Fig 22). Patient mobility, represented by migration [139,140,142] or change of address [140], was a commonly reported barrier. Clinical factors included medication side effects in 6 studies [138–143], lack of symptom improvement [141], and early symptom improvement in 5 studies [139–143]. Nearly half of patients were lost to follow-up due to early symptom improvement in 2 studies [140,143].

**Fig 22.**
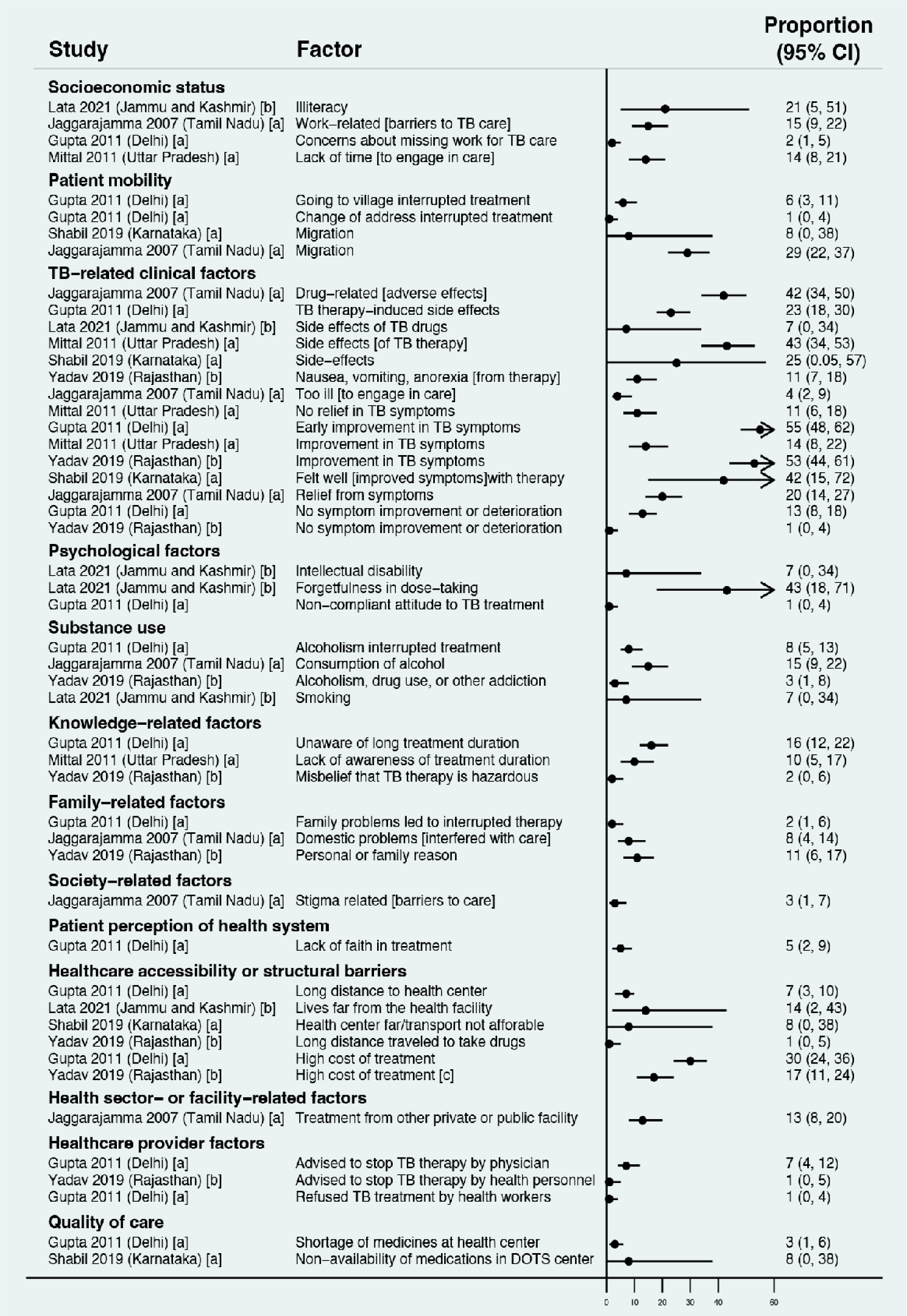
Reasons reported for experiencing unfavorable TB treatment outcomes in quantitative surveys including multiple TB patient populations (Gap 4). Estimates represent the percent of individuals interviewed who reported a given reason for experiencing unfavorable outcomes. CI, confidence interval; TB, tuberculosis.

Other patient-related reasons included psychological barriers such as forgetfulness in dose-taking [138] or non-compliant attitude to treatment [140]; substance use disorders such as alcoholism [139,140] and smoking [138]; knowledge gaps such as lack of awareness of the treatment duration [140,141] or misbelief that TB therapy is hazardous [143]; family or personal problems [139,140,143]; and social stigma [139].

Health-system related reasons included lack of faith in treatment [140]; healthcare accessibility challenges such as long distance to the health center [138,140,142,143] and high treatment costs [140,143]; healthcare provider barriers such as healthcare workers advising patients to stop treatment [140,143] or refusing to give treatment [140]; and shortage or non-availability of medications at the health center [140,142].

#### Factors associated with unfavorable TB treatment outcomes among people with HIV

For studies of people with HIV being treated for active TB, we organized findings into TB-related factors and HIV-related factors associated with unfavorable outcomes across adjusted (Fig 23) and unadjusted analyses (Table D5 in S4 Appendix). TB-related factors included pulmonary (versus extrapulmonary) disease in 2 adjusted analyses [144], irregular treatment (i.e., medication nonadherence) in 1 unadjusted analysis [144], and previous TB treatment history (vs. new TB) in 2 adjusted analyses [144,145]. HIV-related factors included CD4 cell count <=200 in 1 adjusted analysis [145], not being on antiretroviral therapy in 2 adjusted [144] and 1 unadjusted analyses [146], and not initiating cotrimoxazole prophylaxis in 1 adjusted analysis [144].

**Fig 23.**
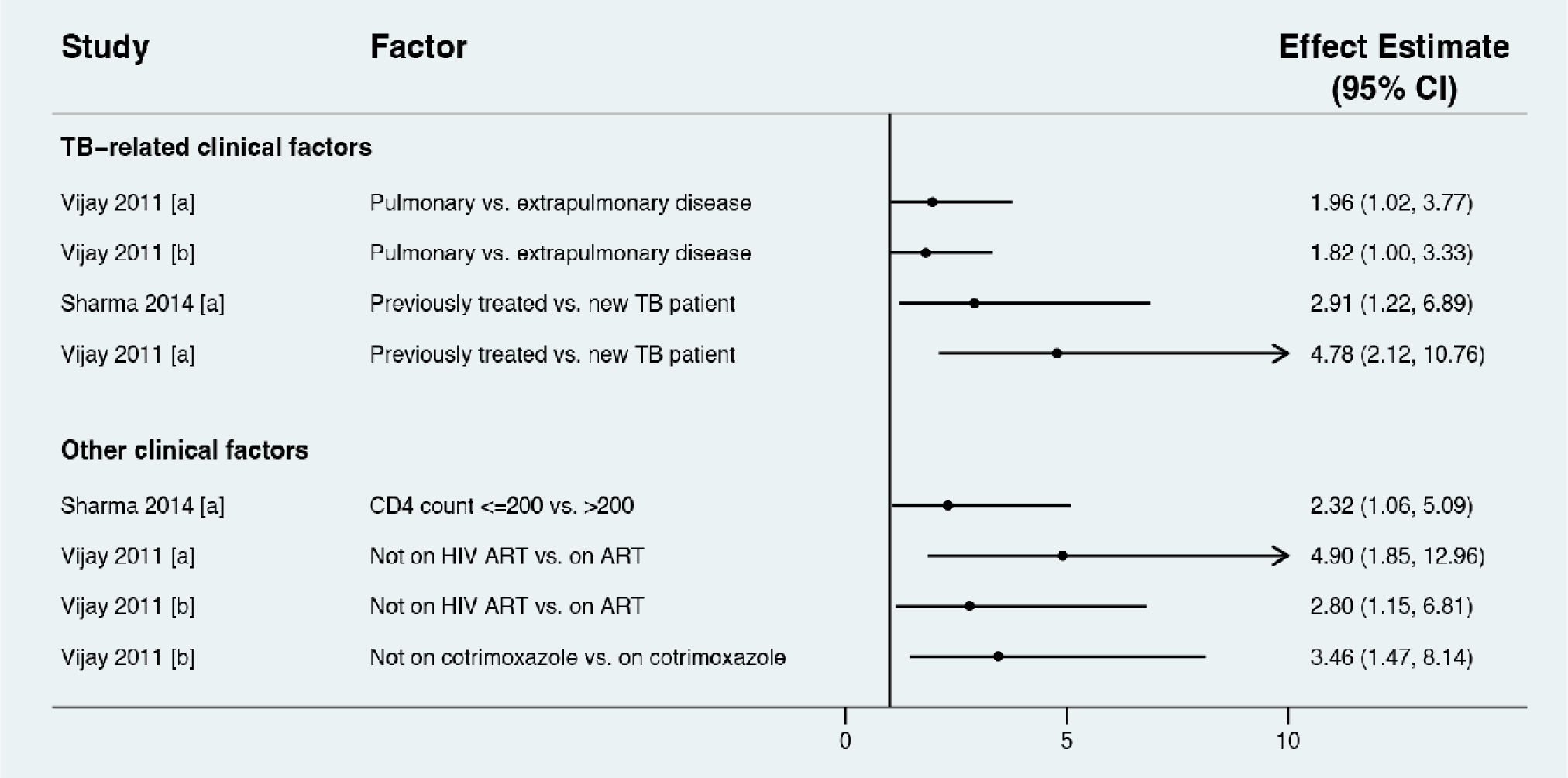
Factors associated with unfavorable outcomes during TB treatment among people with HIV (Gap 4). Estimates greater than 1 represent increased adjusted odds of unfavorable outcomes; estimates less than 1 represent decreased adjusted odds of unfavorable outcomes CI, confidence interval; OR, odds ratio; TB, tuberculosis.

#### Factors associated with unfavorable outcomes during TB treatment among pediatric TB patients

Studies of pediatric TB patients reported few findings from adjusted analyses. Hence, no Forest plot was created for this subpopulation, but unadjusted and adjusted effect estimates are reported in Table D6 in S4 Appendix. With regard to patient-related factors, in 1 unadjusted analysis [147], adolescents (10-19 years of age) had increased risk of unfavorable treatment outcomes compared to children (<10 years of age); however, the effect was non-significant in adjusted analysis. In another study, infants and toddlers (0-4 years of age) had higher unadjusted odds of unfavorable outcomes compared to older children (10-14 years of age) [65]. TB clinical factors associated with unfavorable outcomes included extensively drug resistant tuberculosis (among children with less advanced DR TB) in 1 adjusted analysis [147], pulmonary (vs. extrapulmonary) disease in 1 unadjusted analysis [148], previously TB treatment history (vs. new TB) in 1 unadjusted analysis [148], and presence of TB contact history in 1 unadjusted analysis [149]. Undernourished children had higher risk of unfavorable outcomes in 1 adjusted analysis [147].

### Gap 5—Barriers to achieving recurrence-free survival after TB treatment

#### Characteristics and quality of the included studies

Across searches spanning January 1, 2000, to May 17, 2021, we screened titles and abstracts of 3,799 unique reports and identified 79 for full text review, of which 15 met inclusion criteria (Fig A in S5 Appendix). Of these, 4 studies reported findings on TB recurrence as a single outcome or part of a composite outcome; 4 studies reported findings on post-treatment mortality as a single outcome or part of a composite outcome with on-treatment mortality; and 7 studies reported findings on both recurrence and mortality (Table C in S5 Appendix). Studies were conducted in 8 of India’s states and union territories, including Bihar and Madhya Pradesh, 2 of India’s high-population and low-income states. 3 studies were conducted in multiple states. 3 studies were conducted in rural areas, 7 in urban areas, and 5 in both (Table C in S5 Appendix).

All studies involved a high-quality random or comprehensive sampling (Table C in S5 Appendix). 4 studies were medium quality with regard to sample size and distribution, because they were conducted at a single facility [103,150–152]. 5 studies did not follow patients prospectively and relied on a lower-quality passive surveillance for assessing post-treatment outcomes [89,150,153–155], while 2 studies did not report the surveillance approach [156,157]. For studies evaluating TB recurrence, 3 studies had at least 1 post-treatment follow-up visit with microbiological testing to identify recurrence [80,150,158] (high quality), while 6 studies performed microbiological testing only for patients with persistent symptoms [85,96,103,152,153,159] (medium quality). 1 study evaluated TB recurrence through self-enrollment of patients in treatment [89] (low quality), and 1 did not clearly report the approach to detecting recurrence [151] (low quality).

All studies evaluated adult patients. 4 studies included adolescents (usually >14 years of age) [150,151,156,157] and 1 study included younger children [153]. Except for 1 study of the private sector [89], studies evaluated patients in the public sector. (Table C in S5 Appendix). We present findings separately for studies evaluating TB recurrence (as a single outcome or part of a composite outcome) and post-treatment mortality (as a single outcome or part of a composite outcome along with on-treatment mortality).

#### Factors associated with post-treatment TB recurrence

We identified common findings on factors associated with TB recurrence across adjusted (Fig 24) and unadjusted analyses (Table D in S5 Appendix). In 1 adjusted analysis [80] and 2 unadjusted analyses [96,153], male sex was associated with TB recurrence. Suboptimal medication adherence was associated with TB recurrence in 1 adjusted [158] and 1 unadjusted analysis [80], though this association was nonsignificant after adjusting for male sex in the latter study. Ongoing symptoms after treatment—measured by clinical evaluation or the structured Saint George’s Respiratory Questionnaire—was associated with TB recurrence in 2 adjusted analyses [150,159]. Low body mass index at TB diagnosis was associated with TB recurrence in 1 adjusted [96] and 1 unadjusted analysis [80] as a single risk factor, and in combination with severe alcohol use disorder, measured using the alcohol use disorder identification test C (AUDIT-C), in 1 adjusted analysis [85]. Alcohol use was associated with TB recurrence in 2 adjusted [85,96] and 2 unadjusted analyses [80,158]. Patients with current or past smoking had higher TB recurrence risk in 2 adjusted [151,158] and 1 unadjusted analysis [80].

**Fig 24.**
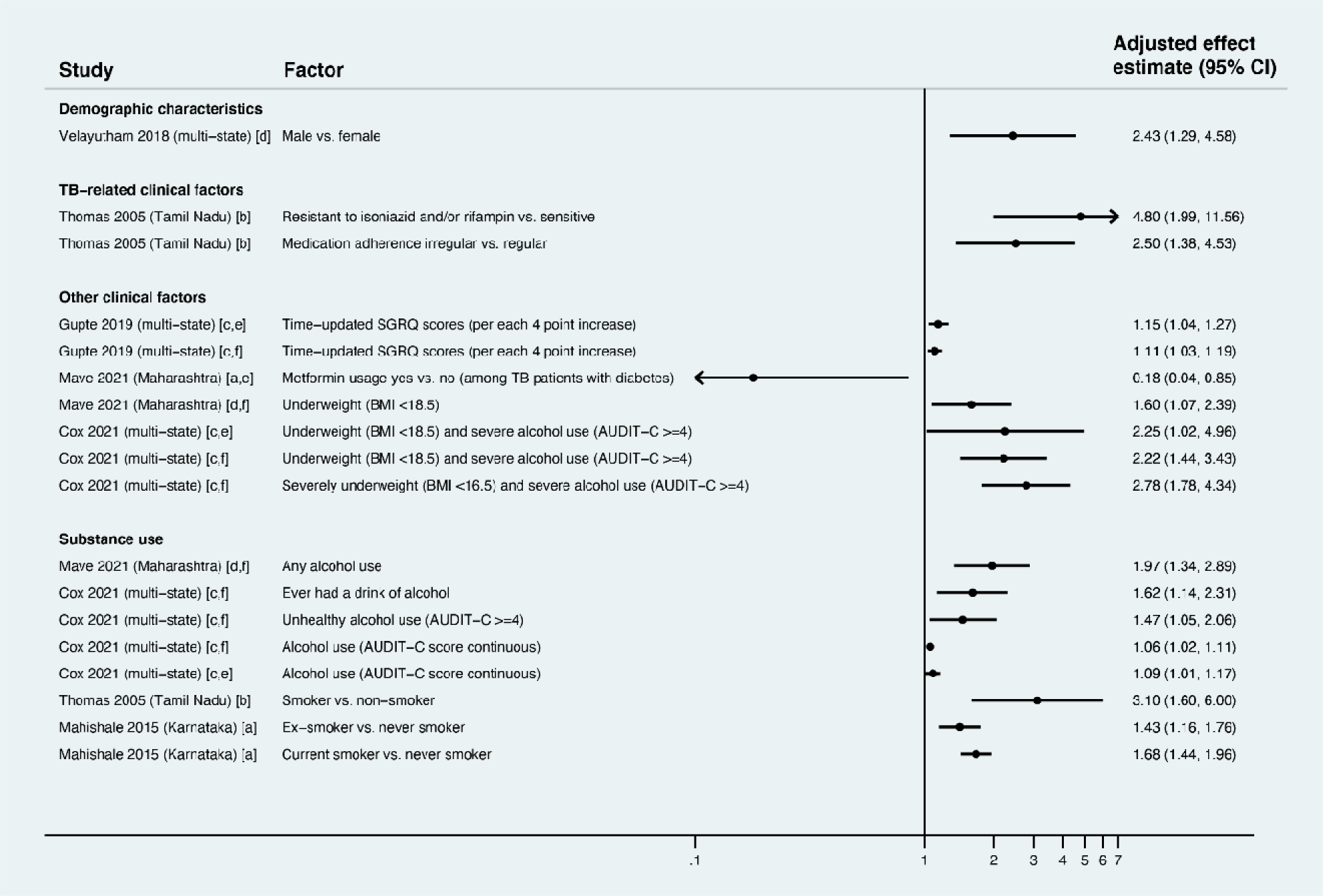
Factors associated with TB recurrence as a single outcome or part of a composite outcome after completion of TB treatment (Gap 5). Only statistically significant findings are presented. Effect estimates greater than 1 represent increased risk of TB recurrence; estimates less than 1 represent decreased risk of TB recurrence. All studies used multivariable regression with findings reported as: [a] adjusted hazard ratios, [b] adjusted odds ratios, [c] adjusted incidence rate ratios, or [d] adjusted relative risk ratios. Studies labeled [e] reported post-treatment TB recurrence as a single outcome, while studies labelled [f] report post-treatment recurrence as part of a composite outcome including on-treatment and post-treatment death and treatment failure. AUDIT, alcohol use disorder identification test; BMI, body mass index; SGRQ, Saint George Respiratory Questionnaire; TB, tuberculosis.

#### Factors associated with mortality after TB treatment

We identified common factors associated with post-treatment mortality across adjusted (Fig 25) and unadjusted analyses (Table D in S5 Appendix). In 4 adjusted [89,154,156,157] and 1 unadjusted analyses [155], older age—people older than 25, 40, 44, or 60 years depending on the study—was associated with post-treatment mortality. In 3 adjusted analyses [154,156,157], unfavorable on-treatment outcomes—including loss to follow-up and treatment failure—were associated with post-treatment mortality. In 1 study that included 2 adjusted analyses [159], higher scores on the Saint George’s Respiratory Questionnaire at TB diagnosis, or updated throughout treatment, were associated with post-treatment mortality, as was dyspnea at TB diagnosis in another unadjusted analysis [150].

**Fig 25.**
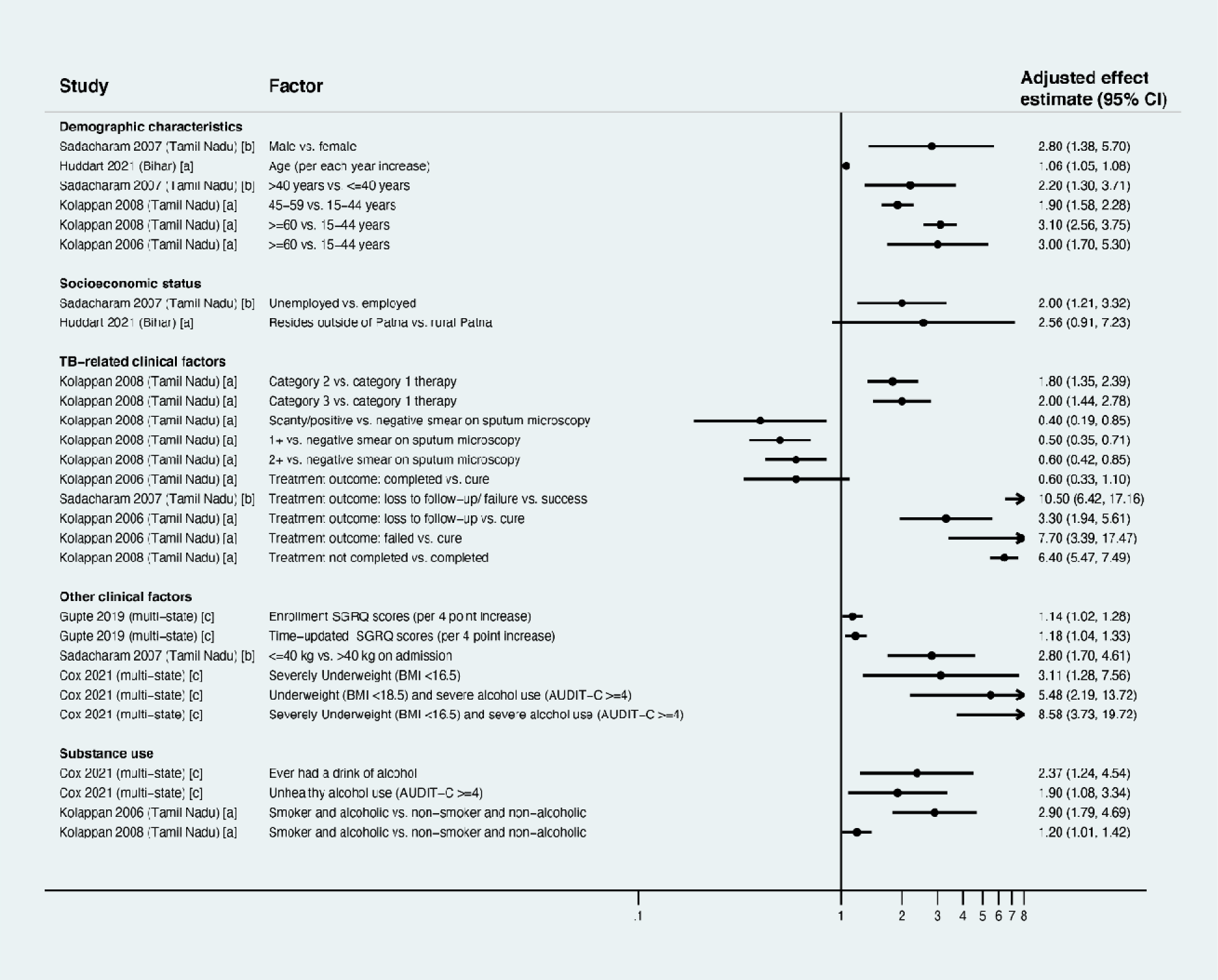
Factors associated with post-treatment tuberculosis mortality (without evaluation of recurrence) (Gap 5). Only statistically significant findings are presented. Effect estimates greater than 1 represent increased risk of post-treatment mortality; estimates less than 1 represent decreased risk of post-treatment mortality. All studies used multivariable regression with findings reported as: [a] adjusted hazard ratios, [b] adjusted odds ratios, and [c] adjusted incidence rate ratios. Studies labeled [d] reported post-treatment mortality as a single outcome, while studies labelled [e] report post-treatment mortality as part of a composite outcome including on-treatment mortality. AUDIT, alcohol use disorder identification test; BMI, body mass index; SGRQ, Saint George Respiratory Questionnaire.

Low body mass index was associated with post-treatment mortality in 2 adjusted analyses as a single factor [85,154], and in combination with severe alcohol use disorder (measured by the AUDIT-C) in 1 adjusted analysis [85]. Alcohol use was associated with post-treatment mortality as an single factor in 1 adjusted [85] and 1 unadjusted analysis [154], and in combination with smoking in 2 adjusted analysis [156,157]. Smoking was associated with post-treatment mortality in 1 adjusted [150] and 1 unadjusted analysis [154]. HIV was associated with post-treatment mortality in 2 unadjusted analyses [103,152].

## Discussion

In this systematic review, we synthesized 2 decades of studies that provide rich insights into barriers to care for people with TB across care cascade stages in India [5,6]. In conducting such an extensive review, we believe we have made several important contributions to knowledge on TB care delivery in India. First, given our disaggregation of findings by care cascade gap and subpopulation, we provide a roadmap for program managers and researchers, who can engage with our results in a targeted manner to understand barriers that may inform intervention development for specific care gaps or specific regions, states, or cities in India [14].

Second, for researchers, our review sheds light on shortcomings of quantitative research conducted to date. Of particular concern across all care cascade gaps is the dearth of studies of children and private sector services, where at least half of people with TB in India receive care [160,161]. In addition, most studies captured data on patient-related factors. Fewer studies captured data on family-, society-, or health system-related factors. In future studies, health system-related factors may be particularly important to measure, as they may directly inform changes in programmatic care delivery. In addition, among patient-related factors, some comorbid conditions that could be intervened upon, such as alcohol use, were frequently measured, while others, such as depression or TB stigma, were measured less frequently, if at all. Understanding the influence of comorbid conditions should be focus of future research to facilitate better integration of TB care with care for other conditions.

Third, we organized and visualized results in a manner that clusters similar factors across studies, bringing into focus common patterns of risk that may affect TB outcomes. Programs may use these findings to prioritize subpopulations at risk for poor outcomes through provision of greater attention or resources. These findings may also inform development of interventions that reduce unfavorable TB outcomes. For the rest of this discussion, we consider in detail common patterns of risk—across and within care cascade gaps—that emerged in this review.

### Continuities in risk across care cascade stages

Our review shows some subpopulations of people with TB have elevated risk of unfavorable outcomes across multiple care cascade stages (Tables 3 and 4). For example, in some studies, men in the community were less likely to seek care for symptoms (Gap 1) [37,59], and less likely to pursue TB evaluation after being referred (Gap 2) [40,41]. Notably, a recent study, which used standardized patients to evaluate private providers, found men and women had similarly low likelihood of receiving appropriate evaluation for TB symptoms, but men had interactions with less provider time, less detailed explanations, and lower satisfaction [162]. Collectively, these findings partly explain the Indian and global phenomenon of “missing men” in TB care [163], in which men are relatively under-represented in case notifications from TB programs [164] even though they have considerably higher TB prevalence in the population [165,166]. After reaching care, men are more likely to suffer unfavorable on-treatment outcomes (Gap 4) [43,44,83,84,86,116–118,129–132,167,168] and post-treatment TB recurrence and death (Gap 5) [80,154]. TB services at every care stage—from active case-finding to post-treatment monitoring—need to incorporate strategies to engage and retain vulnerable men.

**Table 3.** Characteristics and quality of the included studies for each TB care cascade gap investigated in this systematic review.

| Study characteristic | Gap 1 | Gap 2 | Gap 3 | Gap 4 | Gap 5 |
| --- | --- | --- | --- | --- | --- |
| **Total studies or analyses included in each gap** | 15<sup>a</sup> | 20<sup>b</sup> | 25<sup>e</sup> | 104<sup>f</sup> | 15<sup>g</sup> |
| **Type of study design** |  |  |  |  |  |
| Factors associated with unfavorable outcome | 13 | 15<sup>c</sup> | 18 | 97 | 15 |
| Reasons reported by people for unfavorable outcome | 9 | 10<sup>d</sup> | 11 | 14 | 0 |
| **Study setting** |  |  |  |  |  |
| States or union territories covered by included studies | 7 | 11 | 11 | 24 | 8 |
| Rural setting | 5 | 6 | 6 | 24 | 3 |
| Urban setting | 8 | 7 | 9 | 44 | 7 |
| Both rural and urban settings | 3 | 7 | 10 | 29 | 5 |
| **Quality or risk of bias (number of studies with low quality)** |  |  |  |  |  |
| Sampling strategy | 0 | 0 | 0 | 0 | 0 |
| Sample size | 1 | 2 | 8 | 20 | 0 |
| Proportion of estimated population screened for TB | 10 | NA | NA | NA | NA |
| Proportion of interviewed individuals identified as having TB symptoms | 1 | NA | NA | NA | NA |
| Time frame of research fieldwork after start of diagnostic evaluation | NA | 11 | NA | NA | NA |
| Method of evaluating outcome (self-report by the government TB program) | NA | 3 | 7 | NA | NA |
| Time frame of research fieldwork after diagnosis | NA | NA | 16 | NA | NA |
| Assessment of exposures and outcomes (retrospective review) | NA | NA | NA | 47 | NA |
| Surveillance for post-treatment recurrence or mortality (passive) | NA | NA | NA | NA | 7 |
| Method for diagnosing recurrence (clinical diagnosis without microbiology) | NA | NA | NA | NA | 2 |

<sup>a</sup>Of 15 studies total, one study reported data from two geographic locations [37], so we present characteristics across 16 locations.
<sup>b</sup>Of 19 studies total, one study reported two different outcomes [50], so we present characteristics across 20 analyses.
<sup>c</sup>Of these 15 studies, 3 reported on non-pursual of diagnostic workup despite referral [40–42], 4 reported on non-completion of sputum microscopy evaluation [43–46], 2 reported on chest X-ray non-completion [47,48], and 6 reported on non-completion of NAAT, line probe assay, or mycobacterial culture [47,49–53].
<sup>d</sup>Of these 10 studies, 2 reported on non-completion of sputum microscopy evaluation from patient interviews [44,54], 2 reported on chest X-ray non-completion from patient interviews [55,56], and 6 reported on NAAT non-completion from evaluation of health records [49–52,57,58].
<sup>e</sup>18 studies evaluated PTLFU in adults with confirmed or presumed drug-susceptible TB, 6 studies evaluated adults with drug-resistant TB, 1 study evaluated children with drug-susceptible or drug-resistant TB.
<sup>f</sup>We present findings separately for patient subpopulations: those with new confirmed or presumed drug-susceptible TB (n=25), those with a previous TB treatment history being treated for confirmed or presumed drug-susceptible TB (n=17), those involving multiple patient subpopulations—i.e., more than one of the aforementioned subpopulations without disaggregation (n=46), those with RR or MDR TB (n=16), those with HIV being treated for TB (n=4), and children with TB (n=3).
<sup>g</sup>4 studies reported findings on TB recurrence as a single outcome or part of a composite outcome, 4 studies reported finding on post-treatment mortality as a single outcome or part of a composite outcome with on-treatment mortality, and 7 studies reported findings on both recurrence and mortality.
TB, tuberculosis; NA, not applicable; NAAT, nucleic acid amplification test; PTLFU, pre-treatment loss to follow-up; RR, rifampin-resistant; MDR, multidrug-resistant; HIV, Human immunodeficiency virus

**Table 4.**
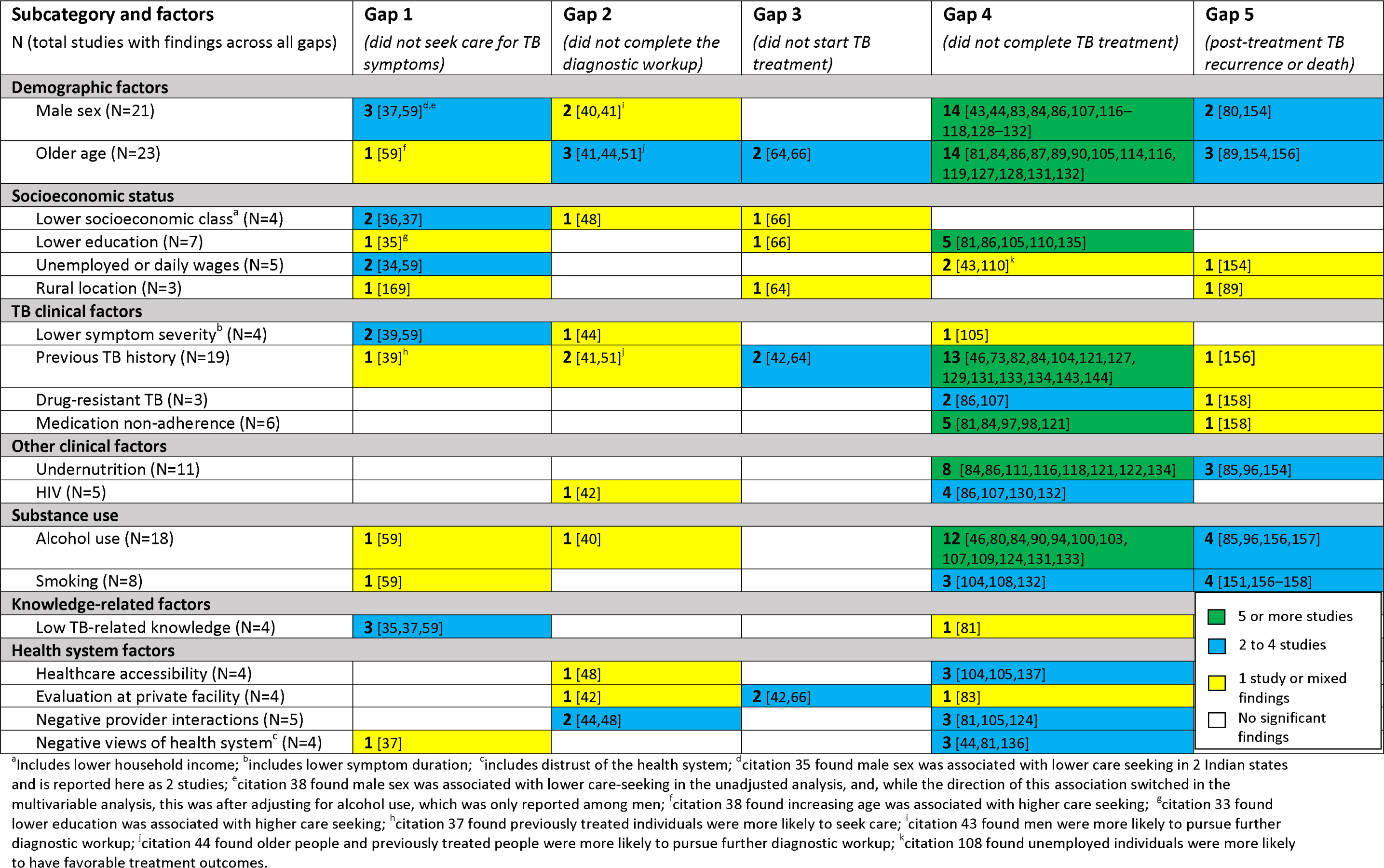
Factors statistically significantly associated with unfavorable outcomes across multiple TB care cascade gaps in multivariable regression analyses. Numbers indicate the number of studies contributing data to each factor for a given gap. Footnotes identify discordant (protective) findings for some factors.

**Table 5.** Reasons reported by people with TB or TB symptoms for not seeking care or being lost from care across multiple care cascade gaps. Numbers indicate the number of studies contributing data to each reason for a given gap. Only findings in which 20% or more of people reported a given reason are presented.

| **Subcategory and reasons**<br>N (total studies with findings across all gaps) | **Gap 1**<br>*(did not seek care for TB symptoms)* | **Gap 2**<br>*(did not complete the diagnostic workup)* | **Gap 3**<br>*(did not start TB treatment)* | **Gap 4**<br>*(did not complete TB treatment)* |
| --- | --- | --- | --- | --- |
| **Socioeconomic status** |  |  |  |  |
| Work constraints or pressure (N=8) | 4 [37,61–63] | 1 [56] | 1 [68] | 2 [99,108] |
| Patient mobility (N=6) |  |  | 1 [68] | 5 [80,93,98] |
| **TB clinical factors** |  |  |  |  |
| Symptoms not severe or resolved early (N=17) | 9 [33,37,38,60–63,170]<sup>a</sup> | 2 [44,55] |  | 6 [80,108,139,140,142,143] |
| Severity of TB illness<sup>b</sup> (N=5) |  | 1 [54] | 4 [57,64,67,77] |  |
| Adverse effects of TB therapy<sup>c</sup> (N=9) |  |  | 1 [72] | 8 [99,108,126,127,139–142] |
| **Psychological factors** |  |  |  |  |
| Stigma related to TB (N=3) |  |  | 2 [44,76] | 1 [108] |
| **Substance use** |  |  |  |  |
| Alcohol use (N=2) |  |  | 1 [76] | 1 [80] |
| **Health system factors** |  |  |  |  |
| Long distance to health facility (N=3) | 2 [38,61] |  | 1 [73] |  |
| Negative views of health system (N=3) |  |  | 2 [44,72] | 1 [108] |
| Negative health provider interactions (N=2) |  | 1 [44] |  | 1 [99] |
| Quality of care: Patient not told or referred for next steps in care or patient untrackable because of poor recording of contact information (N=9) |  | 4 [51,52,56,58] | 5 [57,64,67,74,77] |  |

<sup>a</sup>citation 35 (George et al.) found 'symptoms not severe' as one of the key reasons given by chest symptomatics for not seeking care in two Indian states and is reported here as two studies
<sup>b</sup>includes disease-related barriers and early deaths; <sup>c</sup>includes experiences of adverse effects of prior TB therapy as contributing to pretreatment loss to follow-up.

Other characteristics of people with TB also played a role across multiple gaps. Although variable age categories were used across studies, in general, older individuals—usually those older than 40 compared to younger age categories—were more likely to suffer adverse TB outcomes across multiple gaps. For people previously treated for TB, outcomes varied across care cascade gaps. On the one hand, previously treated individuals were more likely to seek care for TB symptoms (Gap 1) [39] and pursue TB evaluation when referred (Gap 2) [41], which may reflect better TB knowledge. On the other hand, concerningly, these patients were more likely to experience PTLFU (Gap 3) [42,64] and unfavorable treatment outcomes (Gap 4) [43,75,86,105,130,132,134,137,144,145,168], with people who were lost to follow-up during prior treatment experiencing particularly poor outcomes [83,105,111]. These poor outcomes later in the care cascade may reflect underlying drug resistance, chronic pulmonary disease from the prior TB episode, or continuation of behavioral risks that led to unfavorable outcomes during previous treatment. Previously treated patients should be a central focus of the NTEP’s efforts to reduce PTLFU and poor on-treatment outcomes.

TB is a highly socially stratified disease, with Indians in the lowest wealth quintile experiencing a TB prevalence that is fivefold greater than in the highest quintile [171]. Our review further shows that lower socioeconomic status is associated with higher risk of unfavorable outcomes across most care cascade gaps, whether assessed using household income, educational attainment, or job type. Reasons reported by people with TB for unfavorable outcomes help unpack *how* lower socioeconomic status shapes outcomes. For example, findings from multiple gaps suggest the association between lower educational attainment and unfavorable outcomes may partly relate to inadequate TB knowledge [35,37,76,81,140,141,143]. Work constraints were frequently reported as a reason for non-engagement in care [37,38,54,56,60–63,68,76,80,93,99,108,126,139,140], which aligns with findings that people in certain jobs, such as daily wage laborers (informal sector workers who are not paid if they miss work), may be particularly vulnerable to unfavorable outcomes [34,59]. A recent study conducted in Chennai, Vellore, and Mumbai published after the search period for this review similarly found that daily wage laborers have increased adjusted odds of TB medication nonadherence, measured using urine isoniazid testing [29]. Similarly, challenges related to patient mobility [68,77,98,123,139,140,142] align with findings that people seeking care outside of the location they reside [64,172] or migrant laborers (who work in both rural and urban areas during the year) may be more vulnerable to unfavorable outcomes [68,93]. Structural barriers to reaching clinics—including prohibitive distance [29,38,44,48,60–62,64,73,89,99,137,138,140,142,143], costs [29,33,37,59–62,76,104,105,142,170,173], or transportation [29,33,37,38,60,62]—were another pathway by which socioeconomic status was associated with unfavorable outcomes. While the NTEP has attempted to address poverty through direct benefits transfer (DBT; a form of unconditional cash transfer) to TB patients [174], our findings collectively suggest that developing a broader array of strategies to ameliorate the complex impact of extreme poverty on care engagement should be central to achieving the END TB/Sustainable Development Goal targets for India.

India’s 2019-2021 national TB prevalence survey found that about half of people with active TB in the population had no symptoms (or minimal symptoms that did not meet criteria for presumptive TB), and, among those with symptoms, about two-thirds had not sought care, potentially due to lack of symptom severity [166]. Our Gap 1 results evaluating studies of people with TB symptoms in the community affirm these findings from the national prevalence survey. People with shorter symptom duration or fewer symptoms were less likely to have sought care [39,59], and individuals who had not sought care frequently reported low symptom severity or resolving symptoms as reasons [33,37,38,60–63,170]. People with lower symptom duration or severity also had greater risk of being lost across subsequent care cascade stages. In Gap 2, they are less likely to pursue diagnostic workup when referred [40,41] and less likely to complete sputum evaluation [44]. Disappearance of symptoms was a common reason for chest X-ray non-completion [55]. In Gap 4, patients report that mild symptoms at treatment initiation or early symptom resolution with treatment are common reasons for loss to follow-up [80,99,105,108,127,139–143]. These results align with findings suggesting patients diagnosed by active case-finding—which identifies people with TB at a less symptomatic disease stage—were more likely to experience pretreatment and on-treatment loss to follow-up (Gaps 3 and 4) [74,75]. By identifying people with TB early in the disease course, active case-finding has potential for substantial benefits for the individual (averting morbidity and mortality) and public health (preventing transmission); however, our review highlights critical challenges in retaining these individuals in care. Active case-finding strategies should routinely measure care cascade outcomes and consider novel strategies—such as counseling or incentives at each care stage—to ensure early case detection translates into optimal outcomes.

Three conditions associated with higher TB prevalence in India’s national survey—alcohol use, smoking, and undernutrition—were also associated with greater risk of poor outcomes across multiple care cascade stages. Alcohol use was associated with poor outcomes across all gaps [40,43,59,76,80–82,85,86,96,101,104,104,108,110,125,132,134,139,156,157,175], while smoking was associated with poor outcomes across Gaps 1, 4, and 5 [59,104,108,132,138,151,156–158]. Undernutrition was associated with poor outcomes across Gaps 4 and 5 [84–86,96,111,116,118,121,122,154,176], though nutritional status was infrequently measured in earlier care cascade gaps. Public health strategies to reduce these risk factors—such as targeted nutritional supplementation to undernourished individuals in the general population [177], higher taxes on alcohol and tobacco products, or effective implementation of the ban on smoking in public places—may help to reduce TB incidence and improve care engagement among those who contract active TB. Active case-finding strategies could also focus on individuals with these comorbid conditions (e.g., case finding in bars) with incentives to enhance retention at each care cascade stage. At later care cascade stages, TB programs should integrate services—such as nutritional supplementation and counseling or medication-assisted therapy to reduce alcohol use and smoking—to improve treatment outcomes and reduce TB recurrence.

Finally, distrust or negative perceptions of local (usually government) health services—or dissatisfaction after initial contact with government TB services or providers—were associated with unfavorable outcomes or reported as barriers to care engagement across Gaps 1 to 4 [33,37,44,60,61,72,81,99,108,136,140]. While negative perceptions or interactions contributed to losses from government services, when people with TB did trust or prefer local government services (as compared to private services), they were more likely to have sought care for TB symptoms (Gap 1) [59] and had better outcomes during second episodes of TB treatment, as compared to patients previously treated in the private sector (Gap 4) [83]. People with TB evaluated at private sector facilities or laboratories were more likely to not pursue further workup for TB symptoms (Gap 2) [42] or to start TB treatment (Gap 3) [42,66], potentially because government NTEP providers have systems for following-up and linking patients to care that are generally absent in the private sector. These results align with findings of a systematic review of delays in TB care in India, which found initial contact with a private sector provider was independently associated with greater health system delay in diagnosis and treatment [26]. Our findings also align with results of a recent systematic review and standardized patient study in India, which found that, although quality of care is higher in the government TB program (e.g., higher rates of microbiological testing for TB), the client experience is generally better in the private sector [22,178]. Collectively, these findings suggest that government TB services should focus on improving perceptions and the experience of care (e.g., more polite provider behavior, shorter waiting times), while the private sector needs to improve quality of care.

### Findings specific to each care cascade gap

Our review also highlights findings that are more specific to each care cascade gap. Gap 1 findings indicate the importance of TB knowledge in the community to motivate care-seeking [35–37,59]. Using mass communication, including television and social media, to improve public knowledge of TB may be an important strategy to improve care-seeking. Population-level TB knowledge could be routinely measured during TB prevalence surveys or other population-based surveys [5,14]. Gap 1 findings also suggest structural barriers to care—including work-related, financial, and transportation barriers to reaching clinics [33,34,37,38,59–62,170]—must be addressed by increasing clinic accessibility or bringing screening closer to where people live or work through active case-finding. However, to be effective, our findings suggest that active case-finding initiatives must interface with people who are harder-to-reach (e.g., men, daily wage laborers) and incentivize people with lower symptom severity to engage in care.

For Gap 2, health system-related barriers played a central role in non-completion of the diagnostic workup. Accessibility of testing was crucial, with lack of free in-clinic or local access to chest X-rays or NAAT being key obstacles [48,49]. Sputum transportation contributed to NAAT non-completion, with one-third of samples not reaching the reference laboratory for testing in 1 study [49]. Several Gap 2 challenges related to suboptimal quality of care by healthcare providers. For example, providers missed identifying patients who met criteria to undergo NAAT (up to 54%) [51], particularly patients who were not already taking TB treatment and experiencing treatment failure. While algorithms for TB evaluation, including NAAT, have evolved since some of these studies were published [47], our findings suggest that algorithms that target advanced diagnostic testing to patient subgroups run the risk of excluding eligible patients. Communication gaps were also a problem, with many patients not completing the workup because they were not informed they were undergoing sputum testing or chest X-ray due to concern for TB [44,48,56]. Finally, negative interactions with healthcare providers during the diagnostic workup rapidly shaped patient perceptions, with nearly half of patients being lost due to such interactions in one multi-site study [44].

For Gap 3, a meaningful proportion of patients who experienced PTLFU (4% to 40%) [57,58,64,67,72,74,77] died after diagnosis but before starting treatment, likely due to advanced TB resulting from diagnostic delays. This finding suggests that, by identifying patients at earlier disease stages, active case-finding may improve later care cascade outcomes. TB stigma prevented some patients experiencing PTLFU (10% to 23%) [44,72,76] from starting treatment and may have contributed to high treatment refusal (up to 83% of those experiencing PTLFU) in some studies [44,57,58,67,77]. The contribution of TB stigma to losses in Gap 3 highlights the need for robust counseling and education from the start of the TB diagnostic workup. Poor recording of patient contact information by healthcare providers was associated with increased adjusted odds of PTLFU in one study [64] and contributed to the inability to track 10% to 52% of these patients [57,64,67,74,77]. Although India’s NTEP eventually records all patient information in Nikshay, a national case-based electronic medical record and reporting system, in most government TB clinics, patient data are recorded on paper forms before electronic entry, and such information is often incomplete or unreadable [64,172]. Regular auditing and performance feedback on the completeness and readability of TB diagnostic registers may help reduce PTLFU [179].

For Gap 4, given the rich and variable findings by patient population, we highlight key additional high-level points here. First, consistent with findings from clinical trials [20,180,181], non-adherence to TB medications was independently associated with increased risk of unfavorable treatment outcomes (Gap 4) [81,84,93,97,98,121] and TB recurrence (Gap 5) [158] across multiple Indian studies, which suggests that adherence is a mediator of outcomes in programmatic care. However, measuring TB medication adherence in routine care is challenging, with recent studies from India showing that 99DOTS (a digital adherence technology used in the NTEP) and patient self-report have suboptimal accuracy [182–184]. Use of novel approaches for detecting nonadherence, such as urine isoniazid testing [29,185], may facilitate early identification of patients at risk for poor outcomes, so that they can be provided with additional support. A recent study in India found that negative urine isoniazid test results were associated with loss to follow-up and death during treatment [29], which suggests this easy-to-use point-of-care test may be helpful in triaging patients for enhanced support.

Second, in Gap 4, TB medication adverse effects were a contributor to loss to follow-up during treatment across multiple studies for patients with drug-susceptible TB [80,93,98,99,108,138–143] (with up to 42-47% of patients who discontinued therapy doing so due to adverse effects [108,139,141]) and drug-resistant TB [123,127,186] (with up to 75% of patients who discontinued therapy doing so due to adverse effects [186]). Addressing adverse effects and other contributing factors to unfavorable treatment outcomes, such as TB stigma [108,136,168] and insufficient TB knowledge [81,140,141,143], will require development of more intensive counseling and psychosocial support interventions that can be integrated into routine care.

Finally, for Gap 5, post-treatment outcomes are largely shaped by the quality of care received in earlier care cascade stages. As such, Gap 5 findings reflect risk factors identified in earlier gaps—including male sex, older age, undernutrition, alcohol use, smoking, drug resistance, and medication nonadherence. Preventing post-treatment recurrence and death therefore depends on addressing these risk factors upstream in the care cascade. At the same time, close longitudinal post-treatment follow-up could facilitate early identification of new TB cases, thereby serving as a high-yield enhanced case-finding strategy, which may be especially important given the poor outcomes of people with previous TB.

### Limitations that may guide interpretation of review findings

As we juxtapose findings from studies from across India conducted over 20 years, our approach may raise concerns about external validity, or generalizability, given the diversity of India’s population [187]. Some findings may have programmatic relevance at the national level, because India’s NTEP is involved in the care of 2.1 million notified people with TB annually, of whom about 1.4 million were treated directly by public sector services, with the rest notified from the private sector [1]. However, given the importance of local context when developing implementation interventions [188], we discourage bluntly applying review findings without regard for setting. Rather, using causal transportability theory, program implementers can consider the extent to which risk factors identified in this review may be applicable to their setting to inform development of locally-relevant interventions [188]. In addition, for Gap 4, findings from studies in which the research was conducted before 2017 represent a time period before the rollout of daily fixed dose combination medications, when a thrice weekly intermittent medication dosing was used.

A second general limitation is our exclusion of qualitative findings, which will be reported in a future manuscript. Qualitative studies provide insights that may inform intervention development, which are often not obtainable from quantitative studies. Given that open-ended approaches are used to elicit information from patients, qualitative studies may shed greater light on the contribution of health system-related factors to poor TB outcomes [189–191].

A third limitation is relevant to our analysis of Gap 1, which is the largest gap in India’s TB care cascade [6]. Our Gap 1 analysis focused on understanding care-seeking by people with TB symptoms in the community; however, studies of delays in TB care and evaluations of healthcare provider behavior using standardized patients both suggest that heath system-related barriers, specifically poor quality of care, contribute substantially to patient losses in Gap 1 [24–26,162,178,192]. Program managers developing interventions to address Gap 1 should read these standardized patient and delay studies in parallel with our Gap 1 review findings.

## Conclusion

This systematic review organizes findings from 2 decades of studies on India’s TB care cascade to illuminate patterns of risk shaping outcomes for people with TB, while identifying gaps in knowledge to guide future research. In addition to summarizing gap-specific findings, we identify findings contributing to unfavorable outcomes across multiple care cascade gaps. These factors included male sex, older age, poverty-related barriers, lower symptom duration and severity, undernutrition, alcohol use, smoking, and distrust of (or dissatisfaction with) government health services. Our findings also suggest that, while quality of care is better in the government TB program, client experience is generally better in the private sector.

Closing gaps in the active TB care cascade will reduce mortality, enhance well-being and quality of life for people with TB, and help curb TB transmission. For these reasons, developing interventions to close these gaps must be central to India’s ambitious plan to eliminate TB [1,11] and to the WHO’s global End TB agenda. Our review provides rich insights that will hopefully inform the design of future implementation interventions and strategies to accelerate reduction in TB incidence, morbidity, and mortality in India.

## Data Availability

All data produced in the present work are contained in the manuscript or supplemental appendices.

## Acknowledgments

We are grateful to Drs. Rajaram S. and Gururaj Patil for conducting secondary analyses of data informing Gap 1 from the Tuberculosis Health Action Learning Initiative project implemented by the Karnataka Health Promotion Trust, which was funded by the United States Agency for International Development. We are also grateful to Amy Lapidow, librarian at the Tufts Hirsh Health Sciences Library, for assisting with the refresher searches for this review.

## References

1. World Health Organization. Global tuberculosis report 2022. Geneva, Switzerland: World Health Organization; 2022. Available: https://www.who.int/teams/global-tuberculosis-programme/tb-reports/global-tuberculosis-report-2022

2. Central TB Division. India TB report 2020. New Delhi, India: Ministry of Health and Family Welfare; 2020. Available: https://tbcindia.gov.in/showfile.php?lid=3538

3. Pai M, Kasaeva T, Swaminathan S. Covid-19’s Devastating Effect on Tuberculosis Care - A Path to Recovery. N Engl J Med. 2022;386: 1490–1493. doi:10.1056/NEJMp2118145

4. Jhaveri TA, Fung C, LaHood AN, Lindeborg A, Zeng C, Rahman R, et al. Clinical Outcomes of Individuals with COVID-19 and Tuberculosis during the Pre-Vaccination Period of the Pandemic: A Systematic Review. J Clin Med. 2022;11: 5656. doi:10.3390/jcm11195656

5. Subbaraman R, Nathavitharana RR, Mayer KH, Satyanarayana S, Chadha VK, Arinaminpathy N, et al. Constructing care cascades for active tuberculosis: A strategy for program monitoring and identifying gaps in quality of care. PLOS Med. 2019;16: e1002754. doi:10.1371/journal.pmed.1002754

6. Subbaraman R, Nathavitharana RR, Satyanarayana S, Pai M, Thomas BE, Chadha VK, et al. The Tuberculosis Cascade of Care in India’s Public Sector: A Systematic Review and Meta-analysis. PLOS Med. 2016;13: e1002149. doi:10.1371/journal.pmed.1002149

7. Naidoo P, Theron G, Rangaka MX, Chihota VN, Vaughan L, Brey ZO, et al. The South African Tuberculosis Care Cascade: Estimated Losses and Methodological Challenges. J Infect Dis. 2017;216: S702–S713. doi:10.1093/infdis/jix335

8. Lungu P, Kerkhoff AD, Kasapo CC, Mzyece J, Nyimbili S, Chimzizi R, et al. Tuberculosis care cascade in Zambia - identifying the gaps in order to improve outcomes: a population-based analysis. BMJ Open. 2021;11: e044867. doi:10.1136/bmjopen-2020-044867

9. Knoblauch AM, Grandjean Lapierre S, Randriamanana D, Raherison MS, Rakotoson A, Raholijaona BS, et al. Multidrug-resistant tuberculosis surveillance and cascade of care in Madagascar: a five-year (2012-2017) retrospective study. BMC Med. 2020;18: 173. doi:10.1186/s12916-020-01626-6

10. Oga-Omenka C, Boffa J, Kuye J, Dakum P, Menzies D, Zarowsky C. Understanding the gaps in DR-TB care cascade in Nigeria: A sequential mixed-method study. J Clin Tuberc Mycobact Dis. 2020;21: 100193. doi:10.1016/j.jctube.2020.100193

11. Central TB Division. National Strategic Plan for Tuberculosis Elimination in India 2017-2025. New Delhi, India: Ministry of Health and Family Welfare; 2017. Available: https://tbcindia.gov.in/WriteReadData/NSP%20Draft%2020.02.2017%201.pdf

12. World Health Organization. The End TB Strategy. Geneva, Switzerland; 2015 Aug. Available: https://www.who.int/teams/global-tuberculosis-programme/the-end-tb-strategy

13. United Nations Department of Economic and Social Affairs. Transforming our world: the 2030 Agenda for Sustainable Development. [cited 13 Apr 2023]. Available: https://sdgs.un.org/2030agenda

14. Subbaraman R, Jhaveri T, Nathavitharana RR. Closing gaps in the tuberculosis care cascade: an action-oriented research agenda. J Clin Tuberc Mycobact Dis. 2020;19: 100144. doi:10.1016/j.jctube.2020.100144

15. Khatri GR, Frieden TR. Controlling tuberculosis in India. N Engl J Med. 2002;347: 1420–1425. doi:10.1056/NEJMsa020098

16. Shibu V, Daksha S, Rishabh C, Sunil K, Devesh G, Lal S, et al. Tapping private health sector for public health program? Findings of a novel intervention to tackle TB in Mumbai, India. Indian J Tuberc. 2020;67: 189–201. doi:10.1016/j.ijtb.2020.01.007

17. Ananthakrishnan R, Richardson MD, van den Hof S, Rangaswamy R, Thiagesan R, Auguesteen S, et al. Successfully Engaging Private Providers to Improve Diagnosis, Notification, and Treatment of TB and Drug-Resistant TB: The EQUIP Public-Private Model in Chennai, India. Glob Health Sci Pract. 2019;7: 41–53. doi:10.9745/GHSP-D-18-00318

18. National TB Elimination Program of India. Technical and Operational Guidelines for TB Control in India 2016 [Internet]. New Delhi, India: Ministry of Health and Family Welfare; 2016. Available: https://tbcindia.gov.in/index1.php?lang=1&level=2&sublinkid=4573&lid=3177

19. World Health Organization Country Office for India. Standards for TB care in India. New Delhi, India: World Health Organization; 2014. Available: https://tbcindia.gov.in/showfile.php?lid=3061

20. Imperial MZ, Nahid P, Phillips PPJ, Davies GR, Fielding K, Hanna D, et al. A patient-level pooled analysis of treatment-shortening regimens for drug-susceptible pulmonary tuberculosis. Nat Med. 2018;24: 1708–1715. doi:10.1038/s41591-018-0224-2

21. Morgan RL, Whaley P, Thayer KA, Schünemann HJ. Identifying the PECO: A framework for formulating good questions to explore the association of environmental and other exposures with health outcomes. Environ Int. 2018;121: 1027–1031. doi:10.1016/j.envint.2018.07.015

22. Satyanarayana S, Subbaraman R, Shete P, Gore G, Das J, Cattamanchi A, et al. Quality of tuberculosis care in India: a systematic review. Int J Tuberc Lung Dis. 2015;19: 751–763. doi:10.5588/ijtld.15.0186

23. Cazabon D, Alsdurf H, Satyanarayana S, Nathavitharana R, Subbaraman R, Daftary A, et al. Quality of tuberculosis care in high burden countries: the urgent need to address gaps in the care cascade. Int J Infect Dis. 2017;56: 111–116. doi:10.1016/j.ijid.2016.10.016

24. Das J, Kwan A, Daniels B, Satyanarayana S, Subbaraman R, Bergkvist S, et al. Use of standardised patients to assess quality of tuberculosis care: a pilot, cross-sectional study. Lancet Infect Dis. 2015;15: 1305–1313. doi:10.1016/S1473-3099(15)00077-8

25. Kwan A, Daniels B, Saria V, Satyanarayana S, Subbaraman R, McDowell A, et al. Variations in the quality of tuberculosis care in urban India: A cross-sectional, standardized patient study in two cities. PLoS Med. 2018;15: e1002653. doi:10.1371/journal.pmed.1002653

26. Sreeramareddy CT, Qin ZZ, Satyanarayana S, Subbaraman R, Pai M. Delays in diagnosis and treatment of pulmonary tuberculosis in India: a systematic review. Int J Tuberc Lung Dis Off J Int Union Tuberc Lung Dis. 2014;18: 255–266. doi:10.5588/ijtld.13.0585

27. National TB Elimination Program of India. Technical and Operational Guidelines for TB Control in India 2016 [Internet]. New Delhi, India: Ministry of Health and Family Welfare; 2016. Available: https://tbcindia.gov.in/index1.php?lang=1&level=2&sublinkid=4573&lid=3177

28. World Health Organization. Definitions and reporting framework for tuberculosis – 2013 revision. Geneva, Switzerland: World Health Organization; 2013. Report No.: WHO/HTM/TB/2013.2. Available: https://apps.who.int/iris/bitstream/handle/10665/79199/9789241505345_eng.pdf

29. Subbaraman R, Thomas BE, Kumar JV, Thiruvengadam K, Khandewale A, Kokila S, et al. Understanding Nonadherence to Tuberculosis Medications in India Using Urine Drug Metabolite Testing: A Cohort Study. Open Forum Infect Dis. 2021;8: ofab190. doi:10.1093/ofid/ofab190

30. Bronfenbrenner U. Ecological systems theory. Six theories of child development: Revised formulations and current issues. Jessica Kingsley Publishers; 1992. pp. 187–249.

31. Ahmed S, Autrey J, Katz IT, Fox MP, Rosen S, Onoya D, et al. Why do people living with HIV not initiate treatment? A systematic review of qualitative evidence from low- and middle-income countries. Soc Sci Med. 2018;213: 72–84. doi:10.1016/j.socscimed.2018.05.048

32. Stop TB Partnership. Word matter: suggested language and usage for tuberculosis communications. Geneva, Switzerland: Stop TB Partnership; 2022 May. Available: https://www.stoptb.org/words-matter-language-guide

33. Karanjekar V, Gujarati V, Lokare P. Sociodemographic factors associated with health seeking behavior of chest symptomatics in urban slums of Aurangabad city, India. Int J Basic Appl Med Sci. 2014;4: 173–179.

34. Shewade HD, Gupta V, Satyanarayana S, Pandey P, Bajpai UN, Tripathy JP, et al. Patient characteristics, health seeking and delays among new sputum smear positive TB patients identified through active case finding when compared to passive case finding in India. PloS One. 2019;14: e0213345. doi:10.1371/journal.pone.0213345

35. Knowledge about TB and Health Seeking Behaviour among Adult Chest Symptomatic (CS) Population Living in Urban Slums of Hyderabad City. A Baseline Study Report: 2016-17. Karnataka Health Promotion Trust (KHPT) Tuberculosis Health Action Learning Initiative (THALI);

36. Knowledge about TB and Health Seeking Behaviour among Adult Chest Symptomatic (CS) Population Living in Urban Slums of Bengaluru City. A Baseline Study Report: 2016-17. Karnataka Health Promotion Trust (KHPT) Tuberculosis Health Action Learning Initiative (THALI);

37. George O, Sharma V, Sinha A, Bastian S, Santha T. Knowledge and behaviour of chest symptomatics in urban slum populations of two states in India towards care-seeking. Indian J Tuberc. 2013;60: 95–106.

38. Shriraam V, Srihari R, Gayathri T, Murali L. Active case finding for Tuberculosis among migrant brick kiln workers in South India. Indian J Tuberc. 2020;67: 38–42. doi:10.1016/j.ijtb.2019.09.003

39. Fochsen G, Deshpande K, Diwan V, Mishra A, Diwan VK, Thorson A. Health care seeking among individuals with cough and tuberculosis: a population-based study from rural India. Int J Tuberc Lung Dis Off J Int Union Tuberc Lung Dis. 2006;10: 995–1000.

40. Dey A, Thekkur P, Ghosh A, Dasgupta T, Bandopadhyay S, Lahiri A, et al. Active Case Finding for Tuberculosis through TOUCH Agents in Selected High TB Burden Wards of Kolkata, India: A Mixed Methods Study on Outcomes and Implementation Challenges. Trop Med Infect Dis. 2019;4. doi:10.3390/tropicalmed4040134

41. Garg T, Gupta V, Sen D, Verma M, Brouwer M, Mishra R, et al. Prediagnostic loss to follow-up in an active case finding tuberculosis programme: a mixed-methods study from rural Bihar, India. BMJ Open. 2020;10: e033706. doi:10.1136/bmjopen-2019-033706

42. Ismail IM, Kibballi Madhukeshwar A, Naik PR, Nayarmoole BM, Satyanarayana S. Magnitude and Reasons for Gaps in Tuberculosis Diagnostic Testing and Treatment Initiation: An Operational Research Study from Dakshina Kannada, South India. J Epidemiol Glob Health. 2020;10: 326–336. doi:10.2991/jegh.k.200516.001

43. Balasubramanian R, Garg R, Santha T, Gopi PG, Subramani R, Chandrasekaran V, et al. Gender disparities in tuberculosis: report from a rural DOTS programme in south India. Int J Tuberc Lung Dis Off J Int Union Tuberc Lung Dis. 2004;8: 323–332.

44. Dandona R, Dandona L, Mishra A, Dhingra S, Venkatagopalakrishna K, Chauhan LS. Utilization of and barriers to public sector tuberculosis services in India. Natl Med J India. 2004;17: 292–299.

45. Das M, Pasupuleti D, Rao S, Sloan S, Mansoor H, Kalon S, et al. GeneXpert and Community Health Workers Supported Patient Tracing for Tuberculosis Diagnosis in Conflict-Affected Border Areas in India. Trop Med Infect Dis. 2019;5. doi:10.3390/tropicalmed5010001

46. Tripathy JP, Srinath S, Naidoo P, Ananthakrishnan R, Bhaskar R. Is physical access an impediment to tuberculosis diagnosis and treatment? A study from a rural district in North India. Public Health Action. 2013;3: 235–239. doi:10.5588/pha.13.0044

47. Kanakaraju M, Nagaraja SB, Satyanarayana S, Babu YR, Madhukeshwar AK, Narasimhaiah S. Chest Radiography and Xpert MTB/RIF® Testing in Persons with Presumptive Pulmonary TB: Gaps and Challenges from a District in Karnataka, India. Tuberc Res Treat. 2020;2020: 5632810. doi:10.1155/2020/5632810

48. Sarkar J, Murhekar MV. Factors associated with low utilization of x-ray facilities among the sputum negative chest symptomatics in Jalpaiguri district (West Bengal) 2009. Indian J Tuberc. 2011;58: 208–211.

49. Shewade HD, Govindarajan S, Sharath BN, Tripathy JP, Chinnakali P, Kumar AMV, et al. MDR-TB screening in a setting with molecular diagnostic techniques: who got tested, who didn’t and why? Public Health Action. 2015;5: 132–139. doi:10.5588/pha.14.0098

50. Shewade HD, Govindarajan S, Thekkur P, Palanivel C, Muthaiah M, Kumar AMV, et al. MDR-TB in Puducherry, India: reduction in attrition and turnaround time in the diagnosis and treatment pathway. Public Health Action. 2016;6: 242–246. doi:10.5588/pha.16.0075

51. Shewade HD, Kokane AM, Singh AR, Verma M, Parmar M, Chauhan A, et al. High pre-diagnosis attrition among patients with presumptive MDR-TB: an operational research from Bhopal district, India. BMC Health Serv Res. 2017;17: 249. doi:10.1186/s12913-017-2191-6

52. Shewade HD, Nair D, Klinton JS, Parmar M, Lavanya J, Murali L, et al. Low pre-diagnosis attrition but high pre-treatment attrition among patients with MDR-TB: An operational research from Chennai, India. J Epidemiol Glob Health. 2017;7: 227–233. doi:10.1016/j.jegh.2017.07.001

53. Singla N, Satyanarayana S, Sachdeva KS, Van den Bergh R, Reid T, Tayler-Smith K, et al. Impact of introducing the line probe assay on time to treatment initiation of MDR-TB in Delhi, India. PloS One. 2014;9: e102989. doi:10.1371/journal.pone.0102989

54. Chandrasekaran V, Ramachandran R, Cunningham J, Balasubramanian R, Thomas A, Sudha G, et al. Factors leading to tuberculosis diagnostic drop-out and delayed treatment initiation in Chennai, India. Int J Tuberc Lung Dis. 2005;9: S172.

55. Chadha VK, Praseeja P, Hemanthkumar NK, Shivshankara BA, Sharada MA, Nagendra N, et al. Implementation efficiency of a diagnostic algorithm in sputum smear-negative presumptive tuberculosis patients. Int J Tuberc Lung Dis Off J Int Union Tuberc Lung Dis. 2014;18: 1237–1242. doi:10.5588/ijtld.14.0218

56. Thomas A, Gopi PG, Santha T, Jaggarajamma K, Charles N, Prabhakaran E, et al. Course of action taken by smear negative chest symptomatics: A report from a rural area in South India. Indian J Tuberc. 2006;53: 4–6.

57. Chadha SS, Sharath BN, Reddy K, Jaju J, Vishnu PH, Rao S, et al. Operational challenges in diagnosing multi-drug resistant TB and initiating treatment in Andhra Pradesh, India. PloS One. 2011;6: e26659. doi:10.1371/journal.pone.0026659

58. Natrajan S, Singh AR, Shewade HD, Verma M, Bali S. Pre-diagnosis attrition in patients with presumptive MDR-TB in Bhopal, India, 2015: a follow-up study. Public Health Action. 2018;8: 95–96. doi:10.5588/pha.18.0015

59. Helfinstein S, Engl E, Thomas BE, Natarajan G, Prakash P, Jain M, et al. Understanding why at-risk population segments do not seek care for tuberculosis: a precision public health approach in South India. BMJ Glob Health. 2020;5. doi:10.1136/bmjgh-2020-002555

60. Charles N, Thomas B, Watson B, Raja Sakthivel M, Chandrasekeran V, Wares F. Care seeking behavior of chest symptomatics: a community based study done in South India after the implementation of the RNTCP. PloS One. 2010;5. doi:10.1371/journal.pone.0012379

61. Thomas BE, Thiruvengadam K, S R, Rani S, S V, Gangadhar Rao V, et al. Understanding health care-seeking behaviour of the tribal population in India among those with presumptive TB symptoms. PloS One. 2021;16: e0250971. doi:10.1371/journal.pone.0250971

62. Thomas BE, Charles N, Watson B, Chandrasekaran V, Senthil Kumar R, Dhanalakshmi A, et al. Prevalence of chest symptoms amongst brick kiln migrant workers and care seeking behaviour: a study from South India. J Public Health Oxf Engl. 2015;37: 590–596. doi:10.1093/pubmed/fdu104

63. Ghosh S, Sinhababu A, Taraphdar P, Mukhopadhyay DK, Mahapatra BS, Biswas AB. A study on care seeking behavior of chest symptomatics in a slum of Bankura, West Bengal. Indian J Public Health. 2010;54: 42–44. doi:10.4103/0019-557X.70553

64. Thomas BE, Subbaraman R, Sellappan S, Suresh C, Lavanya J, Lincy S, et al. Pretreatment loss to follow-up of tuberculosis patients in Chennai, India: a cohort study with implications for health systems strengthening. BMC Infect Dis. 2018;18: 142. doi:10.1186/s12879-018-3039-3

65. Raizada N, Khaparde SD, Salhotra VS, Rao R, Kalra A, Swaminathan S, et al. Accelerating access to quality TB care for pediatric TB cases through better diagnostic strategy in four major cities of India. PloS One. 2018;13: e0193194. doi:10.1371/journal.pone.0193194

66. Pardeshi G, Deluca A, Agarwal S, Kishore J. Tuberculosis patients not covered by treatment in public health services: findings from India’s National Family Health Survey 2015-16. Trop Med Int Health TM IH. 2018;23: 886–895. doi:10.1111/tmi.13086

67. Sai Babu B, Satyanarayana AVV, Venkateshwaralu G, Ramakrishna U, Vikram P, Sahu S, et al. Initial default among diagnosed sputum smear-positive pulmonary tuberculosis patients in Andhra Pradesh, India. Int J Tuberc Lung Dis Off J Int Union Tuberc Lung Dis. 2008;12: 1055–1058.

68. Mandal A, Basu M, Das P, Mukherjee S, Das S, Roy N. Magnitude and reasons of initial default among new sputum positive cases of pulmonary tuberculosis under RNTCP in a district of West Bengal, India. South East Asia J Public Health. 2014;4: 41–47. doi:10.3329/seajph.v4i1.21839

69. Jain S, Varudkar HG, Julka A, Singapurwala M, Khosla S, Shah B. Socio-economical and Clinico-Radiological Profile of 474 MDR TB Cases of a Rural Medical College. J Assoc Physicians India. 2018;66: 14–18.

70. Khandekar J, Acharya AS, R TH, Sharma A. Do patients with tuberculosis referred from a tertiary care referral centre reach their peripheral health institution? Natl Med J India. 2013;26: 332–334.

71. Kumar D, Goel C, Bansal AK, Bhardwaj AK. Delineating the factors associated with recurrence of tuberculosis in programmatic settings of rural health block, Himachal Pradesh, India. Indian J Tuberc. 2018;65: 303–307. doi:10.1016/j.ijtb.2018.07.001

72. Mehra D, Kaushik RM, Kaushik R, Rawat J, Kakkar R. Initial default among sputum-positive pulmonary TB patients at a referral hospital in Uttarakhand, India. Trans R Soc Trop Med Hyg. 2013;107: 558–565. doi:10.1093/trstmh/trt065

73. Rawat J, Biswas D, Sindhwani G, Kesharwani V, Masih V, Chauhan BS. Diagnostic defaulters: an overlooked aspect in the Indian Revised National Tuberculosis Control Program. J Infect Dev Ctries. 2012;6: 20–22. doi:10.3855/jidc.1895

74. Gopi P, Chandrasekaran V, Subramani R, Narayanan P. Failure to initiate treatment for tuberculosis patients diagnosed in a community survey and at health facilities under a DOTS program in a district of south India. Indian J Tuberc. 2005;52: 153–156.

75. Singh M, Sagili KD, Tripathy JP, Kishore S, Bahurupi YA, Kumar A, et al. Are Treatment Outcomes of Patients with Tuberculosis Detected by Active Case Finding Different From Those Detected by Passive Case Finding? J Glob Infect Dis. 2020;12: 28–33. doi:10.4103/jgid.jgid_66_19

76. Pillai D, Purty A, Prabakaran S, Singh Z, Soundappan G, Anandan V. Initial default among tuberculosis patients diagnosed in selected medical colleges of Puducherry: issues and possible interventions. 2015. doi:10.5455/IJMSPH.2015.30012015196

77. Dave P, Nimavat P, Shah A, Pujara K, Patel P, Modi B. Knowing more about initial default among diagnosed sputum smear-positive pulmonary tuberculosis patients in Gujarat, India [Abstract PC-868-03]. Int J Tuberc Lung Dis. 2013;17: S469.

78. Shewade HD, Shringarpure KS, Parmar M, Patel N, Kuriya S, Shihora S, et al. Delay and attrition before treatment initiation among MDR-TB patients in five districts of Gujarat, India. Public Health Action. 2018;8: 59–65. doi:10.5588/pha.18.0003

79. Ramachandran G, Chandrasekaran P, Gaikwad S, Agibothu Kupparam HK, Thiruvengadam K, Gupte N, et al. Subtherapeutic Rifampicin Concentration Is Associated With Unfavorable Tuberculosis Treatment Outcomes. Clin Infect Dis. 2020;70: 1463–1470. doi:10.1093/cid/ciz380

80. Velayutham B, Chadha VK, Singla N, Narang P, Gangadhar Rao V, Nair S, et al. Recurrence of tuberculosis among newly diagnosed sputum positive pulmonary tuberculosis patients treated under the Revised National Tuberculosis Control Programme, India: A multi-centric prospective study. PloS One. 2018;13: e0200150. doi:10.1371/journal.pone.0200150

81. Vijay S, Kumar P, Chauhan LS, Vollepore BH, Kizhakkethil UP, Rao SG. Risk Factors Associated with Default among New Smear Positive TB Patients Treated Under DOTS in India. Pai M, editor. PLoS ONE. 2010;5: e10043. doi:10.1371/journal.pone.0010043

82. Zhou TJ, Lakshminarayanan S, Sarkar S, Knudsen S, Horsburgh CR, Muthaiah M, et al. Predictors of Loss to Follow-Up among Men with Tuberculosis in Puducherry and Tamil Nadu, India. Am J Trop Med Hyg. 2020;103: 1050–1056. doi:10.4269/ajtmh.19-0415

83. Jha UM, Satyanarayana S, Dewan PK, Chadha S, Wares F, Sahu S, et al. Risk Factors for Treatment Default among Re-Treatment Tuberculosis Patients in India, 2006. Pai M, editor. PLoS ONE. 2010;5: e8873. doi:10.1371/journal.pone.0008873

84. Parmar MM, Sachdeva KS, Dewan PK, Rade K, Nair SA, Pant R, et al. Unacceptable treatment outcomes and associated factors among India’s initial cohorts of multidrug-resistant tuberculosis (MDR-TB) patients under the revised national TB control programme (2007–2011): Evidence leading to policy enhancement. Neyrolles O, editor. PLOS ONE. 2018;13: e0193903. doi:10.1371/journal.pone.0193903

85. Cox SR, Gupte AN, Thomas B, Gaikwad S, Mave V, Padmapriyadarsini C, et al. Unhealthy alcohol use independently associated with unfavorable TB treatment outcomes among Indian men. Int J Tuberc Lung Dis Off J Int Union Tuberc Lung Dis. 2021;25: 182–190. doi:10.5588/ijtld.20.0778

86. Washington R, Potty RS, Rajesham A, Seenappa T, Singarajipura A, Swamickan R, et al. Is a differentiated care model needed for patients with TB? A cohort analysis of risk factors contributing to unfavourable outcomes among TB patients in two states in South India. BMC Public Health. 2020;20: 1158. doi:10.1186/s12889-020-09257-5

87. Isaakidis P, Varghese B, Mansoor H, Cox HS, Ladomirska J, Saranchuk P, et al. Adverse Events among HIV/MDR-TB Co-Infected Patients Receiving Antiretroviral and Second Line Anti-TB Treatment in Mumbai, India. Wilkinson RJ, editor. PLoS ONE. 2012;7: e40781. doi:10.1371/journal.pone.0040781

88. Saha A, Vaidya PJ, Chavhan VB, Pandey KV, Kate AH, Leuppi JD, et al. Factors affecting outcomes of individualised treatment for drug resistant tuberculosis in an endemic region>. Tuberculosis. European Respiratory Society; 2017. p. PA2728. doi:10.1183/1393003.congress-2017.PA2728

89. Huddart S, Singh M, Jha N, Benedetti A, Pai M. Case fatality and recurrent tuberculosis among patients managed in the private sector: A cohort study in Patna, India. PLOS ONE. 2021;16: e0249225. doi:10.1371/journal.pone.0249225

90. Secretary Of Jan Swasthya Sahyog null, Laux TS, Patil S. Predictors of tuberculosis treatment outcomes among a retrospective cohort in rural, Central India. J Clin Tuberc Mycobact Dis. 2018;12: 41–47. doi:10.1016/j.jctube.2018.06.005

91. Sharma V, Thekkur P, Naik PR, Saha BK, Agrawal N, Dinda MK, et al. Treatment success rates among tuberculosis patients notified from the private sector in West Bengal, India. Monaldi Arch Chest Dis. 2021;91. doi:10.4081/monaldi.2021.1555

92. Gopi PG, Chandrasekaran V, Subramani R, Santha T, Thomas A, Selvakumar N, et al. Association of conversion & cure with initial smear grading among new smear positive pulmonary tuberculosis patients treated with Category I regimen. INDIAN J MED RES. 2006; 8.

93. Kulkarni P, Akarte S, Mankeshwar R, Bhawalkar J, Banerjee A, Kulkarni A. Non-Adherence of New Pulmonary Tuberculosis Patients to Anti-Tuberculosis Treatment. Ann Med Health Sci Res. 2013;3: 67. doi:10.4103/2141-9248.109507

94. Joseph N. Treatment outcomes among new smear positive and retreatment cases of tuberculosis in Mangalore, South India – a descriptive study. Australas Med J. 2011;4: 162–167. doi:10.4066/AMJ.2011.585

95. Velayutham BRV, Nair D, Chandrasekaran V, Raman B, Sekar G, Watson B, et al. Profile and Response to Anti-Tuberculosis Treatment among Elderly Tuberculosis Patients Treated under the TB Control Programme in South India. Cattamanchi A, editor. PLoS ONE. 2014;9: e88045. doi:10.1371/journal.pone.0088045

96. Mave V, Gaikwad S, Barthwal M, Chandanwale A, Lokhande R, Kadam D, et al. Diabetes Mellitus and Tuberculosis Treatment Outcomes in Pune, India. Open Forum Infect Dis. 2021;8. doi:10.1093/ofid/ofab097

97. Singla R, Bharty SK, Gupta UA, Khayyam KU, Vohra V, Singla N, et al. Sputum smear positivity at two months in previously untreated pulmonary tuberculosis patients. Int J Mycobacteriology. 2013;2: 199–205. doi:10.1016/j.ijmyco.2013.08.002

98. Singla R, Sarin R, Khalid UK, Mathuria K, Singla N, Jaiswal A, et al. Seven-year DOTS-Plus pilot experience in India: results, constraints and issues. : 6.

99. Zaman F, Sheikh S, Das K, Zaman G, Pal R. An epidemiological study of newly diagnosed sputum positive tuberculosis patients in Dhubri district, Assam, India and the factors influencing their compliance to treatment. J Nat Sci Biol Med. 2014;5: 415. doi:10.4103/0976-9668.136213

100. Tiwari S, Kumar A, Kapoor SK. RELATIONSHIP BETWEEN SPUTUM SMEAR GRADING AND SMEAR CONVERSION RATE AND TREATMENT OUTCOME IN THE PATIENTS OF PULMONARY TUBERCULOSIS UNDERGOING DOTS-A PROSPECTIVE COHORT STUDY. Indian J Tuberc. : 6.

101. M. S, K. M, Marconi S, V. K, S. R, Prasad J. A community based case control study on risk factors for treatment interruptions in people with tuberculosis in Kollam district, Kerala, southern India. Int J Community Med Public Health. 2016; 962–967. doi:10.18203/2394-6040.ijcmph20160937

102. Viswanathan V, Vigneswari A, Selvan K, Satyavani K, Rajeswari R, Kapur A. Effect of diabetes on treatment outcome of smear-positive pulmonary tuberculosis—A report from South India. J Diabetes Complications. 2014;28: 162–165. doi:10.1016/j.jdiacomp.2013.12.003

103. Vashishtha R, Mohan K, Singh B, Devarapu SK, Sreenivas V, Ranjan S, et al. Efficacy and safety of thrice weekly DOTS in tuberculosis patients with and without HIV co-infection: an observational study. BMC Infect Dis. 2013;13: 468. doi:10.1186/1471-2334-13-468

104. Bagchi S, Ambe G, Sathiakumar N. Determinants of Poor Adherence to Anti-Tuberculosis Treatment in Mumbai, India. : 13.

105. Babiarz KS, Suen S, Goldhaber-Fiebert JD. Tuberculosis treatment discontinuation and symptom persistence: an observational study of Bihar, India’s public care system covering >100,000,000 inhabitants. BMC Public Health. 2014;14: 418. doi:10.1186/1471-2458-14-418

106. Ahmed J, Chadha VK, Singh S, Venkatachalappa B, Kumar P. Utilization of RNTCP services in rural areas of Bellary District, Karnataka, by gender, age and distance from health centre. Indian J Tuberc. 2009;56: 62–68.

107. Deepa D, Achanta S, Jaju J, Rao K, Samyukta R, Claassens M, et al. The Impact of Isoniazid Resistance on the Treatment Outcomes of Smear Positive Re-Treatment Tuberculosis Patients in the State of Andhra Pradesh, India. Neyrolles O, editor. PLoS ONE. 2013;8: e76189. doi:10.1371/journal.pone.0076189

108. Sarpal SS. Treatment Outcome Among the Retreatment Tuberculosis (TB) Patients Under RNTCP in Chandigarh, India. J Clin Diagn Res. 2014 [cited 11 Dec 2022]. doi:10.7860/JCDR/2014/6510.4006

109. Srinath S, Sharath B, Santosha K, Chadha SS, Roopa S, Chander K, et al. Tuberculosis “retreatment others”: profile and treatment outcomes in the state of Andhra Pradesh, India. Int J Tuberc Lung Dis Off J Int Union Tuberc Lung Dis. 2011;15: 105–109.

110. Bhagat VM, Gattani PL. FACTORS AFFECTING TUBERCULOSIS RETREATMENT DEFAULTS IN NANDED, INDIA. SOUTHEAST ASIAN J TROP MED PUBLIC Health. 2010;41: 5.

111. Velavan A, Purty AJ, Shringarpure K, Sagili KD, Mishra AK, Selvaraj KS, et al. Tuberculosis retreatment outcomes and associated factors: a mixed-methods study from Puducherry, India. Public Health Action. 2018;8: 187–193. doi:10.5588/pha.18.0038

112. Chandrasekaran V, Gopi PG, Santha T, Subramani R, Narayanan PR. STATUS OF RE-REGISTERED PATIENTS FOR TUBERCULOSIS TREATMENT UNDER DOTS PROGRAMME. Indian J Tuberc. : 5.

113. Mukherjee A, Sarkar A, Saha I, Biswas B, Bhattacharyya P. Outcomes of different subgroups of smear-positive retreatment patients under RNTCP in rural West Bengal, India. Rural Remote Health. 2009 [cited 11 Dec 2022]. doi:10.22605/RRH926

114. Sisodia RS, Wares DF, Sahu S, Chauhan LS, Zignol M. Source of retreatment cases under the Revised National TB Control Programme in Rajasthan, India, 2003. : 7.

115. Burugina Nagaraja S, Satyanarayana S, Chadha SS, Kalemane S, Jaju J, Achanta S, et al. How do patients who fail first-line TB treatment but who are not placed on an MDR-TB regimen fare in South India? PloS One. 2011;6: e25698. doi:10.1371/journal.pone.0025698

116. Sharma N, Khanna A, Chandra S, Basu S, Chopra K, Singla N, et al. Trends & treatment outcomes of multidrug-resistant tuberculosis in Delhi, India (2009-2014): A retrospective record-based study. Indian J Med Res. 2020;151: 598. doi:10.4103/ijmr.IJMR_1048_18

117. Nair D, Navneethapandian PD, Tripathy JP, Harries AD, Klinton JS, Watson B, et al. Impact of rapid molecular diagnostic tests on time to treatment initiation and outcomes in patients with multidrug-resistant tuberculosis, Tamil Nadu, India. Trans R Soc Trop Med Hyg. 2016;110: 534–541. doi:10.1093/trstmh/trw060

118. Bhatt R, Chopra K, Vashisht R. Impact of integrated psycho-socio-economic support on treatment outcome in drug resistant tuberculosis – A retrospective cohort study. Indian J Tuberc. 2019;66: 105–110. doi:10.1016/j.ijtb.2018.05.020

119. Jain K, Desai M, Solanki R, Dikshit RK. Treatment outcome of andardized regimen in patients with multidrug resistant tuberculosis. J Pharmacol Pharmacother. 2014;5: 145–149. doi:10.4103/0976-500X.130062

120. Kandi S, K TK, Kandi SR, Mathur N, D CD, Adepu R. Study of treatment outcomes of multidrug-resistant tuberculosis under programmatic conditions and factors influencing the outcomes in Hyderabad District. Indian J Tuberc. 2021;68: 379–383. doi:10.1016/j.ijtb.2020.12.008

121. Janmeja AK, Aggarwal D, Dhillon R. Factors predicting treatment success in multi-drug resistant tuberculosis patients treated under programmatic conditions. Indian J Tuberc. 2018;65: 135–139. doi:10.1016/j.ijtb.2017.12.015

122. Lohiya S, Tripathy JP, Sagili K, Khanna V, Kumar R, Ojha A, et al. Does Drug-Resistant Extrapulmonary Tuberculosis Hinder TB Elimination Plans? A Case from Delhi, India. Trop Med Infect Dis. 2020;5: 109. doi:10.3390/tropicalmed5030109

123. Dole SS, Waghmare V, Shaikh A. Clinical Profile and Treatment Outcome of Drug Resistant Tuberculosis Patients of Western Maharashtra, India. : 4.

124. Shringarpure KS, Isaakidis P, Sagili KD, Baxi RK. Loss-To-Follow-Up on Multidrug Resistant Tuberculosis Treatment in Gujarat, India: The WHEN and WHO of It. PloS One. 2015;10: e0132543. doi:10.1371/journal.pone.0132543

125. Duraisamy K, Mrithyunjayan S, Ghosh S, Nair SA, Balakrishnan S, Subramoniapillai J, et al. Does Alcohol Consumption during Multidrug-resistant Tuberculosis Treatment Affect Outcome?. A Population-based Study in Kerala, India. Ann Am Thorac Soc. 2014;11: 712–718. doi:10.1513/AnnalsATS.201312-447OC

126. Natarajan S, Singla R, Singla N, Gupta A, Caminero JA, Chakraborty A, et al. Treatment interruption patterns and adverse events among patients on bedaquiline containing regimen under programmatic conditions in India. Pulmonology. 2022;28: 203–209. doi:10.1016/j.pulmoe.2020.09.006

127. Patel SV, Nimavat KB, Patel AB, Mehta KG, Shringarpure K, Shukla LK. Sputum Smear and Culture Conversion in Multidrug Resistance Tuberculosis Patients in Seven Districts of Central Gujarat, India: A Longitudinal Study. : 4.

128. Banerjee S, Bandyopadhyay K, Taraphdar P, Dasgupta A. Perceived discrimination among tuberculosis patients in an urban area of Kolkata City, India. J Glob Infect Dis. 2020;12: 144. doi:10.4103/jgid.jgid_146_19

129. Mundra A, Deshmukh PR, Dawale A. Magnitude and determinants of adverse treatment outcomes among tuberculosis patients registered under Revised National Tuberculosis Control Program in a Tuberculosis Unit, Wardha, Central India: A record-based cohort study. J Epidemiol Glob Health. 2017;7: 111. doi:10.1016/j.jegh.2017.02.002

130. Kv N, Duraisamy K, Balakrishnan S, M S, S JS, Sagili KD, et al. Outcome of Tuberculosis Treatment in Patients with Diabetes Mellitus Treated in the Revised National Tuberculosis Control Programme in Malappuram District, Kerala, India. Wilkinson RJ, editor. PLoS ONE. 2013;8: e76275. doi:10.1371/journal.pone.0076275

131. Patra S, Lukhmana S, Tayler Smith K, Kannan AT, Satyanarayana S, Enarson DA, et al. Profile and treatment outcomes of elderly patients with tuberculosis in Delhi, India: implications for their management. Trans R Soc Trop Med Hyg. 2013;107: 763–768. doi:10.1093/trstmh/trt094

132. Prudhivi R, Challa SR, Rao MV B, Veena G V, Rao N B, Manogna Narne H. Assessment of Success Rate of Directly Observed Treatment Short-Course (DOTS) in Tuberculosis Patients of South India. J Young Pharm. 2018;11: 67–72. doi:10.5530/jyp.2019.11.14

133. Das M, Isaakidis P, Armstrong E, Gundipudi NR, Babu RB, Qureshi IA, et al. Directly-Observed and Self-Administered Tuberculosis Treatment in a Chronic, Low-Intensity Conflict Setting in India. Wilkinson RJ, editor. PLoS ONE. 2014;9: e92131. doi:10.1371/journal.pone.0092131

134. Vasantha M, Gopi PG, Subramani R. SURVIVAL OF TUBERCULOSIS PATIENTS TREATED UNDER DOTS IN A RURAL TUBERCULOSIS UNIT (TU), SOUTH INDIA. Indian J Tuberc. : 6.

135. Gopi PG, Vasantha M, Muniyandi M, Chandrasekaran V, Balasubramanian R, Narayanan PR. RISK FACTORS FOR NON-ADHERENCE TO DIRECTLY OBSERVED TREATMENT (DOT) IN A RURAL TUBERCULOSIS UNIT, SOUTH INDIA. Indian J Tuberc. : 6.

136. Mundra A, Deshmukh P, Dawale A. Determinants of adverse treatment outcomes among patients treated under Revised National Tuberculosis Control Program in Wardha, India: Case–control study. Med J Armed Forces India. 2018;74: 241–249. doi:10.1016/j.mjafi.2017.07.008

137. Gopi PG, Vasantha M, Muniyandi M, Chandrasekaran V, Balasubramanian R, Narayanan PR. RISK FACTORS FOR NON-ADHERENCE TO DIRECTLY OBSERVED TREATMENT (DOT) IN A RURAL TUBERCULOSIS UNIT, SOUTH INDIA. Indian J Tuberc. : 6.

138. Lata S, Khajuria V, Sawhney V, Kumari K. EVALUATION OF NON-ADHERENCE TO ANTITUBERCULAR DRUGS AMONG TUBERCULOSIS PATIENTS: A PROSPECTIVE STUDY. Int J Curr Pharm Res. 2021; 26–28. doi:10.22159/ijcpr.2021v13i2.41550

139. Jaggarajamma K, Sudha G, Chandrasekaran V, Nirupa C, Thomas A, Santha T, et al. REASONS FOR NON-COMPLIANCE AMONG PATIENTS TREATED UNDER REVISED NATIONAL TUBERCULOSIS CONTROL PROGRAMME (RNTCP), TIRUVALLUR DISTRICT, SOUTH INDIA. Indian J Tuberc. : 6.

140. Gupta S, Gupta S, Behera D. REASONS FOR INTERRUPTION OF ANTI-TUBERCULAR TREATMENT AS REPORTED BY PATIENTS WITH TUBERCULOSIS ADMITTED IN A TERTIARY CARE INSTITUTE. Indian J Tuberc. : 7.

141. Mittal C, Gupta S. Noncompliance to DOTS: How it can be decreased. Indian J Community Med. 2011;36: 27. doi:10.4103/0970-0218.80789

142. Shabil M, Rajesh V, Raj KCB, Rajesh KS, Shama KP, Gururaja MP, et al. A Study on Treatment Defaulters in Tuberculosis Patients on Dots Therapy. Res J Pharm Technol. 2019;12: 2245. doi:10.5958/0974-360X.2019.00374.3

143. Yadav GS, Jangid VK, Mathur BB. Study of various reasons for interruption of anti-tubercular treatment in patients of tuberculosis reporting to tertiary care center of west Rajasthan. Int J Res Med Sci. 2019;7: 2220. doi:10.18203/2320-6012.ijrms20192542

144. Vijay S, Kumar P, Chauhan LS, Narayan Rao SV, Vaidyanathan P. Treatment Outcome and Mortality at One and Half Year Follow-Up of HIV Infected TB Patients Under TB Control Programme in a District of South India. Pai M, editor. PLoS ONE. 2011;6: e21008. doi:10.1371/journal.pone.0021008

145. Sharma SK, Soneja M, Prasad KT, Ranjan S. Clinical profile & predictors of poor outcome of adult HIV-tuberculosis patients in a tertiary care centre in north India. : 12.

146. Shastri S, Naik B, Shet A, Rewari B, De Costa A. TB treatment outcomes among TB-HIV co-infections in Karnataka, India: how do these compare with non-HIV tuberculosis outcomes in the province? BMC Public Health. 2013;13: 838. doi:10.1186/1471-2458-13-838

147. Dhakulkar S, Das M, Sutar N, Oswal V, Shah D, Ravi S, et al. Treatment outcomes of children and adolescents receiving drug-resistant TB treatment in a routine TB programme, Mumbai, India. Dholakia YN, editor. PLOS ONE. 2021;16: e0246639. doi:10.1371/journal.pone.0246639

148. Satyanarayana S, Shivashankar R, Vashist RP, Chauhan LS, Chadha SS, Dewan PK, et al. Characteristics and Programme-Defined Treatment Outcomes among Childhood Tuberculosis (TB) Patients under the National TB Programme in Delhi. Madhi SA, editor. PLoS ONE. 2010;5: e13338. doi:10.1371/journal.pone.0013338

149. Sadana P, Verma V, Nagpal M. A study on predictors of treatment outcome among children registered under DOTS in district Tarn Taran, Punjab. Indian J Community Health. 2020;32: 399–403. doi:10.47203/IJCH.2020.v32i02.017

150. Lisha PV, James PT, Ravindran C. Morbidity and mortality at five years after initiating Category I treatment among patients with new sputum smear positive pulmonary tuberculosis. Indian J Tuberc. 2012;59: 83–91.

151. Mahishale V, Patil B, Lolly M, Eti A, Khan S. Prevalence of Smoking and Its Impact on Treatment Outcomes in Newly Diagnosed Pulmonary Tuberculosis Patients: A Hospital-Based Prospective Study. Chonnam Med J. 2015;51: 86–90. doi:10.4068/cmj.2015.51.2.86

152. Tripathy S, Anand A, Inamdar V, Manoj MM, Khillare KM, Datye AS, et al. Clinical response of newly diagnosed HIV seropositive & seronegative pulmonary tuberculosis patients with the RNTCP Short Course regimen in Pune, India. Indian J Med Res. 2011;133: 521–528.

153. Dandekar R, Dixit J, Srinivasan D. The fate of tuberculosis cases after two years of DOTS chemotherapy in Aurangabad city, Maharashtra. Natl J Community Med. 2014;5: 174–178.

154. Sadacharam K, Gopi PG, Chandrasekaran V, Eusuff SI, Subramani R, Santha T, et al. Status of smear-positive TB patients at 2-3 years after initiation of treatment under a DOTS programme. Indian J Tuberc. 2007;54: 199–203.

155. Sharma R, Prajapati S, Patel P, Patel B, Gajjar S, Bapat N. An Outcome-Based Follow-up Study of Cured Category I Pulmonary Tuberculosis Adult Cases from Various Tuberculosis Units under Revised National Tuberculosis Control Program from a Western Indian City. Indian J Community Med Off Publ Indian Assoc Prev Soc Med. 2019;44: 48–52. doi:10.4103/ijcm.IJCM_310_18

156. Kolappan C, Subramani R, Kumaraswami V, Santha T, Narayanan PR. Excess mortality and risk factors for mortality among a cohort of TB patients from rural south India. Int J Tuberc Lung Dis Off J Int Union Tuberc Lung Dis. 2008;12: 81–86.

157. Kolappan C, Subramani R, Karunakaran K, Narayanan PR. Mortality of tuberculosis patients in Chennai, India. Bull World Health Organ. 2006;84: 555–560. doi:10.2471/blt.05.022087

158. Thomas A, Gopi PG, Santha T, Chandrasekaran V, Subramani R, Selvakumar N, et al. Predictors of relapse among pulmonary tuberculosis patients treated in a DOTS programme in South India. Int J Tuberc Lung Dis Off J Int Union Tuberc Lung Dis. 2005;9: 556–561.

159. Gupte AN, Selvaraju S, Paradkar M, Danasekaran K, Shivakumar SVBY, Thiruvengadam K, et al. Respiratory health status is associated with treatment outcomes in pulmonary tuberculosis. Int J Tuberc Lung Dis Off J Int Union Tuberc Lung Dis. 2019;23: 450–457. doi:10.5588/ijtld.18.0551

160. Arinaminpathy N, Batra D, Maheshwari N, Swaroop K, Sharma L, Sachdeva KS, et al. Tuberculosis treatment in the private healthcare sector in India: an analysis of recent trends and volumes using drug sales data. BMC Infect Dis. 2019;19: 539. doi:10.1186/s12879-019-4169-y

161. Satyanarayana S, Nair SA, Chadha SS, Shivashankar R, Sharma G, Yadav S, et al. From where are tuberculosis patients accessing treatment in India? Results from a cross-sectional community based survey of 30 districts. PloS One. 2011;6: e24160. doi:10.1371/journal.pone.0024160

162. Daniels B, Kwan A, Satyanarayana S, Subbaraman R, Das RK, Das V, et al. Use of standardised patients to assess gender differences in quality of tuberculosis care in urban India: a two-city, cross-sectional study. Lancet Glob Health. 2019;7: e633–e643. doi:10.1016/S2214-109X(19)30031-2

163. Chikovore J, Pai M, Horton KC, Daftary A, Kumwenda MK, Hart G, et al. Missing men with tuberculosis: the need to address structural influences and implement targeted and multidimensional interventions. BMJ Glob Health. 2020;5: e002255. doi:10.1136/bmjgh-2019-002255

164. Horton KC, MacPherson P, Houben RMGJ, White RG, Corbett EL. Sex Differences in Tuberculosis Burden and Notifications in Low- and Middle-Income Countries: A Systematic Review and Meta-analysis. PLoS Med. 2016;13: e1002119. doi:10.1371/journal.pmed.1002119

165. Chadha VK, Anjinappa SM, Dave P, Rade K, Baskaran D, Narang P, et al. Sub-national TB prevalence surveys in India, 2006-2012: Results of uniformly conducted data analysis. PloS One. 2019;14: e0212264. doi:10.1371/journal.pone.0212264

166. Indian Council of Medical Research. National TB Prevalence Survey in India 2019-2021 Summary Report. New Delhi, India: Indian Council of Medical Research. Available: https://tbcindia.gov.in/showfile.php?lid=3659 [Accessed 23 December 2022]

167. Deepa D, Achanta S, Jaju J, Rao K, Samyukta R, Claassens M, et al. The Impact of Isoniazid Resistance on the Treatment Outcomes of Smear Positive Re-Treatment Tuberculosis Patients in the State of Andhra Pradesh, India. Neyrolles O, editor. PLoS ONE. 2013;8: e76189. doi:10.1371/journal.pone.0076189

168. Banerjee S, Bandyopadhyay K, Taraphdar P, Dasgupta A. Perceived discrimination among tuberculosis patients in an urban area of Kolkata City, India. J Glob Infect Dis. 2020;12: 144. doi:10.4103/jgid.jgid_146_19

169. Satyanarayana S, Nair SA, Chadha SS, Sharma G, Yadav S, Mohanty S, et al. Health-care seeking among people with cough of 2 weeks or more in India. Is passive TB case finding sufficient? Public Health Action. 2012;2: 157–161. doi:10.5588/pha.12.0019

170. Suganthi P, Chadha VK, Ahmed J, Umadevi G, Kumar P, Srivastava R, et al. Health seeking and knowledge about tuberculosis among persons with pulmonary symptoms and tuberculosis cases in Bangalore slums. Int J Tuberc Lung Dis Off J Int Union Tuberc Lung Dis. 2008;12: 1268–1273.

171. Oxlade O, Murray M. Tuberculosis and poverty: why are the poor at greater risk in India? PloS One. 2012;7: e47533. doi:10.1371/journal.pone.0047533

172. Subbaraman R, Thomas BE, Sellappan S, Suresh C, Jayabal L, Lincy S, et al. Tuberculosis patients in an Indian mega-city: Where do they live and where are they diagnosed? PloS One. 2017;12: e0183240. doi:10.1371/journal.pone.0183240

173. Ghosh R, Roy S, Rashid MK. Assessment of microbiological status after successful completion of intermittent revised national tuberculosis control programme directly observed treatment, short course regimen for microbiologically confirmed pulmonary tuberculosis cases: While new daily regimen going to be implemented in India. Indian J Med Microbiol. 2018;36: 251–256. doi:10.4103/ijmm.IJMM_18_65

174. Patel BH, Jeyashree K, Chinnakali P, Vijayageetha M, Mehta KG, Modi B, et al. Cash transfer scheme for people with tuberculosis treated by the National TB Programme in Western India: a mixed methods study. BMJ Open. 2019;9: e033158. doi:10.1136/bmjopen-2019-033158

175. Gopi PG, Chandrasekaran V, Subramani R, Santha T, Thomas A, Selvakumar N, et al. Association of conversion & cure with initial smear grading among new smear positive pulmonary tuberculosis patients treated with Category I regimen. INDIAN J MED RES. 2006; 8.

176. Vasantha M, Gopi PG, Subramani R. SURVIVAL OF TUBERCULOSIS PATIENTS TREATED UNDER DOTS IN A RURAL TUBERCULOSIS UNIT (TU), SOUTH INDIA. Indian J Tuberc. : 6.

177. Sinha P, Lakshminarayanan SL, Cintron C, Narasimhan PB, Locks LM, Kulatilaka N, et al. Nutritional Supplementation Would Be Cost-Effective for Reducing Tuberculosis Incidence and Mortality in India: The Ration Optimization to Impede Tuberculosis (ROTI-TB) Model. Clin Infect Dis Off Publ Infect Dis Soc Am. 2022;75: 577–585. doi:10.1093/cid/ciab1033

178. Daniels B, Shah D, Kwan AT, Das R, Das V, Puri V, et al. Tuberculosis diagnosis and management in the public versus private sector: a standardised patients study in Mumbai, India. BMJ Glob Health. 2022;7: e009657. doi:10.1136/bmjgh-2022-009657

179. Chaisson LH, Katamba A, Haguma P, Ochom E, Ayakaka I, Mugabe F, et al. Theory-Informed Interventions to Improve the Quality of Tuberculosis Evaluation at Ugandan Health Centers: A Quasi-Experimental Study. PloS One. 2015;10: e0132573. doi:10.1371/journal.pone.0132573

180. Fox WS, Strydom N, Imperial MZ, Jarlsberg L, Savic RM. Examining nonadherence in the treatment of tuberculosis: The patterns that lead to failure. Br J Clin Pharmacol. 2022. doi:10.1111/bcp.15515

181. Stagg HR, Thompson JA, Lipman MC, Sloan DJ, Flook M, Fielding KL, et al. Forgiveness Is the Attribute of the Strong: Nonadherence and Regimen-Shortening in Drug-Sensitive TB. Am J Respir Crit Care Med. 2022. doi:10.1164/rccm.202201-0144OC

182. Subbaraman R, Thomas BE, Kumar JV, Lubeck-Schricker M, Khandewale A, Thies W, et al. Measuring Tuberculosis Medication Adherence: A Comparison of Multiple Approaches in Relation to Urine Isoniazid Metabolite Testing Within a Cohort Study in India. Open Forum Infect Dis. 2021;8: ofab532. doi:10.1093/ofid/ofab532

183. Thomas BE, Kumar JV, Chiranjeevi M, Shah D, Khandewale A, Thiruvengadam K, et al. Evaluation of the Accuracy of 99DOTS, a Novel Cellphone-based Strategy for Monitoring Adherence to Tuberculosis Medications: Comparison of DigitalAdherence Data With Urine Isoniazid Testing. Clin Infect Dis Off Publ Infect Dis Soc Am. 2020;71: e513–e516. doi:10.1093/cid/ciaa333

184. Thomas BE, Kumar JV, Onongaya C, Bhatt SN, Galivanche A, Periyasamy M, et al. Explaining Differences in the Acceptability of 99DOTS, a Cell Phone-Based Strategy for Monitoring Adherence to Tuberculosis Medications: Qualitative Study of Patients and Health Care Providers. JMIR MHealth UHealth. 2020;8: e16634. doi:10.2196/16634

185. Thamineni R, Peraman R, Chenniah J, Meka G, Munagala AK, Mahalingam VT, et al. Level of adherence to anti-tubercular treatment among drug-sensitive tuberculosis patients on a newly introduced daily dose regimen in South India: A cross-sectional study. Trop Med Int Health TM IH. 2022;27: 1013–1023. doi:10.1111/tmi.13824

186. Natarajan S, Singla R, Singla N, Gupta A, Caminero JA, Chakraborty A, et al. Treatment interruption patterns and adverse events among patients on bedaquiline containing regimen under programmatic conditions in India. Pulmonology. 2022;28: 203–209. doi:10.1016/j.pulmoe.2020.09.006

187. Brownson RC, Shelton RC, Geng EH, Glasgow RE. Revisiting concepts of evidence in implementation science. Implement Sci IS. 2022;17: 26. doi:10.1186/s13012-022-01201-y

188. Mehrotra ML, Petersen ML, Geng EH. Understanding HIV Program Effects: A Structural Approach to Context Using the Transportability Framework. J Acquir Immune Defic Syndr 1999. 2019;82 Suppl 3: S199–S205. doi:10.1097/QAI.0000000000002202

189. Thomas BE, Suresh C, Lavanya J, Lindsley MM, Galivanche AT, Sellappan S, et al. Understanding pretreatment loss to follow-up of tuberculosis patients: an explanatory qualitative study in Chennai, India. BMJ Glob Health. 2020;5: e001974. doi:10.1136/bmjgh-2019-001974

190. Deshmukh RD, Dhande DJ, Sachdeva KS, Sreenivas A, Kumar AMV, Satyanarayana S, et al. Patient and Provider Reported Reasons for Lost to Follow Up in MDRTB Treatment: A Qualitative Study from a Drug Resistant TB Centre in India. PloS One. 2015;10: e0135802. doi:10.1371/journal.pone.0135802

191. Mukerji R, Turan JM. Challenges in accessing and utilising health services for women accessing DOTS TB services in Kolkata, India. Glob Public Health. 2020;15: 1718–1729. doi:10.1080/17441692.2020.1751235

192. Satyanarayana S, Kwan A, Daniels B, Subbaraman R, McDowell A, Bergkvist S, et al. Use of standardised patients to assess antibiotic dispensing for tuberculosis by pharmacies in urban India: a cross-sectional study. Lancet Infect Dis. 2016;16: 1261–1268. doi:10.1016/S1473-3099(16)30215-8

