## Supplemental Appendix 1 for "Barriers to engagement by people with active tuberculosis in the care cascade in India: a systematic review of two decades of quantitative research"

**Supplementary Appendix 1**

**Supplement to:**

Barriers to engagement by people with active tuberculosis in the care cascade in India: a systematic review of two decades of quantitative research

Tulip A. Jhaveri, Disha Jhaveri, Amith Galivanche, Dominic Voehler, Maya Lubeck-Schricker, Mei Chung, Pruthu Thekkur, Vineet Chadha, Ruvandhi Nathavitharana, Ajay M.V. Kumar, Hemant Deepak Shewade, Katherine Powers, Kenneth H. Mayer, Jessica E. Haberer, Paul Bain, Madhukar Pai, Srinath Satyanarayana, Ramnath Subbaraman

**Correspondence:**

Ramnath Subbaraman, MD, MSc, FACP

Tufts University School of Medicine

Department of Public Health and Community Medicine

136 Harrison Ave., MV237

Boston, MA 02130, USA

**Methods and findings for the systematic review of barriers to care seeking by individuals with confirmed or presumptive tuberculosis in the population in India (Gap 1)**

### Methods

##### Objectives

The objective of this systematic review was to understand why some individuals in the community with confirmed or presumptive tuberculosis (TB) do not seek care at health facilities (Gap 1 in the care cascade). We defined individuals with presumptive TB as people with cough >2 weeks or other symptoms that could be suggestive of active TB, as defined by the screening methodology used in each study. We included studies of individuals with presumptive TB because there are few robust studies of care seeking by individuals with confirmed TB in the general population. As such, individuals with presumptive TB are the closest surrogate for understanding the care seeking behavior of individuals with confirmed TB.

We searched for population-based surveys that identified individuals with confirmed or presumptive TB. Regarding confirmed TB, we focused on finding relevant TB prevalence surveys, since these surveys detect individuals with active TB in the population who have not been previously diagnosed. Included studies then evaluated these individuals’ preceding care-seeking behavior using questionnaires. Questionnaires usually captured demographic information and other characteristics of these individuals. Some asked additional questions to individuals who had not sought care to understand their reasons for not having visited health facilities.

We extracted two types of quantitative findings that help to understand care seeking behavior by individuals with confirmed or presumptive TB:

(a) *Factors associated with not having sought care in regression analyses*: For studies comparing individuals who had or had not sought care, we extracted effect estimates for independent variables (i.e., exposures or predictors) associated with not having sought care. Effect estimates included odds ratios, risk ratios, hazard ratios, or beta-coefficients, depending on the approach to analysis.
(b) *Reasons reported by individuals for not seeking care in quantitative surveys*: For studies that surveyed patients who had not sought care, we extracted the proportion of individuals who reported a given reason for not going to health facilities.

##### Search strategy

Three separate searches were conducted to identify articles. The first search was conducted as part of a previously published study quantifying gaps in India’s TB care cascade [1]. We used articles related to care seeking by individuals with confirmed or presumptive TB already identified in that prior systematic review. Full details of the search strategy and study selection process are described in that manuscript; however, we provide a summary here. For that review, a medical librarian searched PubMed, Embase, and Web of Science for studies published between January 1, 2000 and October 1, 2015, without language restrictions, using search terms and related variants for “tuberculosis”, “tuberculosis symptoms”, “India”, and “healthcare-seeking behavior” (Table A). In addition, we carried out electronic and hand searches of the following Indian journals that were not indexed for that entire time window: the Indian Journal of Tuberculosis, Lung India, the Indian Journal of Chest and Allied Sciences, the India Journal of Public Health, and the Indian Journal of Community Medicine. We screened all identified studies from this previous review for potential inclusion in our current systematic review; however, different data were extracted from studies that met our inclusion criteria.

###### Table A. Search strategy to identify manuscripts regarding the care-seeking behavior of individuals with confirmed or presumptive TB in India (Gap 1)

| Terms for tuberculosis: | “tuberculosis”[Mesh] OR *Mycobacterium tuberculosis*[tiab] OR TB[tiab] OR TB[tiab] OR MDRTB[tiab] OR XDRTB[tiab] |
| --- | --- |
| Terms for tuberculosis symptoms: | chest symptomatic[tiab] OR chest symptomatics[tiab] OR suspects[tiab] OR chest symptoms[tiab] OR pulmonary symptoms[tiab] OR cough[tiab] OR respiratory symptoms[tiab]; presumptive [tiab] |
| Terms for India: | “India”[Mesh] OR India[tiab] OR India[ad] OR Indian[tiab] OR Indians[tiab] |
| Terms for healthcare-seeking behavior: | “patient acceptance of health care”[Mesh] OR healthcare seeking behavior[tiab]* OR careseeking[tiab] OR health behavior[tiab] OR behavior[tiab] OR “Health Services/utilization”[Mesh] OR referral[tiab] OR behaviour[tiab]* OR health behaviour[tiab] OR behaviour[tiab] |

To update our review, we conducted a second refresher search using the same search terms for October 2, 2015 to October 1, 2019. We did not repeat hand searches of the Indian journals listed above, because all of these journals had been indexed in PubMed prior to the time period of this more recent search. Due to the extensive time required to extract data from the articles identified for this systematic review, we performed a third refresher search using the same search terms for October 2, 2019 to May 17, 2021. Finally, additional studies were identified by looking through the reference lists of the included primary studies and relevant review articles that were identified by the searches and by outreach to experts in the field (Fig A).

##### Inclusion and exclusion criteria

We applied the following criteria for inclusion and exclusion of studies for this systematic review.

*Inclusion criteria* included the following:

(1) Studies that identified individuals with confirmed or presumptive TB using population-based screening, such as household-level TB prevalence surveys or community active case-finding initiatives.
(2) Studies that assessed reasons that individuals identified in these population-based surveys had not sought care, using regression analyses to identify factors associated with not seeking care or questionnaires capturing reasons that patients had not sought care.

*Exclusion criteria* included the following:

(1) Studies that described the proportion of individuals with confirmed or presumptive TB who had not sought care without analyzing reasons why these patients may not have sought care.
(2) Health facility-based studies (i.e., individuals not identified at the population level).
(3) Studies with data collected prior to the year 2000, as India’s Revised National TB Control Programme (now called the National TB Elimination Programme) did not have nationwide coverage until the early 2000s.
(4) Studies only containing qualitative data evaluating why individuals with confirmed or presumptive TB do not seek care. Findings from studies containing qualitative data will be reported in a separate paper.

##### Study selection

Each citations identified by the search was independently assessed by at least two reviewers (authors TJ and DJ) for their eligibility at the title and abstract evaluation stage and again subsequently at the full text evaluation stage (Fig A). Disagreements were resolved by discussion between TJ and DJ or, if necessary, through consultation of a third reviewer (RS). Independent selection of articles at the title and abstract and full text stages was conducted using Covidence software (Veritas Health Innovations, Melbourne, Australia); however, quality assessment and extraction of study findings was conducted using an Excel spreadsheet.

Fig A. PRISMA flowchart: study selection for the systematic review of care seeking by people with confirmed or presumptive TB (Gap 1)

**Search 1: January 1, 2000 to October 1, 2015**

**Search 3: October 2, 2019 to May 17, 2021**

**Search 2: October 2, 2015 to October 1, 2019**

Reports identified from search of PubMed, Embase, and Web of Science and hand search of relevant journals (n=2,373), of which 694 were duplicates.

Duplicates removed before screening

(n = 86)

Records identified from search of PubMed, Embase, and Web of Science (n = 981)

Duplicate records removed before screening (n = 3)

Records identified from search of PubMed, Embase, and Web of Science (n = 2,406)

**Identification**

Records excluded after title and abstract screen

(n = 836)

Records excluded after title and abstract screen

(n = 2,374)

Records that underwent screening of title and abstract

(n = 895)

Records that underwent screening of title and abstract

(n = 2,403)

Total studies included in the systematic review (n = 15)

- Unique (n=14)
- Multiple gaps (n=1)

Reports shortlisted after removal of duplicates (n=1,679) of which 1,460 were excluded after title and abstract review.

Records not retrieved (n = 0)

Records for which full text articles were retrieved (n = 59)

Reports not retrieved

(n = 0)

Records for which full text articles were retrieved (n = 29)

**Screening**

Reports eligible for full text review (n=219), of which 211 were excluded.

Reports excluded after full text review

(n = 56), for the following reasons:

Wrong outcomes evaluated (n = 29)

Wrong patient population (n = 7)

Wrong study design or did not collect data on reasons for not seeking care

(n = 19)

Study protocol with no data (n=1)

Reports assessed for eligibility via full text review (n = 59)

Reports excluded after full text review (n = 28), for the following reasons:

Wrong outcome evaluated (n = 13)

Wrong patient population (n = 8)

Wrong study design or did not collect data on reasons for not seeking care (n = 7)

Reports assessed for eligibility via full text review (n = 29)

Studies included in previous care cascade study with relevant data on reasons for not seeking care (n = 8)

New studies included in the systematic review from the 2019 to 2021 search (n = 3)

New studies included in the systematic review from the 2015 to 2019 search (n = 1)

**Included**

Studies identified by outreach to experts (n = 2)

Studies identified by reviewing references of other articles (n = 1)

##### Quality assessment of quantitative studies

In our prior systematic review, we developed quality criteria relevant to population-based studies focused on identifying and interviewing individuals with presumptive TB (Table B), because there were no existing standardized guidelines for assessing the quality of these types of studies. Although we extracted different quantitative findings from these studies in the current systematic review, we used the same quality criteria because they are still appropriate for describing the overall methodological rigor and risk of bias in these observational studies.

We classified each population-based sampling strategy on whether it was comprehensive, random, or convenience. Studies using convenience sampling were rated as being of very low quality, and their findings were not included in the final analysis. One of the major challenges of population-based sampling is screening all members of a household, because many individuals, such as men, may be working and out of the house at times when researchers visit. Therefore, we also included the proportion of the estimated population screened and the proportion of identified individuals with confirmed or presumptive TB who were interviewed regarding care seeking as quality indicators.

###### Table B. Criteria for assessing quality of quantitative care-seeking studies

| **Criterion** | **Quality level** |
| --- | --- |
| **Sampling strategy** |  |
| Random (i.e., probability-based) or comprehensive sampling of community residents or households | High |
| Convenience sampling of community residents or households | Low (exclude from analysis of findings) |
| **Sample size** |  |
| Multiple villages, slum settlements, or communities with 1000+ residents screened | High |
| Single village, slum settlement or community with 1000+ residents screened | Medium |
| <1000 residents screened | Low |
| **Proportion of estimated population screened for TB symptoms** |  |
| 76-100% of screened | High |
| 50-75% of screened | Medium |
| <50% of screened or not reported | Low |
| **Proportion of identified as having TB symptoms who were interviewed about care seeking** |  |
| 76-100% interviewed | High |
| 50-75% interviewed | Medium |
| <50% interviewed or not reported | Low |

##### Data extraction and analysis

Two reviewers (TJ and DJ) independently extracted data from each included study into a structured form on an Excel spreadsheet. Disagreements were resolved by discussion or, if needed, by consulting a third reviewer (RS). We extracted information on each study’s design, location, setting (i.e., urban versus rural), sample size, and variables of interest (Table C).

For studies that compared individuals who sought care against those who had not sought care, we extracted adjusted and unadjusted effect estimates (odds ratios, risk ratios, hazard ratios, and beta-coefficients) from regression analyses. For studies that had not reported effect estimates, we calculated unadjusted odds ratios from the data provided, if possible. For studies that reported reasons why individuals had not sought care, we extracted the proportion of individuals surveyed who reported a given reason. For effect estimates and proportions, we extracted information on 95% confidence intervals (95% CIs) where available; if 95% CIs were not reported, we calculated these from the data provided, if possible.

For the regression analyses, some studies reported factors associated with having sought care, while others reported factors associated with not having sought care. For consistency, we “flipped” effect estimates and 95% CIs as needed so that all findings represented associations with the outcome of not having sought care. For some variables, we also changed the reference group as needed for consistency of reporting across studies. For example, because most studies compared men to the reference group of women, we “flipped” effect estimates and confidence intervals for studies that presented men as the reference group. This allowed us to consistently present women as the reference group for findings regarding gender to allow better understanding of common or disparate findings across studies.

After “flipping” selected effect estimates from the regression analyses, we reported all unadjusted and adjusted effect estimates, regardless of statistical significance, organized by study (Table D). For the main manuscript and Forest plot, we restricted ourselves to presenting statistically significant adjusted effect estimates from multivariable analyses, as these may represent more meaningful associations from higher-quality analyses. After extracting this subset of findings, we organized findings into categories using our framework of demand- and supply-side factors (main manuscript, Table 1). For findings on reasons why individuals had not sought care, we presented all reported proportions in the Forest Plots in the main manuscript; we also organized these findings using our framework of demand- and supply-side categories.

To visualize quantitative findings, we generated Forest plots of effect estimates (odds ratios, risk ratios, hazard ratios, beta-coefficients, and proportions) using Stata version 16.1 (College Station, TX, USA). We did not conduct meta-analyses of data, because we extracted findings represented a diverse set of variables from every study.

###### Table C. Characteristics of the included studies evaluating why patients with confirmed or presumptive tuberculosis (TB) in the population had not sought care (gap 1 in the TB care cascade)

| Citation  (year) | Location | Urban, rural, both, or unknown | Type of population | Number of community residents screened | % of estimated population screened  n (%) | Number of individuals with confirmed or presumptive TB identified | Number of individuals with confirmed or presumptive TB interviewed | % of individuals with confirmed or presumptive TB identified who were interviewed | Type of findings included in the study (N=sample or denominator of patients for given analysis) |
| --- | --- | --- | --- | --- | --- | --- | --- | --- | --- |
| Charles (2010) [2] | Chennai and Madurai districts, Tamil Nadu | Urban and rural | General | 18,417 | 100% | 640 | 606 (all with presumptive TB) | 95% | Logistic regression (N=606)  Reasons for not seeking care (proportions) (N=162) |
| Fochsen (2006) [3] | Ujjain district, Madhya Pradesh | Rural | General | 45,719 | 100% | 941 | 644 (all with presumptive TB) | 68%^a^ | Logistic regression  (N=644) |
| George (2013, Uttar Pradesh data) [4] | Two districts in Uttar Pradesh | Urban | Urban informal settlements | 68,324 | NR^a^ | NR | 1,526 (all with presumptive TB) | NR^a^ | Logistic regression  (N=1288)  Reasons for not seeking care (proportions) (N=381) |
| George (2013, Karnataka data) [4] | Four districts in Karnataka | Urban | Urban informal settlements | 49,279 | NR^a^ | NR | 1,515 (all with presumptive TB) | NR^a^ | Logistic regression  (N=1506)  Reasons for not seeking care (proportions) (N=636) |
| Ghosh (2010) [5] | Bankura district, West Bengal | Urban | An urban informal settlement* | 1,156* | NR^a^ | 64 | 64 (all with presumptive TB) | 100% | Logistic regression  (N=64)  Reasons for not seeking care (proportions) (N=64) |
| Helfinstein (2020) [6] | Chennai, Tamil Nadu | Urban | Community and slum-based | 135,001 | NR^a^ | 1,922 | 1,667 (all with presumptive TB) | 87% | Logistic regression – Tamil Nadu entire sample (N=1,667); sub-sample from Tamil Nadu slums (N=1,134) |
| Kar (2010) [7] | Kancheepuram, Tamil Nadu | Rural | General | 1,985 | NR^a^ | 65 | 65 (all with presumptive TB) | 100% | Reasons for not seeking care (proportions) (N=65) |
| Karanjekar (2014) [8] | Aurangabad, Maharashtra | Urban | Urban slums | 8,970 | 99% | 105 | 105 (all with presumptive TB) | 100% | Logistic regression  (N=105)  Reasons for not seeking care (proportions) (N=56) |
| KHPT/THALI project (2016-17)^b^ [9] | Hyderabad, Telangana | Urban | Urban slums | 2,007 | NR^a^ | 480 | 427 (all with presumptive TB) | 89% | Logistic regression  (N=427) |
| KHPT/THALI project (2016-17)^b^ [10] | Bengaluru, Karnataka | Urban | Urban slums | 1,909 | NR^a^ | 457 | 413 (all with presumptive TB) | 90% | Logistic regression  (N=413) |
| Satyanarayana (2012) [11] | Thirty districts in four in north, south, east, and west regions of India | Urban and rural | General | 4,562 | NR^a^ | 437 | 437 (all with presumptive TB) | 100% | Logistic regression  (N=437) |
| Shewade (2019)^b^ [12] | Eighteen districts (7 states) throughout India | Urban and Rural | General | NR* | NR^a^ | 573 | 465 (all with confirmed TB) | 81% | Logistic regression (N=465) |
| Shriraam (2020) [13] | Tiruvallur district, Tamil Nadu | Rural | Villages with a large population of brick kiln workers | 650^a^ | 89% | 56 | 56 (all with presumptive TB) | 100% | Reasons for not seeking care (proportions) (N=31) |
| Suganthi (2008) [14] | Bangalore, Karnataka | Urban | Urban informal settlements | 9,676 | 83% | 166 | 124 (all with presumptive TB) | 75%^a^ | Reasons for not seeking care (proportions) (N=63) |
| Thomas (2015) [15] | Tiruvallur district, Tamil Nadu | Rural | Villages with a large population of brick kiln workers | 4,002 | NR^a^ | 377 | 377 (all with presumptive TB) | 100% | Logistic regression  (N=377) |
| Thomas (2021) [16] | 17 states throughout India | Rural | Districts in which >70% of the population was “tribal” (Adivasi) | 74,532 | 81% | 2,675 | 2,675 (all with presumptive TB) | 100% | Logistic regression  (N=2,675)  Reasons for not seeking care (proportions) (N=2,016) |

KHPT=Karnataka Health Promotion Trust; NR=Not reported; THALI=Tuberculosis Health Action Learning Initiative.

^a^Medium or low quality for this indicator

^b^Secondary analysis of data from a previously published study, conducted by the study authors at the request of the systematic review team.

###### Table D. Factors associated with individuals with confirmed or presumptive tuberculosis in the population in India not seeking care

| Study | Exposure / independent variable | Unadjusted odds ratio  (95% confidence Interval) | p-value | Adjusted odds ratio  (95% confidence interval) | p-value |
| --- | --- | --- | --- | --- | --- |
| Charles 2010  (Tamil Nadu)^a,d^ [2] |  |  |  |  |  |
|  | **Sex** |  |  |  |  |
|  | Male | Ref |  | Not reported |  |
|  | Female | 0.89 (0.62—1.27) | 0.51 | Not reported |  |
|  | **Age in years** |  |  |  |  |
|  | <45 | Ref |  | Not reported |  |
|  | >=45 | 0.64 (0.44—0.92)* | 0.017* | Not reported |  |
|  | **Area** |  |  |  |  |
|  | Urban | Ref |  | Not reported |  |
|  | Rural | 0.84 (0.59—1.21) | 0.35 | Not reported |  |
|  | **Literacy** |  |  |  |  |
|  | Literate | Ref |  | Not reported |  |
|  | Illiterate | 0.56 (0.38—0.84)* | 0.005* | Not reported |  |
|  | **Employed** |  |  |  |  |
|  | Yes | Ref |  | Not reported |  |
|  | No | 0.84 (0.59—1.21) | 0.33 | Not reported |  |
|  | **Income (Rs/month)** |  |  |  |  |
|  | <=2000 | Ref |  | Not reported |  |
|  | >2000 | 0.81 (0.47—1.42) | 0.47 | Not reported |  |
| Fochsen 2006 (Madhya Pradesh)^a,e^ [3] |  |  |  |  |  |
|  | **History of TB**^b^ |  |  |  |  |
|  | Yes | Not reported |  | Ref |  |
|  | No | Not reported |  | 2.70 (1.41—5.15)* | Not reported |
|  | **Decreasing duration of cough**^b,c^ |  |  |  |  |
|  | Per each week decrease in cough duration | Not reported |  | 1.05 (1.03—1.07)* | Not reported |
|  | **Fever**^b^ |  |  |  |  |
|  | Yes | Not reported |  | Ref |  |
|  | No | Not reported |  | 1.80 (1.20—2.70)* | Not reported |
| George 2013  (Uttar Pradesh)^a^ [4] |  |  |  |  |  |
|  | **Sex** |  |  |  |  |
|  | Female | Not reported |  | Ref |  |
|  | Male | Not reported |  | 1.75 (1.28—2.40)* | Not reported |
|  | **Age in years** |  |  |  |  |
|  | <=45 | Not reported |  | Ref |  |
|  | 46-65 | Not reported |  | 0.80 (0.57—1.12) | Not reported |
|  | >65 | Not reported |  | 0.93 (0.70—1.22) | Not reported |
|  | **Number of adults in family** |  |  |  |  |
|  | <=3 | Not reported |  | Ref |  |
|  | 4-5 | Not reported |  | 1.20 (0.86—1.70) | Not reported |
|  | >5 | Not reported |  | 0.35 (0.24—0.53)* | Not reported |
|  | **Education** |  |  |  |  |
|  | Illiterate | Not reported |  | Ref |  |
|  | Below primary | Not reported |  | 1.3 (0.76—2.21) | Not reported |
|  | Primary & middle | Not reported |  | 1.15 (0.81—1.63) | Not reported |
|  | Secondary & above | Not reported |  | 1.08 (0.70—1.64) | Not reported |
|  | **Occupation** |  |  |  |  |
|  | Unemployed | Not reported |  | Ref |  |
|  | Unskilled workers | Not reported |  | 1.09 (0.74—1.59) | Not reported |
|  | Skilled worker | Not reported |  | 1.30 (0.91—1.85) | Not reported |
|  | **Socioeconomic class**^b^ |  |  |  |  |
|  | Low vs. medium | Not reported |  | 0.79 (0.58—1.09) | Not reported |
|  | Low vs. high | Not reported |  | 2.24 (1.16—4.33)* | Not reported |
|  | **Knows TB caused by germs**^b^ |  |  |  |  |
|  | Yes | Not reported |  | Ref |  |
|  | No | Not reported |  | 5.89 (3.48—9.96)* | Not reported |
|  | **Knows TB diagnosis can be confirmed by a sputum test**^b^ |  |  |  |  |
|  | Yes | Not reported |  | Ref |  |
|  | No | Not reported |  | 0.56 (0.33—1.12) | Not reported |
|  | **Knows TB spread through air**^b^ |  |  |  |  |
|  | Yes | Not reported |  | Ref |  |
|  | No | Not reported |  | 0.41 (0.21—1.09) | Not reported |
|  | **Knows TB is curable**^b^ |  |  |  |  |
|  | Yes | Not reported |  | Ref |  |
|  | No | Not reported |  | 0.74 (0.38—1.44) | Not reported |
|  | **Knows sputum test as a test for TB diagnosis**^b^ |  |  |  |  |
|  | Yes | Not reported |  | Ref |  |
|  | No | Not reported |  | 2.11 (0.97—4.59) | Not reported |
|  | **Knows persistent cough suggests TB**^b^ |  |  |  |  |
|  | Yes | Not reported |  | Ref |  |
|  | No | Not reported |  | 5.36 (1.89—15.19)* | Not reported |
|  | **Knows TB can affect anybody**^b^ |  |  |  |  |
|  | Yes | Not reported |  | Ref |  |
|  | No | Not reported |  | 1.27 (1.09—1.47)* | Not reported |
|  | **Thinks TB spreads by physical contact, including sharing items, talking with someone, and having sex**^b^ |  |  |  |  |
|  | No | Not reported |  | Ref |  |
|  | Yes | Not reported |  | 1.42 (1.16—1.75)* | Not reported |
|  | **Thinks TB can be confirmed by a blood test**^b^ |  |  |  |  |
|  | No | Not reported |  | Ref |  |
|  | Yes | Not reported |  | 1.19 (1.09—1.29)* | Not reported |
|  | **Negative perceived quality of care** (i.e., unfavorable view of local quality of health care)^b^ |  |  |  |  |
|  | No | Not reported |  | Ref |  |
|  | Yes | Not reported |  | 0.94 (0.66—1.34) | Not reported |
|  | **Lacks self-efficacy in seeking care**^b^ |  |  |  |  |
|  | No | Not reported |  | Ref |  |
|  | Yes | Not reported |  | 1.89 (1.33—2.67)* | Not reported |
|  | **Knows TB is life threatening**^b^ |  |  |  |  |
|  | Yes | Not reported |  | Ref |  |
|  | No | Not reported |  | 1.05 (0.83—1.32) | Not reported |
| George (2013) (Karnataka)^a^ [4] |  |  |  |  |  |
|  | **Sex** |  |  |  |  |
|  | Female | Not reported |  | Ref |  |
|  | Male | Not reported |  | 1.45 (1.01—2.08)* | Not reported |
|  | **Age in years** |  |  |  |  |
|  | <=45 | Not reported |  | Ref |  |
|  | 46-65 | Not reported |  | 0.93 (0.63—1.4) | Not reported |
|  | >65 | Not reported |  | 0.80 (0.60—1.06) | Not reported |
|  | **Number of adults in family** |  |  |  |  |
|  | <=3 | Not reported |  | Ref |  |
|  | 4-5 | Not reported |  | 0.66 (0.46—0.95)* | Not reported |
|  | >5 | Not reported |  | 0.87 (0.56—1.34) | Not reported |
|  | **Education** |  |  |  |  |
|  | Illiterate | Not reported |  | Ref |  |
|  | Below primary | Not reported |  | 0.67 (0.38—1.18) | Not reported |
|  | Primary & middle | Not reported |  | 1.27 (0.81—1.97) | Not reported |
|  | Secondary & above | Not reported |  | 0.7 (0.41—1.19) | Not reported |
|  | **Occupation** |  |  |  |  |
|  | Unemployed | Not reported |  | Ref |  |
|  | Unskilled workers | Not reported |  | 0.85 (0.51—1.40) | Not reported |
|  | Skilled worker | Not reported |  | 1.23 (0.70—2.18) | Not reported |
|  | **Socioeconomic class**^b^ |  |  |  |  |
|  | Low vs. medium | Not reported |  | 1.50 (1.01—2.24)* | Not reported |
|  | Low vs. high | Not reported |  | 1.58 (0.85—2.93) | Not reported |
|  | **Knows TB caused by germs**^b^ |  |  |  |  |
|  | Yes | Not reported |  | Ref |  |
|  | No | Not reported |  | 1.16 (0.74—1.82) | Not reported |
|  | **Knows TB diagnosis can be confirmed by a sputum test**^b^ |  |  |  |  |
|  | Yes | Not reported |  | Ref |  |
|  | No | Not reported |  | 1.53 (1.54—3.75)* | Not reported |
|  | **Knows TB spread through air**^b^ |  |  |  |  |
|  | Yes | Not reported |  | Ref |  |
|  | No | Not reported |  | 1.67 (1.13—2.46)* | Not reported |
|  | **Knows TB is curable**^b^ |  |  |  |  |
|  | Yes | Not reported |  | Ref |  |
|  | No | Not reported |  | 1.65 (1.12—2.43)* | Not reported |
|  | **Knows sputum test as a test for TB diagnosis**^b^ |  |  |  |  |
|  | Yes | Not reported |  | Ref |  |
|  | No | Not reported |  | 2.40 (1.54—3.75)* | Not reported |
|  | **Knows persistent cough suggests TB**^b^ |  |  |  |  |
|  | Yes | Not reported |  | Ref |  |
|  | No | Not reported |  | 1.81 (1.13—2.90)* | Not reported |
|  | **Knows TB can affect anybody**^b^ |  |  |  |  |
|  | Yes | Not reported |  | Ref |  |
|  | No | Not reported |  | 0.96 (0.75—1.23) | Not reported |
|  | **Thinks TB spreads by physical contact, specifically, sharing items, talking with someone, and having sex**^b^ |  |  |  |  |
|  | No | Not reported |  | Ref |  |
|  | Yes | Not reported |  | 0.90 (0.70—1.15) | Not reported |
|  | **Thinks TB can be confirmed by blood test**^b^ |  |  |  |  |
|  | No | Not reported |  | Ref |  |
|  | Yes | Not reported |  | 1.00 (0.77—1.30) | Not reported |
|  | **Negative perceived quality of care** (i.e., unfavorable view of local quality of health care)^b^ |  |  |  |  |
|  | No | Not reported |  | Ref |  |
|  | Yes | Not reported |  | 2.99 (2.06—4.40)* | Not reported |
|  | **Lacks self-efficacy in seeking care**^b^ |  |  |  |  |
|  | No | Not reported |  | Ref |  |
|  | Yes | Not reported |  | 1.84 (1.38—2.46)* | Not reported |
|  | **Knows TB is life threatening**^b^ |  |  |  |  |
|  | Yes | Not reported |  | Ref |  |
|  | No | Not reported |  | 0.65 (0.51—0.83)* | Not reported |
| Ghosh 2010  (West Bengal)^d^ [5] |  |  |  |  |  |
|  | **Sex** |  |  |  |  |
|  | Male | Ref |  |  |  |
|  | Female | 1.46 (0.4—4.8) | 0.53 | Not reported |  |
|  | **Age in years** |  |  |  |  |
|  | 15-29 | Ref |  | Not reported |  |
|  | 30-44 | 1.86 (0.5—6.8) | 0.35 | Not reported |  |
|  | >=45 | 2.17 (0.5—9.6) | 0.31 | Not reported |  |
|  | **Socioeconomic class** |  |  |  |  |
|  | Lower middle | Ref |  | Not reported |  |
|  | Upper lower | 1.88 (0.5—7.0) | 0.35 | Not reported |  |
|  | Upper middle | 1.5 (0.22—10.36) | 0.68 | Not reported |  |
| Helfinstein 2020 (Tamil Nadu, subset of slum households)^a^ [6] |  |  |  |  |  |
|  | **Increasing age in years**^b,c^ |  |  |  |  |
|  | Per every standard deviation increase in age | Not reported |  | 0.82 (0.70—0.96)* | 0.013* |
|  | **Caste** |  |  |  |  |
|  | Refused | Not reported |  | Ref |  |
|  | Scheduled caste or scheduled tribe | Not reported |  | 0.57 (0.23—1.41) | 0.219 |
|  | Other backward caste | Not reported |  | 0.74 (0.3—1.83) | 0.502 |
|  | None of the above | Not reported |  | 0.76 (0.3—1.91) | 0.555 |
|  | **Religion** |  |  |  |  |
|  | Christian | Not reported |  | Ref |  |
|  | Muslim/other | Not reported |  | 0.7 (0.39—1.26) | 0.238 |
|  | Hindu | Not reported |  | 0.96 (0.66—1.41) | 0.851 |
|  | **Literacy** |  |  |  |  |
|  | Illiterate | Not reported |  | Ref |  |
|  | Literate | Not reported |  | 0.77 (0.55—1.08) | 0.121 |
|  | **Decreasing years of education**^b,c^ |  |  |  |  |
|  | Per every standard deviation decrease in years of education | Not reported |  | 1.14 (0.97—1.35) | 0.110 |
|  | **Job** |  |  |  |  |
|  | Homemaker | Not reported |  | Ref |  |
|  | Retired/unemployed/disabled | Not reported |  | 1.04 (0.66—1.65) | 0.876 |
|  | Other | Not reported |  | 1.45 (0.90—2.33) | 0.122 |
|  | Laborer | Not reported |  | 1.64 (1.11—2.43)* | 0.012* |
|  | Professional | Not reported |  | 1.67 (1.03—2.70)* | 0.035* |
|  | **Decreasing wealth score**^b,c^ |  |  |  |  |
|  | Per every standard deviation decrease in wealth score | Not reported |  | 0.94 (0.83—1.07) | 0.358 |
|  | **Decreasing symptom days**^b,c^ |  |  |  |  |
|  | Per every standard deviation decrease in symptom days | Not reported |  | 1.59 (1.37—1.85)* | <0.001* |
|  | **Decreasing symptom count**^b,c^ |  |  |  |  |
|  | Per every standard deviation decrease in symptom count | Not reported |  | 1.28 (1.12—1.48)* | <0.001* |
|  | **Decreasing symptom recurrence score**^b,c^  (decreasing frequency of symptoms) |  |  |  |  |
|  | Per every standard deviation decrease in the frequency of symptoms | Not reported |  | 0.87 (0.76—0.99)* | 0.029* |
| Helfinstein 2020 (Tamil Nadu, all households surveyed)^a^ [6] |  |  |  |  |  |
|  | **Sex** |  |  |  |  |
|  | Female | Not reported |  | Ref |  |
|  | Male | Not reported |  | 0.58 (0.41—0.82)* | 0.002* |
|  | **Increasing age in years**^b,c^ |  |  |  |  |
|  | Per every standard deviation increase in age | Not reported |  | 0.83 (0.72—0.96)* | 0.012* |
|  | **Caste** |  |  |  |  |
|  | Refused | Not reported |  | Ref |  |
|  | Scheduled caste or scheduled tribe | Not reported |  | 0.85 (0.5—1.46) | 0.572 |
|  | Other backward caste | Not reported |  | 1.05 (0.61—1.81) | 0.858 |
|  | None of the above | Not reported |  | 0.87 (0.5—1.52) | 0.618 |
|  | **Religion** |  |  |  |  |
|  | Christian | Not reported |  | Ref |  |
|  | Muslim/other | Not reported |  | 0.99 (0.61—1.6) | 0.699 |
|  | Hindu | Not reported |  | 0.87 (0.63—1.20) | 0.387 |
|  | **Sample** |  |  |  |  |
|  | Slum-based | Not reported |  | Ref |  |
|  | Original Chennai-representative | Not reported |  | 1.35 (1.07—1.71)* | 0.010* |
|  | **Literacy** |  |  |  |  |
|  | Illiterate | Not reported |  | Ref |  |
|  | Literate | Not reported |  | 0.88 (0.67—1.16) | 0.343 |
|  | **Decreasing years of education**^b,c^ |  |  |  |  |
|  | Per each standard deviation decrease in years of education | Not reported |  | 0.99 (0.86—1.14) | 0.851 |
|  | **Job** |  |  |  |  |
|  | Homemaker | Not reported |  | Ref |  |
|  | Retired, unemployed, or disabled | Not reported |  | 0.78 (0.52—1.15) | 0.202 |
|  | Other | Not reported |  | 1.35 (0.92—1.98) | 0.130 |
|  | Laborer | Not reported |  | 1.35 (0.97—1.88) | 0.072 |
|  | Professional | Not reported |  | 1.72 (1.15—2.58)* | 0.008* |
|  | **Decreasing wealth score**^b,c^ |  |  |  |  |
|  | Per each standard deviation decrease in score | Not reported |  | 0.99 (0.88—1.10) | 0.831 |
|  | **Decreasing worry about healthcare cost**^b,c^ |  |  |  |  |
|  | Per each standard deviation decrease in worry | Not reported |  | 1.07 (0.94—1.21) | 0.313 |
|  | **Decreasing general financial stress**^b,c^ |  |  |  |  |
|  | Per each standard deviation decrease in financial stress | Not reported |  | 1.01 (0.88—1.15) | 0.917 |
|  | **Pre-existing illness (comorbidity)** |  |  |  |  |
|  | Yes | Not reported |  | Ref |  |
|  | No | Not reported |  | 1.28 (1.02—1.61)* | 0.035* |
|  | **Value good health** (the extent to which the individual places value on being healthy)^b,c^ |  |  |  |  |
|  | Per each standard deviation decrease in personal valuing of good health | Not reported |  | 1.19 (1.07—1.33)* | 0.001* |
|  | **Facility preference** |  |  |  |  |
|  | Government | Not reported |  | Ref |  |
|  | Private | Not reported |  | 0.85 (0.66—1.08) | 0.197 |
|  | No difference (i.e., no preference) | Not reported |  | 1.52 (1.11—2.08)* | 0.01* |
|  | **Facility preference mismatch with community preference** (i.e., personal preference for government or private facility is at odds with what family or community members would recommend) |  |  |  |  |
|  | No | Not reported |  | Ref |  |
|  | Yes | Not reported |  | 1.85 (1.37—2.50)* | <0.001* |
|  | **Smoker** |  |  |  |  |
|  | No | Not reported |  | Ref |  |
|  | Yes | Not reported |  | 1.67 (1.14—2.44)* | 0.007* |
|  | **Problem Drinker** |  |  |  |  |
|  | No | Not reported |  | Ref |  |
|  | Yes | Not reported |  | 1.89 (1.25—2.85)* | 0.002* |
|  | **Hours of sleep** |  |  |  |  |
|  | <6 | Not reported |  | Ref |  |
|  | 6+ | Not reported |  | 1.28 (1.03—1.60)* | 0.029* |
|  | **Exercise** |  |  |  |  |
|  | <1 day weekly | Not reported |  | Ref |  |
|  | 1+ days weekly | Not reported |  | 1.45 (1.09—1.93)* | 0.012* |
|  | **Gets health information from doctors**^b^ |  |  |  |  |
|  | Yes | Not reported |  | Ref |  |
|  | No | Not reported |  | 1.43 (1.02—2.01)* | 0.03* |
|  | **Gets health information from the government**^b^ |  |  |  |  |
|  | Yes | Not reported |  | Ref |  |
|  | No | Not reported |  | 1.16 (0.91—1.49) | 0.23 |
|  | **Other disease knowledge** (i.e., knowledge of malnutrition, HIV/AIDS, pneumonia, malaria, or typhoid)^b,c^ |  |  |  |  |
|  | Per each standard deviation decrease in knowledge | Not reported |  | 1.12 (0.98—1.28) | 0.111 |
|  | **Gets health information from the community**^b^ |  |  |  |  |
|  | Yes | Not reported |  | Ref |  |
|  | No | Not reported |  | 0.94 (0.83—1.07) | 0.351 |
|  | **Gets health information from the media**^b^ |  |  |  |  |
|  | Yes | Not reported |  | Ref |  |
|  | No | Not reported |  | 0.89 (0.78—1.01) | 0.062 |
|  | **Aware of TB** (i.e., individual has ever heard of TB) |  |  |  |  |
|  | No | Not reported |  | Ref |  |
|  | Yes | Not reported |  | 1.02 (0.71—1.47) | 0.919 |
|  | **Family push to seek care** |  |  |  |  |
|  | No | Not reported |  | Ref |  |
|  | Yes | Not reported |  | 1.06 (0.95—1.20) | 0.343 |
|  | **Relative with pre-existing illness** |  |  |  |  |
|  | No | Not reported |  | Ref |  |
|  | Yes | Not reported |  | 0.93 (0.73—1.18) | 0.510 |
|  | **Level of perceived family/community care when sick**^b,c^ |  |  |  |  |
|  | Per each standard deviation decrease in perception of care | Not reported |  | 1.07 (0.95—1.22) | 0.282 |
|  | **Decreasing worry about transportation cost**^b,c^ |  |  |  |  |
|  | Per each standard deviation decrease in worry | Not reported |  | 1.17 (1.02—1.34)* | 0.021* |
|  | **Perceived number of people who help with obligations**^b,c^ |  |  |  |  |
|  | Per each standard deviation decrease in number of perceive people who help | Not reported |  | 1.03 (0.92—1.16) | 0.574 |
|  | **Tendency to delay gratification**^b,c^ |  |  |  |  |
|  | Per each standard deviation decrease in tendency to delay gratification | Not reported |  | 0.95 (0.85—1.06) | 0.341 |
| Karanjekar 2014 (Maharashtra)^d^ [8] |  |  |  |  |  |
|  | **Sex** |  |  |  |  |
|  | Female | Ref |  |  |  |
|  | Male | 1.03 (0.42—2.53) | 0.95 | Not reported |  |
|  | **Age in years** |  |  |  |  |
|  | 15-34 | Ref |  | Not reported |  |
|  | 35-54 | 1.55 (0.58—4.16) | 0.39 | Not reported |  |
|  | 55-74 | 1.38 (0.36—5.28) | 0.64 | Not reported |  |
|  | >74 | 1.27 (0.12—13.52) | 0.85 | Not reported |  |
|  | **Religion** |  |  |  |  |
|  | Hindu | Ref |  | Not reported |  |
|  | Muslim | 1.13 (0.26—4.93) | 0.87 | Not reported |  |
|  | Buddhist | 10.2 (2.95—35.32)* | 0.0002* | Not reported |  |
|  | **Marital status** |  |  |  |  |
|  | Married | Ref |  | Not reported |  |
|  | Unmarried | 2.46 (0.94—6.44) | 0.07 | Not reported |  |
|  | **Type of family** |  |  |  |  |
|  | Nuclear | Ref |  | Not reported |  |
|  | Joint | 0.93 (0.36—2.39) | 0.87 | Not reported |  |
|  | Three generation | 0.46 (0.05—4.39) | 0.5 | Not reported |  |
|  | **Education** |  |  |  |  |
|  | Literate | Ref |  | Not reported |  |
|  | Illiterate | 30.7 (8.98—105.04)* | <0.0001* | Not reported |  |
|  | **Socioeconomic class** |  |  |  |  |
|  | I, II, III | Ref |  | Not reported |  |
|  | IV, V | 2.05 (0.77—5.43) | 0.15 | Not reported |  |
| KHPT/THALI project 2016-17 (Telangana)^f^ [9] |  |  |  |  |  |
|  | **Sex** |  |  |  |  |
|  | Female | Not reported |  | Ref |  |
|  | Male | Not reported |  | 1.15 (0.54—2.42) | 0.718 |
|  | **Age in years** |  |  |  |  |
|  | <40 | Not reported |  | Ref |  |
|  | 40-59 | Not reported |  | 1.58 (0.79—3.16) | 0.194 |
|  | 60+ | Not reported |  | 1.28 (0.52—3.12) | 0.583 |
|  | **Marital status** |  |  |  |  |
|  | Currently married | Not reported |  | Ref |  |
|  | Marriage annulled | Not reported |  | 0.81 (0.43—1.53) | 0.510 |
|  | Never married | Not reported |  | 1.34 (0.56—3.21) | 0.511 |
|  | **Religion** |  |  |  |  |
|  | Hindu | Not reported |  | Ref |  |
|  | Non-Hindu | Not reported |  | 1.89 (0.97—3.68) | 0.06 |
|  | **Caste or tribe** |  |  |  |  |
|  | Scheduled caste or scheduled tribe | Not reported |  | Ref |  |
|  | Non-scheduled caste or scheduled tribe | Not reported |  | 0.79 (0.42—1.49) | 0.462 |
|  | **Literacy and education**^b^ |  |  |  |  |
|  | Illiterate vs. primary completed | Not reported |  | 0.46 (0.24—0.88)* | 0.020* |
|  | Illiterate vs. middle school completed | Not reported |  | 0.42 (0.21—0.85)* | 0.017* |
|  | **Occupation** |  |  |  |  |
|  | No regular income | Not reported |  | Ref |  |
|  | Regular income | Not reported |  | 1.20 (0.57—2.51) | 0.629 |
|  | Irregular income | Not reported |  | 1.50 (0.65—3.46) | 0.338 |
|  | **Personal monthly income** |  |  |  |  |
|  | <5000 | Not reported |  | Ref |  |
|  | 5000 | Not reported |  | 1.05 (0.52—2.15) | 0.885 |
|  | **Household monthly income** |  |  |  |  |
|  | <5000 | Not reported |  | Ref |  |
|  | 5000+ | Not reported |  | 1.10 (0.70—1.75) | 0.673 |
|  | **Has comprehensive TB knowledge**^b^ |  |  |  |  |
|  | Yes | Not reported |  | Ref |  |
|  | No | Not reported |  | 2.44 (1.48—4.02)* | 0.001* |
| KHPT/THALI project 2016-17 (Karnataka)^f^ [10] |  |  |  |  |  |
|  | **Sex** |  |  |  |  |
|  | Female | Not reported |  | Ref |  |
|  | Male | Not reported |  | 0.81 (0.50—1.30) | 0.377 |
|  | **Age in years** |  |  |  |  |
|  | <40 | Not reported |  | Ref |  |
|  | 40-59 | Not reported |  | 0.73 (0.40—1.31) | 0.288 |
|  | 60+ | Not reported |  | 0.58 (0.28—1.17) | 0.126 |
|  | **Marital status** |  |  |  |  |
|  | Currently married | Not reported |  | Ref |  |
|  | Marriage annulled | Not reported |  | 1.11 (0.55—2.24) | 0.76 |
|  | Never married | Not reported |  | 0.86 (0.42—1.75) | 0.668 |
|  | **Religion** |  |  |  |  |
|  | Hindu | Not reported |  | Ref |  |
|  | Non-Hindu | Not reported |  | 1.24 (0.64—2.41) | 0.518 |
|  | **Caste/Tribe** |  |  |  |  |
|  | Scheduled caste or scheduled tribe | Not reported |  | Ref |  |
|  | Non-scheduled caste or scheduled tribe | Not reported |  | 1.40 (0.69—2.81) | 0.346 |
|  | **Literacy and Education** |  |  |  |  |
|  | Illiterate | Not reported |  | Ref |  |
|  | Primary & middle school incomplete | Not reported |  | 0.82 (0.45—1.50) | 0.512 |
|  | Middle school complete | Not reported |  | 1.10 (0.60—2.01) | 0.762 |
|  | **Occupation** |  |  |  |  |
|  | No regular income | Not reported |  | Ref |  |
|  | Regular income | Not reported |  | 1.27 (0.49—3.27) | 0.617 |
|  | Irregular income | Not reported |  | 1.02 (0.51—2.02) | 0.965 |
|  | **Personal monthly income** |  |  |  |  |
|  | <5000 | Not reported |  | Ref |  |
|  | 5000 | Not reported |  | 1.28 (0.56—2.90) | 0.556 |
|  | **Household monthly income**^b^ |  |  |  |  |
|  | 15000+ | Not reported |  | Ref |  |
|  | <15000 | Not reported |  | 2.22 (1.27—3.88)* | 0.006* |
|  | **Has comprehensive TB knowledge** |  |  |  |  |
|  | No | Not reported |  | Ref |  |
|  | Yes | Not reported |  | 0.86 (0.51—1.46) | 0.58 |
| Satyanarayana 2012  (Multi-site, 30 Indian districts) [11] |  |  |  |  |  |
|  | **Sex** |  |  |  |  |
|  | Male | Ref |  | Ref |  |
|  | Female | 1.2 (0.7—2.0) |  | 1.0 (0.6—1.6) | 0.88 |
|  | **Age in years** |  |  |  |  |
|  | 18-24 | Ref |  | Ref |  |
|  | 25-34 | 1.4 (0.7—3.0) |  | 0.9 (0.4—2.0) | 0.94 |
|  | 35-44 | 1.2 (0.6—2.4) |  | 0.8 (0.4—1.6) | 0.57 |
|  | 45-54 | 0.9 (0.4—1.9) |  | 0.5 (0.2—1.1) | 0.12 |
|  | >=55 | 0.5 (0.2—1.4) |  | 0.4 (0.1—1.1) | 0.09 |
|  | **Residence** |  |  |  |  |
|  | Urban | Ref |  | Ref |  |
|  | Rural | 2.4 (1.1—5.2)* |  | 1.70 (1.05—2.9)* | 0.03* |
|  | **Zone of the country** |  |  |  |  |
|  | South | Ref |  | Ref |  |
|  | North | 3.4 (1.7—6.6)* |  | 2.50 (1.30—4.80)* | <0.01* |
|  | East | 5.2 (2.7—9.8)* |  | 3.90 (2.0—7.3)* | <0.01* |
|  | West | 1.4 (0.6—2.7) |  | 1.3 (0.6—2.6) | 0.45 |
|  | **Literacy status** |  |  |  |  |
|  | Literate | Ref |  | Ref |  |
|  | Illiterate | 1.4 (1.0—1.7) |  | 1.6 (1.0—2.7) | 0.05 |
|  | **Household income (INR)** |  |  |  |  |
|  | >=4000 | Ref |  | Ref |  |
|  | <4000 | 1.9 (1.2—3.1) |  | 1.3 (0.8—2.1) | 0.29 |
| Shewade 2019 (Multi-site, 18 Indian districts)^f^ [12] |  |  |  |  |  |
|  | **Sex** |  |  |  |  |
|  | Male | Ref |  | Ref |  |
|  | Female | 1.35 (0.54—3.37) | 0.52 | 1.76 (0.59—5.27) | 0.313 |
|  | **Age in years**^c^ |  |  |  |  |
|  | Per every year increase in age |  |  | 1.02 (0.97—1.03) | 0.104 |
|  | **Residence** |  |  |  |  |
|  | Urban | Ref |  | Ref |  |
|  | Rural | 2.21 (0.21—22.91) | 0.51 | 2.20 (0.22—21.74) | 0.5 |
|  | **Education** |  |  |  |  |
|  | Higher Secondary and above | Ref |  | Ref |  |
|  | No Formal Education | 1.05 (0.16—7.11) | 0.95 | 0.50 (0.14—1.84) | 0.30 |
|  | Less than Primary | 0.85 (0.18—4.08) | 0.834 | 045 (0.14—1.48) | 0.19 |
|  | Up to Secondary | 0.46 (0.07—2.99) | 0.41 | 0.29 (0.07—1.31) | 0.11 |
|  | **Occupation** |  |  |  |  |
|  | Unemployed | Ref |  | Ref |  |
|  | Homemaker | 1.14 (0.55—2.38) | 0.73 | 0.92 (0.46—1.83) | 0.81 |
|  | Daily wage laborer | 1.68 (1.01—2.81)* | 0.048 | 2.17 (1.11—4.22)* | 0.023* |
|  | Employed but not daily wage | 0.96 (0.22—4.25) | 0.96 | 1.30 (0.29—6.11) | 0.74 |
|  | **TB death in household** |  |  |  |  |
|  | No | Ref |  | Ref |  |
|  | Yes | 1.90 (0.34—10.7) | 0.468 | 2.02 (0.42—9.7) | 0.38 |
|  | **Patient delay in care seeking (a measure of symptom duration)**^c^ |  |  |  |  |
|  | Per each day increase in symptom duration | 1.004 (1.002—1.006)* | <0.001* | 1.004 (1.001—1.008)* | 0.007* |
| Thomas 2015  (Tamil Nadu)^a^ [15] |  |  |  |  |  |
|  | **Duration of stay in brick kilns** |  |  |  |  |
|  | <=4 months | Ref |  |  |  |
|  | 5-6 months | 0.59 (0.38—0.92)* | 0.018* | Not reported |  |
|  | >6 months | 0.18 (0.08—0.44)* | <0.01* | Not reported |  |
| Thomas 2021  (Multi-site, 17 Indian states)^a^ [16] |  |  |  |  |  |
|  | **Sex** |  |  |  |  |
|  | Female | Ref |  |  |  |
|  | Male | 0.97 (0.81-1.17) | 0.713 | Not Reported |  |
|  | **Age in years** |  |  |  |  |
|  | 15-34 | 0.63 (0.5-0.78)* | <0.001* | Not Reported |  |
|  | 35-54 | 0.71 (0.57-0.88)* | 0.001* | Not Reported |  |
|  | 55 | Ref |  |  |  |
|  | **Occupation** |  |  |  |  |
|  | Employed | Ref |  |  |  |
|  | Unemployed | 0.58 (0.38-0.9)* | 0.015* | Not Reported |  |
|  | **Education** |  |  |  |  |
|  | Literate | Ref |  |  |  |
|  | Illiterate | 1.38 (1.15-1.64)* | <0.001* | Not Reported |  |
|  | **Cost incurred** |  |  |  |  |
|  | No | Ref |  |  |  |
|  | Yes | 1.58 (1.30-1.91)* | <0.001* | Not Reported |  |
|  | **Cough** |  |  |  |  |
|  | Yes | Ref |  |  |  |
|  | No | 0.92 (0.69-1.23) | 0.578 | Not Reported |  |
|  | **Expectoration** |  |  |  |  |
|  | Yes | Ref |  |  |  |
|  | No | 0.99 (0.83-1.19) | 0.924 | Not Reported |  |
|  | **Chest pain** |  |  |  |  |
|  | Yes | Ref |  |  |  |
|  | No | 0.88 (0.73-1.07) | 0.203 | Not Reported |  |
|  | **Fever** |  |  |  |  |
|  | Yes | Ref |  |  |  |
|  | No | 1.22 (1.02-1.46)* | 0.03* | Not Reported |  |
|  | **Loss of Appetite** |  |  |  |  |
|  | Yes | Ref |  |  |  |
|  | No | 1.19 (0.99-1.43) | 0.053 | Not Reported |  |
|  | **Blood in Sputum** |  |  |  |  |
|  | Yes | Ref |  |  |  |
|  | No | 1.69 (1.32-2.16)* | <0.001* | Not Reported |  |
|  | **Night sweats** |  |  |  |  |
|  | Yes | Ref |  |  |  |
|  | No | 0.96 (0.78-1.19) | 0.705 | Not Reported |  |
|  | **Weight loss** |  |  |  |  |
|  | Yes | Ref |  |  |  |
|  | No | 1.59 (1.33-1.89)* | <0.001* | Not Reported |  |
|  | **Shortness of breath** |  |  |  |  |
|  | Yes | Ref |  |  |  |
|  | No | 1.43 (1.19-1.72)* | <0.001* | Not Reported |  |
|  | **Tiredness** |  |  |  |  |
|  | Yes | Ref |  |  |  |
|  | No | 1.27 (1.06-1.52)* | 0.008* | Not Reported |  |
|  | **Diabetes** |  |  |  |  |
|  | Yes | Ref |  |  |  |
|  | No | 1.41 (0.69-2.89) | 0.350 | Not Reported |  |
|  | **Hypertension** |  |  |  |  |
|  | Yes | Ref |  |  |  |
|  | No | 2.59 (1.75-3.85)* | <0.001* | Not Reported |  |
|  | **HIV** |  |  |  |  |
|  | Yes | Ref |  |  |  |
|  | No | 3.56 (2.02-6.28)* | <0.001* | Not Reported |  |
|  | **Asthma** |  |  |  |  |
|  | Yes | Ref |  |  |  |
|  | No | 1.80 (1.38-2.35)* | <0.001* | Not Reported |  |
|  | **Malaria** |  |  |  |  |
|  | Yes | Ref |  |  |  |
|  | No | 0.81 (0.6-1.09) | 0.157 | Not Reported |  |
|  | **Alcohol consumption** |  |  |  |  |
|  | No | Ref |  |  |  |
|  | Yes | 0.93 (0.77-1.12) | 0.463 | Not Reported |  |
|  | **Smoking** |  |  |  |  |
|  | No | Ref |  |  |  |
|  | Yes | 0.92 (0.75-1.12) | 0.426 | Not Reported |  |
|  | **Substance Abuse** |  |  |  |  |
|  | No | Ref |  | Not Reported |  |
|  | Yes | 2.70 (1.86-3.92)* | <0.001* |  |  |
|  | **Knowledge on TB** |  |  |  |  |
|  | Have not heard about TB vs. low TB knowledge (score <6) | 2.83 (2.2-3.62)* | <0.001* | Not Reported |  |
|  | Have not heard about TB vs. high TB knowledge (score =6) | 4.64 (3.7-5.83)* | <0.001* | Not Reported |  |
|  | **Long distance** |  |  |  |  |
|  | No | Ref |  | Not Reported |  |
|  | Yes | 1.54 (1.27-1.86)* | <0.001* |  |  |
|  | **Poor attitude of healthcare workers** |  |  |  |  |
|  | No | Ref |  |  |  |
|  | Yes | 1.06 (0.85-1.33) | 0.601 | Not Reported |  |
|  | **Lack of Services** |  |  |  |  |
|  | No | Ref |  | Not Reported |  |
|  | Yes | 1.21 (0.90-1.63) | 0.216 |  |  |

KHPT, Karnataka Health Promotion Trust; TB, tuberculosis; THALI, Tuberculosis Health Action Learning Initiative.

^a^Study reported having sought care as the outcome, so effect estimates (odds ratios and 95% confidence intervals) were “flipped” to present the odds of not having sought care for all studies.

^b^Reference group of the variable was switched, resulting in flipping of the effect estimate (odds ratio and 95% confidence interval), to facilitate consistency in reference groups across studies or to allow for more intuitive interpretation of the study finding. For example, we sometimes did this to show how the absence (rather than the presence) of a symptom might result in increased risk of not seeking care or to present an odds ratio of greater than one (rather than less than one) for a particular association.

^c^Variable was included in the analysis as a continuous variable. We have specified the unit of change in the variable associated with the effect estimate.

^d^Unadjusted odds ratios and/or p-values were estimated by the systematic review team from the raw data, as these were not provided in the original study.

^e^For the Foschen et al. (2006) study, sex, age, marital status, caste, and hemoptysis were also included in the multivariable model; however, results were not reported for these variables in the original paper because the associations were not statistically significant.

^f^These findings were not reported in the original studies; however, new analyses were performed by the study authors using the original datasets, based on a request from the systematic review team.

*Indicates statistical significance at the 5% level.

### References

1.	Subbaraman R, Nathavitharana RR, Satyanarayana S, Pai M, Thomas BE, Chadha VK, et al. The Tuberculosis Cascade of Care in India’s Public Sector: A Systematic Review and Meta-analysis. PLoS Med. 2016;13: e1002149. doi:10.1371/journal.pmed.1002149

2.	Charles N, Thomas B, Watson B, Raja Sakthivel M, Chandrasekeran V, Wares F. Care seeking behavior of chest symptomatics: a community based study done in South India after the implementation of the RNTCP. PLoS One. 2010;5. doi:10.1371/journal.pone.0012379

3.	Fochsen G, Deshpande K, Diwan V, Mishra A, Diwan VK, Thorson A. Health care seeking among individuals with cough and tuberculosis: a population-based study from rural India. Int J Tuberc Lung Dis. 2006;10: 995–1000.

4.	George O, Sharma V, Sinha A, Bastian S, Santha T. Knowledge and behaviour of chest symptomatics in urban slum populations of two states in India towards care-seeking. Indian Journal of Tuberculosis. 2013;60: 95–106.

5.	Ghosh S, Sinhababu A, Taraphdar P, Mukhopadhyay DK, Mahapatra BS, Biswas AB. A study on care seeking behavior of chest symptomatics in a slum of Bankura, West Bengal. Indian J Public Health. 2010;54: 42–44. doi:10.4103/0019-557X.70553

6.	Helfinstein S, Engl E, Thomas BE, Natarajan G, Prakash P, Jain M, et al. Understanding why at-risk population segments do not seek care for tuberculosis: a precision public health approach in South India. BMJ Glob Health. 2020;5. doi:10.1136/bmjgh-2020-002555

7.	Kar M, Logaraj M. Awareness, attitude and treatment seeking behaviour regarding tuberculosis in a rural area of Tamil Nadu. Indian J Tuberc. 2010;57: 226–229.

8.	Karanjekar V, Gujarati V, Lokare P. Sociodemographic factors associated with health seeking behavior of chest symptomatics in urban slums of Aurangabad city, India. International Journal of Basic and Applied Medical Sciences. 2014;4: 173–179.

9.	Knowledge about TB and Health Seeking Behaviour among Adult Chest Symptomatic (CS) Population Living in Urban Slums of Hyderabad City. A Baseline Study Report: 2016-17. Karnataka Health Promotion Trust (KHPT) Tuberculosis Health Action Learning Initiative (THALI);

10.	Knowledge about TB and Health Seeking Behaviour among Adult Chest Symptomatic (CS) Population Living in Urban Slums of Bengaluru City. A Baseline Study Report: 2016-17. Karnataka Health Promotion Trust (KHPT) Tuberculosis Health Action Learning Initiative (THALI);

11.	Satyanarayana S, Nair SA, Chadha SS, Sharma G, Yadav S, Mohanty S, et al. Health-care seeking among people with cough of 2 weeks or more in India. Is passive TB case finding sufficient? Public Health Action. 2012;2: 157–161. doi:10.5588/pha.12.0019

12.	Shewade HD, Gupta V, Satyanarayana S, Pandey P, Bajpai UN, Tripathy JP, et al. Patient characteristics, health seeking and delays among new sputum smear positive TB patients identified through active case finding when compared to passive case finding in India. PLoS One. 2019;14: e0213345. doi:10.1371/journal.pone.0213345

13.	Shriraam V, Srihari R, Gayathri T, Murali L. Active case finding for Tuberculosis among migrant brick kiln workers in South India. Indian J Tuberc. 2020;67: 38–42. doi:10.1016/j.ijtb.2019.09.003

14.	Suganthi P, Chadha VK, Ahmed J, Umadevi G, Kumar P, Srivastava R, et al. Health seeking and knowledge about tuberculosis among persons with pulmonary symptoms and tuberculosis cases in Bangalore slums. Int J Tuberc Lung Dis. 2008;12: 1268–1273.

15.	Thomas BE, Charles N, Watson B, Chandrasekaran V, Senthil Kumar R, Dhanalakshmi A, et al. Prevalence of chest symptoms amongst brick kiln migrant workers and care seeking behaviour: a study from South India. J Public Health (Oxf). 2015;37: 590–596. doi:10.1093/pubmed/fdu104

16.	Thomas BE, Thiruvengadam K, S R, Rani S, S V, Gangadhar Rao V, et al. Understanding health care-seeking behaviour of the tribal population in India among those with presumptive TB symptoms. PLoS One. 2021;16: e0250971. doi:10.1371/journal.pone.0250971
