## Supplemental Appendix 2 for "Barriers to engagement by people with active tuberculosis in the care cascade in India: a systematic review of two decades of quantitative research"

**Supplementary Appendix 2**

**Supplement to:**

Barriers to engagement by people with active tuberculosis in the care cascade in India: a systematic review of two decades of quantitative research

Tulip A. Jhaveri, Disha Jhaveri, Amith Galivanche, Dominic Voehler, Maya Lubeck-Schricker, Mei Chung, Pruthu Thekkur, Vineet Chadha, Ruvandhi Nathavitharana, Ajay M.V. Kumar, Hemant Deepak Shewade, Katherine Powers, Kenneth H. Mayer, Jessica E. Haberer, Paul Bain, Madhukar Pai, Srinath Satyanarayana, Ramnath Subbaraman

**Correspondence:**

Ramnath Subbaraman, MD, MSc, FACP

Tufts University School of Medicine

Department of Public Health and Community Medicine

136 Harrison Ave., MV237

Boston, MA 02130, USA

**Methods and findings for the systematic review of barriers to completion of the diagnostic workup by individuals with presumptive tuberculosis (Gap 2)**

### Methods

##### Objectives

The objective of this systematic review was to understand into why some individuals with presumptive tuberculosis (TB) presenting to TB diagnostic facilities—government designated microscopy centers (DMCs) or private facilities—do not complete the appropriate diagnostic workup for TB (Gap 2 in the care cascade). We defined individuals with presumptive TB as people with cough >2 weeks or other symptoms that could be suggestive of active TB, as defined by the participant inclusion criteria used in each study. We defined individuals with presumptive drug-resistant TB (DR TB) as comprising individuals who met criteria for undergoing drug-susceptibility testing and other workup for DR TB in India’s TB program, including being a person living with HIV (PLHIV), having a prior history of TB treatment, or experiencing failure during treatment for drug-susceptible TB.

Notably, Stage 2 of the care cascade comprises multiple steps in the diagnostic workup that vary depending on the type of presumed TB (e.g., smear-positive TB, smear-negative TB, DR TB, etc.). In general, for each type of presumed TB, a different diagnostic algorithm was used, usually culminating in the completion of a specific diagnostic test that would diagnose that form of TB. For example, sputum smear microscopy had to be completed to diagnose smear-positive TB; chest X-ray had to be completed to diagnose smear-negative TB; and cartridge-based nucleic acid amplification testing (CBNAAT, e.g., Xpert MTB/RIF testing) or mycobacterial culture had to be completed to diagnose rifampin-resistant TB (RR TB) or multidrug-resistant TB (MDR TB), respectively. As such, an assumption of our approach to reporting findings is that non-completion of the diagnostic workup varies based on the specific diagnostic test required.

We therefore break down our reporting of Gap 2 findings as follows:

(a) *Non-pursual of diagnostic workup despite referral*: These studies evaluated patients recognized as having presumptive TB who were referred for, but did not pursue, any diagnostic workup.
(b) *Non-completion of sputum microscopy evaluation*: These studies evaluated non-completion of sputum microscopy testing, which comprised not completing submission of either two or three sputum samples, depending on the study year. Prior to April 2009, the workup for TB in India started with collection of three sputum samples. After April 2009, the workup for TB in India started with collection of only two sputum samples.
(c) *Non-completion of chest X-ray*: Nearly all of these studies related to the diagnostic workup for presumed smear-negative TB. While the recommended diagnostic algorithm for presumptive smear-negative TB evolved over time, all such algorithms culminated in the need to complete a chest X-ray to confirm the diagnosis. In addition, one recent study evaluated non-completion of chest X-ray after 2017, when the diagnostic algorithm in India’s National TB Elimination Program (NTEP) was changed such that all individuals with presumptive TB were supposed to undergo chest X-ray. Given that the availability of chest X-ray facilities (rather than the specific diagnostic algorithm) seemed to drive non-completion, we compare findings across all of these studies that evaluated non-completion of chest X-ray, regardless of the diagnostic algorithm used.
(d) *Non-completion of CBNAAT, line probe assay, or mycobacterial culture*: Several studies evaluated non-completion of the diagnostic algorithm for presumed DR TB or RR TB. While the recommended diagnostic algorithm for DR TB evolved over time, with the exception of two papers that evaluated completion of mycobacterial culture [1] or line probe assay, most such algorithms culminated in completion of CBNAAT testing for the papers included in our review. In addition, one recent study evaluated non-completion of CBNAAT after 2017, when the diagnostic algorithm in India’s National TB Elimination Program (NTEP) was changed such that patients with positive sputum microscopy tests, chest X-ray evidence of TB, or who were PLHIV were supposed to undergo CBNAAT. Again, given that the availability of the testing modality seemed to drive non-completion of this workup, we compare findings across all of these studies that evaluated non-completion of CBNAAT, line probe assay, or mycobacterial culture, regardless of the diagnostic algorithm used.

We extracted three types of quantitative data that help to understand non-completion of the TB diagnostic workup:

(a) *Factors associated with not having completed a specific diagnostic test*: For studies comparing individuals who had or had not completed a specific diagnostic test (e.g., sputum microscopy, chest X-ray, or CBNAAT), we extracted effect estimates for variables associated with non-completion. Effect estimates included odds ratios, risk ratios, hazard ratios, or beta-coefficients, depending on the approach to analysis.
(b) *Reasons reported by patients for non-completion of the diagnostic workup*: For studies that surveyed patients who had not completed a specific diagnostic test, we extracted the proportion of individuals who reported a given reason for not completing the workup. *Notably, the denominator for these analyses was the number of patients who did not complete a specific workup. The numerator in these analyses was the number of patients who reported a given reason for not completing the workup.*
(c) *Reasons from health system records for non-completion of the diagnostic workup*: For individuals with presumptive DR or RR TB, multiple studies extracted data from medical records that describe health system-related reasons for non-completion of tests (e.g., healthcare providers not recognizing patients had presumptive DR TB, misplacement of sputum samples during transportation to a testing center, etc.). Notably, these analyses reported findings differently than studies that directly interviewed patients who had not completed the diagnostic workup. *Specifically, the denominator in these analyses was the overall number of patients with presumptive DR TB—rather than the number of patients who did not complete the diagnostic workup. The numerator in these analyses was the number of patients who experienced a specific health system problem contributing to non-completion of the diagnostic workup.*

##### Search strategy

Three separate searches were conducted to identify articles. The first search was conducted as part of a previously published study quantifying gaps in India’s TB care cascade [2]. We used articles identified for that review that evaluated Gap 2 in the TB care cascade but that also reported factors and reasons associated with not completing the TB diagnostic workup. For that review, a medical librarian searched PubMed, Embase, and Web of Science for studies published between January 1, 2000 and February 26, 2015, without language restrictions, using search terms and related variants for “tuberculosis”, “India”, and “loss to follow-up”, including “pretreatment loss to follow-up” and “initial default” to include early losses preceding treatment initiation, including during the diagnostic workup (Table A). We also carried out electronic searches of key Indian journals that were not indexed for that entire time window: the Indian Journal of Tuberculosis, Lung India, the Indian Journal of Chest and Allied Sciences, the India Journal of Public Health, and the Indian Journal of Community Medicine. Additional studies were identified by searching reference lists of the primary studies and relevant review articles. Notably, given similarities in the search terms for identifying patient losses during the diagnostic workup (Gap 2), pretreatment loss to follow-up (Gap 3), and poor outcomes during treatment (Gap 4), this single search was used to identify studies related to all of these gaps. We screened all identified studies from this previous review for potential inclusion in our current review; however, different data were extracted from studies that met our inclusion criteria.

To update our review, we conducted a second refresher search using the same search terms for October 2, 2015 to October 1, 2019. We did not repeat hand searches of the Indian journals listed above, because all of these journals had been indexed in PubMed prior to the time period of this more recent search. Due to the extensive time required to extract data from the articles identified for this systematic review, we performed a third refresher search using the same search terms for October 2, 2019 to May 17, 2021. Finally, additional studies were identified by looking through the reference lists of the included primary studies and relevant review articles that were identified by the searches and by outreach to experts in the field.

###### Table A. Search strategy to identify manuscripts regarding non-completion of the diagnostic workup for TB patients in India (Gap 2). This same search was also used to identify relevant articles for Gaps 3 and 4.

| Terms for tuberculosis: | “tuberculosis”[Mesh] OR *Mycobacterium tuberculosis*[tiab] OR TB[tiab] OR MDRTB[tiab] OR XDRTB[tiab] |
| --- | --- |
| Terms for India: | “India”[Mesh] OR India[tiab] OR India[ad] OR Indian[tiab] OR Indians[tiab] |
| Terms for loss to follow-up or other poor outcomes: | “patient dropouts”[tiab] OR “treatment refusal”[Mesh] OR “patient compliance”[Mesh] OR lost to follow up[tiab] OR loss to follow up[tiab] OR default*[tiab] OR compliance[tiab] OR adherence[tiab] OR noncompliance[tiab] OR nonadherence[tiab] OR diagnostic dropout [tiab] OR patient cooperation[tiab] OR dropout*[tiab] OR linkage to care[tiab] OR retention[tiab] OR attrition[tiab] OR cascade of care[tiab] OR treatment cascade[tiab] OR treatment success*[tiab] OR treatment completion[tiab] OR cure[tiab] OR pretreatment loss to follow-up[tiab] OR initial default[tiab]; treatment failure [tiab] |

##### Inclusion and exclusion criteria

We applied the following criteria for inclusion and exclusion of studies for this systematic review.

*Inclusion criteria* included the following:

(1) Studies that followed individuals who were recognized by healthcare providers as having presumptive TB and then referred for diagnostic evaluation. As describe above, depending on the patient population being studied, these studies may have followed patients to see if they pursued TB diagnostic workup after initial referral, completed sputum microscopy evaluation, completed chest X-ray, or complete CBNAAT, line probe assay, or mycobacterial culture.
(2) Studies also had to have assessed reasons that individuals in the study may not have completed the diagnostic workup, by comparing characteristics of those who did or did not complete the workup (e.g., regression analyses) or by follow-up structured interviews with patients who did not complete the workup.

*Exclusion criteria* included the following:

(1) Studies that only described the proportion of TB patients who did not complete a given diagnostic workup, without evaluating reasons for non-completion.
(2) Studies with data collected prior to the year 2000, as India’s Revised National TB Control Programme (now called the National TB Elimination Programme) did not achieve nationwide coverage until the early 2000s.
(3) Studies only containing qualitative data evaluating why individuals did not complete the TB diagnostic workup. Findings from studies containing qualitative data will be reported in a separate paper.

##### Study selection

Each citation identified by the search was independently assessed by at least two reviewers (among TJ, DJ, AG, DV, MLS and KP) for their eligibility at the title and abstract evaluation stage and again subsequently at the full text evaluation stage (Fig A). Disagreements between the two reviewers were resolved by discussion or, if necessary, through consultation of a third reviewer (RS). Independent selection of articles at the title and abstract and full text stages was conducted using Covidence software (Veritas Health Innovations, Melbourne, Australia); however, quality assessment and extraction of study findings was conducted using an Excel spreadsheet.

###### Fig A. PRISMA flowchart: study selection for the systematic review of non-completion of the diagnostic workup, pretreatment loss to follow-up, and on-treatment loss to follow-up for TB patients in India (Gaps 2, 3 and 4)

**Search 1: January 1, 2000 to October 1, 2015**

**Search 3: October 2, 2019 to May 17, 2021**

**Search 2: October 2, 2015 to October 1, 2019**

Reports identified from search of PubMed, Embase, and Web of Science and hand search of relevant journals (n=3,160), of which 1,243 were duplicates.

Duplicates removed before screening

(n=29)

Records identified from search of PubMed, Embase, and Web of Science (n=496)

Duplicate records removed before screening (n=0)

Records identified from search of PubMed, Embase, and Web of Science (n=860)

**Identification**

Records excluded after title and abstract screen

(n=429)

Records excluded after title and abstract screen

(n=762)

Records that underwent screening of title and abstract

(n=467)

Records that underwent screening of title and abstract

(n=860)

Reports shortlisted after removal of duplicates (n=1,917) of which 1,751 were excluded after title and abstract review.

Records not retrieved (n=0)

Records for which full text articles were retrieved (n=38)

Reports not retrieved

(n=0)

Records for which full text articles were retrieved (n=98)

**Screening**

Reports eligible for full text review (n=166), of which 88 were excluded.

Reports excluded after full text review

(n = 20), for the following reasons:

Wrong outcomes evaluated (n=7)

Wrong patient population (n=2)

Wrong study design

(n=3)

Wrong setting (n=1)

No information on reasons for bad outcome (n=7)

Reports assessed for eligibility via full text review (n=38)

Reports excluded after full text review (n = 71), for the following reasons:

Wrong outcome evaluated (n=52)

Wrong patient population (n=1)

Wrong study design (n=7)

No information on reasons for bad outcome (n=11)

Reports assessed for eligibility via full text review (n=98)

Studies included in previous care cascade study with relevant data on reasons for gap 2, 3 and 4 (n=78)

New studies included in the systematic review from the 2019 to 2021 search (n=18)

New studies included in the systematic review from the 2015 to 2019 search (n=27)

**Included**

Total studies included in the systematic review (n=123)

Gap 2 (n=10)

Gap 3 (n=12)

Gap 4 (n=82)

Multiple gaps (n=19)

##### Quality assessment of quantitative studies

In our previous systematic review, we developed quality criteria relevant to studies focused on identifying patients who did not complete the TB diagnostic workup (Table B), because there were no existing standardized guidelines for assessing the quality of these types of studies. Although we extracted different data from these studies in the current systematic review, we used the same quality criteria for studies that included relevant quantitative information, because these criteria are still appropriate for assessing the overall quality of these types of observational studies.

We classified the health facility-based strategy for sampling patients in each study based on whether it was comprehensive, random, or convenience. Studies using convenience sampling were excluded from analysis. Studies that assessed more than 150 patients and at over one government or private sector diagnostic facility were rated as being higher in quality than studies that assessed less than 150 patients at a single diagnostic facility. Studies that assessed outcomes within a shorter time frame after the start of diagnostic workup (e.g., within 1-2 months) were rated as being higher in quality than studies that assessed these indicators 3 or more months after the start of diagnostic workup, since tracking patients to find out accurate outcomes becomes much harder with time due to patient mobility. Specifically, as the time increases between the start of patient’s diagnostic evaluation and follow-up of the patient’s outcome by a study team, there is a higher likelihood of misclassifying a patient’s outcome as “non-completion of diagnostic workup” simply because it is not possible to find the patient because they have moved, obtained a new cell phone, etc. For the same reason, studies in which a dedicated research team was used to track patients were rated as being higher in quality than those relying on self-report by local TB programs to evaluate non-completion of the diagnostic workup.

###### Table B. Criteria for assessing quality of quantitative studies evaluating failure to complete the diagnostic workup for TB patients in India

| **Criterion** | **Quality level** |
| --- | --- |
| **Sampling strategy** | |
| Random (i.e., probability-based) or comprehensive sampling at selected TB diagnostic facilities | High |
| Convenience sampling or not reported | Low (exclude from analysis of findings) |
| **Sample size** | |
| >1 diagnostic center and 150+ patients | High |
| Single diagnostic center study with 150+ patients | Medium |
| <150 patients or not reported | Low |
| **Time frame of research fieldwork after start of diagnostic evaluation** | |
| 2 weeks to 1 month after start of diagnostic evaluation | High |
| 1-3 months after start of diagnostic evaluation | Medium |
| >3 months after start of diagnostic evaluation or not reported | Low |
| **Method of evaluating outcome** | |
| Patient tracking by a dedicated research team | High |
| Relying on self-report by government TB program or not reported | Medium to Low |

##### Data extraction and analysis

Multiple reviewers (TJ, AG, DV, MLS, and DJ) independently extracted data from each included study into a structured form on an Excel spreadsheet; however, we ensured that every article had data independently extracted by at least two or more reviewers. Disagreements were resolved by discussion or, if necessary, by consulting a third reviewer (RS). For each study, we extracted information on the study design, location, setting (i.e., urban versus rural), sample size, and variables of interest (Table C).

For studies that compared individuals who had completed a specific part of the TB diagnostic workup against those who had not completed that part of the workup, we extracted adjusted and unadjusted effect estimates (odds ratios, risk ratios, hazard ratios, and beta-coefficients) from regression analyses. For studies that did not report effect estimates, we calculated unadjusted odds ratios from the data provided, if possible. For studies that reported reasons why individuals had not completed part of the diagnostic workup, we extracted the proportion of individuals surveyed who reported a given reason for noncompletion or the proportion of patients or patients samples who did not complete part of the workup due to a specific health system problem (e.g., misplaced or untested sputum samples). For effect estimates and proportions, we extracted information on 95% confidence intervals (95% CIs) where available; if 95% CIs were not reported, we calculated these from the data provided, if possible.

For the regression analyses, some studies reported factors associated with having completed part of the TB diagnostic workup, while others reported factors associated with not having completed part of the diagnostic workup. For consistency, we “flipped” effect estimates and 95% CIs as needed so that all findings represented associations with the outcome of not having completed the diagnostic workup. For some variables, we also changed the reference group as needed for consistency of reporting across studies. For example, because most studies compared men to the reference group of women, we “flipped” effect estimates and confidence intervals for studies that presented men as the reference group. This allowed us to consistently present women as the reference group for findings regarding gender.

After “flipping” selected effect estimates from the regression analyses, we reported all unadjusted and adjusted effect estimates, regardless of statistical significance, organized by study (Table D). For the main manuscript and Forest plot, we restricted ourselves to presenting statistically significant adjusted effect estimates from multivariable analyses, as these may represent more meaningful associations from higher-quality analyses. After extracting this subset of findings, we organized findings into categories using our framework of demand- and supply-side factors (main manuscript, Table 1). For findings on reasons why individuals had not completed part of the TB diagnostic workup, we presented all reported proportions in the Forest Plots in the main manuscript; we also organized these findings using our framework of demand- and supply-side categories.

To visualize quantitative findings, we generated Forest plots of effect estimates odds ratios, risk ratios, hazard ratios, beta-coefficients, and proportions using Stata version 16.1 (College Station, TX, USA). As described earlier, we present these Forest plots organized by the specific type of diagnostic modality that was not completed (i.e., noncompletion of sputum microscopy, chest X-ray, or CBNAAT). We did not conduct meta-analyses of data, because we extracted findings represented a diverse set of variables from every study.

###### Table C. Characteristics of the included studies for individuals with presumptive tuberculosis who did not complete specific steps of the diagnostic workup (Gap 2)

| **Citation**  (year) | **Location** | **Urban, rural, both, or unknown** | **Type of population** | **Single or multiple designated microscopy centers** (DMCs) | **Number of overall individuals with presumptive TB followed in the study** | **Time frame of research fieldwork after patient diagnosis** | **Methodology** (Patient tracking by a dedicated research team versus self-report by the government TB program) | **Type of findings included in the study**  (N=sample or denominator of patients for given analysis) |
| --- | --- | --- | --- | --- | --- | --- | --- | --- |
| **Non-pursual of diagnostic workup despite referral** |  |  |  |  |  |  |  |  |
| Dey (2019)[3] | West Bengal | Urban | Individuals with symptoms identified via active case finding in high TB burden wards | Multiple | 1,132 | 0-4 weeks | Patient tracking by dedicated research team | Relative risk regression (N=1,132) |
| Garg (2020) [4] | Bihar | Rural | Individuals with symptoms identified via active case finding in the general population | Multiple | 11,146 | 0-4 weeks | Patient tracking by dedicated research team | Relative risk regression (N=11,146) |
| Ismail (2020) [5] | Karnataka | Both | Individuals with symptoms identified via passive case finding (i.e., routine care) at government clinics | Multiple | 8,822 | 0-4 weeks | Self-report by government TB program* | Relative risk regression (N=8,822) |
| **Non-completion of sputum microscopy evaluation** |  |  |  |  |  |  |  |  |
| Balasubramanian (2004) [6] | Tamil Nadu | Rural | Adults >14 years | Multiple | 8,646 | 1-3 months* | Patient tracking by dedicated research team | Logistic regression (N=8,646)^a^ |
| Chandrasekaran (2005) [7] | Tamil Nadu | Urban | Adults >15 years | Multiple | 1,000 | 2-4 weeks | Patient tracking by dedicated research team | Reasons for non-completion by patient interview (proportions) (N=92) |
| Dandona (2004) [8] | Andhra Pradesh, Maharashtra, Rajasthan, and Tamil Nadu | Both | General | Multiple | 4,310 | NR* | Patient tracking by dedicated research team | Reasons for non-completion by patient interview (proportions) (N=314); Logistic regression (N=4,310) |
| Das (2019)^b^ [9] | Chhattisgarh | Rural | Conflict-Affected | Multiple | 763 | 2-4 weeks | Patient tracking by dedicated research team | Logistic regression (N=763)^a^ |
| Tripathy (2013) [10] | Punjab | Rural | General | Multiple | 1,708 | 2-4 weeks | Self-report by government TB program* | Logistic regression (N=1,708)^a^ |
| **Non-completion of chest X-ray** |  |  |  |  |  |  |  |  |
| Chadha (2013) [11] | Karnataka | Both | General | Multiple | 256 | 2-4 weeks and 1-3 months | Patient tracking by dedicated research team | Reasons for non-completion by patient interview (proportions) (N=243) |
| Kanakaraju (2020) [12] | Karnataka | Rural | General | Multiple | 732 | NR | Patient tracking by dedicated research team | Relative risk regression (N=732) |
| Sarkar (2011) [13] | West Bengal | Both | General | Multiple | 4,875 | NR | Patient tracking by dedicated research team | Logistic regression (N=4,875) |
| Thomas (2006) [14] | Tamil Nadu | Both | General | Multiple | 423 | NR | Patient tracking by dedicated research team | Reasons for non-completion by patient interview (proportions) (N=148) |
| **Non-completion of CBNAAT, line probe assay, or mycobacterial culture** |  |  |  |  |  |  |  |  |
| Chadha (2011)^c^ [1] | Andhra Pradesh | Both | General | Multiple | 559 | >3 months* | Self-report by government* | Reasons for non-completion from health system records (proportions) (N=559) |
| Kanakaraju (2020)^d^ [12] | Karnataka | Rural | General | Multiple | 732 | NR | Patient tracking by dedicated research team | Logistic regression (N=732)^a^ |
| Natrajan (2018) [15] | Madhya Pradesh | Both | General | Multiple | 318 | NR | Patient tracking by dedicated research team | Reasons for non-completion from health system records (proportions) (N=318) |
| Shewade (2017) [16] | Madhya Pradesh | Urban | General | Multiple | 770 | >3 months* | Patient tracking by dedicated research team | Reasons for non-completion from health system records (proportions) (N=770); Relative risk regression (N=770) |
| Shewade (2017) [17] | Tamil Nadu | Urban | General | Multiple | 628 | >3 months* | Patient tracking by dedicated research team | Reasons for non-completion from health system records (proportions) (N=628); Relative risk regression (N=628) |
| Shewade (2015) [18] | Puducherry | Urban | General | Multiple | 341 | 1-3 months* | Patient tracking by dedicated research team | Reasons for non-completion from health system records (proportions) (N=341); Relative risk regression (N=341) |
| Shewade (2016) [19] | Puducherry | Urban | General | Multiple | 212 | NR* | Patient tracking by dedicated research team | Reasons for non-completion from health system records (proportions) (N=212); Relative risk regression (N=212) |
| Singla (2014)^e^ [20] | Delhi | Urban | General | Single* | 3,814 | NR* | Patient tracking by dedicated research team | Logistic regression (N=3,814) |

CBNAAT=cartridge-based nucleic acid amplification testing (e.g., Xpert MTB/RIF, Trunat), NR=Not reported

*Medium or low quality for this indicator

^a^Unadjusted odds ratios and/or p-values were estimated by the systematic review team from the raw data, as these were not provided in the original study.

^b^This study compared rates of completion of the diagnostic workup between initial testing with sputum microscopy as compared to a later time period after Xpert MTB/RIF had been implemented as the initial test.

^c^This study evaluated non-completion of mycobacterial culture for DR TB evaluation and was conducted before widespread rollout of CBNAAT testing.

^d^This study evaluated non-completion of CBNAAT implemented a part of evaluation of all individuals with presumptive TB, rather than as part of focused evaluation of individuals with presumptive DR or RR TB.

^e^This study evaluated non-completion of DR TB evaluation before and after introduction of line probe assay.

###### Table D. Factors associated with non-completion of the tuberculosis (TB) diagnostic workup by individuals with presumptive TB or presumptive drug-resistant TB in India (Gap 2)

| Study | Exposure / independent variable | Unadjusted Effect Estimate (95% Confidence Interval) | P-Value | Adjusted Effect Estimate (95% Confidence Interval) | P-Value |
| --- | --- | --- | --- | --- | --- |
| Non-pursual of diagnostic workup despite referral |  |  |  |  |  |
| Dey 2019 (West Bengal)^a^ [3] |  | Values below are relative risk ratios |  | Values below are adjusted relative risk ratios |  |
|  | **Age (years)** |  |  |  |  |
|  | <=14 | Ref |  | Ref |  |
|  | 15-29 | 1.1 (0.6-1.9) | Not reported | 1.3 (0.7-2.2) | Not reported |
|  | 30-44 | 1.3 (0.8-2.3) | Not reported | 1.5 (0.8-2.5) | Not reported |
|  | 45-59 | 1.2 (0.7-2.1) | Not reported | 1.4 (0.8-2.3) | Not reported |
|  | 60-74 | 1.3 (0.7-2.2) | Not reported | 1.4 (0.8-2.5) | Not reported |
|  | >=75 | 0.5 (0.2-1.3) | Not reported | 0.5 (0.2-1.5) | Not reported |
|  | Age not recorded | 3.4 (2.0-5.9)* | Not reported | 3.3 (1.9-5.6)* | Not reported |
|  | **Sex^e^** |  |  |  |  |
|  | Male | Ref |  | Ref |  |
|  | Female | 0.8 (0.7-0.9)* | Not reported | 0.77 (0.63-0.83)* | Not reported |
|  | **Presenting symptoms** |  |  |  |  |
|  | No cough but other symptoms | Ref |  | Ref |  |
|  | Cough with other symptoms | 1.1 (0.7-1.6) | Not reported | 1.1 (0.8-1.6) | Not reported |
|  | Only cough | 1.9 (1.6-2.4)* | Not reported | 1.8 (1.5-2.2)* | Not reported |
|  | Note, the risk ratio was flipped for the Forest plot to show:  No cough but other symptoms vs. only cough | 0.52 (0.42-0.63)* | Not reported | 0.56 (0.45-0.67)* | Not reported |
|  | **Previous history of TB** |  |  |  |  |
|  | Yes | Ref |  | Ref |  |
|  | No | 1.2 (0.8-1.6) | Not reported | 1.0 (0.8-1.4) | Not reported |
|  | **Family history of TB** |  |  |  |  |
|  | No | Ref |  | Ref |  |
|  | Yes | 1.7 (1.5-2.0)* | Not reported | 1.5 (1.3-1.8)* | Not reported |
|  | **Diabetes** |  |  |  |  |
|  | Yes | Ref |  | Ref |  |
|  | No/Unknown | 1.6 (1.2-1.9) | Not reported | 1.3 (1.0-1.6) | Not reported |
|  | **HIV** |  |  |  |  |
|  | Yes | Ref |  | Ref |  |
|  | No/Unknown | 4.9 (0.7-32.0) | Not reported | 6.0 (0.9-39.8) | Not reported |
|  | **Alcohol use** |  |  |  |  |
|  | No | Ref |  | Ref |  |
|  | Yes | 1.7 (1.4-2.2)* | Not reported | 1.7 (1.3-2.2)* | Not reported |
|  | **Tobacco use** |  |  |  |  |
|  | No | Ref |  | Ref |  |
|  | Yes | 1.2 (1.0-1.4) | Not reported | 1.2 (1.0-1.4) | Not reported |
| Garg 2020 (Bihar)^a^ [4] |  | Values below are relative risk ratios |  | Values below are adjusted relative risk ratios |  |
|  | **Age (years)** |  |  |  |  |
|  | >=65 | Ref |  | Ref |  |
|  | <15 | 1.4 (1.3-1.4)* | <0.001 | 1.2 (1.2-1.3)* | <0.001 |
|  | 15-44 | 1.0 (1.0-1.1) | 0.139 | 1.0 (1.0-1.1) | 0.366 |
|  | 45-64 | 1.0 (1.0-1.1) | 0.212 | 1.0 (1.0-1.1) | 0.164 |
|  | Note, the risk ratio was flipped for the Forest plot to show:  >=65 vs. <15 | 0.71 (0.71-0.77)* | <0.001 | 0.83 (0.77-0.83)* | <0.001 |
|  | **Block** |  |  |  |  |
|  | Sarairanjan | Ref |  |  |  |
|  | Ujiarpur | 1.0 (1.0-1.1) | 0.022 |  |  |
|  | Bibhutipur | 1.0 (1.0-1.1) | 0.033 |  |  |
|  | **Sex** |  |  |  |  |
|  | Female | Ref |  | Ref |  |
|  | Male | 1.1 (1.0-1.1)* | <0.001 | 1.1 (1.0-1.1)* | 0.001 |
|  | **Previous history of anti-TB treatment** |  |  |  |  |
|  | No | Ref |  | Ref |  |
|  | Yes | 0.7 (0.6-0.7)* | <0.001 | 0.7 (0.7-0.8)* | <0.001 |
|  | **Presenting symptoms** |  |  |  |  |
|  | Absence of listed signs | Ref |  |  |  |
|  | Hemoptysis in last 6 months | 0.6 (0.6-0.7)* | <0.001 | 0.7 (0.6-0.7)* | <0.001 |
|  | Cough ≥2 weeks | 0.7 (0.6-0.7)* | <0.001 |  |  |
|  | Sputum | 0.6 (0.6-0.7)* | <0.001 |  |  |
|  | Chest pain in last 1 month | 0.6 (0.6-0.7)* | <0.001 |  |  |
|  | Fever ≥2 weeks | 0.8 (0.8-0.8)* | <0.001 |  |  |
|  | Night sweats | 0.8 (0.7-0.8)* | <0.001 |  |  |
|  | Severe weight loss in last 3 months | 0.7 (0.7-0.8)* | <0.001 |  |  |
|  | Swelling in a lymph node | 1.4 (1.4-1.5)* | <0.001 |  |  |
|  | **Alcohol use** |  |  |  |  |
|  | No | Ref |  |  |  |
|  | Yes | 1.0 (0.9-1.2) | 0.862 |  |  |
|  | **Tobacco use** |  |  |  |  |
|  | No | Ref |  |  |  |
|  | Yes | 0.8 (0.8-0.9)* | <0.001 |  |  |
|  | **Source of referral** |  |  |  |  |
|  | Registered medical practitioner | Ref |  | Ref |  |
|  | Accredited Social Health Activist (ASHA) | 1.3 (1.2-1.4)* | <0.001 | 1.2 (1.1-1.3)* | <0.001 |
|  | Anganwadi worker | 1.3 (1.1-1.6) | 0.003 | 1.2 (1.0-1.5)* | 0.025 |
|  | Community practitioner | 1.4 (1.3-1.5)* | <0.001 | 1.3 (1.2-1.4)* | <0.001 |
| Ismail 2020 (Karnataka)^a^ [5] |  | Values below are relative risk ratios |  | Values below are adjusted relative risk ratios |  |
|  | **Age (years)** |  |  |  |  |
|  | 35-44 | Ref |  | Ref |  |
|  | <15 | 1.19 (0.85-1.67) | Not reported | 1.07 (0.77-1.5) | 0.66 |
|  | 15-24 | 0.94 (0.69-1.28) | Not reported | 0.93 (0.69-1.25) | 0.651 |
|  | 25-34 | 1.17 (0.91-1.51) | Not reported | 1.16 (0.91-1.48) | 0.211 |
|  | 45-54 | 0.97 (0.77-1.23) | Not reported | 1.0 (0.80-1.26) | 0.951 |
|  | 55-64 | 0.97 (0.76-1.22) | Not reported | 0.97 (0.77-1.22) | 0.82 |
|  | >=65 | 0.92 (0.73-1.16) | Not reported | 0.88 (0.69-1.10) | 0.276 |
|  | **Sex** |  |  |  |  |
|  | Male | Ref |  | Ref |  |
|  | Female | 1.05 (0.91-1.20) | Not reported | 1.04 (0.91-1.19) | 0.521 |
|  | Transgender | 2.35 (0.68-8.17) | Not reported | 1.97 (0.54-7.19) | 0.301 |
|  | **Contact of TB case** |  |  |  |  |
|  | No | Ref |  | Ref |  |
|  | Yes | 0.4 (0.17-0.95) | Not reported | 0.42 (0.17-1.00) | 0.052 |
|  | **Diabetes** |  |  |  |  |
|  | No | Ref |  | Ref |  |
|  | Yes | 0.51 (0.28-0.92) | Not reported | 0.76 (0.42-1.37) | 0.368 |
|  | **HIV** |  |  |  |  |
|  | Negative | Ref |  | Ref |  |
|  | Unknown | 2.27 (1.95-2.65)* | Not reported | 2.06 (1.76-2.41)* | <0.001 |
|  | Positive | 1.79 (0.97-3.27) | Not reported | 1.79 (0.97-3.28) | 0.059 |
|  | **Tobacco use** |  |  |  |  |
|  | No | Ref |  | Ref |  |
|  | Yes | 0.65 (0.42-1.01) | Not reported | 0.9 (0.91-1.39) | 0.665 |
|  | **Enrollment center** |  |  |  |  |
|  | Peripheral health institute | Ref |  | Ref |  |
|  | District | 1.6 (1.03-2.48) | Not reported | 1.41 (0.91-2.18) | 0.122 |
|  | Private chemist | 2.87 (0.84-9.77) | Not reported | 1.62 (0.45-5.83) | 0.459 |
|  | Private health facility | 2.09 (1.46-2.97)* | Not reported | 1.86 (1.31-2.65)* | <0.001 |
|  | Private lab | 0.76 (0.11-5.09) | Not reported | 0.62 (0.09-4.16) | 0.625 |
|  | TB unit | 4.34 (3.59-5.25)* | Not reported | 3.66 (3.0-4.42)* | <0.001 |
| Non-completion of sputum microscopy evaluation |  |  |  |  |  |
| Balasubramanian 2004 (Tamil Nadu)^b^ [6] |  | Values below are odds ratios |  |  |  |
|  | **Age (years)** |  |  |  |  |
|  | 15-24 | Ref |  |  |  |
|  | 25-34 | 0.93 (0.77-1.14) | 0.487 |  |  |
|  | 35-44 | 0.96 (0.79-1.16) | 0.644 |  |  |
|  | 45-54 | 0.78 (0.64-0.95) | 0.013 |  |  |
|  | 55-64 | 0.85 (0.70-1.04) | 0.106 |  |  |
|  | 65+ | 0.89 (0.71-1.12) | 0.322 |  |  |
|  | **Sex** |  |  |  |  |
|  | Female | Ref |  |  |  |
|  | Male | 1.15 (1.02-1.30)* | 0.024 |  |  |
| Dandona 2004 (Multi-site, 8 Indian districts)^c^ [8] |  |  |  | Values below are adjusted odds ratios |  |
|  | **Age (years)** |  |  |  |  |
|  | 16-30 |  |  | Ref |  |
|  | 31-50 |  |  | 0.99 (0.68-1.45) | Not reported |
|  | >50 |  |  | 2.07 (1.41-3.04)* | Not reported |
|  | **Sex** |  |  |  |  |
|  | Male |  |  | Ref |  |
|  | Female |  |  | 1.17 (0.88-1.56) | Not reported |
|  | **Marital status** |  |  |  |  |
|  | Others (separated/widowed) |  |  | Ref |  |
|  | Never married |  |  | 2.63 (1.12-6.16)* | Not reported |
|  | Married |  |  | 2.67 (1.31-5.45)* | Not reported |
|  | **Literacy** |  |  |  |  |
|  | Illiterate |  |  | Ref |  |
|  | Literate |  |  | 1.09 (0.82-1.45) | Not reported |
|  | **Monthly family income (Rs)** |  |  |  |  |
|  | <=3000 |  |  | Ref |  |
|  | 3001-5000 |  |  | 0.99 (0.60-1.64) | Not reported |
|  | >5000 |  |  | 1.43 (0.89-2.29) | Not reported |
|  | **Duration of symptoms** |  |  |  |  |
|  | >15 days |  |  | Ref |  |
|  | <=15 days |  |  | 3.02 (2.31-3.95)* | Not reported |
|  | **Family-related factors** |  |  |  |  |
|  | Person accompanying patient to clinic visit |  |  |  |  |
|  | Health worker |  |  | Ref |  |
|  | None |  |  | 2.33 (1.47-3.68)* | Not reported |
|  | Family/friend |  |  | 0.86 (0.54-1.38) | Not reported |
|  | **Person informed of suspicion of TB by health personnel** |  |  |  |  |
|  | Yes |  |  | Ref |  |
|  | No |  |  | 7.75 (5.66-10.60)* | Not reported |
|  | Do not remember |  |  | 4.83 (2.24-10.39)* | Not reported |
| Das 2019 (Chhattisgarh)^b^ [9] |  | Values below are odds ratios |  |  |  |
|  | **Diagnostic modality—sputum microscopy versus Xpert MTB/RIF** |  |  |  |  |
|  | Post-Xpert MTB/RIF testing | Ref |  |  |  |
|  | Pre-Xpert MTB/RIF testing (testing with sputum microscopy) | 6.68 (3.48-12.79)* | <0.0001 |  |  |
| Tripathy 2013 (Punjab)^b^ [10] |  | Values below are odds ratios |  |  |  |
|  | **Distance to DMC** |  |  |  |  |
|  | Distance <= 10km | Ref |  |  |  |
|  | Distance > 10km | 4.65 (1.65-13.11)* | 0.0036 |  |  |
| Non-completion of chest X-ray |  |  |  |  |  |
| Kanakaraju 2020 (Karnataka)^a,d^ [12] |  | Values below are relative risk ratios |  | Values below are adjusted relative risk ratios |  |
|  | **Age (years)** |  |  |  |  |
|  | 25-34 | Ref |  | Ref |  |
|  | <25 | 1.19 (0.91-1.56) | 0.775 | 1.06 (0.72-1.57) | Not reported |
|  | 35-44 | 1.72 (1.26-2.36)* | 0.281 | 1.23 (0.82-1.85) | Not reported |
|  | 45-54 | 1.47 (1.12-1.93)* | 0.664 | 0.93 (0.63-1.36) | Not reported |
|  | 55-64 | 1.56 (1.19-2.05)* | 0.558 | 1.12 (0.77-1.63) | Not reported |
|  | >64 | 1.54 (1.19-1.99)* | 0.761 | 1.06 (0.75-1.52) | Not reported |
|  | **Sex** |  |  |  |  |
|  | Male | Ref |  | Ref |  |
|  | Female | 1.14 (0.94-1.37) | 0.559 | 1.09 (0.85-1.39) | Not reported |
|  | Transgender | N/A |  | N/A |  |
|  | **Place of residence** |  |  |  |  |
|  | Rural | Ref |  |  |  |
|  | Urban | 0.47 (0.44-0.49)* | N/A |  |  |
|  | **Predominant symptom** |  |  |  |  |
|  | Cough | Ref |  | Ref |  |
|  | Cough and fever | 0.23 (0.18-0.3)* | 0.116 | 0.52 (0.23-1.18) | Not reported |
|  | Hemoptysis | 0.20 (0.16-0.26)* | 0.115 | 0.20 (0.03-1.49) | Not reported |
|  | Not recorded | 2.04 (1.26-3.31)* | 0.985 | 1.01 (0.4-2.56) | Not reported |
|  | **Sputum examination results** |  |  |  |  |
|  | Negative | Ref |  | Ref |  |
|  | Positive | 0.79 (0.61-1.04) | 0.135 | 0.75 (0.50-1.11) | Not reported |
|  | **Designated microscopy center** |  |  |  |  |
|  | District hospital | Ref |  | Ref |  |
|  | Taluka hospital 1 | 0.25 (0.19-0.31)* | 0.047 | 0.44 (0.19-0.99)* | Not reported |
|  | Taluka hospital 2 | 3.03 (1.52-6.06)* | 0.059 | 3.03 (0.94-9.77) | Not reported |
|  | Primary health center 1 | 2.38 (0.78-7.23) | 0.122 | 2.56 (0.79-8.37) | Not reported |
|  | Primary health center 2 | 0 |  | 0 | Not reported |
|  | Primary health center 3 | 0 |  | 0 | Not reported |
|  | Primary health center 4 | 0.52 (0.32-0.82)* | 0.776 | 0.77 (0.31-1.90) | Not reported |
| Sarkar 2011 (West Bengal)^c^ [13] |  | Values below are odds ratios |  | Values below are adjusted odds ratios |  |
|  | **Income** |  |  |  |  |
|  | Above Poverty line | Ref |  |  |  |
|  | Below poverty line | 132.7 (103-7-168.5)* |  |  |  |
|  | **Can afford x-rays from private hospital** |  |  |  |  |
|  | Yes | Ref |  | Ref |  |
|  | No | 63.3(42.8-93.5)* |  | 22.1 (13-37.7)* | Not reported |
|  | **Knew that x-rays are needed for diagnosis** |  |  |  |  |
|  | Yes | Ref |  | Ref |  |
|  | No | 4.2 (2.9-6.1)* |  | 22.7 (8.4-61)* | Not reported |
|  | **Distance from nearest public health facility with x-ray** |  |  |  |  |
|  | <30 km | Ref |  |  |  |
|  | >30 km | 37.7 (29.2-48.7)* |  |  |  |
|  | **Below poverty line and >30 km from nearest public health facility with chest x-ray** |  |  | 82.9 (64.7-106.3)* | Not reported |
| Non-completion of CBNAAT, line probe assay, or mycobacterial culture |  |  |  |  |  |
| Kanakaraju 2020 (Karnataka)^b,d^ [12] |  | Values below are odds ratios |  |  |  |
|  | **Age** |  |  |  |  |
|  | 25-34 | Ref | 0.74 |  |  |
|  | <25 | 1.2031 (0.39-3.68) | 0.42 |  |  |
|  | 35-44 | 1.5780 (0.52-4.76) | 0.75 |  |  |
|  | 45-54 | 0.8594 (0.34-2.18) | 0.86 |  |  |
|  | 55-64 | 0.9297 (0.37-2.32) | 0.72 |  |  |
|  | >64 | 1.1892 (0.47-3.03) |  |  |  |
|  | **Sex** |  |  |  |  |
|  | Male | Ref |  |  |  |
|  | Female | 1.3607 (0.72-2.56) | 0.338 |  |  |
|  | Transgender | 0.03 (0.003-0.22)* | 0.027 |  |  |
|  | **Place of residence** |  |  |  |  |
|  | Urban | Ref |  |  |  |
|  | Rural | 2.78 (1.16-6.65)* | 0.02 |  |  |
|  | **Predominant symptom** |  |  |  |  |
|  | Cough | Ref |  |  |  |
|  | Cough and fever | 0.62 (0.29-1.31) | 0.21 |  |  |
|  | Hemoptysis | 0.14 (0.0056-3.71) | 0.24 |  |  |
|  | Not recorded | 0.44 (0.21-0.93)* | 0.03 |  |  |
|  | **Sputum examination results** |  |  |  |  |
|  | Positive | Ref |  |  |  |
|  | Negative | 102.04 (45.63-228.17)* | 0.0001 |  |  |
|  | **Designated microscopy center** |  |  |  |  |
|  | District hospital | Ref |  |  |  |
|  | Taluka hospital 1 | 0.47 (0.21-1.05) | 0.07 |  |  |
|  | Taluka hospital 2 | 0.30 (0.13-0.73)* | 0.01 |  |  |
|  | Primary health center 1 | 0.37 (0.10-1.45) | 0.15 |  |  |
|  | Primary health center 2 | 2.71 (0.15-48.86) | 0.50 |  |  |
|  | Primary health center 3 | 0.62 (0.07-5.21) | 0.66 |  |  |
|  | Primary health center 4 | 0.38 (0.10-1.50) | 0.17 |  |  |
|  | **Underwent chest radiography** |  |  |  |  |
|  | Yes | Ref |  |  |  |
|  | No | 1.6773 (0.93-3.02) | 0.08 |  |  |
| Shewade 2017 (Madhya Pradesh)^a^ [16] |  | Values below are relative risk ratios |  | Values below are adjusted relative risk ratios |  |
|  | **Age** |  |  |  |  |
|  | <14 | 0.9 (0.5-1.9) | Not reported | 1.0 (0.6-1.9) | Not reported |
|  | 14-44 | 1.0 (0.9-1.2) | Not reported | 1.0 (0.9-1.2) | Not reported |
|  | 45-64 | Ref |  | Ref |  |
|  | >=65 | 1.3 (1.1-1.6)* | Not reported | 1.3 (1.1-1.7)* | Not reported |
|  | **Sex** |  |  |  |  |
|  | Female | Ref |  | Ref |  |
|  | Male | 1.0 (0.9-1.2) | Not reported | 1.1 (1.0-1.2) | Not reported |
|  | **Presumptive MDR-TB criteria** |  |  |  |  |
|  | Follow up smear positive | Ref |  | Ref |  |
|  | Previously treated-recurrent | 1.2 (0.9-1.5)* | Not reported | 1.3 (1.0-1.6)* | Not reported |
|  | Treatment after failure | 1.2 (0.7-1.9) | Not reported | 1.3 (0.8-2.0) | Not reported |
|  | Treatment after LTFU | 1.5 (1.2-2.0)* | Not reported | 1.5 (1.1-2.1)* | Not reported |
|  | Previously treated-others | 1.7 (1.4-2.1)* | Not reported | 1.6 (1.1-2.3)* | Not reported |
|  | New patient with TB-HIV | 1.6 (0.7-3.6) | Not reported | 1.8 (0.8-4.2) | Not reported |
|  | **Site of TB** |  |  |  |  |
|  | Pulmonary smear positive | Ref |  | Ref |  |
|  | Extrapulmonary | 1.6 (1.4-1.9)* | Not reported | 1.5 (1.0-2.2)* | Not reported |
|  | Pulmonary smear negative | 1.4 (1.2-1.6) | Not reported | 1.2 (0.8-1.7) | Not reported |
|  | **Health facility** |  |  |  |  |
|  | District level | Ref |  | Ref |  |
|  | Primary/Secondary level | 1.0 (0.9-1.1) | Not reported | 1.0 (0.9-1.1) | Not reported |
|  | Medical College | 1.2 (1.0-1.4)* | Not reported | 1.2 (1.02-1.4)* | Not reported |
| Shewade 2017 (Tamil Nadu)^a^ [17] |  | Values below are relative risk ratios |  |  |  |
|  | **Age** |  |  |  |  |
|  | 45-64 | Ref |  |  |  |
|  | <14 | 3.6 (0.7-18.7) |  |  |  |
|  | 14-44 | 1.5 (0.5-2.3) |  |  |  |
|  | >=65 | 2.0 (0.8-4.8) |  |  |  |
|  | **Sex** |  |  |  |  |
|  | Male | Ref |  |  |  |
|  | Female | 1.4 (0.9-2.4) |  |  |  |
|  | **Presumptive MDR-TB criteria** |  |  |  |  |
|  | Retreatment- LTFU | Ref |  |  |  |
|  | Retreatment-relapse | 1.2 (0.7-2.2) |  |  |  |
|  | Retreatment- failure | 0.7 (0.2-3.0) |  |  |  |
|  | Retreatment- other | 2.7 (1.5-5.1)* |  |  |  |
|  | Follow up smear positive | 1.0 (0.5-2.3) |  |  |  |
|  | New Patient with TB/HIV | 1.5 (0.7-3.2) |  |  |  |
|  | **Site of TB** |  |  |  |  |
|  | Pulmonary smear positive | Ref |  |  |  |
|  | Pulmonary smear negative | 2.2 (1.3-3.6)* |  |  |  |
|  | Extrapulmonary | 2.9 (1.6-5.4)* |  |  |  |
|  | **Health facility** |  |  |  |  |
|  | Primary/Secondary level | Ref |  |  |  |
|  | District level | 1.5 (0.9-2.4) |  |  |  |
|  | Medical College/other | 2.2 (0.9-5.4) |  |  |  |
| Shewade 2016 (Puducherry)^a^ [19] |  | Values below are relative risk ratios |  |  |  |
|  | **Age** |  |  |  |  |
|  | 45-64 | Ref |  |  |  |
|  | <14 | None |  |  |  |
|  | 14-44 | 1.1 (0.7-1.8) |  |  |  |
|  | >=65 | 0.3 (0.04-1.9) |  |  |  |
|  | **Sex** |  |  |  |  |
|  | Male | Ref |  |  |  |
|  | Female | 1.6 (0.9-2.7) |  |  |  |
|  | **Presumptive MDR-TB criteria** |  |  |  |  |
|  | Retreatment | Ref |  |  |  |
|  | Follow up smear positive | 1.4 (0.8-2.4) |  |  |  |
|  | New Patient with TB/HIV | 3.6 (2.0-6.5)* |  |  |  |
|  | **Sputum status** |  |  |  |  |
|  | Smear-positive pulmonary | Ref |  |  |  |
|  | Smear-negative pulmonary | 3.0 (1.1-8.3)* |  |  |  |
|  | Smear-negative extrapulmonary | 5.6 (4.0-7.8)* |  |  |  |
|  | **Health facility** |  |  |  |  |
|  | District level | Ref |  |  |  |
|  | Primary/Secondary level | 1.4 (0.6-3.0) |  |  |  |
|  | Medical College | 1.7 (0.9-3.1) |  |  |  |
|  | Other facilities | 4.2 (2.6-6.9)* |  |  |  |
| Shewade 2015 (Puducherry)^a^ [18] |  | Values below are relative risk ratios |  |  |  |
|  | **Age** |  |  |  |  |
|  | 45-64 | Ref |  |  |  |
|  | <14 | None |  |  |  |
|  | 14-44 | 1.08 (0.84-1.39) |  |  |  |
|  | >=65 | 1.38 (0.93-2.04) |  |  |  |
|  | **Sex** |  |  |  |  |
|  | Male | Ref |  |  |  |
|  | Female | 1.46 (1.13-1.88)* |  |  |  |
|  | **Presumptive MDR-TB criteria** |  |  |  |  |
|  | Retreatment | Ref |  |  |  |
|  | Follow-up smear positive | 1.05 (0.82-1.36) |  |  |  |
|  | New patient with TB/HIV | 1.71 (1.26-2.31)* |  |  |  |
|  | **Extrapulmonary TB** |  |  |  |  |
|  | No | Ref |  |  |  |
|  | Yes | 2.31 (1.97-2.73)* |  |  |  |
|  | **Health facility** |  |  |  |  |
|  | District level | Ref |  |  |  |
|  | PHC/CHC | 1.80 (1.35-2.40)* |  |  |  |
|  | Medical College | 1.59 (1.19-3.27)* |  |  |  |
|  | Other facilities | 2.43 (1.81-3.27)* |  |  |  |
| Singla 2014 (Delhi)^b^ [20] |  | Values below are odds ratios |  |  |  |
|  | **Line-probe assay** |  |  |  |  |
|  | Post-line probe assay availability | Ref |  |  |  |
|  | Pre-line probe assay availability | 6.94 (5.49-8.76)* | <0.0001 |  |  |

CBNAAT, cartridge-based nucleic acid amplification test

^a^Effect estimates comprise relative risk or adjusted relative risk ratios

^b^Unadjusted odds ratios and p-values were estimated by the systematic review team from the raw data, as these were not provided in the original study.

^c^Effect estimates comprise odds ratios or adjusted odds ratios

^d^Study reported completing the workup or test as the outcome, so effect estimates (odds ratio or relative risk and 95% confidence interval) were flipped to show effect estimates for the outcome of not completing the diagnostic test or workup.
