## Supplemental Appendix 3 for "Barriers to engagement by people with active tuberculosis in the care cascade in India: a systematic review of two decades of quantitative research"

**Supplementary Appendix 3**

**Supplement to:**

Barriers to engagement by people with active tuberculosis in the care cascade in India: a systematic review of two decades of quantitative research

Tulip A. Jhaveri, Disha Jhaveri, Amith Galivanche, Dominic Voehler, Maya Lubeck-Schricker, Mei Chung, Pruthu Thekkur, Vineet Chadha, Ruvandhi Nathavitharana, Ajay M.V. Kumar, Hemant Deepak Shewade, Katherine Powers, Kenneth H. Mayer, Jessica E. Haberer, Paul Bain, Madhukar Pai, Srinath Satyanarayana, Ramnath Subbaraman

**Correspondence:**

Ramnath Subbaraman, MD, MSc, FACP

Tufts University School of Medicine

Department of Public Health and Community Medicine

136 Harrison Ave., MV237

Boston, MA 02130, USA

**Methods and findings for the systematic review of barriers to patients diagnosed with TB getting successfully started on, or registered in, treatment (Gap 3)**

### Methods

##### Objectives

The objective of this systematic review is to understand why some patients who are diagnosed with TB—in either public or private sector facilities in India—do not get successfully registered in treatment, a problem also known as pretreatment loss to follow-up (PTLFU) (Gap 3 in the care cascade). We broadly defined patients diagnosed with TB as comprising individuals identified by India’s National TB Elimination Program (NTEP) or private sector healthcare providers as having active TB based on a microbiological diagnosis (e.g., positive sputum smear microscopy result, positive Xpert MTB/RIF result, or histological finding) or empirical diagnosis (e.g., based on consistent clinical symptoms, clinical findings, or findings on a non-specific test such as chest X-ray).

We searched for studies that followed such patients after their diagnosis to determine whether they subsequently started treatment for active TB and, ideally, got notified (or registered) as being on treatment to India’s NTEP. When such information is available, we use notification or registration to define a successful outcome, as notification indicates that the patient has been reported to the NTEP as having been started on treatment, such that healthcare providers are accountable for the patient’s treatment outcome [1].

We extracted two types of quantitative findings that help to understand PTLFU:

(a) *Factors associated with PTLFU in regression analyses:* For studies comparing patients who did or did not start on, or get registered in, TB treatment, we extracted effect estimates for independent variables (i.e., exposures or predictors) associated with PTLFU. Effect estimates included odds ratios, risk ratios, hazard ratios, or beta-coefficients, depending on the approach to analysis.
(b) *Reasons reported by patients for PTLFU*: For studies that surveyed patients who experienced PTLFU regarding reasons they had not started on, or registered in, TB treatment, we extracted the proportion of patients who reported a given reason for PTLFU.

###### Table A. Search strategy to identify manuscripts regarding diagnosed TB patients not getting successfully started on, or registered in, treatment (Gap 3). This same search was also used to identify relevant articles for Gaps 2 and 4.

| Terms for tuberculosis: | “tuberculosis”[Mesh] OR *Mycobacterium tuberculosis*[tiab] OR TB[tiab] OR MDRTB[tiab] OR XDRTB[tiab] |
| --- | --- |
| Terms for India: | “India”[Mesh] OR India[tiab] OR India[ad] OR Indian[tiab] OR Indians[tiab] |
| Terms for loss to follow-up or other poor outcomes: | “patient dropouts”[tiab] OR “treatment refusal”[Mesh] OR “patient compliance”[Mesh] OR lost to follow up[tiab] OR loss to follow up[tiab] OR default*[tiab] OR compliance[tiab] OR adherence[tiab] OR noncompliance[tiab] OR nonadherence[tiab] OR diagnostic dropout [tiab] OR patient cooperation[tiab] OR dropout*[tiab] OR linkage to care[tiab] OR retention[tiab] OR attrition[tiab] OR cascade of care[tiab] OR treatment cascade[tiab] OR treatment success*[tiab] OR treatment completion[tiab] OR cure[tiab] OR pretreatment loss to follow-up[tiab] OR initial default[tiab]; treatment failure [tiab] |

##### Inclusion and exclusion criteria

We applied the following criteria for inclusion and exclusion of studies for this systematic review.

*Inclusion criteria* included the following:

(1) Studies that followed patients who were diagnosed with any form of active TB to evaluate whether these patients got successfully started on, or registered in, treatment.
(2) Studies also had to have assessed reasons that individuals in the study did not get started on, or registered in, treatment. These studies could have compared characteristics of those who did or did not experience PTLFU (e.g., regression analyses) or conducted structured interviews with patients who experienced PTLFU to understand reasons for this outcome.

*Exclusion criteria* included the following:

(1) Studies that only described the proportion of patients who experienced PTLFU without stating reasons for not starting on, or registering in, treatment.
(2) Studies with data collected prior to the year 2000, as India’s Revised National TB Control Programme (now called the NTEP) did not achieve nationwide coverage until the early 2000s.
(3) Studies only containing qualitative data evaluating on PTLFU. Findings from studies containing qualitative data will be reported in a separate paper.

We classified the health facility-based strategy for sampling patients in each study based on whether it was comprehensive, random, or convenience. Studies using convenience sampling were excluded from analysis. Studies that assessed more than 150 patients and at over one government DMC were rated as being higher in quality than studies that assessed less than 150 patients at a single government DMC. Studies that assessed PTLFU within a shorter time frame after diagnosis (e.g., within 1-2 months) were rated as being higher in quality than studies that assessed this outcome 3 or more months after diagnosis, since tracking patients to find out accurate outcomes becomes much harder with time due to patient mobility. Studies in which a dedicated research team was used to track PTLFU patients were rated as being higher in quality than studies that relied on self-report by local TB programs to evaluate PTLFU.

###### Table B. Criteria for assessing quality of quantitative studies evaluating failure of diagnosed TB patients not getting successfully registered in treatment.

| **Criterion** | **Quality level** |
| --- | --- |
| **Sampling strategy** | |
| Random or comprehensive sampling at selected facilities | High |
| Convenience sampling or not reported | Low (exclude findings from analysis) |
| **Sample size** | |
| >1 center and 150+ patients | High |
| Single center study with 150+ patients | Medium |
| <150 patients or not reported | Low |
| **Time frame of research fieldwork after patient diagnosis** | |
| 2 weeks to 1 month after diagnosis | High |
| 1-3 months after diagnosis | Medium |
| >3 months after diagnosis or not reported | Low |
| **Method of evaluating outcome** | |
| Patient tracking by a dedicated research team | High |
| Relying on self-report by government TB program or not reported | Medium to Low |

##### Data extraction and analysis

Multiple reviewers (TJ, AG, DV, MLS, and DJ) independently extracted data from each included study into a structured form on an Excel spreadsheet; however, we ensured that every article had data independently extracted by at least two or more reviewers. Disagreements were resolved by discussion or, if necessary, by consulting a supervising reviewer (RS). From each study, we extracted information on the study design, location, setting (i.e., urban versus rural), sample size, and variables of interest (Table C). We also divided studies based on whether they evaluated PTLFU for patients with drug-susceptible versus drug-resistant TB.

For studies that compared patients who did or did not complete PTLFU, we extracted adjusted and unadjusted effect estimates (odds ratios, risk ratios, hazard ratios, and beta-coefficients) from regression analyses. For studies that did not report effect estimates, we calculated unadjusted odds ratios from the data provided, if possible. For studies that reported reasons why patients experienced PTLFU, we extracted the proportion of patients surveyed who reported a given reason for PTLFU. Of note, some studies considered initiation of TB treatment but where reporting was delayed or who initiated treatment at other government centers or in the private sector to be a negative outcome; however, we considered treatment initiation in such situations to be a potentially successful outcome. As such, for such studies, we modified the denominator of patients surveyed to exclude those who had initiated treatment in these other scenarios. We also excluded patients who had positive sputum smear microscopy results as part of treatment follow-up from the denominator. For effect estimates and proportions, we extracted information on 95% confidence intervals (95% CIs) where available; if 95% CIs were not reported, we calculated these from the data provided, if possible. We also recalculated 95% Cis for studies in which the denominator had been modified to excluded patients who had positive sputum smear microscopy results as part of treatment follow-up, or who initiated treatment with delayed reporting, at other government sites, or in the private sector.

For some variables, we also changed the reference group as needed for consistency of reporting across studies. For example, because most studies compared men to the reference group of women, we “flipped” effect estimates and confidence intervals for studies that presented men as the reference group. This allowed us to consistently present women as the reference group for findings regarding gender.

After “flipping” selected effect estimates from the regression analyses, we reported all unadjusted and adjusted effect estimates, regardless of statistical significance, organized by study (Table D). For the main manuscript and Forest plot, we restricted ourselves to presenting statistically significant adjusted effect estimates from multivariable analyses, as these may represent more meaningful associations from higher-quality analyses. After extracting this subset of findings, we organized findings into categories using our framework of demand- and supply-side factors (main manuscript, Table 1). For findings on reasons why patients experienced PTLFU, we presented all reported proportions in the Forest Plots in the main manuscript; we also organized these findings using our framework of demand- and supply-side categories.

To visualize quantitative findings, we generated Forest plots of effect estimates odds ratios, risk ratios, hazard ratios, beta-coefficients, and proportions using Stata version 16.1 (College Station, TX, USA). We did not conduct meta-analyses of data, because we extracted findings represented a diverse set of variables from every study.

###### Table C. Characteristics of the included studies for patients diagnosed with tuberculosis who did not start on, or get registered in, treatment (Gap 3)

| **Citation** (year) | **Location** | **Urban, rural, both, or unknown** | **Type of population** (type of TB or other aspects of the patient population) | **Single or multiple designated microscopy centers (DMCs)** | **Sample size** (overall sample studied to identify PTLFU patients) | **Time frame of research fieldwork after patient diagnosis** | **Methodology** (Patient tracking by a dedicated research team versus self-report by the government TB program) | **Type of findings included in the study**  (N=sample or denominator of patients for given analysis) |
| --- | --- | --- | --- | --- | --- | --- | --- | --- |
| Drug-susceptible adult TB patients |  |  |  |  |  |  |  |  |
| Ahmed (2009) [3] | Karnataka | Rural | General population (mostly drug-susceptible TB) | Multiple | 232 | 0-4 weeks | Self-report by the government TB program* | Logistic regression (N=232) (only summary of findings presented) |
| Balasubramanian (2004) [4] | Tamil Nadu | Rural | General population (mostly drug-susceptible TB) | Multiple | 1,064 | 1-3 months* | Both | Logistic regression (N=1,064)^a^ |
| Dandona (2004) [5] | Andhra Pradesh, Maharashtra, Rajasthan, Tamil Nadu | Both | General population (mostly drug-susceptible TB) | Multiple | 3,994 | Not reported | Patient tracking by a dedicated research team | Logistic regression (N=3,994)^a^  Reasons for PTLFU (proportions) (N=43)^b^ |
| Das (2019) [6] | Chhattisgarh | Rural | General population (mostly drug-susceptible TB); Conflict-affected border areas | Multiple | 297 | 0-4 weeks | Patient tracking by a dedicated research team | Logistic regression (N=297)^a^ |
| Dave (2013) [7] | Gujarat | Both | General population (mostly drug-susceptible TB) | Multiple | 3,159 | Not reported | Patient tracking by a dedicated research team | Reasons for PTLFU (proportions) (N=2,494)^b^ |
| Gopi (2005) [8] | Tamil Nadu | Rural | General population (mostly drug-susceptible TB) | Multiple | 1,292 | 1-3 months* | Patient tracking by a dedicated research team | Logistic regression (N=1,292)^a^  Reasons for PTLFU (N=200 of whom 90 were interviewed)^b^ |
| Ismail (2020) [9] | Karnataka | Both | General population (mostly drug-susceptible TB) | Multiple | 822 | >3 months* | Patient tracking by a dedicated research team | Relative risk regression (N=822) |
| Khandekar (2013) [10] | Delhi | Urban | General population (mostly drug-susceptible TB) | Single* | 1,361 | Not reported | Patient tracking by a dedicated research team | Logistic regression (N=1,283)^a^ |
| Kumar (2013) [11] | Delhi | Urban | General population (mostly drug-susceptible TB) | Single* | 184 | 1-3 months* | Self-report by the government TB program* | Logistic regression (N=184)^a^ |
| Mandal (2014) [12] | West Bengal | Both | General population (mostly drug-susceptible TB) | Multiple | 132* | 0-4 weeks | Patient tracking by a dedicated research team | Reasons for PTLFU (proportions) (N=132) |
| Mehra (2013) [13] | Uttarakhand | Urban | General population (mostly drug-susceptible TB) | Single* | 555 | 0-4 weeks | Patient tracking by a dedicated research team | Logistic regression (N=555)^a^  Reasons for PTLFU (proportions) (N=98) |
| Pardeshi (2018) [14] | Multi-state (representative population-based sample across 29 states and 7 union territories) | Both | General population (mostly drug-susceptible TB) | Multiple | 5,607 | Not reported | Self-report by the government TB program* | Logistic regression (N=5,607) |
| Pillai (2015) [15] | Puducherry | Urban | General population (mostly drug-susceptible TB) | Multiple | 950 | Not reported | Patient tracking by a dedicated research team | Reasons for PTLFU (proportions) (N=20) |
| Rawat (2012) [16] | Uttarakhand | Rural | General population (mostly drug-susceptible TB) | Single* | 466 | Not reported | Self-report by the government TB program* | Reasons for PTLFU (proportions) (N=21) (only summary of findings presented for some reasons) |
| Sai Babu (2008) [17] | Andhra Pradesh and Telangana | Both | General population (mostly drug-susceptible TB) | Multiple | 15,631 | Not reported | Patient tracking by a dedicated research team | Reasons for PTLFU (proportions) (N=552)^b^ |
| Singh (2020) [18] | Uttarakhand | Both | General population (mostly drug-susceptible TB) | Multiple | 256 | Not reported | Self-report by the government TB program* | Logistic regression (N=256)^a^ |
| Thomas (2018) [1] | Tamil Nadu | Urban | General population (mostly drug-susceptible TB) | Multiple | 344 | 0-4 weeks | Patient tracking by a dedicated research team | Logistic regression (N=344)  Reasons for PTLFU (proportions) (N=76) |
| Tripathy (2013) [19] | Punjab | Rural | General population (mostly drug-susceptible TB) | Multiple | 156 | Not reported | Self-report by the government TB program* | Logistic regression (N=156)^a^ |
| Drug-resistant adult TB patients |  |  |  |  |  |  |  |  |
| Chadha (2011) [20] | Andhra Pradesh | Both | Drug-resistant TB patients | Multiple | 169 | >3 months* | Self-report by the government TB program* | Reasons for PTLFU (proportions) (N=48)^b^ |
| Jain (2018) [21] | Madhya Pradesh | Urban | Drug-resistant TB patients | Single* | 474 | 0-4 weeks | Patient tracking by a dedicated research team | Logistic regression (N=474) (only summary of findings presented) |
| Natrajan (2018) [22] | Madhya Pradesh | Both | Drug-resistant TB patients | Multiple | 74* | Not reported | Patient tracking by a dedicated research team | Reasons for PTLFU (proportions) (N=6)^b^ |
| Shewade (2017) [23] | Tamil Nadu | Urban | Drug-resistant TB patients | Multiple | 74 | Not reported | Patient tracking by a dedicated research team | Relative risk regression (N=74) |
| Shewade (2018) [24] | Gujarat | Both | Drug-resistant TB patients | Multiple | 257 | >3 months* | Patient tracking by a dedicated research team | Relative risk regression (N=257) |
| Singla (2014) [25] | Delhi | Urban | Drug-resistant TB patients | Single* | 688 | Not reported | Patient tracking by a dedicated research team | Logistic regression (N=688)^a^ |
| Pediatric TB patients |  |  |  |  |  |  |  |  |
| Raizada (2018) [26] | Tamil Nadu, Delhi, Andhra Pradesh, West Bengal | Urban | Children (0-14 years) with drug-susceptible and drug-resistant TB | Multiple | 3,340 | Not reported | Patient tracking by a dedicated research team | Logistic regression for drug-susceptible TB (N=3,045)^a^  Logistic regression for drug-resistant TB (N=295)^a^ |

PTLFU, pretreatment loss to follow-up; TB, tuberculosis,

*Medium or low quality for this indicator

^a^Unadjusted odds ratios and p-values were estimated by the systematic review team from the raw data, as these were not provided in the original study.

^b^Denominator reported here varies from what is reported in the original study, as we excluded patients from the denominator who had initiated treatment with delayed reporting or who initiated treatment at other government sites or in the private sector, as these represented potentially successful outcomes. We also excluded patients who had positive sputum smear microscopy results as part of follow-up testing while already on treatment.

###### Table D. Factors associated with patients diagnosed with tuberculosis not getting started on, or registered in, treatment (Gap 3)

| Study | Exposure / Independent variable | Unadjusted Effect Estimate (95% Confidence Interval) | P-value | Adjusted Effect Estimate (95% Confidence Interval) | P-value |
| --- | --- | --- | --- | --- | --- |
| Drug-susceptible TB |  |  |  |  |  |
| Ahmed 2009 (Karnataka) [3] |  |  |  |  |  |
|  | **Sex** | Study reported higher PTLFU among men (23%) than women (9%) but data not provided to estimate odds ratio. | 0.004* |  |  |
| Balasubramanian 2004 (Tamil Nadu)^a,b^ [4] |  | Values below are odds ratios |  |  |  |
|  | **Sex** |  |  |  |  |
|  | Female | Ref |  |  |  |
|  | Male | 1.11 (0.77-1.67) | 0.6 |  |  |
|  | **Age (Years)** |  | Note: paper reports p=0.02 using Chi-Squared for trend for age |  |  |
|  | 15-24 | Ref |  |  |  |
|  | 25-34 | 0.74 (0.35-1.57) | 0.44 |  |  |
|  | 35-44 | 1.57 (0.80-3.06) | 0.19 |  |  |
|  | 45-54 | 1.51 (0.78-2.95) | 0.22 |  |  |
|  | 55-64 | 1.75 (0.89-3.44) | 0.11 |  |  |
|  | 65 and above | 2.65 (1.27-5.51)* | 0.009* |  |  |
| Dandona 2004 (multi-state)^a,b^ [5] |  | Values below are odds ratios |  |  |  |
|  | **Sex** |  |  |  |  |
|  | Female | Ref |  |  |  |
|  | Male | 1.27 (0.91-1.78) | 0.16 |  |  |
|  | **Age (Years)** |  |  |  |  |
|  | 16-30 | Ref |  |  |  |
|  | 31-50 | 0.88 (0.6-1.29) | 0.51 |  |  |
|  | Over 50 | 1.13 (0.75-1.71) | 0.54 |  |  |
|  | **Marital status** |  |  |  |  |
|  | Never married | Ref |  |  |  |
|  | Married | 1.39 (0.82-2.36) | 0.22 |  |  |
|  | Others | 0.94 (0.40-2.23) | 0.90 |  |  |
|  | **Literacy** |  |  |  |  |
|  | Literate | Ref |  |  |  |
|  | Illiterate | 0.86 (0.62-1.18) | 0.35 |  |  |
|  | **Income** |  |  |  |  |
|  | <3000 monthly | Ref |  |  |  |
|  | 3001-5000 monthly | 0.81 (0.42-1.56) | 0.53 |  |  |
|  | >5000 monthly | 1.69 (0.96-2.99) | 0.72 |  |  |
|  | **Who informed patient of diagnosis** |  |  |  |  |
|  | Health personnel other than doctor | Ref |  |  |  |
|  | Doctor | 1.68 (0.75-3.74) |  |  |  |
|  | Patient did not collect report | 15697.8 (887.92-277524.8)* | <0.0001* |  |  |
|  | Family member | 11.16 (4.79-26.01)* | <0.0001* |  |  |
| Das 2019 (Chhattisgarh)^a,b^ [6] |  | Values below are odds ratios |  |  |  |
|  | **Availability of GenXpert** |  |  |  |  |
|  | Post GenXpert | Ref |  |  |  |
|  | Pre GenXpert | 1.94 (0.43-8.84) | 0.39 |  |  |
| Gopi 2005 (Tamil Nadu)^a,b^ [8] |  | Values below are odds ratios |  |  |  |
|  | **Sex** |  |  |  |  |
|  | Female | Ref |  |  |  |
|  | Male | 2.17 (1.42-3.31)* | 0.0003* |  |  |
|  | **Age** |  |  |  |  |
|  | 15-44 | Ref |  |  |  |
|  | 45 and above | 1.55 (1.15-2.7)* | 0.004* |  |  |
|  | **Case finding approach** |  |  |  |  |
|  | Passive case finding (i.e., diagnosed in health facility) | Ref |  |  |  |
|  | Active case finding (i.e., diagnosed in community survey) | 1.75 (1.25-2.47)* | 0.001* |  |  |
| Ismail 2020 (Karnataka)^c^ [9] |  | Values below are relative risk ratios |  | Values below are adjusted relative risk ratios |  |
|  | **Sex** |  |  |  |  |
|  | Male | Ref |  | Ref |  |
|  | Female | 1.34 (0.62-2.93) |  | 1.38 (0.67-2.85) | 0.374 |
|  | Transgender | N/A |  | N/A |  |
|  | **Age (Years)** |  |  |  |  |
|  | 35-44 | Ref |  | Ref |  |
|  | <15 | N/A |  | N/A |  |
|  | 15-24 | 0.8 (0.19-3.27) |  | 0.74 (0.17-3.23) | 0.691 |
|  | 25-34 | 1.12 (0.33-3.80) |  | 1.21 (0.32-4.58) | 0.775 |
|  | 45-54 | 0.87 (0.25-2.97) |  | 1.05 (0.32-3.45) | 0.94 |
|  | 55-64 | 0.76 (0.18-3.11) |  | 0.88 (0.20-3.73) | 0.87 |
|  | 65 and above | 1.43 (0.42-4.83) |  | 1.46 (0.36-5.81) | 0.59 |
|  | **Type of TB** |  |  |  |  |
|  | New | Ref |  | Ref |  |
|  | Recurrent | 3.04 (1.55-7.78)* |  | 3.2 (1.38-7.46)* | 0.007* |
|  | **Contact of TB case** |  |  |  |  |
|  | No | Ref |  | Ref |  |
|  | Yes | 1.33 (0.18-9.41) |  | 0.83 (0.85-8.24) | 0.88 |
|  | **Diabetes mellitus** |  |  |  |  |
|  | No | Ref |  | Ref |  |
|  | Yes | 0.26 (0.03-1.93) |  | 0.33 (0.04-2.7) | 0.304 |
|  | **Type of enrollment center** |  |  |  |  |
|  | Peripheral health institute | Ref |  | Ref |  |
|  | District | Not reported |  | Not reported | Not reported |
|  | Private chemist | Not reported |  | Not reported | Not reported |
|  | Private health facility | 3.01 (1.26-7.22)* |  | 2.73 (1.08-6.9)* | 0.034* |
|  | Private lab | 5.65 (1.40-22.70)* |  | 5.12 (1.38-18.92)* | 0.014* |
|  | TB unit | Not reported |  | Not reported | Not reported |
|  | Unknown | Not reported |  | Not reported | Not reported |
| Khandekar 2013 (Delhi)^a,b^ [10] |  | Values below are odds ratios |  |  |  |
|  | **Sex** |  |  |  |  |
|  | Female | Ref |  |  |  |
|  | Male | 1.03 (0.8-1.31) | 0.83 |  |  |
|  | **Age (Years)** |  |  |  |  |
|  | 0-14 | Ref |  |  |  |
|  | 15-24 | 0.75 (0.54-1.03) | 0.07 |  |  |
|  | 25-44 | 0.87 (0.64-1.2) | 0.4 |  |  |
|  | 45-59 | 0.71 (0.43-1.2) | 0.2 |  |  |
|  | 60 and above | 1.04 (0.55-2.0) | 0.9 |  |  |
|  | **Type of TB** |  |  |  |  |
|  | Pulmonary TB | Ref |  |  |  |
|  | Extrapulmonary TB | 0.82 (0.63-1.06) | 0.14 |  |  |
|  | Both pulmonary and extrapulmonary | 0.90 (0.52-1.57) | 0.71 |  |  |
|  | **Type of patient** |  |  |  |  |
|  | New | Ref |  |  |  |
|  | Relapse | 1.27 (0.51-3.16) | 0.6 |  |  |
|  | Treatment after default | 1.36 (0.25-7.49) | 0.72 |  |  |
|  | Failure | 0.99 (0.31-3.14) | 0.99 |  |  |
|  | Others | 1.25 (0.83-1.88) | 0.28 |  |  |
| Kumar 2013 (Delhi)^a,b^ [11] |  | Values below are odds ratios |  |  |  |
|  | **Sex** |  |  |  |  |
|  | Female | Ref |  |  |  |
|  | Male | 0.65 (0.31-1.34) | 0.24 |  |  |
|  | **Age (Years)** |  |  |  |  |
|  | 0-14 | Ref |  |  |  |
|  | 15-24 | 1.00 (0.11-9.07) | 1 |  |  |
|  | 25-34 | 2.49 (0.29-21.66) | 0.41 |  |  |
|  | 35-44 | 2.21 (0.24-20.35) | 0.49 |  |  |
|  | 45-54 | 2.11 (0.21-21.01) | 0.53 |  |  |
|  | 55-64 | 1.14 (0.09-14.68) | 0.92 |  |  |
|  | 65 and above | 1.60 (0.08-31.77) | 0.76 |  |  |
| Mehra 2013 (Uttarakhand)^a,b^ [13] |  | Values below are odds ratios |  |  |  |
|  | **Sex** |  |  |  |  |
|  | Female | Ref |  |  |  |
|  | Male | 1.00 (0.65-1.55) | 0.99 |  |  |
|  | **Age (Years)** |  |  |  |  |
|  | 40 and above | Ref |  |  |  |
|  | 18-40 | 1.27 (0.86-1.93) | 0.26 |  |  |
|  | **Type of residence** |  |  |  |  |
|  | Urban | Ref |  |  |  |
|  | Rural | 2.72 (1.72-4.30)* | <0.0001* |  |  |
|  | **Education** |  |  |  |  |
|  | Literate | Ref |  |  |  |
|  | Illiterate | 4.4 (2.55-7.61)* | <0.0001* |  |  |
|  | **Socioeconomic status** |  |  |  |  |
|  | Middle | Ref |  |  |  |
|  | Low | 1.35 (0.86-2.10) | 0.19 |  |  |
|  | **Treatment category** |  |  |  |  |
|  | Category II | Ref |  |  |  |
|  | Category I | 1.06 (0.71-1.59) | 0.77 |  |  |
|  | **Smoking status** |  |  |  |  |
|  | Non-smoker | Ref |  |  |  |
|  | Smoker | 1.11 (0.71-1.74) | 0.65 |  |  |
|  | **Smokeless tobacco use status** |  |  |  |  |
|  | Non-user | Ref |  |  |  |
|  | User | 1.96 (1.31-2.95)* | 0.0012* |  |  |
|  | **Alcohol use** |  |  |  |  |
|  | Non-alcoholic | Ref |  |  |  |
|  | Alcoholic | 1.00 (0.66-1.52) | 0.99 |  |  |
| Pardeshi 2018 (multi-state)^a^ [14] |  | Values below are odds ratios |  | Values below are adjusted odds ratios |  |
|  | **Sex** |  |  |  |  |
|  | Male | Ref |  | Ref |  |
|  | Female | 1.38 (1.04-1.84) | 0.03 | 1.24 (0.93-1.67) | 0.15 |
|  | **Age (Years)** |  |  |  |  |
|  | 21-30 | Ref |  | Ref |  |
|  | 0-10 | 3.2 (1.51-6.81)* | 0.002* | 3.43 (1.52-7.77)* | 0.003* |
|  | 11 to 20 | 1.06 (0.47-2.36) | 0.89 | 1.25 (0.56-2.79) | 0.59 |
|  | 31-40 | 1.77 (1.12-2.82)* | 0.02 | 1.67 (1.05-2.65)* | 0.03* |
|  | 41-50 | 1.68 (1.01-2.81)* | 0.04* | 1.50 (0.88-2.57) | 0.14 |
|  | 51-60 | 1.44 (0.89-2.35) | 0.14 | 1.24 (0.70-2.19) | 0.45 |
|  | 60 and above | 1.82 (1.15-2.87)* | 0.01* | 1.59 (0.96-2.63) | 0.07 |
|  | **Type of residence** |  |  |  |  |
|  | Urban | Ref |  | Not included in the multivariable model |  |
|  | Rural | 1.19 (0.82-1.72) | 0.36 | Not included in the multivariable model |  |
|  | **Education** |  |  |  |  |
|  | Secondary | Ref |  | Ref |  |
|  | No education or preschool | 2.42 (1.57-3.69)* | 0.00* | 1.82 (1.10-3.01)* | 0.02* |
|  | Primary | 1.10 (0.61-2.01) | 0.75 | 0.89 (0.45-1.79) | 0.76 |
|  | Higher than secondary | 1.74 (0.88-3.44) | 0.11 | 2.01 (0.91-4.45) | 0.08 |
|  | **Wealth index** |  |  |  |  |
|  | Rich | Ref |  | Ref |  |
|  | Middle | 1.37 (0.76-2.44) | 0.29 | 1.38 (0.73-2.59) | 0.32 |
|  | Poor | 2.0 (1.19-3.37)* | 0.009* | 1.86 (1.01-3.34)* | 0.04* |
|  | **Household structure** |  |  |  |  |
|  | Non-nuclear | Ref |  | Not included in the multivariable model |  |
|  | Nuclear | 0.87 (0.65-1.17) | 0.37 | Not included in the multivariable model |  |
|  | **Type of HCP where household members usually go for other illness** |  |  |  |  |
|  | Public health services | Ref |  | Ref |  |
|  | Other health services | 1.75 (1.31-2.33)* | 0.00* | 1.69 (1.26-2.26)* | 0.00* |
| Singh 2020 (Uttarakhand)^a,b^ [18] |  | Values below are odds ratios |  |  |  |
|  | **Method of Case Finding** |  |  |  |  |
|  | Active Case Finding | 125.3 (7.43-2112.52) | 0.0008* |  |  |
|  | Passive Case Finding | Ref |  |  |  |
| Thomas 2018 (Tamil Nadu)^b^ [1]  *Outcome of this analysis is non-registration in the TB program* |  | Values below are odds ratios |  | Values below are adjusted odds ratios |  |
|  | **Sex** |  |  |  |  |
|  | Male | Ref |  | Ref |  |
|  | Female | 0.88 (0.45-1.72) | 0.7 | 1.33 (0.59-2.88) | 0.49 |
|  | **Age (Years)** |  |  |  |  |
|  | 18-35 | Ref |  | Ref |  |
|  | 36-50 | 1.34 (0.65-2.77) | 0.43 | 0.97 (0.44-2.20) | 0.93 |
|  | 51 and above | 2.88 (1.42-5.85)* | 0.003* | 2.94 (1.40-6.49)* | 0.04* |
|  | **From Chennai** |  |  |  |  |
|  | Yes | Ref |  | Ref |  |
|  | No | 1.25 (0.66-2.37) | 0.48 | 1.18 (0.56-2.38) | 0.66 |
|  | **Trackability** |  |  |  |  |
|  | Probably trackable | Ref |  | Ref |  |
|  | Possibly trackable | 1.78 (1.05-3.04)* | 0.03* | 1.66 (0.91-3.05) | 0.1 |
|  | Untrackable | 3.16 (1.06-9.46)* | 0.04* | 4.49 (1.29-15.06)* | 0.02* |
|  | **Prior TB treatment history** |  |  |  |  |
|  | No prior TB treatment history | Ref |  | Ref |  |
|  | Prior TB treatment history | 3.54 (2.08-6.05)* | <0.0001* | 3.88 (2.15-7.09)* | <0.0001* |
|  | **Site of initial microscopy test** |  |  |  |  |
|  | Moderate or low volume patient microscopy center | Ref |  | Ref |  |
|  | High volume patient microscopy center | 2.91 (1.59-5.30)* | 0.0005* | 3.18 (1.69-6.32)* | 0.0002* |
| Thomas 2018 (Tamil Nadu)^b^ [1]  *Outcome of this analysis is non-initiation of TB therapy* |  | Values below are odds ratios |  | Values below are adjusted odds ratios |  |
|  | **Sex** |  |  |  |  |
|  | Male | Ref |  | Ref |  |
|  | Female | 0.59 (0.22-1.58) | 0.3 | 0.82 (0.26-2.21) | 0.7 |
|  | **Age (Years)** |  |  |  |  |
|  | 18-35 | Ref |  | Ref |  |
|  | 36-50 | 1.42 (0.52-3.89) | 0.49 | 0.99 (0.35-3.09) | 0.99 |
|  | 51 and above | 3.20 (1.23-8.30)* | 0.02* | 2.70 (1.06-7.84)* | 0.04* |
|  | **From Chennai** |  |  |  |  |
|  | Yes | Ref |  | Ref |  |
|  | No | 3.20 (1.57-6.51)* | 0.0013* | 3.01 (1.37-6.52)* | 0.007* |
|  | **Trackability** |  |  |  |  |
|  | Probably trackable | Ref |  | Ref |  |
|  | Possibly trackable | 1.89 (0.94-3.78) | 0.07 | 1.45 (0.68-3.08) | 0.33 |
|  | Untrackable | 3.94 (1.13-13.70)* | 0.03 | 4.53 (1.08-16.52)* | 0.04* |
|  | **Prior TB treatment history** |  |  |  |  |
|  | No prior TB treatment history | Ref |  | Ref |  |
|  | Prior TB treatment history | 1.62 (0.82-3.23) | 0.17 | 1.79 (0.83-3.77) | 0.13 |
|  | **Site of initial microscopy test** |  |  |  |  |
|  | Moderate or low volume patient microscopy center | Ref |  | Ref |  |
|  | High volume patient microscopy center | 2.04 (0.96-4.32) | 0.06 | 2.02 (0.94-4.68) | 0.07 |
| Tripathy 2013 (Punjab)^a,b^ [19] |  | Values below are odds ratios |  |  |  |
|  | **Distance to DMC** |  |  |  |  |
|  | Distance less than 10 KM | Ref |  |  |  |
|  | Distance more than 10 KM | 192.82 (40.96-907.59)* | <0.0001* |  |  |
| Drug-resistant TB |  |  |  |  |  |
| Jain 2018 (Madhya Pradesh)^a,b^ [21] |  |  |  |  |  |
|  | **Distance from drug-resistant TB treatment center** | Study reported that residing a distance of >150 kilometers from the drug-resistant TB treatment center associated with increased risk of PTLFU (i.e., non-initiation of treatment >19 days after diagnosis), but data were not reported to allow calculation of odds ratio or other effect estimates. | 0.009* |  |  |
| Shewade 2017 (Tamil Nadu)^c^ [23] |  | Values below are relative risk ratios |  |  |  |
|  | **Sex** |  |  |  |  |
|  | Female | Ref |  |  |  |
|  | Male | 1.4 (0.6-3.1) |  |  |  |
|  | **Age (Years)** |  |  |  |  |
|  | 45-64 | Ref |  |  |  |
|  | <14 | Not reported (no participants in this category) |  |  |  |
|  | 14-44 | 1.3 (0.7-2.4) |  |  |  |
|  | 65 and above | Not reported (no participants in this category) |  |  |  |
|  | **Presumptive drug-resistant TB criteria** (i.e., criteria for drug-susceptibility testing) |  |  |  |  |
|  | Follow-up smear positive | Ref |  |  |  |
|  | Retreatment | 2.0 (0.5-7.1) |  |  |  |
|  | New patient with TB/HIV | Not reported |  |  |  |
|  | **Site of TB** |  |  |  |  |
|  | Pulmonary smear positive | Ref |  |  |  |
|  | Extrapulmonary TB | Not reported |  |  |  |
|  | Pulmonary smear negative | Not reported |  |  |  |
|  | Pulmonary missing | Not reported |  |  |  |
|  | **Type of health facility** |  |  |  |  |
|  | District level | Ref |  |  |  |
|  | Primary/Secondary | 1.2 (0.5-2.6) |  |  |  |
|  | Medical college/others | Not reported (no participants in this category) |  |  |  |
|  | **Quarter** |  |  |  |  |
|  | Jan-March 2014 | Ref |  |  |  |
|  | April-Jun 2014 | 14.5 (1.9-107.9)* | <0.05* |  |  |
|  | Jul-Sep 2014 | 18.5 (2.6-129.9)* | <0.05* |  |  |
|  | Oct-Dec 2014 | 18.5 (2.6-133.3)* | <0.05* |  |  |
| Shewade 2018 (Gujarat)^c^ [24] |  | Values below are relative risk ratios |  | Values below are adjusted relative risk ratios |  |
|  | **Sex** |  |  |  |  |
|  | Male | Ref |  | Ref |  |
|  | Female | 1.2 (0.5-3.0) |  | 1.2 (0.6-2.8) |  |
|  | **Age (Years)** |  |  |  |  |
|  | 15-44 | Ref |  | Ref |  |
|  | <15 | Not reported (no participants in this category) |  | Not reported (no participants in this category) |  |
|  | 45-64 | 1.0 (0.4-2.8) |  | 1.5 (0.7-3.4) |  |
|  | 65 and above | 1.5 (0.2-9.9) |  | 2.4 (0.3-17.4) |  |
|  | **Presumptive MDR TB criteria** |  |  |  |  |
|  | Retreatment | Ref |  | Ref |  |
|  | Follow-up smear positive | 5.1 (2.1-12.2)* |  | 6.0 (2.3-15.2)* |  |
|  | New patient with TB/HIV | Not reported (no participants in this category) |  | Not reported (no participants in this category) |  |
|  | Pulmonary TB with MDRTB contact | Not reported (no participants in this category) |  | Not reported (no participants in this category) |  |
|  | **Site of TB** |  |  |  |  |
|  | Pulmonary smear positive | Ref |  | Ref |  |
|  | Pulmonary smear missing | 3.2 (1.2-8.3)* |  | 17.1 (7.7-39.3)* |  |
|  | Extrapulmonary TB | Not reported (no participants in this category) |  | Not reported (no participants in this category) |  |
|  | Pulmonary smear negative | Not reported (few participants in this category) |  | Not reported (few participants in this category) |  |
|  | **Health facility** |  |  |  |  |
|  | Primary/Secondary | Ref |  | Ref |  |
|  | District level | Not reported (few participants in this category) |  | Not reported (few participants in this category) |  |
|  | Medical college/others | 2.0 (0.5-7.8) |  | 0.7 (0.3-1.5) |  |
| Singla 2014 (Delhi)^a,b^ [25] |  | Values below are odds ratios |  |  |  |
|  | **Line probe assay** |  |  |  |  |
|  | After line probe assay implementation | Ref |  |  |  |
|  | Before line probe assay implementation | 4.69 (3.15-6.97)* | <0.0001* |  |  |
| Pediatric TB – drug-susceptible |  |  |  |  |  |
| Raizada 2018 (multi-state)^a,b,d^ [26] |  | Values below are odds ratios |  |  |  |
|  | **Sex** |  |  |  |  |
|  | Female | Ref |  |  |  |
|  | Male | 1.11 (0.87-1.41) | 0.41 |  |  |
|  | **Age (Years)** |  |  |  |  |
|  | 10 to 14 | Ref |  |  |  |
|  | 0-4 | 2.81 (2.14-3.68)* | <0.0001* |  |  |
|  | 5 to 9 | 1.33 (0.97-1.82) | 0.08 |  |  |
|  | **Sector** |  |  |  |  |
|  | Public | Ref |  |  |  |
|  | Private | 0.70 (0.44-1.11) | 0.13 |  |  |
| Pediatric TB – drug-resistant |  |  |  |  |  |
| Raizada 2018 (multi-state)^a,b,d^ [26] |  | Values below are odds ratios |  |  |  |
|  | **Sex** |  |  |  |  |
|  | Female | Ref |  |  |  |
|  | Male | 1.56 (0.79-3.10) | 0.20 |  |  |
|  | **Age (Years)** |  |  |  |  |
|  | 10 to 14 | Ref |  |  |  |
|  | 0-4 | 3.33 (1.44-7.73)* | 0.0051* |  |  |
|  | 5 to 9 | 1.49 (0.64-3.49) | 0.37 |  |  |
|  | **Sector** |  |  |  |  |
|  | Public | Ref |  |  |  |
|  | Private | 2.33 (0.87-6.23) | 0.09 |  |  |

^a^Effect estimates comprise odds ratios or adjusted odds ratios.

^b^Unadjusted odds ratios and p-values were estimated by the systematic review team from the raw data, as these were not provided in the original study.

^c^Effect estimates comprise relative risk or adjusted relative risk ratios.

^d^Study reported starting or registering in treatment as the outcome, so effect estimates (odds ratio or relative risk and 95% confidence interval) were flipped to show effect estimates for the outcome of not starting or registering in treatment.

10.	Khandekar J, Acharya AS, R TH, Sharma A. Do patients with tuberculosis referred from a tertiary care referral centre reach their peripheral health institution? Natl Med J India. 2013;26: 332–334.

11.	Kumar S. A retrospective cohort study of the magnitude of initial default among sputum smear-positive TB patients diagnosed at NITRD New Delhi, 4th quarter 2012. New Delhi, India; 2013.
