## Supplemental Appendix 4 for "Barriers to engagement by people with active tuberculosis in the care cascade in India: a systematic review of two decades of quantitative research"

**Supplementary Appendix 4**

**Supplement to:**

Barriers to engagement by people with active tuberculosis in the care cascade in India: a systematic review of two decades of quantitative research

Tulip A. Jhaveri, Disha Jhaveri, Amith Galivanche, Dominic Voehler, Maya Lubeck-Schricker, Mei Chung, Pruthu Thekkur, Vineet Chadha, Ruvandhi Nathavitharana, Ajay M.V. Kumar, Hemant Deepak Shewade, Katherine Powers, Kenneth H. Mayer, Jessica E. Haberer, Paul Bain, Madhukar Pai, Srinath Satyanarayana, Ramnath Subbaraman

**Correspondence:**

Ramnath Subbaraman, MD, MSc, FACP

Tufts University School of Medicine

Department of Public Health and Community Medicine

136 Harrison Ave., MV237

Boston, MA 02130, USA

**Methods and findings for the systematic review of barriers to achieving treatment success in patients diagnosed with active tuberculosis (Gap 4)**

### Methods

##### Objectives

The objective of this systematic review was to understand why some people with TB who start and get registered for TB treatment do not achieve treatment success (Gap 4 in the care cascade). We included patients who started TB treatment either in India’s National TB Elimination Programme (NTEP) or at private sector facilities. We included studies that evaluated patient outcomes starting from TB treatment initiation. Favorable outcomes in this analysis were assumed to include both cure and treatment completion, which together are referred to as “treatment success.” Unfavorable outcomes in this analysis generally comprised the standard case definitions, as defined by India’s NTEP and the World Health Organization (WHO), for the following outcomes during the treatment period: loss to follow-up, treatment failed, died, not evaluated, or a composite of more than one of these. In addition, situations in which studies reported loss to follow-up of less than two months or nonadherence to medications using other measures, we also analyzed these separately as unfavorable outcomes representing medication nonadherence.

Given the large number of studies identified in this review, as well as the heterogeneity in treatment outcomes based on the type of TB, we report our data using subgroups related to drug-resistance profile or proxy measures previously used to presume risk of drug resistance in the NTEP (e.g., “new” patients versus patients with a prior TB treatment history; drug-susceptible versus drug-resistant patients). We also analyzed studies that only included people with HIV or pediatric patients as separate subgroups. We break down our findings as follows:

(a) *New TB patients*: These studies evaluated outcomes among patients with a new diagnosis of active TB disease at any site, including pulmonary or extrapulmonary TB with or without a bacteriological diagnosis. Up until the last few years, “new” patients were often presumed to have drug-susceptible TB and treated with a standardized regimen of isoniazid, rifampin, pyrazinamide, and ethambutol. Note that studies of sputum smear-positive patients treated using a “Category 1” regimen or smear-negative or extrapulmonary patients treated using a “Category 3” regimen, under prior nomenclature, are included in this subgroup.
(b) *Patients with a prior TB treatment history*: These studies evaluated outcomes among patients with a prior TB treatment history at any site, including pulmonary or extrapulmonary TB with or without a bacteriological diagnosis. These patients were categorized separately for a few reasons. Up until about 10 to 15 years ago, these patients were treated in the NTEP using a “Category 2” regimen, which included isoniazid, rifampin, pyrazinamide, and ethambutol, along with the addition of streptomycin by injection. Over the last decade or more, patients with a prior TB treatment history were supposed to undergo drug-susceptibility testing given their elevated risk for having drug-resistant TB. Notably, even with increased drug-susceptibility testing in recent years, these patients continue to have considerably poorer treatment outcomes in the NTEP than new patients.
(c) *Rifampin-resistant or multidrug-resistant (MDR) TB patients*: These studies evaluated outcomes among patients confirmed to have rifampin-resistant TB (generally diagnosed using Xpert MTB/Rif) or MDR TB (generally diagnosed using culture or line probe assay).
(d) *Multiple populations*: These studies evaluated involved cohort studies that included more that one of the subgroups above and did not report outcomes separately for these subgroups.
(e) *People with HIV treated for active TB*: These studies evaluated outcomes among people with HIV who were being treated for active TB disease.
(f) *Pediatric TB patients*: These studies reported outcomes separately for children with TB.

We extracted two types of quantitative findings that help to understand reasons for unfavorable TB treatment outcomes:

(a) *Factors associated with unfavorable treatment outcomes in regression analyses*: For studies comparing individuals who had favorable versus unfavorable outcomes, we extracted effect estimates for independent variables (i.e., exposures or predictors) associated with experiencing an unfavorable treatment outcome. Effect estimates included odds ratios, risk ratios, hazard ratios, or beta-coefficients, depending on the approach to analysis.
(b) *Reasons reported by individuals for experiencing unfavorable treatment outcomes in quantitative surveys*: For studies that surveyed patients who experienced an unfavorable treatment outcome (or family members of patients who had died), we extracted the proportion of individuals who reported a given reason for an unfavorable outcome.

##### Search strategy

Three separate searches were conducted to identify articles. The first search was conducted as part of a previously published study quantifying gaps in India’s TB care cascade [2]. We used articles identified for that review that evaluated Gap 4 in the TB care cascade but that also reported factors and reasons associated with not achieving treatment success. For that review, a medical librarian searched PubMed, Embase, and Web of Science for studies published between January 1, 2000 and February 26, 2015, without language restrictions, using search terms and related variants for “tuberculosis”, “India”, and “loss to follow-up”, including “treatment success” and “treatment failure” to include losses after treatment initiation (Table A). We also carried out electronic searches of key Indian journals that were not indexed for that entire time window: the Indian Journal of Tuberculosis, Lung India, the Indian Journal of Chest and Allied Sciences, the India Journal of Public Health, and the Indian Journal of Community Medicine. Additional studies were identified by searching reference lists of the primary studies and review articles. Notably, given similarities in the search terms for identifying losses during the diagnostic workup (Gap 2), pretreatment loss to follow-up (Gap 3), and poor outcomes on treatment (Gap 4), this single search was used to identify studies related to all of these gaps. We screened all identified studies from this previous review for potential inclusion in our current review; however, studies met inclusion criteria for the current review only if they included information on reasons for not achieving TB treatment success.

To update our review, we conducted a second refresher search using the same search terms for October 2, 2015 to October 1, 2019. We did not repeat hand searches of the Indian journals listed above, because all of these journals had been indexed in PubMed prior to the time period of this more recent search. Due to the extensive time required to extract data from the articles identified for this systematic review, we performed a third refresher search using the same search terms for October 2, 2019 to May 17, 2021. Finally, additional studies were identified by looking through the reference lists of the included primary studies and relevant review articles that were identified by the searches and by outreach to experts in the field.

###### Table A. Search strategy to identify manuscripts regarding tuberculosis (TB) patients not achieving treatment success in India (Gap 4). This same search was also used to identify relevant articles for Gaps 2 and 3.

| Terms for tuberculosis: | “tuberculosis”[Mesh] OR *Mycobacterium tuberculosis*[tiab] OR TB[tiab] OR MDRTB[tiab] OR XDRTB[tiab] |
| --- | --- |
| Terms for India: | “India”[Mesh] OR India[tiab] OR India[ad] OR Indian[tiab] OR Indians[tiab] |
| Terms for loss to follow-up or other poor outcomes: | “patient dropouts”[tiab] OR “treatment refusal”[Mesh] OR “patient compliance”[Mesh] OR lost to follow up[tiab] OR loss to follow up[tiab] OR default*[tiab] OR compliance[tiab] OR adherence[tiab] OR noncompliance[tiab] OR nonadherence[tiab] OR patient cooperation[tiab] OR dropout*[tiab] OR linkage to care[tiab] OR retention[tiab] OR attrition[tiab] OR cascade of care[tiab] OR treatment cascade[tiab] OR treatment success*[tiab] OR treatment completion[tiab] OR cure[tiab] OR pretreatment loss to follow-up[tiab] OR initial default[tiab]; treatment failure [tiab] |

##### Inclusion and exclusion criteria

We applied the following criteria for inclusion and exclusion of studies for this systematic review.

*Inclusion criteria* included the following:

(1) Studies that followed patients who were started on TB treatment to evaluate whether these patients achieved treatment success, or, conversely, whether they experienced suboptimal treatment outcomes, including loss to follow-up, treatment failed, not evaluated, or death. These studies had to have been conducted in programmatic settings—representing routine care in the public or private sector—and could have addressed any form of TB (i.e., TB at any body site, of any severity, or with any drug-resistance profile).
(2) Studies also had to have assessed reasons for individuals with TB in the study not achieving treatment success using a quantitative analysis. These studies could have compared characteristics of those who did or did not experience treatment success (e.g., regression analyses) or conducted structured interviews with patients who did not achieve treatment success to understand reasons for this outcome.

*Exclusion criteria* included the following:

(1) Studies that only described the proportion of patients who did or did not achieve treatment success without describing reasons why these outcomes occurred.
(2) Studies with data collected prior to the year 2000, as India’s Revised National TB Control Programme (now called the NTEP) did not achieve nationwide coverage until the early 2000s.
(3) Studies only containing qualitative data evaluating inability to achieve treatment success. Findings from studies containing qualitative data will be reported in a separate paper.
(4) Studies that enrolled patients who would not be representative of the broader patient population, or in which study personnel were involved to help retain the patient in care, beyond what would have been provided in routine care. For example, we excluded randomized trials of new drug regimens, as these trials generally involve selection of patients who are more likely to adhere to treatment and mechanisms external to routine care for retaining these patients in care.

##### Study selection

Each citation identified by the search was independently assessed by at least two reviewers (among TJ, DJ, AG, DV, MLS and KP) for their eligibility at the title and abstract evaluation stage and again subsequently at the full text evaluation stage (Fig A). Disagreements between the two reviewers were resolved by discussion or, if necessary, through consultation of a third reviewer (RS). Independent selection of articles at the title and abstract and full text stages was conducted using Covidence software (Veritas Health Innovations, Melbourne, Australia); however, quality assessment and extraction of study findings was conducted using an Excel spreadsheet.

###### Fig A. PRISMA flowchart: study selection for the systematic review of non-completion of the diagnostic workup, pretreatment loss to follow-up, and on-treatment loss to follow-up for TB patients in India (Gaps 2, 3 and 4)

**Search 2: October 2, 2015 to October 1, 2019**

**Identification**

**Included**

Records identified from search of PubMed, Embase, and Web of Science (n=860)

Duplicate records removed before screening (n=0)

Records that underwent screening of title and abstract

(n=860)

Records excluded after title and abstract screen

(n=762)

Records for which full text articles were retrieved (n=98)

Reports not retrieved

(n=0)

Reports assessed for eligibility via full text review (n=98)

Reports excluded after full text review (n = 71), for the following reasons:

Wrong outcome evaluated (n=52)

Wrong patient population (n=1)

Wrong study design (n=7)

No information on reasons for bad outcome (n=11)

Records identified from search of PubMed, Embase, and Web of Science (n=496)

Reports assessed for eligibility via full text review (n=38)

Reports excluded after full text review

(n = 20), for the following reasons:

Wrong outcomes evaluated (n=7)

Wrong patient population (n=2)

Wrong study design

(n=3)

Wrong setting (n=1)

No information on reasons for bad outcome (n=7)

Studies included in previous care cascade study with relevant data on reasons for gap 2, 3 and 4 (n=78)

Records for which full text articles were retrieved (n=38)

Records not retrieved (n=0)

Records that underwent screening of title and abstract

(n=467)

Duplicates removed before screening

(n=29)

Records excluded after title and abstract screen

(n=429)

Reports eligible for full text review (n=166), of which 88 were excluded.

Reports shortlisted after removal of duplicates (n=1,917) of which 1,751 were excluded after title and abstract review.

Reports identified from search of PubMed, Embase, and Web of Science and hand search of relevant journals (n=3,160), of which 1,243 were duplicates.

**Search 1: January 1, 2000 to October 1, 2015**

**Screening**

**Search 3: October 2, 2019 to May 17, 2021**

New studies included in the systematic review from the 2019 to 2021 search (n=18)

New studies included in the systematic review from the 2015 to 2019 search (n=27)

Total studies included in the systematic review (n=123)

Gap 2 (n=10)

Gap 3 (n=12)

Gap 4 (n=82)

Multiple gaps (n=19)

##### Quality assessment of quantitative studies

In our previous systematic review, we had developed quality criteria relevant to studies focused on identifying patients who did not complete the diagnostic workup for TB (Gap 2) or who were diagnosed with TB but did not get successfully started on, or registered in, TB treatment (Gap 3). Notably, in that prior review, we did not extract data from studies addressing patient outcomes during TB treatment (Gap 4), because district-, state-, and national-level treatment outcomes are routinely publicly reported by India’s NTEP.

Given that the goal of the current review is to identify factors (or exposures) associated with unfavorable TB treatment outcomes, we could not rely on aggregate national-level reporting of TB treatment outcomes and had to review individual studies that evaluated exposures contributing to patient outcomes for Gap 4, as with other care cascade gaps. Notably, unlike with other gaps, studies contributing to Gap 4 were more likely to use cohort or case-control designs that may be more amenable to standardized quality evaluation approaches for non-randomized observational studies, such as the Newcastle-Ottawa scale.

However, we still used a more limited quality evaluation approach for Gap 4 for a few reasons. First, the overwhelming majority of studies included in this gap were not observational studies with specific inclusion and exclusion criteria designed to evaluate specific hypotheses; rather, most Gap 4 studies, whether conducted in the public or private sector, involved evaluation of exposures and outcomes under routine programmatic care. Second, given that our systematic review is focused on identifying exposures associated with unfavorable outcomes—rather than the outcomes themselves—we felt that study quality was often related to research approaches that were more likely to enrich the diversity of exposures characterized that might contribute to outcomes. For example, many Gap 4 studies retrospectively extracted data from routine programmatic records, which considerably limited the exposures available for analysis, especially given use of standardized records in the NTEP. In contrast, studies that prospectively collected baseline data directly from patients using structured surveys were able to explore the role of broader and more diverse exposures not captured in routine programmatic records. In addition, given challenges with recording of programmatic TB outcomes in many low- and middle-income country settings, studies that prospectively verify patient outcomes through triangulation with local TB clinic staff or patients themselves may be of higher quality.

As such, we systematically evaluated and reported findings on the following criteria that may provide further insights into the characteristics and quality of the included studies. We classified the health facility-based strategy for sampling patients in each study based on whether it was comprehensive, random, or convenience. Studies using convenience sampling were excluded from analysis. Notably, this was the only criterion that was used to exclude studies from this review, because we felt that non-random sampling was a major threat to the validity of study findings. For the remaining criteria below, we report them to allow readers to better understand factors that may shape the quality of individual studies but did not exclude studies from the review based on these criteria.

Studies that assessed more than 150 patients and at over one clinical site were rated as being higher in quality than studies that assessed less than 150 patients at a single clinical site. Studies that used prospective data collection were rates as being higher in quality than studies that only relied on retrospective data extraction from program records. Furthermore, for the reasons articulated above, we also report on whether studies more specifically used prospective data collection to assess exposures and/or outcomes.

###### Table B. Criteria for assessing quality of quantitative studies evaluating failure of diagnosed TB patients not getting successfully registered in treatment.

| **Criterion** | **Quality level** |
| --- | --- |
| **Sampling strategy** | |
| Random or comprehensive sampling at selected facilities | High |
| Convenience sampling or not reported | Low (exclude findings from analysis) |
| **Sample size** | |
| >1 center and 150+ patients | High |
| Single center study with 150+ patients | Medium |
| <150 patients or not reported | Low |
| **Type of study design** | |
| Prospective data collection | High |
| Retrospective data collection | Medium to low |
| **Assessment of exposures** | |
| Patient interviews or tracking by a dedicated research team | High |
| Relying on extraction from medical records alone (limited exposures available) | Medium to low |
| **Assessment of outcomes** | |
| Patient interviews or tracking by a dedicated research team | High |
| Relying on extraction from medical records alone | Medium to low |

##### Data extraction and analysis

Multiple reviewers (TJ, AG, DV, MLS, and DJ) independently extracted data from each included study into a structured form on an Excel spreadsheet; however, we ensured that every article had data independently extracted by at least two or more reviewers. Disagreements were resolved by discussion or, if necessary, by consulting a supervising reviewer (RS). From each study, we extracted information on the study design, location, setting (i.e., urban versus rural), sample size, quality measures (e.g., prospective or retrospective data collection), and variables of interest (Table C).

For studies that compared patients who did or did not achieve treatment success or experience medication nonadherence, we extracted adjusted and unadjusted effect estimates (odds ratios, risk ratios, hazard ratios, and beta-coefficients) from regression analyses. For studies that did not report effect estimates, we calculated unadjusted odds ratios from the data provided, if possible. For studies that reported reasons why patients experienced unfavorable treatment outcomes, we extracted the proportion of patients surveyed who reported a given reason for unfavorable outcomes. For effect estimates and proportions, we extracted information on 95% confidence intervals (95% CIs) where available; if 95% CIs were not reported, we calculated these from the data provided, if possible.

For some variables, we also changed the reference group as needed for consistency of reporting across studies. For example, because most studies compared men to the reference group of women, we “flipped” effect estimates and confidence intervals for studies that presented men as the reference group. This allowed us to consistently present women as the reference group for findings regarding gender.

After “flipping” selected effect estimates from the regression analyses, we reported all unadjusted and adjusted effect estimates, regardless of statistical significance, organized by study (Table D). For the main manuscript and Forest plot, we restricted ourselves to presenting statistically significant adjusted effect estimates from multivariable analyses, as these may represent more meaningful associations from higher-quality analyses. After extracting this subset of findings, we organized findings into categories using our framework of demand- and supply-side factors (main manuscript, Table 1). For findings on reasons why patients did not achieve treatment success or experienced medication nonadherence, we presented all reported proportions in the Forest Plots in the main manuscript; we also organized these findings using our framework of demand- and supply-side categories.

To visualize quantitative findings, we generated Forest plots of effect estimates odds ratios, risk ratios, hazard ratios, beta-coefficients, and proportions using Stata version 16.1 (College Station, TX, USA). We did not conduct meta-analyses of data, because we extracted findings represented a diverse set of variables from every study.

###### Table C. Characteristics of the included studies for patients who did not achieve treatment success (Gap 4)

| **Citation** (year) | **Location**  (state/union territory) | **Urban, rural, or both** | **Public or private sector** | **Type of population** | **Type of unfavorable outcomes assessed** | **Single or multiple designated microscopy centers (DMCs)** | **Sample size** | **Methodology for assessing exposures and outcomes** | **Type of findings included in the study** (sample size for each type of analysis) |
| --- | --- | --- | --- | --- | --- | --- | --- | --- | --- |
| New TB patients |  |  |  |  |  |  |  |  |  |
| Ahmed (2009) | Karnataka | Rural | Public sector | New sputum smear positive pulmonary TB patients | Death, treatment failure, loss to follow-up, and transferred out as a composite outcome | Multiple healthcare facilities | 186 | Retrospective data collection from the government TB program* | Logistic regression (N=186)^a^ |
| Babiarz (2014) | Bihar | Rural | Public sector | New TB patients (sputum smear positive pulmonary, sputum smear negative pulmonary, and extrapulmonary) | Loss to follow-up as a single outcome (i.e., treatment discontinuation <25 weeks after initiation) | Multiple healthcare facilities | 811 | Retrospective data collection on outcomes from the government TB program with follow-up patient interview for more information on exposures* | Logistic regression (N=811) |
| Bagchi (2010) | Maharashtra | Urban | Public sector | New sputum smear positive pulmonary TB patients in the first two months of therapy | Medication non-adherence as a single outcome (i.e., at least one week's worth of missed TB medication doses in any treatment month) | Multiple healthcare facilities | 100 | Case-control study using retrospective data collection from the government TB program with cross-sectional data collection from patient interviews by a dedicated research team | Logistic Regression (N=100) |
| Balasubramanian (2004) | Tamil Nadu | Rural | Public sector | New sputum smear positive pulmonary TB patients | Loss to follow-up as a single outcome | Multiple healthcare facilities | 1,015 | Prospective data collection for both outcomes and exposures using a dedicated research team | Logistic regression (N=1,015) |
| Bhatt (2017) | Tamil Nadu | Urban and rural | Public sector | New sputum smear positive pulmonary TB patients | (1) Death, treatment failure, and loss to follow-up as single outcomes; and (2) unfavorable treatment outcomes (i.e., not achieving treatment success) as a composite outcome | Single healthcare facility* | 191 | Prospective data collection from the government TB program with longitudinal patient follow-up and interviews by a dedicated research team | Logistic regression (N=191) |
| Chakrabarti (2012) | West Bengal | Rural (tribal and non-tribal populations) | Public sector | New pulmonary TB patients (sputum smear positive pulmonary, sputum smear negative pulmonary, and extrapulmonary) | Death, treatment failure, loss to follow-up, and transferred out as a composite outcome | Single healthcare facility* | 399 | Retrospective data collection from the government TB program* | Logistic regression (N=399)^a^ |
| Gopi (2006) | Tamil Nadu | Rural | Public sector | New sputum smear positive pulmonary TB patients | Not achieving microbiological cure assessed by sputum smear microscopy | Single healthcare facility* | 1,463 | Prospective data collection from the government TB program with baseline interview of patients by a dedicated research team | Logistic regression (N=1,463) |
| Joseph (2011) | Karnataka | Rural | Public sector | New sputum smear positive pulmonary TB patients | Treatment failure and loss to follow-up as a composite outcome | Multiple healthcare facilities | 212 | Retrospective data collection from the government TB program* | Logistic regression (N=212)^a^ |
| Kulkarni (2013) | Maharashtra | Urban | Public Sector | New sputum smear positive pulmonary TB patients | Medication nonadherence (i.e., interruption of treatment for >=1 month) | Multiple healthcare facilities | 150 | Prospective data collection from the government TB program with longitudinal patient follow-up and interviews by a dedicated research team | Logistic regression (N=150); Reasons for medication nonadherence (N=78) |
| Mave (2021) | Maharashtra | Urban and rural | Public sector | New bacteriologically confirmed pulmonary TB patients (i.e., by sputum smear microscopy, Xpert MTB/RIF testing, or mycobacterial culture) | (1) Death as a single outcome, and (2) treatment failure as a single outcome | Multiple healthcare facilities | 832 | Prospective data collection for both exposures and outcomes by a dedicated research team | Logistic Regression (N=832) |
| Mukherjee (2009) | West Bengal | Urban and rural | Public sector | New pulmonary TB patients (sputum smear positive and sputum smear negative) | Death, treatment failure, and loss to follow-up as a composite outcome | Multiple healthcare facilities | 2,870 | Retrospective data collection from the government TB program* | Logistic Regression (N=2,870)^a^ |
| Mukherjee (2012) | West Bengal | Rural | Public sector | New sputum smear positive pulmonary TB patients | Death, treatment failure, and loss to follow-up as a composite outcome | Multiple healthcare facilities | 1,393 | Retrospective data collection from the government TB program* | Logistic regression (N=1,393)^a^ |
| Ramachandran (2020) | Tamil Nadu and Maharashtra | Urban | Public sector | New pulmonary TB patients (sputum smear positive and sputum smear negative) | Treatment failure and death as single outcomes | Multiple healthcare facilities | 404 | Prospective data collection from the government TB program with baseline evaluation and then longitudinal follow-up of patients by a dedicated research team | Poisson regression (N=404) |
| Shameer (2016) | Kerala | Not reported | Public sector | New TB patients (sputum smear positive pulmonary, sputum smear negative pulmonary, and extrapulmonary) | Medication nonadherence (i.e., missing 3 or more consecutive doses) | Multiple healthcare facilities | 141* | Case-control study using retrospective data collection from the government TB program with cross-sectional data collection from patient interviews by a dedicated research team | Logistic regression (N=141) |
| Shewade (2019) | Tamil Nadu, Kerala, Maharashtra, Madhya Pradesh, Chhattisgarh, Bihar, Jharkhand, West Bengal, Punjab | Urban and rural | Public sector | New sputum smear positive pulmonary TB patients | Death, treatment failure, loss to follow-up, and not evaluated as a composite outcome | Multiple healthcare facilities | 572 | Prospective data collection from the government TB program with baseline patient interviews by a dedicated research team | Logistic regression (N=572) |
| Shivam (2014) | West Bengal | Rural | Public sector | New TB patients (sputum smear positive pulmonary, sputum smear negative pulmonary, and extrapulmonary) | Death, treatment failure, and loss to follow-up as a composite outcome | Multiple healthcare facilities | 758 | Retrospective data collection from the government TB program* | Logistic regression (N=758)^a^ |
| Singla (2009) | Delhi | Urban | Public sector | New sputum smear positive pulmonary TB patients | Treatment failure as a single outcome as compared to patients who achieved cure | Single healthcare facility* | 118* | Prospective data collection from the government TB program with baseline patient interviews by a dedicated research team | Logistic regression (N=118) |
| Singla (2013) | Delhi | Urban | Public sector | New sputum smear positive pulmonary TB patients | (1) Sputum smear positive at 2 months as a single outcome; (2) sputum culture positive at 2 months as a single outcome; and (3) death, treatment failure, and loss to follow-up as a composite outcome | Single healthcare facility* | 148* | Prospective data collection from the government TB program with baseline patient interviews by a dedicated research team | Logistic regression (N=148) |
| Tiwari (2012) | Delhi | Urban | Public sector | New sputum smear positive pulmonary TB patients | Death, treatment failure, loss to follow-up, and transferred out as a composite outcome | Multiple healthcare facilities | 338 | Prospective data collection from the government TB program with baseline patient interviews by a dedicated research team | Logistic regression (N=338)^a^ |
| Trivedi (2019) | Gujarat | Urban and rural | Public sector | New sputum smear positive pulmonary TB patients | Not achieving cure (based on follow-up sputum microscopy) as a single outcome | Single healthcare facility* | 76* | Prospective data collection from the government TB program with baseline patient interviews by a dedicated research team | Logistic regression (N=76)^a^ |
| Vashishtha (2013) | Delhi | Urban | Public sector | New TB patients (sputum smear positive pulmonary, sputum smear negative pulmonary, and extrapulmonary) with and without HIV | Death, treatment failure, loss to follow-up, and treatment modified as a composite outcome | Single healthcare facility* | 305 (150 with HIV and 155 without HIV) | Prospective data collection from the government TB program by a dedicated research team | Logistic regression (N=305)^a^ |
| Velayutham (2014) | Tamil Nadu | Urban and rural | Public sector | New TB patients (sputum smear positive pulmonary, sputum smear negative pulmonary, and extrapulmonary) | (1) Death as a single outcome; (2) treatment failure as a single outcome; and (3) loss to follow-up as a single outcome. | Multiple healthcare facilities | 2,602 | Retrospective data collection from the government TB program* | Logistic regression (N=2,602) |
| Velayutham (2014) | Tamil Nadu | Urban and rural | Public sector | New sputum smear positive pulmonary TB patients | (1) Death as a single outcome; (2) treatment failure as a single outcome; and (3) loss to follow-up as a single outcome. | Multiple healthcare facilities | 2,285 | Retrospective data collection from the government TB program* | Logistic regression (N=2,285) |
| Velayutham (2018) | Madhya Pradesh, Kerala, Delhi, Tamil Nadu, Maharashtra, Karnataka | Urban and rural | Public sector | New sputum smear positive pulmonary TB patients | Death, treatment failure, loss to follow-up, treatment modified, and not evaluated as a composite outcome | Multiple healthcare facilities | 1,543 | Prospective data collection from the government TB program with baseline and follow-up patient interviews by a dedicated research team | Logistic Regression (N=1,543) |
| Vijay (2010) | Tamil Nadu, Kerala, Rajashan, Himachal Pradesh, Delhi, Manipur | Urban and rural | Public sector | New sputum smear positive pulmonary TB patients | Loss to follow-up as a single outcome | Multiple healthcare facilities | 929 | Case-control study using retrospective data collection from the government TB program with cross-sectional data collection from patient interviews by a dedicated research team | Logistic Regression (N=929) |
| Viswanathan (2014) | Tamil Nadu | Urban and rural | Public sector | New sputum smear positive pulmonary TB patients | (1) Death, treatment failure, treatment modified, and loss to follow-up as a composite outcome; and (2) treatment failure and treatment modified as a composite outcome | Multiple healthcare facilities | 245 | Prospective data collection from the government TB program with baseline patient interviews by a dedicated research team | Logistic regression (N=245)^a^ including diabetics and non-diabetes; Logistic regression (N=78)^a^ including diabetics only |
| Zaman (2014) | Assam | Not reported | Public sector | New sputum smear positive pulmonary TB patients | Death, treatment failure, and loss to follow-up as a composite outcome | Multiple healthcare facilities | 54* | Prospective data collection from the government TB program with baseline and follow-up patient interviews by a dedicated research team | Logistic regression (N=54)^a^ for death, treatment failure, or loss to follow-up as a composite outcome; Logistic regression (N=28)^a^ for loss to follow-up as a single outcome among patients with irregular adherence; Reasons for medication nonadherence including loss to follow-up (N=28) |
| Zhou (2020) | Puducherry, Tamil Nadu | Urban and rural | Public sector | New sputum smear positive pulmonary TB patients (confirmed by positive mycobacterial culture) | Loss to follow-up as a single outcome | Multiple healthcare facilities | 425 | Prospective data collection from the government TB program with baseline interviews by a dedicated research team | Logistic Regression (N=425) |
| Patients with a prior TB treatment history |  |  |  |  |  |  |  |  |  |
| Babiarz (2014) | Bihar | Rural | Public sector | Previously treated TB patients (sputum smear positive pulmonary, sputum smear negative pulmonary, and extrapulmonary) | Loss to follow-up as a single outcome (i.e., treatment discontinuation <25 weeks after initiation) | Multiple healthcare facilities | 196 | Retrospective data collection on outcomes from the government TB program with follow-up patient interview for more information on exposures* | Logistic Regression (N=196) |
| Bhagat (2010) | Maharashtra | Urban | Public sector | Previously treated TB patients (sputum smear positive pulmonary, sputum smear negative pulmonary, and extrapulmonary) | Loss to follow-up as a single outcome | Multiple healthcare facilities | 112* | Retrospective data collection from the government TB program* | Logistic Regression (N=97) |
| Chakrabarti (2012) | West Bengal | Rural (tribal and non-tribal populations) | Public sector | Previously treated TB patients (sputum smear positive pulmonary, sputum smear negative pulmonary, and extrapulmonary) | Death, treatment failure, loss to follow-up, and transferred out as a composite outcome | Single healthcare facility* | 75 | Retrospective data collection from the government TB program* | Logistic Regression (N=75)^a^ |
| Chandrasekaram (2006) | Tamil Nadu | Rural | Public sector | Previously treated smear positive pulmonary TB patients | Death, treatment failure, loss to follow-up, and others (e.g., transferred out) as a composite outcome | Multiple healthcare facilities | 696 | Retrospective data collection from the government TB program* | Logistic regression (N=696)^a^ |
| Deepa (2013) | Andhra Pradesh | Urban | Public sector | Previously treated sputum smear positive pulmonary TB patients | Death, treatment failure, loss to follow-up, and transferred out as a composite outcome | Multiple healthcare facilities | 1,077 | Retrospective data collection from the government TB program* | Relative risk regression (N=1,077) |
| Joseph (2011) | Karnataka | Rural | Public sector | Previously sputum smear positive pulmonary TB patients | Treatment failure and loss to follow-up as a composite outcome | Multiple healthcare facilities | 74* | Retrospective data collection from the government TB program* | Logistic Regression (N=74)^a^ |
| Jha (2010) | Nationally representative sample | Urban and rural | Public sector | Previously treated TB patients (sputum smear positive pulmonary, sputum smear negative pulmonary, and extrapulmonary) | Loss to follow-up as a single outcome compared to patients with treatment success or treatment failure | Multiple healthcare facilities | 8,790 | Case-control study using retrospective data from the government TB program* | Logistic Regression (N=2,330 for univariable analysis, N=1,160 for multivariable analysis) |
| Mukherjee (2009) | West Bengal | Rural | Public sector | Previously sputum smear positive pulmonary TB patients | Death, treatment failure, loss to follow-up, and transferred out as a composite outcome | Multiple healthcare facilities | 234 | Retrospective data collection from the government TB program* | Logistic Regression (N=234)^a^ |
| Nagaraja (2011) | Andhra Pradesh | Not reported | Public sector | Previously treated patients who experienced treatment failure and were not placed on regimens to treat drug-resistant TB | Death, treatment failure, loss to follow-up, and transferred out as a composite outcome | Multiple healthcare facilities | 202 | Retrospective data collection from the government TB program* | Logistic Regression (N=202)^a^ |
| Pardeshi (2010) | Maharashtra | Urban | Public sector | Previously treated sputum smear positive pulmonary TB patients | Loss to follow-up as a single outcome | Multiple healthcare facilities | 716 | Retrospective data collection from the government TB program* | Logistic Regression (N=95)^a^ |
| Sarpal (2014) | Chandigarh | Urban | Public sector | Previously treated TB patients (sputum smear positive pulmonary, sputum smear negative pulmonary, and extrapulmonary) | Death, treatment failure, and loss to follow-up as a composite outcome | Multiple healthcare facilities | 545 | Prospective data collection from the government TB program with longitudinal patient follow-up and interviews by a dedicated research team | Logistic Regression (N=545)^a^ |
| Sarpal (2014) | Chandigarh | Urban | Public sector | Previously treated TB patients (sputum smear positive pulmonary, sputum smear negative pulmonary, and extrapulmonary) | Loss to follow-up as a single outcome | Multiple healthcare facilities | 545 | Prospective data collection from the government TB program with longitudinal patient follow-up and interviews by a dedicated research team | Logistic Regression (N=545);^a^ Reasons for loss to follow-up (N=32) |
| Shivam (2014) | West Bengal | Rural | Public sector | Previously treated TB patients (sputum smear positive pulmonary, sputum smear negative pulmonary, and extrapulmonary) | Death, treatment failure, and loss to follow-up as a composite outcome | Multiple healthcare facilities | 149 | Retrospective data collection from the government TB program* | Logistic Regression (N=149)^a^ |
| Singla (2009) | New Delhi | Urban | Public sector | Previously treated sputum smear positive pulmonary TB patients | Death, treatment failure, loss to follow-up, and transferred out as a composite outcome | Single healthcare facility* | 38* | Prospective data collection from the government TB program with baseline patient interviews by a dedicated research team | Logistic Regression (N=38)^a^ for all previously treated patients; logistic regression (N=19)^a^ for previously treated patients with any drug resistance detected |
| Sisodia (2006) | Rajasthan | Not reported |  | Previously treated sputum smear positive pulmonary TB patients | Death, treatment failure, loss to follow-up, and transferred out as a composite outcome | Multiple healthcare facilities | 2,215 | Prospective data collection from the government TB program by a dedicated research team | Logistic Regression (N=2,215)^a^ |
| Srinath (2010) | Andhra Pradesh | Not reported | Public sector | Previously treated TB patients, specifically focusing on "other" TB previously treated patients (generally sputum smear negative pulmonary or extrapulmonary patients) | Death, treatment failure, loss to follow-up, and transferred out as a composite outcome | Multiple healthcare facilities | 4,067 previously treated TB patients, including 1,029 "other" previously treated patients | Retrospective data collection from the government TB program* | Logistic Regression (N=4,067)^a^ for all previously treated patients; logistic regression (N=1,009) for "other" previously treated patients |
| Velavan (2018) | Puducherry | Urban and rural | Public sector | Previously treated TB patients (sputum smear positive pulmonary, sputum smear negative pulmonary, and extrapulmonary) | Death, treatment failure, and loss to follow-up as a composite outcome | Multiple healthcare facilities | 392 | Retrospective data collection from the government TB program* | Logistic Regression (N=392)^a^ |
| Velayutham (2014) | Tamil Nadu | Urban and rural | Public sector | (1) All previously treated TB patients; and (2) previously treated sputum smear positive pulmonary TB patients | (1) Death as a single outcome; (2) treatment failure as a single outcome; and (3) loss to follow-up as a single outcome | Multiple healthcare facilities | 803 previously treated TB patients, including 699 previously treated smear positive pulmonary TB patients | Retrospective data collection from the government TB program* | Logistic Regression (N=803) previously treated patients; logistic regression (N=699) previously treated smear positive pulmonary TB patients |
| Rifampin-resistant or multidrug-resistant (MDR) TB patients |  |  |  |  |  |  |  |  |  |
| Bhatt (2018) | New Delhi | Urban | Public sector | Rifampin-resistant TB patients confirmed by Xpert MTB/RIF or culture | (1) Death as a single outcome; (2) treatment failure as a single outcome; and (3) loss to follow-up as a single outcome | Single healthcare facility* | 123* | Retrospective data collection from the government TB program* | Logistic Regression (N=123); Cox regression (N=123) |
| Dela (2017) | Gujarat | Not reported | Public sector | Multidrug-resistant TB patients registered in the government program | Death, progression to extensively drug-resistant TB, loss to follow-up, and transfer out as a composite outcome | Single healthcare facility* | 125* | Retrospective data collection from the government TB program* | Logistic Regression (N=125)^a^ |
| Dole (2017) | Maharashtra | Urban | Public sector | Rifampin-resistant TB patients confirmed by line probe assay or Xpert MTB/RIF | Death, treatment failure, and loss to follow-up as a composite outcome | Single healthcare facility* | 146* | Retrospective data collection from the government TB program* | Logistic regression (N=146); Reasons for loss to follow-up (N=28) |
| Duraisamy (2014) | Kerala | Urban and rural | Public sector | Multidrug-resistant TB patients confirmed by culture | (1) Death, treatment failure, progression to extensively drug-resistant TB, loss to follow-up, treatment interruption due to adverse drug reaction, and transferred out as a composite outcome; and (2) medication non-adherence (mean number of missed doses) as a single outcome | Multiple healthcare facilities | 179 | Retrospective data collection from the government TB program* | Cox regression (N=179) |
| Isaakidis (2012) | Maharashtra | Urban | Private sector  (non-profit non-governmental organization) | Multidrug-resistant TB patients, confirmed by culture or diagnosed empirically, with HIV | Death, treatment failure, and loss to follow-up as a composite outcome | Single healthcare facility* | 67* | Prospective data collection by clinical staff and a dedicated research team with data entered into an electronic medical record and separate clinical research database | Logistic Regression (N=67) |
| Jain (2014) | Gujarat | Urban | Public sector | Multidrug-resistant TB patients registered in the government program and treated with a standardized regimen | Death, treatment failure, and loss to follow-up as a composite outcome | Single healthcare facility* | 130* | Prospective data collection from the government TB program with baseline patient interview by dedicated research staff | Logistic Regression (N=130)^a^ |
| Janmeja (2017) | Chandigarh | Urban | Public sector | Rifampin resistant or multidrug-resistant TB diagnosed by cartridge-based nucleic acid amplification testing or culture, respectively | Death, treatment failure, modification of therapy, and loss to follow-up as a composite outcome | Single healthcare facility* | 256 | Retrospective data collection from the government TB program* | Logistic Regression (N=256) |
| Kandi (2021) | Telangana | Urban | Public sector | Multidrug-resistant TB patients registered in the government program | Death, treatment failure, switch to extensively drug-resistant TB treatment, and loss to follow-up as a composite outcome | Single healthcare facility* | 377 | Retrospective data collection from the government TB program* | Logistic Regression (N=377)^a^ |
| Lohiya (2020) | New Delhi | Urban | Public sector | Extrapulmonary rifampin-resistant or multidrug-resistant TB diagnosed by cartridge-based nucleic acid amplification testing, culture, or empiric diagnosis | Death, treatment failure, modification of therapy, treatment discontinuation for reasons other than adverse drug reactions, and loss to follow-up as a composite outcome | Multiple healthcare facilities | 203 | Retrospective data collection from the government TB program* | Relative Risk Regression (N=203) |
| Nair (2016) | Tamil Nadu | Urban and rural | Public sector | Multidrug-resistant or rifampin-resistant TB diagnosed by culture vs. line probe assay or Xpert MTB/RIF | Death, treatment failure, switched to extensively drug resistant TB treatment, interrupted treatment due to reasons other than adverse drug reaction, transfer out, and loss to follow-up as a composite outcome | Multiple healthcare facilities | 524 | Retrospective data collection from the government TB program* | Relative risk regression (N=524) |
| Natarajan (2020) | Delhi | Urban and rural | Public sector | Multidrug-resistant or rifampin-resistant pulmonary TB who required the addition of bedaquiline with/without other newer/repurposed drugs | Medication non-adherence as a single outcome | Single healthcare facility* | 275 | Prospective data collection by clinical staff and a dedicated research team with data entered into an electronic medical record and separate clinical research database | Reasons for treatment interruption (N=275) |
| Parmar (2018) | 7 Indian states | Urban and rural | Public sector | Multidrug-resistant TB patients diagnosed by culture or line probe assay | (1) Death, treatment failure, and loss to follow-up as a composite outcome; (2) death as a single outcome; (3) treatment failure as a single outcome; and (4) loss to follow-up as a single outcome. | Multiple healthcare facilities | 2,264 | Retrospective data collection from the government TB program* | Logistic Regression (N=2,264) |
| Patel (2018) | Gujarat | Urban | Public sector | Multidrug-resistant TB patients registered in the government program | Death, treatment failure (i.e., smear positivity, culture positivity, or smear/culture unavailability), and loss to follow-up as a composite outcome | Single healthcare facility* | 145 | Prospective data collection from the government TB program with follow-up interviews by dedicated research staff | Reasons for loss to follow-up (N=32) |
| Rupani (2020) | Gujarat | Urban | Public sector | Multidrug-resistant TB patients diagnosed by cartridge-based nucleic acid amplification testing | Loss to follow-up (i.e., discontinuation of MDR TB treatment) | Single healthcare facility* | 94* | Prospective data collection from the government TB program with follow-up interviews by dedicated research staff | Logistic Regression (N=94) |
| Saha (2017) | Maharashtra | Urban | Private sector  (for-profit hospital) | Multidrug-resistant TB, pre-extensively drug-resistant TB, or extensively drug-resistant TB diagnosed by Xpert MTB/RIF and culture | Death, treatment failure, and loss to follow-up as a composite outcome | Single healthcare facility* | 59* | Retrospective data collection from medical record of private hospital* | Logistic regression (N=59)^a^ |
| Sharma (2020) | Delhi | Urban | Public sector | Multidrug-resistant TB patients registered in the government program | Death, treatment failure, switched to extensively drug-resistant TB therapy, treatment stopped due to adverse drug reaction, and loss to follow-up as a composite outcome | Multiple healthcare facilities | 2,958 | Retrospective data collection from the government TB program* | Logistic regression (N=2,958) |
| Shringarpure (2015) | Gujarat | Urban and rural | Public sector | Multidrug-resistant TB patients registered in the government program | Loss to follow-up as a single outcome | Single healthcare facility* | 322 | Retrospective data collection from the government TB program* | Cox regression (N=322) |
| Multiple populations |  |  |  |  |  |  |  |  |  |
| Babiarz (2014) | Bihar | Rural | Public sector | New and previously treated TB patients (sputum smear positive pulmonary, sputum smear negative pulmonary, and extrapulmonary) as a combined population | Loss to follow-up as a single outcome (i.e., treatment discontinuation <25 weeks after initiation) | Multiple healthcare facilities | 1,007 | Retrospective data collection on outcomes from the government TB program with follow-up patient interview for more information on exposures* | Logistic regression (N=1,007) |
| Bagchi (2010) | Maharashtra | Urban | Public sector | New and previously treated sputum smear positive pulmonary TB patients, excluding new patients in the first two months of therapy | Medication non-adherence as a single outcome (i.e., at least one week's worth of missed TB medication doses in any treatment month) | Multiple healthcare facilities | 438 | Case-control study using retrospective data collection from the government TB program with cross-sectional data collection from patient interviews by a dedicated research team | Logistic regression (N=438) |
| Balasubramanian (2004) | Tamil Nadu | Rural | Public sector | New and previously treated TB patients (sputum smear positive pulmonary, sputum smear negative pulmonary, and extrapulmonary) as a combined population | Loss to follow-up as a single outcome | Multiple healthcare facilities | 2,371 | Prospective data collection for both exposures and outcomes by a dedicated research team | Logistic regression (N=2,371);  Reasons for loss to follow-up (N=1,086) |
| Banerjee (2020) | West Bengal | Urban | Public sector | New and previously treated TB patients (pulmonary and extrapulmonary) as a combined population | Death, treatment failure, and loss to follow-up as a composite outcome | Multiple healthcare facilities | 140* | Prospective data collection for both exposures and outcomes by a dedicated research team | Logistic regression (N=140) |
| Bhagyalaxmi (2010) | Gujarat | Urban | Public sector | New and previously treated TB patients (pulmonary and extrapulmonary) as a combined population | Death, treatment failure, and loss to follow-up as a composite outcome | Multiple healthcare facilities | 200 | Prospective data collection for both exposures and outcomes by a dedicated research team | Logistic regression (N=200)^a^ |
| Brahmapurkar (2017) | Chhattisgarh | Urban | Public sector | New and previously treated TB patients (sputum smear positive pulmonary, sputum smear negative pulmonary, and extrapulmonary) as a combined population | Death, treatment failure, loss to follow-up, and transferred out as a composite outcome | Multiple healthcare facilities | 496 | Retrospective data collection from the government TB program* | Logistic regression (N=496)^a^ |
| Cox (2021) | Maharashtra and Tamil Nadu | Urban and rural | Public sector | Men with drug-susceptible or presumed drug-susceptible pulmonary TB confirmed by Xpert MTB/RIF or culture or through empirical diagnosis for those with test-negative TB | Treatment failure as a single outcome | Multiple healthcare facilities | 751 | Prospective data collection for both exposures and outcomes by a dedicated research team | Logistic regression (N=751) |
| Dandona (2004) | Andhra Pradesh, Maharastra, Rajasthan, Tamil Nadu | Urban and rural | Public sector | New and previously treated pulmonary TB patients as a combined population | Loss to follow-up as a single outcome | Multiple healthcare facilities | 744 | Prospective data collection for both exposures and outcomes by a dedicated research team | Logistic regression (N=744); Reported barriers to treatment completion among those who did and did not complete treatment (N=729) |
| Das (2014) | Nagaland | Rural | Public sector with private sector support from a non-profit philanthropic organization | New and previously treated TB patients (sputum smear positive pulmonary, sputum smear negative pulmonary, and extrapulmonary) as a combined population | Death, treatment failure, and loss to follow-up as a composite outcome | Single healthcare facility* | 238 | Retrospective data collection from the government TB program* | Logistic regression (N=238)^a^ |
| Gopi (2007) | Tamil Nadu | Rural | Public sector | New and previously treated pulmonary TB patients (sputum smear positive and sputum smear negative) during the intensive phase of therapy as a combined population | Medication non-adherence as a single outcome (i.e., partial or complete non-observation of doses by healthcare providers during the intensive phase of therapy) | Multiple healthcare facilities | 1,666 | Prospective data collection for both exposures and outcomes by a dedicated research team | Logistic regression (N=1,666) |
| Gupta (2011) | Delhi | Urban | Public sector | New and previously treated TB patients (including smear-positive pulmonary, smear-negative pulmonary, and extrapulmonary disease) with history of treatment interruption as a combined population | Medication non-adherence as a single outcome | Single healthcare facility* | 201 | Retrospective cohort-based analysis | Reasons for treatment interruption (N=201) |
| Huddart (2021) | Bihar | Both | Private sector | New and previously treated TB patients as a combined population | Death as a single outcome | Multiple healthcare facilities | 4,000 | Retrospective assessment of outcomes using phone surveys by a dedicated research team* | Logistic regression (N=4,000, with n=2,240 observed [i.e., answered the survey] and n=1,760 unobserved [i.e., did not answer the survey but accounted for using inverse probability weighting]) |
| Jaggarajamma (2007) | Tamil Nadu | Rural | Public sector | New and previously treated TB patients (sputum smear positive pulmonary, sputum smear negative pulmonary, and extrapulmonary) as a combined population | Loss to follow-up as a single outcome | Multiple healthcare facilities | 1,124 | Prospective data collection for both exposures and outcomes by a dedicated research team | Logistic regression (N=1,124);^a^  Reasons reported by patients for loss to follow-up (N=141) |
| Jan Swasthya Sahyog (2018) | Chattisgharh | Rural | Private sector (non-profit community-based organization) | New and previously treated TB patients (sputum smear positive pulmonary, sputum smear negative pulmonary, and extrapulmonary) as a combined population | Death, treatment failure, and loss to follow-up as a composite outcome | Multiple healthcare facilities | 4,979 | Retrospective data collection from health system records of a community-based program* | Relative risk regression and logistic regression (N=2,607)^a^ |
| Jonnalagada (2011) | Andra Pradesh | Urban and rural | Public sector | New and previously treated TB patients (sputum smear positive pulmonary, sputum smear negative pulmonary, and extrapulmonary) as a combined population | Death, treatment failure, loss to follow-up, and transferred out as a composite outcome | Multiple healthcare facilities | 8,240 | Retrospective data collection from the government TB program* | Logistic regression (N=8,240)^a^ |
| Joseph (2011) | Karnataka | Rural | Public sector | New and previously treated sputum smear positive pulmonary TB patients as a combined population | Treatment failure and loss to follow-up as a composite outcome | Single healthcare facility* | 286 | Retrospective data collection from the government TB program* | Logistic regression (N=286)^a^ |
| Karanjekar (2014) | Maharashtra | Rural | Public sector | New and previously treated TB patients (sputum smear positive pulmonary, sputum smear negative pulmonary, and extrapulmonary) as a combined population | Death, loss to follow-up, and transferred out as a composite outcome | Single healthcare facility* | 125* | Prospective data collection for both exposures and outcomes by a dedicated research team | Logistic regression (N=125)^a^ |
| Kumar (2018) | Madhya Pradesh | Urban | Public sector | New and previously treated TB patients (sputum smear positive pulmonary, sputum smear negative pulmonary, and extrapulmonary) as a combined population | Death, treatment failure, loss to follow-up, and transferred out as a composite outcome | Multiple healthcare facilities | 454 | Retrospective data collection from the government TB program* | Logistic regression (N=454) |
| Lata (2021) | Jammu and Kashmir | Rural | Public sector | New and previously treated TB patients who completed at least 2 months of TB therapy as a combined population | Medication non-adherence as a single outcome (i.e., self-reported non-ingestion of at least one medication dose as measured by the Morisky adherence scale) | Single healthcare facility* | 72* | Prospective data collection for both exposures and outcomes by a dedicated research team | Logistic regression (N=72);^a^  Reasons for medication nonadherence (N=14) |
| Mittal (2011) | Uttar Pradesh | Urban | Public sector | New and previously treated TB patients (sputum smear positive pulmonary, sputum smear negative pulmonary, and extrapulmonary) as a combined population | Death, treatment failure, loss to follow-up, and transferred out as a composite outcome | Multiple healthcare facilities | 900 | Retrospective data collection from the government TB program* | Logistic regression (N=900)^a^ |
| Mittal (2011) | Uttar Pradesh | Urban | Public sector | New and previously treated TB patients (sputum smear positive pulmonary, sputum smear negative pulmonary, and extrapulmonary) as a combined population | Loss to follow-up as a single outcome | Multiple healthcare facilities | 900 | Retrospective data collection on outcomes from the government TB program;* prospective interview of patients who became lost to follow-up | Logistic regression (N=900);^a^  Reasons for loss to follow-up (N=111) |
| Mukhopadhyay (2011) | West Bengal | Urban and rural | Public sector | New and previously treated TB patients (sputum smear positive pulmonary, sputum smear negative pulmonary, and extrapulmonary) as a combined population | Death, treatment failure, and loss to follow-up as a composite outcome | Multiple healthcare facilities | 898 | Retrospective data collection from the government TB program* | Logistic regression (N=898)^a^ |
| Mundra (2017) | Maharashtra | Urban and rural | Public sector | New and previously treated TB patients (sputum smear positive pulmonary, sputum smear negative pulmonary, and extrapulmonary) as a combined population | (1) Death, treatment failure, treatment modification, and loss to follow-up as a composite outcome; (2) death as a single outcome; (3) treatment failure as a single outcome; (4) treatment modification as a single outcome; and (5) loss to follow-up as a single outcome | Multiple healthcare facilities | 503 | Retrospective data collection from the government TB program* | Cox regression (N=503) |
| Mundra (2018) | Maharashtra | Urban and rural | Public sector | New and previously treated TB patients (sputum smear positive pulmonary, sputum smear negative pulmonary, and extrapulmonary) as a combined population | Death, treatment failure, treatment modification, and loss to follow-up as a composite outcome | Single healthcare facility* | 275 | Case-control study using retrospective data collection from the government TB program with cross-sectional data collection from patient interviews by a dedicated research team | Logistic regression (N=275) |
| Nahar (2014) | Madhya Pradesh | Urban | Public sector | TB patients being treated in the government TB program (no further description, but presumed to comprise new and previously treated patients) | Loss to follow-up as a single outcome | Multiple healthcare facilities | Not fully clarified, presumed to be 386 | Case-control study using retrospective data collection from the government TB program with cross-sectional data collection from patient interviews by a dedicated research team | Logistic regression (Sample size not fully clarified, presumed to be N=386) |
| Nandakumar (2013) | Kerala | Urban and rural | Public sector | New and previously treated TB patients (sputum smear positive pulmonary, sputum smear negative pulmonary, and extrapulmonary) as a combined population | Death, treatment failure, loss to follow-up, and transferred out as a composite outcome | Multiple healthcare facilities | 3,116 | Retrospective data collection from the government TB program* | Relative risk regression (N=3,116) |
| Pardeshi (2007) | Maharashtra | Urban | Public sector | New and previously treated sputum smear positive pulmonary TB patients as a combined population | Death, treatment failure, and loss to follow-up as a composite outcome | Single healthcare facility* | 1,646 | Retrospective data collection from the government TB program* | Logistic regression (N=1,646)^a^ |
| Pardeshi (2010) | Maharashtra | Urban | Public sector | New and previously treated sputum smear positive pulmonary TB patients as a combined population | Death, treatment failure, loss to follow-up, and transferred out as a composite outcome | Single healthcare facility* | 1,925 | Retrospective data collection from the government TB program* | Logistic regression (N=1,925)^a^ |
| Patra (2013) | Delhi | Urban | Public sector | New and previously treated TB patients (sputum smear positive pulmonary, sputum smear negative pulmonary, and extrapulmonary) as a combined population among individuals >=60 years old | Death, treatment failure, loss to follow-up, and transferred out as a composite outcome | Single healthcare facility* | 2,401 | Retrospective data collection from the government TB program* | Logistic regression (N=2,401) |
| Pauniikar (2019) | Maharashtra | Both | Public sector | New and previously treated TB patients (sputum smear positive pulmonary, sputum smear negative pulmonary, and extrapulmonary) as a combined population | Loss to follow-up as a single outcome | Single healthcare facility* | 440 | Prospective data collection for both exposures collected by a dedicated research team with retrospective data on outcomes collected from the government TB program | Cox regression (N=440) |
| Pore (2020) | Maharashtra | Rural | Public sector | Presumed drug-susceptible new and previously treated TB patients and multidrug-resistant TB patients (sputum smear positive pulmonary, sputum smear negative pulmonary, and extrapulmonary) as a combined population | Medication non-adherence as a single outcome (i.e., missing one or more medications for 7 consecutive days anytime during the treatment period) | Single healthcare facility* | 88* | Prospective data collection for both exposures and outcomes by a dedicated research team | Logistic regression (N=88);^a^ Reasons for medication nonadherence (N=34) |
| Prudhivi (2019) | Andhra Pradesh | Urban and rural | Public sector | New and previously treated pulmonary TB patients (sputum smear positive and sputum smear negative) as a combined population | Death, treatment failure, loss to follow-up, and transferred out as a composite outcome | Single healthcare facility* | 1,113 | Retrospective data collection from the government TB program* | Logistic regression (N=1,113) |
| Ratnesh (2020) | Uttar Pradesh | Urban | Public sector | New and previously treated TB patients (sputum smear positive pulmonary, sputum smear negative pulmonary, and extrapulmonary) as a combined population | Loss to follow-up as a single outcome | Multiple healthcare facilities | 2,010 | Prospective data collection for both exposures and outcomes by a dedicated research team | Logistic regression (N=2,010) |
| Shabil (2019) | Karnataka | Urban and rural | Public sector | New and previously treated TB patients as a combined population, without further description of the population | Loss to follow-up as a single outcome | Single healthcare facility* | 90* | Prospective data collection for both exposures and outcomes by a dedicated research team | Logistic regression (N=90);^a^ Reasons for loss to follow-up (N=12); Reasons for medication nonadherence (i.e., treatment interruption) (N=9) |
| Sharma (2003) | Delhi | Urban | Public sector | New and previously treated TB patients (sputum smear positive pulmonary, sputum smear negative pulmonary, and extrapulmonary) as a combined population | Death, treatment failure, loss to follow-up, and transferred out as a composite outcome | Single healthcare facility* | 67 | Retrospective data collection from the government TB program* | Logistic regression (N=67)^a^ |
| Sharma (2021) | West Bengal | Urban | Private sector | New and previously treated TB patients (sputum smear positive pulmonary, sputum smear negative pulmonary, and extrapulmonary) as a combined population | Death, treatment failure, treatment modified to drug-resistant TB therapy, loss to follow-up, and not evaluated as a composite outcome | Multiple healthcare facilities | 2,347 | Retrospective data collection from an electronic database created to capture outcomes on patients treated in the private sector* | Logistic regression (N=2,347)^a^ |
| Shivam (2014) | West Bengal | Rural | Public sector | New and previously treated TB patients (sputum smear positive pulmonary, sputum smear negative pulmonary, and extrapulmonary) as a combined population | Death, treatment failure, and loss to follow-up as a composite outcome | Single healthcare facility* | 758 | Retrospective data collection from the government TB program* | Logistic regression (N=758) |
| Siddiqui (2016) | Delhi | Urban | Public sector | New and previously treated TB patients (sputum smear positive pulmonary, sputum smear negative pulmonary, and extrapulmonary) as a combined population | Death, treatment failure, treatment modified to drug-resistant TB therapy, and loss to follow-up as a composite outcome | Multiple healthcare facilities | 316 | Retrospective data collection from the government TB program* | Logistic regression (316) |
| Singh (2020) | Uttarakhand | Urban | Public sector | New and previously treated TB patients (sputum smear positive pulmonary, sputum smear negative pulmonary, and extrapulmonary) as a combined population | Death, treatment failure, treatment regimen modified, loss to follow-up, transferred out and not evaluated as a composite outcome | Multiple healthcare facilities | 433 | Retrospective data collection from the government TB program* | Logistic regression (N=238) |
| Vasantha (2008) | Tamil Nadu | Rural | Public sector | New and previously treated TB patients (sputum smear positive pulmonary, sputum smear negative pulmonary, and extrapulmonary) as a combined population | Death as a single outcome | Multiple healthcare facilities | 3,513 | Prospective data collection for both exposures and outcomes by a dedicated research team | Cox regression (N=3,513) |
| Vashishtha (2013) | Delhi | Urban | Public sector | New and previously treated TB patients (sputum smear positive pulmonary, sputum smear negative pulmonary, and extrapulmonary) as a combined population among people with and without HIV | Death, treatment failure, treatment regimen modified, and loss to follow-up as a composite outcome | Multiple healthcare facilities | 305 | Prospective data collection for both exposures and outcomes by a dedicated research team | Logistic regression (N=305)^a^ |
| Vasudevan (2014) | Puducherry | Urban | Public sector | New and previously treated TB patients (sputum smear positive pulmonary, sputum smear negative pulmonary, and extrapulmonary) as a combined population | Loss to follow-up as a single outcome | Multiple healthcare facilities | 4,421 | Retrospective data collection from the government TB program* | Logistic regression (N=4,421)^a^ |
| Viswanathan (2014) | Tamil Nadu | Urban | Public sector | New and previously treated TB patients (sputum smear positive pulmonary, sputum smear negative pulmonary, and extrapulmonary) as a combined population | Death, treatment failure, and loss to follow-up as a composite outcome | Multiple healthcare facilities | 209 | Retrospective data collection from the government TB program* | Logistic regression (N=209)^a^ |
| Washington (2020) | Karnataka and Telangana | Rural | Public sector with private sector support from a non-profit philanthropic organization | Presumed drug-susceptible new and previously treated TB patients and multidrug-resistant TB patients (sputum smear positive pulmonary, sputum smear negative pulmonary, and extrapulmonary) as a combined population | (1) Death, treatment failure, and loss to follow-up as a composite outcome; and (2) death as a single outcome | Multiple healthcare facilities | 4,749 | Prospective data collection for both exposures and outcomes by a dedicated research team | Logistic regression (N=4,749) |
| Yadav (2019) | Rajasthan | Urban and rural | Public sector | New and previously treated TB patients (including smear-positive pulmonary, smear-negative pulmonary, and extrapulmonary disease) with history of treatment interruption as a combined population | Medication non-adherence as a single outcome | Single healthcare facility* | 150 | Descriptive cross-sectional questionnaire-based | Reasons for medication nonadherence (i.e., treatment interruption) (N=9) |
| People with HIV treated for active TB |  |  |  |  |  |  |  |  |  |
| Sharma (2014) | New Delhi | Urban | Public sector | New and previously treated TB patients (including smear-positive pulmonary, smear-negative pulmonary, and extrapulmonary disease) with HIV | Death, treatment failure, and loss to follow-up as a composite outcome | Single healthcare facility* | 431 | Retrospective data collection from the government TB program* | Logistic Regression (N=431) |
| Shastri (2013) | Karnataka | Urban and rural | Public sector | New and previously treated TB patients (presumably including smear-positive pulmonary, smear-negative pulmonary, and extrapulmonary disease) with HIV | Death, treatment failure, and loss to follow-up as a composite outcome | Multiple healthcare facilities | 5,079 | Retrospective data collection from the government TB program* | Logistic Regression (N=5,040)^a^ |
| Vijay (2011) | Karnataka | Urban and rural | Public sector | New and previously treated TB patients (including smear-positive pulmonary, smear-negative pulmonary, and extrapulmonary disease) with HIV | (1) Death, treatment failure, and loss to follow-up as a composite outcome; and (2) death as a single outcome | Multiple healthcare facilities | 281 | Retrospective data collection from the government TB program* (a dedicated study team collected interviewed patients, but these data did not inform the regression analyses) | Logistic Regression (N=281) |
| Pediatric TB patients |  |  |  |  |  |  |  |  |  |
| Dhakulkar (2021) | Maharashtra | Urban | Public sector with collaborative support from the private sector (non-profit non-governmental organization) | Children (0-9 years old) and adolescents (10-19 years old) with multidrug-resistant TB, pre-extensively drug-resistant TB, and extensively drug-resistant TB | Death, treatment failure, and loss to follow-up as a composite outcome | Single healthcare facility* | 268 | Retrospective data collection from the government TB program* | Logistic Regression (N=268) |
| Raizada (2018) | New Delhi, Tamil Nadu, Telangana, West Bengal | Urban | Public sector and private sector | Children (0-14 years old) with drug-susceptible TB diagnosed by Xpert MTB/RIF testing | Death as a single outcome | Multiple healthcare facilities | 1,164 | Retrospective data collection from the government TB program or private sector* | Logistic Regression (N=1,164)^a^ |
| Sadana (2020) | Punjab | Urban and rural | Public sector | Children aged 0-14 years old | Death, treatment modified, and loss to follow-up as a composite outcome in children <15 years age | Single healthcare facility* | 62* | Evaluationg of exposures through baseline interview by a dedicated research team and assessment of outcomes through retrospective data collection from the government TB program* | Logistic Regression (N=62)^a^ |
| Satyanarayana (2010) | New Delhi | Urban | Public sector | Children 0-14 years old | Death, treatment failure, loss to follow-up, amd transferred out as a composite outcome | Multiple healthcare facilities | 1,074 | Retrospective data collection from the government TB program* | Logistic Regression (N=1,074)^a^ |

TB, tuberculosis

*Medium or low quality for this indicator

^a^Unadjusted odds ratios and/or p-values were estimated by the systematic review team from the raw data, as these were not provided in the original study.

###### Table D. Factors associated with patients diagnosed with tuberculosis not achieving treatment success (Gap 4)

| Study and specific outcome | Exposure / Independent variable | Unadjusted Effect Estimate (95% Confidence Interval) | P-value | Adjusted Effect Estimate (95% Confidence Interval) | P-value |
| --- | --- | --- | --- | --- | --- |
| Studies in new smear positive pulmonary TB patients who do not achieve treatment success as a single outcome or part of a composite outcome |  |  |  |  |  |
| Ahmed, 2009^a^ (Karnataka) *Outcome: Death, treatment failure, loss to follow-up, and transferred out as a composite outcome* |  | Values below are odds ratios |  |  |  |
|  | **Sex** |  |  |  |  |
|  | Female | Ref |  |  |  |
|  | Male | 2.16 (0.89-5.28) | 0.09 |  |  |
|  | **Distance from village of residence to treating health center** |  |  |  |  |
|  | <4 km | Ref |  |  |  |
|  | 5-9 km | 1.45 (0.6-3.5) | 0.41 |  |  |
|  | 10-14 km | 0.61 (0.13-2.92) | 0.53 |  |  |
|  | 15-19 km | 0.47 (0.06-3.92) | 0.48 |  |  |
|  | >20 km | 4.5 (1.40-14.42)* | 0.01* |  |  |
| Babiarz, 2014 (Bihar) *Outcome: Loss to follow-up as a single outcome (i.e., treatment discontinuation <25 weeks after initiation)* |  |  |  | Values below are adjusted odds ratios |  |
|  | **Gender** |  |  |  |  |
|  | Female |  |  | Ref |  |
|  | Male |  |  | 1.34 (0.81-2.22) |  |
|  | **Age^c^** |  |  |  |  |
|  | Per each year increase in age |  |  | 0.98 (0.93-1.03) |  |
|  | **Religion** |  |  |  |  |
|  | Non-Hindu |  |  | Ref |  |
|  | Hindu |  |  | 0.97 (0.51-1.84) |  |
|  | **Caste/tribe** |  |  |  |  |
|  | Other |  |  | Ref |  |
|  | Scheduled caste/tribe/OBC |  |  | 1.02 (0.51-2.02) |  |
|  | **Number of children in household^c^** |  |  |  |  |
|  | Per each increase in number of children |  |  | 1.08 (0.89-1.31) |  |
|  | **Education** |  |  |  |  |
|  | Yes |  |  | Ref |  |
|  | No |  |  | 1.01 (0.95-1.07) |  |
|  | **Poor** |  |  |  |  |
|  | No |  |  | Ref |  |
|  | Yes |  |  | 1.27 (0.73-2.19) |  |
|  | **Middle income** |  |  |  |  |
|  | No |  |  | Ref |  |
|  | Yes |  |  | 0.95 (0.56-1.61) |  |
|  | **Household size^c^** |  |  |  |  |
|  | Per each person increase in household |  |  | 0.92 (0.81-1.04) |  |
|  | **Total weeks from symptom onset to treatment initiation^c^** |  |  |  |  |
|  | Per each week increase in symptoms |  |  | 1.02 (0.97-1.08) |  |
|  | **Number of symptoms at treatment initiation** |  |  |  |  |
|  | >=5 |  |  | Ref |  |
|  | 2 or fewer |  |  | 1.29 (0.76-2.21) |  |
|  | 3 to 4 |  |  | 1.16 (0.65-2.09) |  |
|  | **Travel cost as a barrier** |  |  |  |  |
|  | No |  |  | Ref |  |
|  | Yes |  |  | 4.39 (1.36-14.13)* |  |
|  | **Number of providers visited^d^** |  |  |  |  |
|  | Per each provider increased |  |  | 5.62 (2.32-13.66)* |  |
|  | **Treatment or medication fees** |  |  |  |  |
|  | No |  |  | Ref |  |
|  | Yes |  |  | 6.32 (1.43-28.0)* |  |
| Bagchi, 2010^a^ (Maharashtra) *Population: New sputum smear positive pulmonary TB patients in the first two months of therapy Outcome: Medication non-adherence as a single outcome (i.e., at least one week's worth of missed TB medication doses in any treatment month)* |  | Values below are odds ratios |  | Values below are adjusted odds ratios |  |
|  | **Sex** |  |  |  |  |
|  | Female | Ref |  |  |  |
|  | Male | 0.7 (0.2-2.4) | Not reported |  |  |
|  | **Household members** |  |  |  |  |
|  | 0-3 | Ref |  |  |  |
|  | >3 | 0.3 (0.0-2.7) | Not reported | 0.3 (0.03-2.9) | Not reported |
|  | **Smoking status** |  |  |  |  |
|  | Never smoked | Ref |  | Ref |  |
|  | Ever smoked | 8.2 (1.4-46)* | Not reported | 7.8 (1.2-49)* | Not reported |
|  | **Tobacco chewing** |  |  |  |  |
|  | Never used | Ref |  | Ref |  |
|  | Ever used | 2.0 (0.-6.9) | Not reported | 1.6 (0.3-2.8) | Not reported |
|  | **Alcohol use** |  |  |  |  |
|  | Never used | Ref |  |  |  |
|  | Ever used | 1.3 (0.1-12.4) | Not reported |  |  |
|  | **Hid TB disease from family** |  |  |  |  |
|  | No | Ref |  |  |  |
|  | Yes | 0.6 (0.1-3.1) | Not reported |  |  |
|  | **Treatment duration perceived by patient as being too long** | ` |  |  |  |
|  | No | Ref |  | Ref |  |
|  | Yes | 2.9 (0.9-9.9) | Not reported | 1.4 (0.3-7.4) | Not reported |
|  | **Treatment discontinued once symptoms resolved** |  |  |  |  |
|  | No | Ref |  | Ref |  |
|  | Yes | 2.9 (0.8-11) | Not reported | 0.9 (0.1-5.8) | Not reported |
|  | **Knows about problems with stopping treatment earlly** |  |  |  |  |
|  | Yes | Ref |  | Ref |  |
|  | No | 2.1 (0.6-6.7) | Not reported | 0.8 (0.2-3.9) | Not reported |
|  | **Feels confident about completing treatment** |  |  |  |  |
|  | Somewhat or very sure | Ref |  | Ref |  |
|  | Not at all | 0.5 (0.1-1.8) | Not reported | 0.6 (0.0-11) | Not reported |
|  | **Travel mode to health center** |  |  |  |  |
|  | Walking | Ref |  |  |  |
|  | Other | 4.7 (1.7-12)* | Not reported |  |  |
|  | **Travel to health center is a problem** |  |  |  |  |
|  | No | Ref |  |  |  |
|  | Yes | 7.1 (1.6-31)* | Not reported |  |  |
|  | **Has concerns about transportation to the health center** |  |  |  |  |
|  | No | Ref |  | Ref |  |
|  | Somewhat or very concerned | 4.3 (1.1-17)* | Not reported | 0.9 (0.7-1.9) | Not reported |
|  | **Concerned about distance to the health center** |  |  |  |  |
|  | No | Ref |  |  |  |
|  | Somewhat or very concerned | 4.3 (1.1-17)* | Not reported |  |  |
|  | **Concerned about the time to reach the health center** |  |  |  |  |
|  | No | Ref |  |  |  |
|  | Somewhat or very concerned | 4.2 (1.2-15)* | Not reported |  |  |
|  | **Costs-related barriers to travel to health center** |  |  |  |  |
|  | No | Not reported |  | Ref |  |
|  | Yes | Not reported | Not reported | 5.1 (1.4-19)* | Not reported |
|  | **Doctor communicated problems related to stopping medication early** |  |  |  |  |
|  | Yes | Ref |  | Ref |  |
|  | No | 2.2 (0.8-5.9) | Not reported | 2.8 (0.5-14) | Not reported |
|  | **Where patient gets most TB information** |  |  |  |  |
|  | DOTS center | Ref |  | Ref |  |
|  | Other sources | 2.0 (0.7-5.8) | Not reported | 0.8 (0.1-6.9) | Not reported |
| Balasubramanian, 2004 (Tamil Nadu) *Outcome: Loss to follow-up as a single outcome* |  | Values below are odds ratios |  | Values below are adjusted odds ratios |  |
|  | **Gender** |  |  |  |  |
|  | Female | Ref |  |  |  |
|  | Male | 2.5 (1.4-4.3)* | <0.001* |  |  |
|  | **Age (years)** |  |  |  |  |
|  | <45 | Ref |  |  |  |
|  | >=45 | 1.6 (1.1-2.3)* | <0.001* |  |  |
| Bhatt, 2017 (Tamil Nadu) *Outcome: Death as a single outcome* |  | Values below are odds ratios |  |  |  |
|  | **Type of treatment** |  |  |  |  |
|  | DOTS | Ref |  |  |  |
|  | SAT | 1.30 (0.37-4.62) |  |  |  |
| Bhatt, 2017 (Tamil Nadu) *Outcome: Treatment failure as a single outcome* |  | Values below are odds ratios |  |  |  |
|  | **Type of treatment** |  |  |  |  |
|  | DOTS | Ref |  |  |  |
|  | SAT | 1.13 (0.27-4.66) |  |  |  |
| Bhatt, 2017 (Tamil Nadu) *Outcome: Loss to follow-up as a single outcome* |  | Values below are odds ratios |  |  |  |
|  | **Type of treatment** |  |  |  |  |
|  | DOTS | Ref |  |  |  |
|  | SAT | 0.61 (0.25-1.50) |  |  |  |
| Bhatt, 2017^b^ (Tamil Nadu) *Outcome: Not achieving treatment success as a composite outcome* |  | Values below are odds ratios |  |  |  |
|  | **Type of treatment** |  |  |  |  |
|  | DOTS | Ref |  |  |  |
|  | SAT | 0.74 (0.36-1.51) |  |  |  |
| Chakrabarti, 2012^a^ (West Bengal) *Outcome: Death, treatment failure, loss to follow-up, and transferred out as a composite outcome* |  | Values below are odds ratios |  |  |  |
|  | **Population** |  |  |  |  |
|  | Non-tribal | Ref |  |  |  |
|  | Tribal | 1.14 (0.71-1.84) | 0.58 |  |  |
| Gopi, 2006^b^ (Tamil Nadu) *Outcome: Not achieving microbiological cure as assessed by sputum microscopy* |  | Values below are odds ratios |  | Values below are adjusted odds ratios |  |
|  | **Age (years)** |  |  |  |  |
|  | <45 | Ref |  | Ref |  |
|  | >=45 | 1.8 (1.4-2.3)* | <0.001* | 1.5 (1.1-2.1)* | <0.05* |
|  | **Sex** |  |  |  |  |
|  | Male | Ref |  | Ref |  |
|  | Female | 0.42 (0.29-0.59)* | <0.001* | 0.71 (0.45-1.14) |  |
|  | **Education** |  |  |  |  |
|  | Illiterate | Ref |  | Ref |  |
|  | Literate | 0.77 (0.59-1)* | <0.05* | 0.83 (0.59-1.18) |  |
|  | **Cough** |  |  |  |  |
|  | Cough >=4 weeks | Ref |  | Ref |  |
|  | Cough <4 weeks | 0.63 (0.45-0.86)* | <0.01* | 0.77 (0.53-1.12) |  |
|  | **Smear grade** |  |  |  |  |
|  | High | Ref |  | Ref |  |
|  | Low | 0.71 (0.56-0.91)* | <0.01* | 0.91 (0.66-1.24) |  |
|  | **Conversion** |  |  |  |  |
|  | Yes | Ref |  | Ref |  |
|  | No | 4.1 (3.1-5.5)* | <0.001* | 3.5 (2.6-4.8)* | <0.05* |
|  | **Body weight** |  |  |  |  |
|  | >40 kg | Ref |  |  |  |
|  | <=40 kg | 0.83 (0.63-1.11) | 0.2 |  |  |
|  | **Smoking** |  |  |  |  |
|  | Yes | Ref |  | Ref |  |
|  | No | 0.42 (0.32-0.55)* | <0.001* | 0.91 (0.62-1.33) |  |
|  | **Alcoholism** |  |  |  |  |
|  | No | Ref |  | Ref |  |
|  | Yes | 2.7 (2.1-3.6)* | <0.001* | 1.7 (1.2-2.4)* | <0.05* |
|  | **Patient delay** |  |  |  |  |
|  | <4 weeks | Ref |  |  |  |
|  | >=4 weeks | 0.83 (0.65-1.08) | 0.3 |  |  |
|  | **Diagnosis** |  |  |  |  |
|  | Community survey | Ref |  |  |  |
|  | Health facility | 0.83 (0.63-1.11) | 0.3 |  |  |
| Joseph, 2011^a^ (Karnataka) *Outcome:* *Treatment failure and loss to follow-up as a composite outcome* |  | Values below are odds ratios |  |  |  |
|  | **Sex** |  |  |  |  |
|  | Female | Ref |  |  |  |
|  | Male | 1.31 (0.60-2.85) | 0.5 |  |  |
|  | **Age (years)** |  |  |  |  |
|  | <=30 | Ref |  |  |  |
|  | 30-60 | 1.71 (0.77-3.78) | 0.18 |  |  |
|  | >60 | 5.27 (1.78-15.65)* | 0.003* |  |  |
|  | **Residence** |  |  |  |  |
|  | Urban | Ref |  |  |  |
|  | Rural | 1.25 (0.63-2.46) | 0.52 |  |  |
| Kulkarni, 2013 (Maharashtra) *Outcome: Medication nonadherence as a single outcome* |  | Values below are odds ratios |  |  |  |
|  | **Age (years)** |  |  |  |  |
|  | Other than 15-49 | Ref |  |  |  |
|  | 15-49 | 1.56 (0.93-2.63) | 0.06 |  |  |
|  | **Gender** |  |  |  |  |
|  | Female | Ref |  |  |  |
|  | Male | 2.51 (1.51-4.18)* | <0.001* |  |  |
|  | **Migrant** |  |  |  |  |
|  | No | Ref |  |  |  |
|  | Yes | 1.97 (1.38-2.82)* | <0.001* |  |  |
|  | **Education** |  |  |  |  |
|  | Post-high school diploma or more | Ref |  |  |  |
|  | Literate with any schooling until high school | 18.33 (1.02-328.99)* | 0.05* |  |  |
|  | Illiterate | 16.82 (0.89-317.71) | 0.06 |  |  |
|  | **Employment** |  |  |  |  |
|  | Unemployed | Ref |  |  |  |
|  | Employed | 1.63 (1.17-2.27)* | 0.004* |  |  |
|  | **Social class** |  |  |  |  |
|  | II | Ref |  |  |  |
|  | III | 2.62 (0.82-8.39) | 0.1 |  |  |
|  | IV and V | 6.32 (2.53-15.84)* | 0.0001* |  |  |
|  | **Female sex worker** |  |  |  |  |
|  | Not female sex worker | Ref |  |  |  |
|  | Female sex worker | 4.89 (2.73-8.76)* | 0.001* |  |  |
|  | **Smoking** |  |  |  |  |
|  | No | Ref |  |  |  |
|  | Yes | 1.88 (1.41-2.50)* | <0.001* |  |  |
|  | **Alcohol consumption** |  |  |  |  |
|  | No | Ref |  |  |  |
|  | Yes | 1.85 (1.38-2.48)* | <0.001* |  |  |
|  | **Knowledge about importance of regular treatment** |  |  |  |  |
|  | Satisfactory | Ref |  |  |  |
|  | Unsatisfactory | 2.20 (1.64-2.95)* | <0.001* |  |  |
|  | **Living with own family** |  |  |  |  |
|  | Yes | Ref |  |  |  |
|  | No | 3.20 (1.93-5.30)* | <0.001* |  |  |
| Mave, 2021 (Maharashtra) *Outcome: Death as a single outcome* |  | Values below are hazard ratios |  | Values below are adjusted hazard ratios |  |
|  | **Diabetes** |  |  |  |  |
|  | TB only without diabetes mellitus | Ref |  | Ref |  |
|  | Any diabetes mellitus | 5.06 (2.26-11.35)* | <0.001* | 4.36 (1.62-11.76)* | 0.004* |
|  | Newly diagnosed diabetes mellitus | 7.17 (2.67-19.27)* | <0.001* | 6.56 (2.18-19.71)* | 0.001* |
|  | Known diabetes mellitus before TB diagnosis | 4.20 (1.70-10.33)* | 0.002* | 3.14 (1.03-9.61)* | 0.045* |
|  | Diabetes mellitus being treated with metformin | 3.30 (1.18-9.28) * | 0.02* | 2.32 (0.67-8.08) | 0.2 |
|  | Diabetes mellitus not being treated with metformin | 7.13 (2.96-17.21)* | <0.001* | 6.17 (2.24-17.04)* | <0.001* |
| Mave, 2021 (Maharashtra) *Outcome: Treatment failure as a single outcome* |  | Values below are odds ratios |  | Values below are adjusted odds ratios |  |
|  | **Diabetes** |  |  |  |  |
|  | TB only without diabetes mellitus | Ref |  | Ref |  |
|  | Any diabetes mellitus | 0.56 (0.30-1.06) | 0.08 | 0.75 (0.36-1.58) | 0.46 |
| Mukherjee, 2009^a^ (West Bengal) *Outcome: Death, treatment failure, loss to follow-up, and transferred out as a composite outcome* |  | Values below are odds ratios |  |  |  |
|  | **Sputum smear status** |  |  |  |  |
|  | New sputum smear negative pulmonary TB patients | Ref |  |  |  |
|  | New sputum smear positive pulmonary TB patients | 0.72 (0.58-0.89)* | 0.002* |  |  |
| Mukherjee, 2012^a^ (West Bengal) *Outcome: Death, treatment failure, and loss to follow-up as a composite outcome* |  | Values below are odds ratios |  |  |  |
|  | **Sex** |  |  |  |  |
|  | Female | Ref |  |  |  |
|  | Male | 1.44 (0.95-2.22) | 0.09 |  |  |
| Ramachandran, 2020 (Maharashtra and Tamil Nadu) *Outcome: Treatment failure as a single outcome* |  | Values below are incident rate ratios |  | Values below are adjusted incident rate ratios |  |
|  | **Rifampicin^c^** |  |  |  |  |
|  | 1 unit decrease in drug concentration | 1.36 (1.04-1.26) | 0.005* | 1.16 (1.05-1.28)* | 0.003* |
|  | **Isoniazid^c^** |  |  |  |  |
|  | 1 unit decrease in drug concentration | 1.06 (0.00-1.13) | 0.076 | 1.06 (0.99-1.14) | 0.1 |
|  | **Pyrazinamide^c^** |  |  |  |  |
|  | 1 unit decrease in drug concentration | 1.01 (0.99-1.04) | 0.255 | 1.02 (0.99-1.04) | 0.13 |
| Ramachandran, 2020 (Maharashtra and Tamil Nadu) *Outcome: Death as a single outcome* |  |  |  | Values below are adjusted incident rate ratios |  |
|  | **Rifampicin^c^** |  |  |  |  |
|  | 1 unit decrease in drug concentration | 1.07 (0.96-1.18) | 0.205 | 1.04 (0.94-1.15) | 0.47 |
|  | **Isoniazid^c^** |  |  |  |  |
|  | 1 unit decrease in drug concentration | 1.05 (0.97-1.14) | 0.212 | 1.04 (0.72-1.13) | 0.28 |
|  | **Pyrazinamide^c^** |  |  |  |  |
|  | 1 unit decrease in drug concentration | 1.01 (0.98-1.03) | 0.821 | 1.01 (0.98-1.04) | 0.62 |
| Shameer, 2016 (Kerala) *Outcome: Medication nonadherence (i.e., missing 3 or more consecutive doses)* |  | Values below are odds ratios |  | Values below are adjusted odds ratios |  |
|  | **Age (years)** |  |  |  |  |
|  | >45 | Ref |  |  |  |
|  | <45 | 1.69 (0.83-3.4) | 0.15 |  |  |
|  | **Sex** |  |  |  |  |
|  | Female | Ref |  |  |  |
|  | Male | 1.53 (0.67-3.70) | 0.33 |  |  |
|  | **Education** |  |  |  |  |
|  | Above primary school | Ref |  | Ref |  |
|  | Up to primary school | 3.5 (1.56-8.03)* | 0.003* | 2.25 (0.83-6.13) |  |
|  | **Occupation** |  |  |  |  |
|  | Others | Ref |  |  |  |
|  | Nil/unskilled | 0.82 (0.34-1.97) | 0.82 |  |  |
|  | **SES** |  |  |  |  |
|  | Middle | Ref |  | Ref |  |
|  | Lower | 2.72 (0.87-8.53) | 0.09 | 1.02 (0.25-4.07) |  |
|  | **Type of TB** |  |  |  |  |
|  | Extrapulmonary | Ref |  |  |  |
|  | Pulmonary | 1.36 (0.57-3.25) | 0.52 |  |  |
|  | **Adverse drug reactions** |  |  |  |  |
|  | Absent | Ref |  | Ref |  |
|  | Present | 2.91 (1.41-6.02)* | 0.006* | 2.46 (1.07-6.14)* | <0.05* |
|  | **Initial counseling** |  |  |  |  |
|  | Not received | Ref |  |  |  |
|  | Received | 1 (0.23-4.18) | 1 |  |  |
|  | **Selection of DOT center after consulting with patient** |  |  |  |  |
|  | Yes | Ref |  |  |  |
|  | No | 2.67 (0.68-10.48) | 0.16 |  |  |
|  | **Current smoker** |  |  |  |  |
|  | No | Ref |  | Ref |  |
|  | Yes | 8.34 (2.8-24.81)* | <0.001* | 3.84 (0.92-16.06) |  |
|  | **Alcohol** |  |  |  |  |
|  | Never used | Ref |  | Ref |  |
|  | Non-hazardous use | 1.84 (0.75-4.01) | 0.21 |  |  |
|  | Hazardous use | 22.67 (5.76-89.16)* | <0.001* | 16.67 (3.22-61.42)* | <0.05* |
|  | **TB stigma perceived by the patient** |  |  |  |  |
|  | Lower degree | Ref |  |  |  |
|  | Higher degree | 3.04 (1.40-6.51)* | 0.004* |  |  |
|  | **Shared TB status with family** |  |  |  |  |
|  | No | Ref |  | Ref |  |
|  | Yes | 2.90 (1.06-7.94)* | 0.058* | 2.22 (0.52-9.50) |  |
|  | **Family support** |  |  |  |  |
|  | No | Ref |  |  |  |
|  | Yes | 0.64 (0.24-1.73) | 0.43 |  |  |
|  | **Distance to nearest PHI** |  |  |  |  |
|  | <=2 km | Ref |  | Ref |  |
|  | >2 km | 2.76 (1.15-6.58)* | 0.02* | 2.99 (0.99-8.99) |  |
|  | **Any conflict with DOT provider** |  |  |  |  |
|  | No | Ref |  |  |  |
|  | Yes | 2.2 (0.77-6.30) | 0.17 |  |  |
| Shewade, 2019 (multi-state) *Outcome: Death, treatment failure, loss to follow-up, and not evaluated as a composite outcome* |  |  |  | Values below are adjusted relative risk ratios |  |
|  | **Cohort including 572 patients** |  |  |  |  |
|  | Non-Axshya SAMVAD (passive case finding) |  |  | Ref |  |
|  | Axshya SAMVAD (active case finding) |  |  | 0.83 (0.56-1.21) |  |
|  | **Cohort including 465 patients** |  |  |  |  |
|  | Non-Axshya SAMVAD (passive case finding) |  |  | Ref |  |
| Shivam, 2014^a^ (West Bengal) *Outcome: Death, treatment failure, and loss to follow-up as a composite outcome* |  | Values below are odds ratios |  |  |  |
|  | **Sex** |  |  |  |  |
|  | Female | Ref |  |  |  |
|  | Male | 0.81 (0.51-1.30) | 0.38 |  |  |
| Singla, 2009 (Delhi) *Outcome: Treatment failure as a single outcome as compared to patients who achieved cure* |  | Values below are odds ratios |  |  |  |
|  | **Gender** |  |  |  |  |
|  | Female | Ref |  |  |  |
|  | Male | 1.13 (0.39-3.25) | 0.83 |  |  |
|  | **Age^c^** |  |  |  |  |
|  | Per each year increase in age | 1.02 (0.98-1.05) | 0.41 |  |  |
|  | **SES** |  |  |  |  |
|  | Upper | Ref |  |  |  |
|  | Upper-middle | 9.00 (0.10-831.85) | 0.34 |  |  |
|  | Lower-middle | 3.86 (0.12-126.74) | 0.45 |  |  |
|  | Upper-lower | 1.49 (0.06-37.50) | 0.81 |  |  |
|  | Lower | 5.00 (0.11-220.64) | 0.4 |  |  |
|  | **Duration of illness** |  |  |  |  |
|  | <2 months | Ref |  |  |  |
|  | >2 months | 14.8 (3.12-70.14)* | <0.001* |  |  |
|  | **Cavity** |  |  |  |  |
|  | Absent | Ref |  | Ref |  |
|  | Present | 16.02 (5.92-43.97)* | <0.001* | 18.35 (2.52-133)* | 0.004* |
|  | **Radiological extent of disease** |  |  |  |  |
|  | Less advanced | Ref |  |  |  |
|  | Far advanced | 4.63 (2.05-10.46)* | <0.001* |  |  |
|  | **Initial sputum grade** |  |  |  |  |
|  | Non-3+ | Ref |  |  |  |
|  | 3+ | 2.89 (1.33-6.23)* | 0.01* |  |  |
|  | **Interruptions in treatment (i.e., missing at least one medication dose)** |  |  |  |  |
|  | No | Ref |  | Ref |  |
|  | Yes | 1.76 (1.09-2.82)* | 0.03* | 2.018 (1.03-3.94)* | 0.04* |
|  | **Culture-positive at 5 months** |  |  |  |  |
|  | No | Ref |  |  |  |
|  | Yes | Infinite (0-infinity)* | <0.001* |  |  |
|  | **Smear-positive at 2 months** |  |  |  |  |
|  | No | Ref |  | Ref |  |
|  | Yes | 275.00 (33.47-2250)* | <0.001* | 219.9 (17.6-2603.2)* | <0.001* |
|  | **BMI^d^** |  |  |  |  |
|  | Per each unit increase in BMI | 0.91 (0.78-1.05) | 0.18 |  |  |
|  | **Alcohol use** |  |  |  |  |
|  | Non-drinker | Ref |  |  |  |
|  | Alcohol consumption | 1.42 (0.56-3.59) | 0.5 |  |  |
|  | **Smoking** |  |  |  |  |
|  | Non-smoker | Ref |  |  |  |
|  | Smoker | 1.24 (0.57-2.70) | 0.69 |  |  |
| Singla, 2013 (Delhi) *Outcome: Smear positive at 2 months as a single outcome* |  |  |  | Values below are adjusted odds ratios |  |
|  | **Duration of illness** |  |  |  |  |
|  | <=2 months |  |  | Ref |  |
|  | >2 months |  |  | 8.29 (2.10-32.70)* | 0.003* |
|  | **Cavity** |  |  |  |  |
|  | Absent |  |  | Ref |  |
|  | Present |  |  | 10.81 (2.42-48.22)* | 0.002* |
|  | **Radiological extent of disease** |  |  |  |  |
|  | Less advanced |  |  | Ref |  |
|  | Far advanced |  |  | 10.80 (3.12-37.39)* | <0.001* |
|  | **Number of interruptions (missed doses) in treatment in IP** |  |  |  |  |
|  | No interruptions |  |  | Ref |  |
|  | 1 to 2 |  |  | 4.74 (1.21-18.49)* | 0.03* |
|  | 3 or more |  |  | 15.56 (2.15-112.88)* | 0.007* |
| Singla, 2013 (Delhi) *Outcome: Culture positive at 2 months as a single outcome* |  | Values below are odds ratios |  |  |  |
|  | **Duration of illness** |  |  |  |  |
|  | <=2 months | Ref |  |  |  |
|  | >2 months | 2.38 (1.03-5.50)* | <0.001* |  |  |
|  | **Cavity** |  |  |  |  |
|  | Absent | Ref |  |  |  |
|  | Present | 10.81 (2.42-48.22)* | 0.002* |  |  |
|  | **Extensive disease** |  |  |  |  |
|  | No | Ref |  |  |  |
|  | Yes | 12.19 (3.98-37.37)* | <0.001* |  |  |
|  | **Sputum smear** |  |  |  |  |
|  | 1 or 2 + | Ref |  |  |  |
|  | 3+ | 4.85 (1.84-12.75)* | <0.001* |  |  |
|  | **Interruptions** |  |  |  |  |
|  | No interruptions | Ref |  |  |  |
|  | Occasional interrupter | 1.39 (0.54-3.56) |  |  |  |
|  | Frequent interrupter | 4.31 (1.43-12.97) * | 0.02* |  |  |
| Singla, 2013^a^ (Delhi) *Outcome: Death, treatment failure, and loss to follow-up as a composite outcome* |  | Values below are odds ratios |  |  |  |
|  | **Smear status at 2 months** |  |  |  |  |
|  | Smear-negative at 2 months | Ref |  |  |  |
|  | Smear-positive at 2 months | 86.26 (5.14-1447.98)* | 0.002* |  |  |
|  | **Smear status at 3 months** |  |  |  |  |
|  | Smear-negative at 3 months | Ref |  |  |  |
|  | Smear-positive at 3 months | 19.05 (5.60-64.20)* | <0.001* |  |  |
|  | **IP interruption** |  |  |  |  |
|  | No IP interruption | Ref |  |  |  |
|  | Any IP interruption | 1.98 (0.84-4.66) | 0.12 |  |  |
|  | **IP interruption (more categories)** |  |  |  |  |
|  | Non-interrupter | Ref |  |  |  |
|  | Occasional interrupter | 1.12 (0.41-3.08) | 0.83 |  |  |
|  | Frequent interrupter | 4.30 (1.40-12.90)* | 0.009* |  |  |
| Tiwari, 2012^a^ (Delhi) *Outcome: Treatment failure, LTFU, transferred out and death as a composite outcome* |  | Values below are odds ratios |  |  |  |
|  | **High/low positive** |  |  |  |  |
|  | High positive | Ref |  |  |  |
|  | Low positive | 0.87 (0.49-1.57) | 0.65 |  |  |
|  | **Conversion at 2 months** |  |  |  |  |
|  | Converted at 2 months | Ref |  |  |  |
|  | Not converted at 2 months | 3.86 (1.03-7.32)* | <0.0001* |  |  |
|  | **Conversion at 3 months** |  |  |  |  |
|  | Converted at 3 months | Ref |  |  |  |
|  | Not converted at 3 months | 4.25 (1.66-10.90)* | 0.003* |  |  |
|  | Not converted at 2 months | 3.86 (1.03-7.32)* | <0.0001* |  |  |
|  | **Conversion at 3 months** |  |  |  |  |
|  | Converted at 3 months | Ref |  |  |  |
|  | Not converted at 3 months | 4.25 (1.66-10.90)* | 0.003* |  |  |
| Trivedi, 2019^a^ (Gujarat) *Outcome: Unable to achieve cure as a single outcome* |  | Values below are odds ratios |  |  |  |
|  | **Gender** |  |  |  |  |
|  | Female | Ref |  |  |  |
|  | Male | 1.09 (0.26-4.49) | 0.91 |  |  |
|  | **Age (years)** |  |  |  |  |
|  | <=40 | Ref |  |  |  |
|  | >40 | 1.92 (0.55-6.70) | 0.31 |  |  |
|  | **Residence** |  |  |  |  |
|  | Urban | Ref |  |  |  |
|  | Rural | 2.05 (0.51-8.31) | 0.31 |  |  |
|  | **Initial sputum colony** |  |  |  |  |
|  | 1+ or scanty | Ref |  |  |  |
|  | 2+ or 3+ | 0.58 (0.17-1.97) | 0.38 |  |  |
|  | **TB/HIV coinfected** |  |  |  |  |
|  | No | Ref |  |  |  |
|  | Yes | 1.54 (0.06-39.97) | 0.79 |  |  |
|  | **Personal habit (i.e., smoking, tobacco, or alcohol use)** |  |  |  |  |
|  | No | Ref |  |  |  |
|  | Yes | 3.57 (0.97-13.14) | 0.06 |  |  |
| Vashishtha, 2013^a^ (Delhi) *Outcome: Death, treatment failure, loss to follow-up, and treatment modified as a composite outcome* |  | Values below are odds ratios |  |  |  |
|  | **Outcome at end of initial ATT** |  |  |  |  |
|  | HIV-negative | Ref |  |  |  |
|  | HIV-positive | 8.89 (3.04-26.02)* | 0.0001* |  |  |
| Velayutham, 2014 (Tamil Nadu) *Outcome: Death as a single outcome* |  | Values below are odds ratios |  |  |  |
|  | **Age** |  |  |  |  |
|  | Younger | Ref |  |  |  |
|  | Elderly | 2.60 (1.70-3.90)* | <0.001* |  |  |
| Velayutham, 2014 (Tamil Nadu) *Outcome: Treatment failure as a single outcome* |  | Values below are odds ratios |  |  |  |
|  | **Age** |  |  |  |  |
|  | Younger | Ref |  |  |  |
|  | Elderly | 1.97 (0.54-1.66) | 0.92 |  |  |
| Velayutham, 2014 (Tamil Nadu) *Outcome: Loss to follow-up as a single outcome* |  | Values below are odds ratios |  |  |  |
|  | **Age** |  |  |  |  |
|  | Younger | Ref |  |  |  |
|  | Elderly | 1.30 (0.90-1.80) | 0.09 |  |  |
| Velayutham, 2014 (Tamil Nadu) *Outcome: Death as a single outcome* |  | Values below are odds ratios |  |  |  |
|  | **Age** |  |  |  |  |
|  | Younger | Ref |  |  |  |
|  | Elderly | 2.60 (1.6-4.20)* | <0.001* |  |  |
| Velayutham, 2014 (Tamil Nadu) *Outcome: Treatment failure as a single outcome* |  | Values below are odds ratios |  |  |  |
|  | **Age** |  |  |  |  |
|  | Younger | Ref |  |  |  |
|  | Elderly | 0.91 (0.49-1.58) | 0.74 |  |  |
|  |  | Values below are odds ratios |  |  |  |
| Velayutham, 2014 (Tamil Nadu) *Outcome: Loss to follow-up as a single outcome* | **Age** |  |  |  |  |
|  | Younger | Ref |  |  |  |
|  | Elderly | 1.40 (1.0-1.97)* | 0.03* |  |  |
| Velayutham, 2018 (6 Indian States) *Outcome: Death, treatment failure, loss to follow-up, treatment modified, and not evaluated as a composite outcome* |  | Values below are relative risk ratios |  | Values below are adjusted relative risk ratios |  |
|  | **Sex** |  |  |  |  |
|  | Female | Ref |  |  |  |
|  | Male | 1.08 (0.98-1.20) | 0.12 |  |  |
|  | **Age** |  |  |  |  |
|  | 18-24 | Ref |  |  |  |
|  | 25-34 | 1.03 (0.89-1.19) | 0.69 |  |  |
|  | 35-44 | 1.04 (0.89-1.21) | 0.61 |  |  |
|  | 45-54 | 1.03 (0.89-1.20) | 0.66 |  |  |
|  | 55-64 | 1.06 (0.90-1.24) | 0.48 |  |  |
|  | >65 | 1.01 (0.84-1.23) | 0.86 |  |  |
|  | **Baseline sputum smear grade** |  |  |  |  |
|  | Scanty/1+ | Ref |  |  |  |
|  | 2+/3+ | 1.02 (0.93-1.12) | 0.66 |  |  |
|  | **Baseline sputum culture grade** |  |  |  |  |
|  | Cols/1+ | Ref |  |  |  |
|  | 2+/3+ | 0.98 (0.89-1.08) | 0.64 |  |  |
|  | **Baseline drug susceptibility test** |  |  |  |  |
|  | Sensitive | Ref |  | Ref |  |
|  | Resistant to one or more drugs | 1.15 (0.98-1.34) | 0.08 | 1.14 (0.96-1.35) | 0.14 |
|  | **Body mass index** |  |  |  |  |
|  | 18.5-22.9 | Ref |  |  |  |
|  | >23 | 0.89 (0.72-1.09) | 0.28 |  |  |
|  | 16-18.4 | 1.04 (0.92-1.17) | 0.54 |  |  |
|  | <16 | 1.06 (0.94-1.19) | 0.35 |  |  |
|  | **Diabetes** |  |  |  |  |
|  | No diabetes | Ref |  |  |  |
|  | Not on anti-diabetic treatment | 0.92 (0.77-1.11) | 0.39 |  |  |
|  | On anti-diabetic treatment | 0.92 (0.79-1.06) | 0.25 |  |  |
|  | **HIV status** |  |  |  |  |
|  | Non-reactive | Ref |  | Ref |  |
|  | Reactive, not on ART | 1.51 (0.97-2.34) | 0.07 | 1.44 (0.68-3.03) | 0.34 |
|  | Reactive, on ART | 1.11 (0.84-1.47) | 0.45 | 1.04 (0.73-1.46) | 0.84 |
|  | **Smoker** |  |  |  |  |
|  | Non-smoker | Ref |  |  |  |
|  | Past smoker | 1.08 (0.98-1.18) | 0.12 |  |  |
|  | Current smoker | 1.16 (0.96-1.41) | 0.12 |  |  |
|  | **Alcohol use** |  |  |  |  |
|  | None | Ref |  | Ref |  |
|  | Past | 1.09 (1.00-1.20)* | 0.04* | 1.08 (0.97-1.20) | 0.17 |
|  | Current | 1.02 (0.72-1.46) | 0.9 | 1.00 (0.68-1.47) | 1 |
|  | **Duration of IP** |  |  |  |  |
|  | 2 months | Ref |  |  |  |
|  | 3 months | 1.09 (0.95-1.24) | 0.24 |  |  |
|  | **Type of DOT** |  |  |  |  |
|  | Health center-based | Ref |  |  |  |
|  | Community-based | 1.03 (0.94-1.34) | 0.53 |  |  |
|  | **Duration of symptoms to treatment initiation** |  |  |  |  |
|  | <3 weeks | Ref |  |  |  |
|  | 3-6 weeks | 0.98 (0.87-1.12) | 0.79 |  |  |
|  | 6-9 weeks | 1.00 (0.87-1.15) | 0.99 |  |  |
|  | 9-12 weeks | 0.98 (0.84-1.14) | 0.79 |  |  |
|  | >12 weeks | 1.03 (0.90-1.18) | 0.63 |  |  |
|  | **Missed doses in IP of treatment** |  |  |  |  |
|  | None | Ref |  | Ref |  |
|  | 1 to 6 | 1.10 (0.96-1.27) | 0.18 | 1.10 (0.95-1.29) | 0.2 |
|  | 7 to 12 | 1.28 (0.99-1.65) | 0.06 | 1.21 (0.91-1.59) | 0.19 |
|  | >12 | 1.31 (0.93-1.84) | 0.12 | 1.29 (0.91-1.81) | 0.15 |
| Vijay, 2010 (6 Indian States) *Outcome: Loss to follow-up as a single outcome* |  | Values below are odds ratios |  | Values below are adjusted odds ratios |  |
|  | **Age (years)** |  |  |  |  |
|  | 41 and over | Ref |  |  |  |
|  | <41 | 1.02 (0.79-1.32) | 0.88 |  |  |
|  | **Residency time** |  |  |  |  |
|  | 1 year or more | Ref |  |  |  |
|  | <1 year | 0.94 (0.53-1.66) | 0.82 |  |  |
|  | **Marital status** |  |  |  |  |
|  | Not married | Ref |  |  |  |
|  | Married | 0.87 (0.63-1.19) | 0.39 |  |  |
|  | **Literacy** |  |  |  |  |
|  | Literate | Ref |  | Ref |  |
|  | Not literate | 1.47 (1.12-1.92)* | 0.004* | 1.41 (1.03-1.92)* | 0.03* |
|  | **Employment** |  |  |  |  |
|  | Not employed | Ref |  |  |  |
|  | Employed | 0.95 (0.73-1.24) | 0.73 |  |  |
|  | **Number of earners** |  |  |  |  |
|  | Patient not sole earner | Ref |  |  |  |
|  | Patient sole earner | 0.97 (0.72-1.33) | 0.88 |  |  |
|  | **Drug side effects** |  |  |  |  |
|  | No side effects to drugs | Ref |  | Ref |  |
|  | Side effects to drugs | 3.14 (2.39-4.14)* | <0.001* | 2.55 (1.87-3.47)* | <0.001* |
|  | **Associated illness** |  |  |  |  |
|  | Did not have associated illness | Ref |  |  |  |
|  | Had associated illness | 0.77 (0.54-1.09) | 0.14 |  |  |
|  | **Satisfaction** |  |  |  |  |
|  | Satisfied with services | Ref |  | Ref |  |
|  | Unsatisfied with services | 12.04 (5.92-25.2)* | <0.001* | 1.73 (1.14-2.60)* | 0.009* |
|  | **Alcoholism** |  |  |  |  |
|  | Not alcoholic | Ref |  | Ref |  |
|  | Alcoholic | 1.93 (1.48-2.52)* | 0.01* | 1.72 (1.23-2.44)* | 0.002* |
|  | **Smoking** |  |  |  |  |
|  | Not a smoker | Ref |  | Ref |  |
|  | Smoker | 1.48 (1.14-1.92)* | 0.003* | 1.12 (0.77-1.64) | 0.55 |
|  | **Knowledge of TB and treatment** |  |  |  |  |
|  | Adequate | Ref |  | Ref |  |
|  | Inadequate or poor | 2.22 (1.70-3.00)* | <0.001* | 1.88 (1.35-2.63)* | <0.001* |
|  | **Nuclear family** |  |  |  |  |
|  | Not having a nuclear family | Ref |  |  |  |
|  | Having a nuclear family | 1.04 (0.79-1.37) | 0.77 |  |  |
|  | **Family support** |  |  |  |  |
|  | Not having family support | Ref |  |  |  |
|  | Having family support | 0.59 (0.23-1.49) | 0.22 |  |  |
|  | **Other commitments** |  |  |  |  |
|  | Did not have other commitments | Ref |  | Ref |  |
|  | Had other commitments | 2.44 (1.72-3.45)* | <0.01* | 3.22 (1.12-9.09)* | 0.03* |
|  | **Distance to DOT center** |  |  |  |  |
|  | >2 km | Ref |  |  |  |
|  | <=2 km | 0.91 (0.66-1.26) | 0.57 |  |  |
|  | **DOT timing** |  |  |  |  |
|  | No overlapping work hours and DOT timing | Ref |  |  |  |
|  | Overlapping work hours and DOT timing | 0.93 (0.61-1.42) | 0.71 |  |  |
|  | **Out station duties** |  |  |  |  |
|  | No out station duties during treatment | Ref |  |  |  |
|  | Out station duties during treatment | 2.23 (1.05-4.72)* | 0.03* |  |  |
|  | **Patient-provider interaction** |  |  |  |  |
|  | Adequate | Ref |  | Ref |  |
|  | Inadequate | 2.0 (1.48-2.71)* | <0.001* | 1.72 (1.23-2.44)* | <0.001* |
|  | **Health staff support** |  |  |  |  |
|  | Adequate | Ref |  | Ref |  |
|  | Inadequate | 8.52 (3.40-22.83)* | <0.001* | 1.93 (1.41-2.64)* | <0.001* |
|  | **Address verification** |  |  |  |  |
|  | Address verification done | Ref |  | Ref |  |
|  | Address verification not done | 1.34 (1.01-1.77)* | 0.03* | 1.37 (1.00-1.88) | 0.053 |
|  | **DOT location** |  |  |  |  |
|  | DOT not at health/sub center | Ref |  |  |  |
|  | DOT at health/sub center | 0.84 (0.59-1.19) | 0.32 |  |  |
|  | **DOT used** |  |  |  |  |
|  | DOT done | Ref |  | Ref |  |
|  | DOT not done | 1.35 (1.03-1.80)* | 0.03* | 1.101 (0.73-1.39) | 0.93 |
|  | **Instances of missed doses** |  |  |  |  |
|  | Had not missed any doses | Ref |  | Ref |  |
|  | Instances of missed doses | 3.25 (2.42-3.47)* | <0.001* | 2.56 (1.82-3.57)* | <0.001* |
| Viswanathan, 2014^a^ (Tamil Nadu) *Outcome: Death, treatment failure, treatment modified, and loss to follow-up as a composite outcome* |  | Values below are odds ratios |  |  |  |
|  | **Diabetic status** |  |  |  |  |
|  | Non-diabetic | Ref |  |  |  |
|  | Diabetic | 0.89 (0.25-3.13) | 0.86 |  |  |
| Viswanathan, 2014^a^ (Tamil Nadu) *Outcome: Treatment failure or treatment modified as a composite outcome* |  | Values below are odds ratios |  |  |  |
|  | **Diabetes treatment status** |  |  |  |  |
|  | Diabetes not treated | Ref |  |  |  |
|  | Diabetes treated with oral medications or insulin | 36.0 (3.23-401.5) | 0.004* |  |  |
| Zaman, 2014^a^ (Assam) *Outcome: Death, treatment failure, and loss to follow-up as a composite outcome* |  | Values below are odds ratios |  |  |  |
|  | **Time lag between onset of systems and start of treatment** |  |  |  |  |
|  | Began treatment within 6 weeks after onset of symptoms | Ref |  |  |  |
|  | Began treatment between 6 weeks and 6 months after onset of symptoms | 16.88 (2.15-132.51)* | 0.007* |  |  |
| Zaman, 2014^a^ (Assam) *Outcome: Loss to follow-up as a single outcome* |  | Values below are odds ratios |  |  |  |
|  | **Distance from house to DOTS center and association with loss to follow-up among patients with irregular adherence** |  |  |  |  |
|  | Within 1 km | Ref |  |  |  |
|  | >1 km | 114.33 (3.8-3395.10)* | 0.006* |  |  |
| Zhou, 2020 (Tamil Nadu) *Outcome: Loss to follow-up as a single outcome* |  | Values below are odds ratios |  | Values below are adjusted odds ratios |  |
|  | **Age**^c^ |  |  |  |  |
|  | Increase of 1 year | 1.01 (0.99-1.03) | Not reported | 1.01 (0.99-1.03) | Not reported |
|  | **Marital status** |  |  |  |  |
|  | Married/single/widowed | Ref |  | Ref |  |
|  | Separated/divorced | 4.04 (1.51-10.83)* | Not reported | 3.80 (1.39-10.38)* | Not reported |
|  | **Religion** |  |  |  |  |
|  | Hindu | Ref |  |  |  |
|  | Christian/Muslim | 0.57 (0.23-1.39) | Not reported |  |  |
|  | **Monthly income** |  |  |  |  |
|  | 5500 or less | Ref |  |  |  |
|  | More than 5500 | 1.53 (0.94-2.50) | Not reported |  |  |
|  | **Weight** |  |  |  |  |
|  | Normal weight or overweight | Ref |  |  |  |
|  | Underweight or severely underweight | 1.77 (1.03-3.05)* | Not reported |  |  |
|  | **Diabetic status** |  |  |  |  |
|  | Non-diabetic | Ref |  | Ref |  |
|  | Diabetic | 0.50 (0.29-0.87)* | Not reported | 0.52 (0.29-0.92)* | Not reported |
|  | **Smoking tobacco use** |  |  |  |  |
|  | Nonsmoker | Ref |  |  |  |
|  | Current smoker or former smoker | 1.92 (1.12-3.30)* | Not reported |  |  |
|  | **At risk alcohol use** |  |  |  |  |
|  | Not at risk | Ref |  | Ref |  |
|  | At risk | 2.20 (1.31-3.69)* | Not reported | 1.92 (1.12-3.27)* | Not reported |
|  | **Knowledge TB transmitted by cough** |  |  |  |  |
|  | No | Ref |  |  |  |
|  | Yes | 0.61 (0.36-1.06) | Not reported |  |  |
|  | Began treatment between 6 weeks and 6 months after onset of symptoms | 16.88 (2.15-132.51)* | 0.007* |  |  |
| Zaman, 2014^a^ (Assam) *Outcome: Loss to follow-up as a single outcome* |  | Values below are odds ratios |  |  |  |
|  | **Distance from house to DOTS center and association with loss to follow-up among patients with irregular adherence** |  |  |  |  |
|  | Within 1 km | Ref |  |  |  |
|  | >1 km | 114.33 (3.8-3395.10)* | 0.006* |  |  |
| Zhou, 2020 (Tamil Nadu) *Outcome: Loss to follow-up as a single outcome* |  | Values below are odds ratios |  | Values below are adjusted odds ratios |  |
|  | **Age**^c^ |  |  |  |  |
|  | Increase of 1 year | 1.01 (0.99-1.03) | Not reported | 1.01 (0.99-1.03) | Not reported |
|  | **Marital status** |  |  |  |  |
|  | Married/single/widowed | Ref |  | Ref |  |
|  | Separated/divorced | 4.04 (1.51-10.83)* | Not reported | 3.80 (1.39-10.38)* | Not reported |
|  | **Religion** |  |  |  |  |
|  | Hindu | Ref |  |  |  |
|  | Christian/Muslim | 0.57 (0.23-1.39) | Not reported |  |  |
|  | **Monthly income** |  |  |  |  |
|  | 5500 or less | Ref |  |  |  |
|  | More than 5500 | 1.53 (0.94-2.50) | Not reported |  |  |
|  | **Weight** |  |  |  |  |
|  | Normal weight or overweight | Ref |  |  |  |
|  | Underweight or severely underweight | 1.77 (1.03-3.05)* | Not reported |  |  |
|  | **Diabetic status** |  |  |  |  |
|  | Non-diabetic | Ref |  | Ref |  |
|  | Diabetic | 0.50 (0.29-0.87)* | Not reported | 0.52 (0.29-0.92)* | Not reported |
|  | **Smoking tobacco use** |  |  |  |  |
|  | Nonsmoker | Ref |  |  |  |
|  | Current smoker or former smoker | 1.92 (1.12-3.30)* | Not reported |  |  |
|  | **At risk alcohol use** |  |  |  |  |
|  | Not at risk | Ref |  | Ref |  |
|  | At risk | 2.20 (1.31-3.69)* | Not reported | 1.92 (1.12-3.27)* | Not reported |
|  | **Knowledge TB transmitted by cough** |  |  |  |  |
|  | No | Ref |  |  |  |
|  | Yes | 0.61 (0.36-1.06) | Not reported |  |  |
| Studies in patients with a prior TB treatment history who do not achieve treatment success as a single outcome or part of a composite outcome |  |  |  |  |  |
| Babiarz, 2014 (Bihar) *Outcome: Loss to follow-up as a single outcome (i.e., treatment discontinuation <25 weeks after initiation)* |  |  |  | Values below are adjusted odds ratios |  |
|  | **Gender** |  |  |  |  |
|  | Female |  |  | Ref |  |
|  | Male |  |  | 1.02 (0.39-2.69) |  |
|  | **Age^c^** |  |  |  |  |
|  | Per each year increase in age |  |  | 0.84 (0.70-1.01) |  |
|  | **Religion** |  |  |  |  |
|  | Non-Hindu |  |  | Ref |  |
|  | Hindu |  |  | 0.25 (0.08-0.78)* |  |
|  | **Caste/tribe** |  |  |  |  |
|  | Other |  |  | Ref |  |
|  | Scheduled caste, scheduled tribe, or other backward class |  |  | 0.36 (0.08-1.71) |  |
|  | **Number of children in household^c^** |  |  |  |  |
|  | Per each increase in number of children |  |  | 0.80 (0.53-1.20) |  |
|  | **Education** |  |  |  |  |
|  | Per each year increase in education |  |  | 0.75 (0.58-0.98)* |  |
|  | **Poor** |  |  |  |  |
|  | No |  |  | Ref |  |
|  | Yes |  |  | 0.76 (0.14-4.11) |  |
|  | **Middle income** |  |  |  |  |
|  | No |  |  | Ref |  |
|  | Yes |  |  | 0.99 (0.30-3.20) |  |
|  | **Household size^c^** |  |  |  |  |
|  | Per each person increase in household |  |  | 1.35 (0.97-1.88) |  |
|  | **Completed previous TB treatment** |  |  |  |  |
|  | No |  |  | Ref |  |
|  | Yes |  |  | 0.15 (0.03-0.73)* |  |
|  | **Total weeks from symptom onset to treatment initiation^c^** |  |  |  |  |
|  | Per each week increase in symptoms |  |  | 1.04 (0.90-1.20) |  |
|  | **Number of symptoms at treatment initiation** |  |  |  |  |
|  | >=5 |  |  | Ref |  |
|  | 2 or fewer |  |  | 4.20 (1.26-13.92)* |  |
|  | 3 to 4 |  |  | 2.55 (0.66-9.95) |  |
|  | **Travel cost as a barrier** |  |  |  |  |
|  | No |  |  | Ref |  |
|  | Yes |  |  | 1.43 (0.13-16.08) |  |
|  | **Number of providers visited^c^** |  |  |  |  |
|  | Per each provider increased |  |  | 3.08 (0.46-20.45) |  |
|  | **Treatment or medication fees** |  |  |  |  |
|  | No |  |  | Ref |  |
|  | Yes |  |  | 1.13 (0.08-16.48) |  |
| Bhagat, 2010^a^ (Maharashtra) *Outcome:* *Loss to follow-up as a single outcome* |  | Values below are odds ratios |  | Values below are adjusted odds ratios | Maybe move to "all comers"? |
|  | **Age (years)** |  |  |  |  |
|  | <24 | Ref |  |  |  |
|  | 25-34 | 1.74 (0.4-7.62) | 0.46 |  |  |
|  | 35-44 | 2.69 (0.58-12.6) | 0.21 |  |  |
|  | 45-54 | 1.39 (0.28-6.8) | 0.68 |  |  |
|  | **Sex** |  |  |  |  |
|  | Female | Ref |  | Ref |  |
|  | Male | 3.34 (0.71-15.67) | 0.13 | 3.96 (0.61-25.77) | 0.15 |
|  | **Religion** |  |  |  |  |
|  | Hindu | Ref |  |  |  |
|  | Muslim | 2.25 (0.79-6.43) | 0.13 |  |  |
|  | Others | 1.23 (0.33-4.52) | 0.75 |  |  |
|  | **Marital status** |  |  |  |  |
|  | Unmarried | Ref |  |  |  |
|  | Married | 2.02 (0.61-6.62) | 0.25 |  |  |
|  | **Literacy** |  |  |  |  |
|  | Literate | Ref |  | Ref |  |
|  | Illiterate | 5.28 (1.92-14.48)* | 0.001* | 3.51 (1.10-11.24)* | 0.03* |
|  | **Employment** |  |  |  |  |
|  | Unemployed | Ref |  | Ref |  |
|  | Employed | 3.86 (1.47-10.15)* | 0.006* | 3.52 (1.09-11.33)* | 0.04* |
|  | **Overcrowding** |  |  |  |  |
|  | Absent | Ref |  | Ref |  |
|  | Present | 1.79 (0.66-4.85) | 0.25 | 1.26 (0.37-4.25) | 0.71 |
|  | **History of TB** |  |  |  |  |
|  | Absent | Ref |  | Ref |  |
|  | Present | 5.07 (0.79-32.39) | 0.09 | 1.15 (0.11-11.98) | 0.91 |
|  | **Alcohol use** |  |  |  |  |
|  | Not alcoholic | Ref |  | Ref |  |
|  | Alcoholic | 4.66 (1.71-12.73)* | 0.003* | 3.41 (1.04-11.22)* | 0.04* |
|  | **Type of family** |  |  |  |  |
|  | Joint family | Ref |  | Ref |  |
|  | Nuclear family | 0.68 (0.24-1.94) | 0.47 | 0.67 (0.19-2.37) | 0.53 |
| Chakrabarti, 2012^a^ (West Bengal) *Outcome:* *Death, treatment failure, loss to follow-up, and transferred out as a composite outcome* |  | Values below are odds ratios |  |  |  |
|  | **Population** |  |  |  |  |
|  | Non-tribal | Ref |  |  |  |
|  | Tribal | 1.23 (0.41-3.72) | 0.71 |  |  |
| Chandrasekaram^a^, 2006 (Tamil Nadu) *Outcome: Death, treatment failure, loss to follow-up, and others (e.g., transferred out) as a composite outcome* |  | Values below are odds ratios |  |  |  |
|  | **Category of previously treated patient based on prior treatment outcome** |  |  |  |  |
|  | Relapse (completion of prior treatment) | Ref |  |  |  |
|  | Loss to follow-up during previous treatment | 2.65 (1.55-4.52)* | 0.0004* |  |  |
|  | Treatment failure | 1.73 (1.02-2.93)* | 0.04* |  |  |
| Deepa, 2013 (Andhra Pradesh) *Outcome: Death, treatment failure, loss to follow-up, and transferred out as a composite outcome* |  | Values below are relative risk ratios |  | Values below are adjusted relative risk ratios |  |
|  | **Sex** |  |  |  |  |
|  | Female | Ref |  | Ref |  |
|  | Male | 1.36 (1.1-1.69)* | Not reported | 1.36 (1.09-1.68)* | Not reported |
|  | **Age (years)** |  |  |  |  |
|  | <40 | Ref |  |  |  |
|  | >=40 | 1.2 (0.94-1.33) | Not reported |  |  |
|  | **INH-resistance** |  |  |  |  |
|  | No | Ref |  | Ref |  |
|  | Yes | 1.44 (1.18-1.78)* | Not reported | 1.46 (1.19-1.78)* | Not reported |
|  | **Type** |  |  |  |  |
|  | Relapse | Ref |  | Ref |  |
|  | TAD | 1.2 (0.99-1.44) | Not reported | 1.18 (0.98-1.42) | Not reported |
|  | Failure | 1.71 (1.35-2.16)* | Not reported | 1.62 (1.28-2.04)* | Not reported |
|  | **HIV status** |  |  |  |  |
|  | Negative | Ref |  | Ref |  |
|  | Positive | 1.34 (1.0-1.79) | Not reported | 1.34 (1.0-1.77) | Not reported |
|  | Unknown | 1.66 (1.11-2.48)* | Not reported | 1.68 (1.13-2.51)* | Not reported |
|  | **ART** |  |  |  |  |
|  | Received | Ref |  |  |  |
|  | Not received | 1.93 (1.14-3.29)* | Not reported |  |  |
| Joseph, 2011^a^ (Karnataka) *Outcome: Treatment failure and loss to follow-up as a composite outcome* |  | Values below are odds ratios |  |  |  |
|  | **Sex** |  |  |  |  |
|  | Female | Ref |  |  |  |
|  | Male | 0.71 (0.18-2.76) | 0.62 |  |  |
|  | **Age (years)** |  |  |  |  |
|  | <=30 | Ref |  |  |  |
|  | 30-60 | 1.05 (0.37-2.97) | 0.93 |  |  |
|  | >60 | 0.91 (0.20-4.10) | 0.9 |  |  |
|  | **Residence** |  |  |  |  |
|  | Urban | Ref |  |  |  |
|  | Rural | 1.48 (0.58-3.80) | 0.41 |  |  |
| Jha, 2010 (Nationally Representative Sample) *Outcome: Loss to follow-up as a single outcome* *as compared to treatment success or treatment failure* |  | Values below are odds ratios |  | Values below are adjusted odds ratios |  |
|  | **Sex** |  |  |  |  |
|  | Female | Ref |  | Ref |  |
|  | Male | 1.56 (1.28-1.89)* | <0.01* | 1.42 (1.16-1.73)* | <0.01* |
|  | **Age (years)** |  |  |  |  |
|  | 25-34 | Ref |  | 0.87 (0.68-1.09) | 0.23 |
|  | <15 | 0.44 (0.17-1.07) | 0.05 | 0.49 (0.19-1.22) | 0.12 |
|  | 15-24 | 0.85 (0.63-1.14) | 0.26 | 0.79 (0.59-1.04) | 0.1 |
|  | 35-44 | 1.19 (0.94-1.51) | 0.13 | Ref |  |
|  | 45-54 | 0.98 (0.76-1.26) | 0.84 | 0.81 (0.63-1.03) | 0.09 |
|  | 55-64 | 1.39 (1.03-1.89) | 0.26 | 1.12 (0.84-1.51) | 0.43 |
|  | >=65 | 0.72 (0.47-1.10) | 0.11 | 0.60 (0.40-0.91)* | 0.02* |
|  | **Classification** |  |  |  |  |
|  | Smear-positive | Ref |  |  |  |
|  | Smear-negative | 0.92 (0.74-1.14) | 0.42 |  |  |
|  | Smear-unknown | 0.65 (0.26-1.6) | 0.31 |  |  |
|  | Extrapulmonary | 0.63 (0.37-1.08) | 0.07 |  |  |
|  | **Outcome of previous TB treatment** |  |  |  |  |
|  | Relapse (previous treatment completed) | Ref |  | Ref |  |
|  | Previous treatment failure | 1.16 (0.85-1.60) | 0.33 | 1.14 (0.84-1.56) | 0.39 |
|  | Loss to follow-up during previous treatment | 1.41 (1.15-1.72)* | <0.01* | 1.31 (1.07-1.61)* | <0.01* |
|  | "Other" previously treated patient** | 1.04 (0.83-1.30) | 0.74 | 0.98 (0.77-1.24) | 0.86 |
|  | **Adverse Reaction** |  |  |  |  |
|  | Not documented defaulter | Ref |  |  |  |
|  | Documented defaulter | 27.6 (10.8-76.7)* | <0.01* |  |  |
|  | **Source of previous treatment** |  |  |  |  |
|  | RNTCP (i.e., public sector) | Ref |  | Ref |  |
|  | Non-RNTCP (i.e., private sector) | 1.31 (1.07-1.6)* | <0.01* | 1.28 (1.04-1.57)* | <0.01* |
|  | Data missing | 1.18 (0.96-1.44) | 0.1 | 1.14 (0.92-1.40) | 0.21 |
|  | **Nature of DOT provider** |  |  |  |  |
|  | Public health facility | Ref |  |  |  |
|  | Community provider | 0.92 (0.72-1.19) | 0.52 |  |  |
|  | Medical college | 0.44 (0.28-0.71)* | <0.01* |  |  |
|  | Private provider | 0.66 (0.49-0.88)* | <0.01* |  |  |
|  | NGO | 0.66 (0.41-1.05) | 0.06 |  |  |
|  | Data missing | 1.26 (0.24-7.11) | 0.75 |  |  |
|  | **Nature of DOT provider** |  |  |  |  |
|  | Other facility (e.g., community providers, medical providers, private practitioners, or non-governmental organizations) |  |  | Ref |  |
|  | Public health facility |  |  | 1.33 (1.11-1.60)* | <0.01* |
|  | **Missed doses during IP** |  |  |  |  |
|  | None | Ref |  |  |  |
|  | 1 or more | 1.66 (1.39-1.99)* | <0.01* |  |  |
|  | 2 or more | 1.69 (1.40-2.04)* | <0.01* |  |  |
|  | 3 or more | 1.68 (1.37-2.05)* | <0.01* |  |  |
|  | 4 or more | 1.86 (1.50-2.30)* | <0.01* |  |  |
|  | 5 or more | 1.93 (1.54-2.43)* | <0.01* |  |  |
|  | 6 or more | 1.9 (1.49-2.42)* | <0.01* |  |  |
|  | 10 or more | 1.93 (1.41-2.63)* | <0.01* |  |  |
| Mukherjee, 2009^a^ (West Bengal) *Outcome:* *Death, treatment failure, loss to follow-up, and transferred as a composite outcome* |  | Values below are odds ratios |  |  |  |
|  | **Outcome of previous TB treatment** |  |  |  |  |
|  | Relapse (previous treatment completed) | Ref |  |  |  |
|  | Previous treatment failure | 2.77 (1.42-5.38)* | 0.003* |  |  |
|  | Loss to follow-up during previous treatment | 2.55 (1.17-5.54)* | 0.02* |  |  |
|  | **Sputum grade (initial)** |  |  |  |  |
|  | Low grade (1+) | Ref |  |  |  |
|  | High grade (2+, 3+) | 1.73 (0.94-3.2) | 0.08 |  |  |
| Nagaraja, 2011^a^ (Andhra Pradesh) *Outcome: Death, treatment failure, loss to follow-up, and transferred as a composite outcome* |  | Values below are odds ratios |  |  |  |
|  | **Treatment category during prior treatment episode** |  |  |  |  |
|  | Category I | Ref |  |  |  |
|  | Category II | 2.31 (1.22-4.38)* | 0.009* |  |  |
|  | Category III | 0.58 (0.09-3.74) | 0.57 |  |  |
|  | **Sensitivity pattern** |  |  |  |  |
|  | Pan sensitive | Ref |  |  |  |
|  | Any resistance | 3.22 (1.39-7.5)* | 0.007* |  |  |
|  | Resistance to 'S' only | 0.29 (0.07-1.25) | 0.09 |  |  |
|  | Resistance to 'H' only | 3.29 (0.89-12.18) | 0.08 |  |  |
|  | Resistance to 'H' and 'S' | 16.94 (0.97-294.51) | 0.05 |  |  |
|  | Resistance to 'H' and 'E' | 5.26 (0.27-101.14) | 0.27 |  |  |
|  | Resistance to 'S' and 'E' | 1.74 (0.17-17.51) | 0.64 |  |  |
|  | Resistance to 'S,' 'H,' and 'E' | 1.74 (0.33-9.19) | 0.51 |  |  |
|  | Negative culture | 0.64 (0.32-1.29) | 0.21 |  |  |
|  | Non-tuberculous mycobacteria | 2.92 (0.14-162.93) | 0.49 |  |  |
| Pardeshi, 2010^a^ (Maharashtra) *Outcome:* *Loss to follow-up as a single outcome* |  | Values below are odds ratios |  |  |  |
|  | **Outcome of previous TB treatment** |  |  |  |  |
|  | Relapse (previous treatment completed) | Ref |  |  |  |
|  | Previous treatment failure | 0.95 (0.14-6.46) | 0.96 |  |  |
|  | Loss to follow-up during previous treatment | 2.91 (0.76-11.10) | 0.12 |  |  |
| Sarpal, 2014^a^ (Punjab) *Outcome: Death, treatment failure, and loss to follow-up as a composite outcome* |  | Values below are odds ratios |  |  |  |
|  | **Outcome of previous TB treatment** |  |  |  |  |
|  | Relapse (previous treatment completed) | Ref |  |  |  |
|  | Previous treatment failure | 3.58 (1.78-7.21)* | 0.0004* |  |  |
|  | Loss to follow-up during previous treatment | 2.09 (1.18-3.69)* | 0.01* |  |  |
|  | "Other" previously treated patient** | 0.18 (0.08-0.41)* | <0.0001* |  |  |
| Sarpal 2014^a^ (Punjab) *Outcome: Loss to follow-up as a single outcome* |  | Values below are odds ratios |  | Values below are adjusted odds ratios |  |
|  | **Age (years)** |  |  |  |  |
|  | <=35 | Ref |  | Ref |  |
|  | >35 | 1.41 (0.65-3.04) | 0.44 | 0.74 (0.32-1.73) | 0.49 |
|  | **Sex** |  |  |  |  |
|  | Female | Ref |  | Ref |  |
|  | Male | 19.17 (2.71-380.6)* | <0.001* | 6.84 (0.73-64.41) | 0.09 |
|  | **Religion** |  |  |  |  |
|  | Other than Hindu | Ref |  | Ref |  |
|  | Hindu | 1.73 (0.49-7.32) | 0.52 | 1.63 (0.46-5.72) | 0.45 |
|  | **Marital status** |  |  |  |  |
|  | Single (unmarried) | 0.57 (0.21-1.51) | 0.31 | 1.01 (0.34-2.98) | 0.99 |
|  | Not single | Ref |  | Ref |  |
|  | **Type of family** |  |  |  |  |
|  | Nuclear | 0.91 (0.36-0.38) | 0.99 | 0.85 (0.33-2.18) | 0.74 |
|  | Other than nuclear | Ref |  | Ref |  |
|  | **Socioeconomic status** |  |  |  |  |
|  | Low | Ref |  |  |  |
|  | Others | 1.63 (0.75-3.53) | 0.24 |  |  |
|  | **Place of residence** |  |  |  |  |
|  | Slum Urban | Ref |  | Ref |  |
|  | Rural | 1.13 (0.47-2.79) | 0.93 | 0.87 (0.35-2.18) | 0.76 |
|  | **Education** |  |  |  |  |
|  | Illiterate | Ref |  | 0.941 (0.32-2.76) | 0.91 |
|  | Literate | 1.21 (0.39-3.46) | 0.7 | Ref |  |
|  | **Substance use** |  |  |  |  |
|  | No substance use disorder | Not reported |  | Ref |  |
|  | Substance use disorder ("addicted") | Not reported | Not reported | 4.45 (1.28-15.48)* | 0.019* |
| Shivam, 2014^a^ (West Bengal) *Outcome:* *Death, treatment failure, and loss to follow-up as a composite outcome* |  | Values below are odds ratios |  |  |  |
|  | **Sex** |  |  |  |  |
|  | Female | Ref |  |  |  |
|  | Male | 1.55 (0.58-4.16) | 0.39 |  |  |
| Singla, 2009^a^ (Delhi) *Outcome: Death, treatment failure, loss to follow-up, and transferred out as a composite outcome* |  | Values below are odds ratios |  |  |  |
|  | **Culture status among sputum smear positive patients** |  |  |  |  |
|  | Smear-positive, culture negative | Ref |  |  |  |
|  | Smear-positive, culture positive | 18.75 (3.25-108.23)* | 0.001* |  |  |
|  | **Resistance** |  |  |  |  |
|  | Drug resistance but not multidrug resistance | Ref |  |  |  |
|  | Multidrug resistance | 0.25 (0.02-2.58) | 0.24 |  |  |
| Sisodia, 2006^a^ (Rajasthan) *Outcome: Death, treatment failure, loss to follow-up, and transferred out as a composite outcome* |  |  |  |  |  |
|  | **Outcome of previous TB treatment** | Values below are odds ratios |  |  |  |
|  | Relapse (previous treatment completed) | Ref |  |  |  |
|  | Loss to follow-up during previous treatment | 1.77 (1.28-2.45)* | 0.0006* |  |  |
|  | Previous treatment failure | 1.75 (0.80-3.86) | 0.16 |  |  |
|  | **Source of previous TB treatment** |  |  |  |  |
|  | Government (TB program or other government sector facility) | Ref |  |  |  |
|  | Private sector | 16.91 (7.02-40.72)* | <0.0001* |  |  |
| Srinath, 2010^a^ (Andhra Pradesh) *Population: all previously treated TB patients; Outcome: Death, treatment failure, loss to follow-up, transferred out, and not recorded as a composite outcome* |  | Values below are odds ratios |  |  |  |
|  | **Type of retreatment** |  |  |  |  |
|  | Others treated with Cat II | Ref |  |  |  |
|  | Smear-positive relapse | 1.43 (1.19-1.71)* | 0.0001* |  |  |
|  | Smear-positive failures | 3.39 (2.68-4.29)* | <0.0001* |  |  |
|  | Smear-positive treatment after default | 1.87 (1.50-2.20)* | <0.0001* |  |  |
| Srinath, 2010^a^ (Andhra Pradesh) *Population: previously treated "other" TB patients; Outcome: Death, treatment failure, loss to follow-up, transferred out, and not recorded as a composite outcome* |  | Values below are odds ratios |  |  |  |
|  | **Gender** |  |  |  |  |
|  | Female | Ref |  |  |  |
|  | Male | 1.43 (1.02-2.02)* | 0.04* |  |  |
|  | **Age (years)** |  |  |  |  |
|  | <15 | Ref |  |  |  |
|  | 15-64 | 1.24 (0.27-5.70) | 0.78 |  |  |
|  | **Site of TB** |  |  |  |  |
|  | Pulmonary | Ref |  |  |  |
|  | Extra-pulmonary | 0.56 (0.34-0.90)* | 0.02* |  |  |
|  | **HIV status** |  |  |  |  |
|  | HIV-negative | Ref |  |  |  |
|  | HIV-positive | 3.10 (1.39-6.90)* | 0.006* |  |  |
|  | HIV-unknown | 1.74 (0.97-3.12) | 0.06 |  |  |
| Velavan, 2018^a^ (Puducherry) *Outcome: Death, treatment failure, and loss to follow-up as a composite outcome* |  | Values below are relative risk ratios |  | Values below are adjusted relative risk ratios |  |
|  | **Sex** |  |  |  |  |
|  | Female | Ref |  | Ref |  |
|  | Male | 1.7 (0.9-3.1) |  | 1.5 (0.8-2.5) | 0.17 |
|  | **Age** |  |  |  |  |
|  | <15 | Ref |  |  |  |
|  | 15-29 | 1.8 (0.4-6.6) | Not reported |  |  |
|  | 30-44 | 2.3 (0.6-8.5) | Not reported |  |  |
|  | 45-59 | 1.5 (0.4-6.5) | Not reported |  |  |
|  | >60 | 1.6 (0.2-14.2) | Not reported |  |  |
|  | **Place of residence** |  |  |  |  |
|  | Urban | Ref |  |  |  |
|  | Rural | 0.8 (0.6-1.3) | Not reported |  |  |
|  | Peri-urban | 1.2 (0.7-2.2) | Not reported |  |  |
|  | **Outcome of previous TB treatment** |  |  |  |  |
|  | Relapse (previous treatment completed) | Ref |  | Ref |  |
|  | Previous treatment failure | 1.7 (1.1-2.4)* | Not reported | 1.7 (1.04-2.8)* | 0.03* |
|  | Loss to follow-up during previous treatment | 1.7 (1.1-2.5)* | Not reported | 1.6 (1.1-2.4)* | 0.01* |
|  | "Other" previously treated patient** | 0.4 (0.1-0.9)* | Not reported | 0.7 (0.3-1.8) | 0.4 |
|  | **Site of TB** |  |  |  |  |
|  | Pulmonary | Ref |  | Ref |  |
|  | Extrapulmonary | 0.1 (0.02-0.9)* | Not reported | 0.3 (0.03-2.3) | 0.22 |
|  | **HIV status** |  |  |  |  |
|  | Negative | Ref |  |  |  |
|  | Positive | 0.9 (0.2-5.3) | Not reported |  |  |
|  | **Pre-treatment weight** |  |  |  |  |
|  | >=40 kg | Ref |  | Ref |  |
|  | <40 kg | 1.9 (1.3-2.6)* | Not reported | 1.8 (1.3-2.5)* | 0.001* |
| Velayutham, 2014 (Tamil Nadu) *Patient population: all previously treated TB patients; Outcome: Loss to follow-up as a single outcome* |  | Values below are odds ratios |  |  |  |
|  | **Age** |  |  |  |  |
|  | Younger | Ref |  |  |  |
|  | Elderly | 0.90 (0.60-1.40) | 0.61 |  |  |
| Velayutham, 2014 (Tamil Nadu) *Patient population: all previously treated TB patients; Outcome: Death as a single outcome* |  | Values below are odds ratios |  |  |  |
|  | **Age** |  |  |  |  |
|  | Younger | Ref |  |  |  |
|  | Elderly | 1.50 (0.70-3.00) | 0.25 |  |  |
| Velayutham, 2014 (Tamil Nadu) *Patient population: all previously treated TB patients; Outcome: Treatment failure as a single outcome* |  | Values below are odds ratios |  |  |  |
|  | **Age** |  |  |  |  |
|  | Younger | Ref |  |  |  |
|  | Elderly | 0.41 (0.14-1.18) | 0.1 |  |  |
| Velayutham, 2014 (Tamil Nadu) *Patient population: previously treated smear-positive pulmonary TB patients; Outcome: Loss to follow-up as a single outcome* |  | Values below are odds ratios |  |  |  |
|  | **Age** |  |  |  |  |
|  | Younger | Ref |  |  |  |
|  | Elderly | 0.90 (0.50-1.40) | 0.5 |  |  |
| Velayutham, 2014 (Tamil Nadu) *Patient population: previously treated smear-positive pulmonary TB patients; Outcome: Treatment failure as a single outcome* |  | Values below are odds ratios |  |  |  |
|  | **Age** |  |  |  |  |
|  | Younger | Ref |  |  |  |
|  | Elderly | 0.42 (0.11-1.18) | 0.09 |  |  |
| Velayutham, 2014 (Tamil Nadu) *Patient population: previously treated smear-positive pulmonary TB patients; Outcome: Death as a single outcome* |  | Values below are odds ratios |  |  |  |
|  | **Age** |  |  |  |  |
|  | Younger | Ref |  |  |  |
|  | Elderly | 1.60 (0.60-3.30) | 0.28 |  |  |
| Studies in MDR-TB patients who do not achieve treatment success as a single outcome or part of a composite outcome |  |  |  |  |  |
| Bhatt, 2018 (Delhi) *Outcome: Death as a single outcome* |  | Values below are hazard ratios |  | Values below are adjusted hazard ratios |  |
|  | **Age^c^** |  |  |  |  |
|  | Per each year increase in age | Not reported |  | 1.007 (0.98-1.04) | 0.63 |
|  | **Sex** |  |  |  |  |
|  | Female | Not reported |  | Ref |  |
|  | Male | Not reported |  | 4.07 (1.47-11.24)* | 0.007* |
|  | **Support duration^c^** |  |  |  |  |
|  | Per each month increase in duration of a support package including counseling, nutritional supplementation, and cash transfer | Not reported |  | 0.88 (0.81-0.95)* | 0.0009* |
|  | **Initial BMI^c,***^** |  |  |  |  |
|  | Per each unit decrease in BMI | Not reported |  | 1.22 (1.07-1.39)* | 0.005* |
|  | **Family size^c^** |  |  |  |  |
|  | Per each increase in member of family | Not reported |  | 0.91 (0.72-1.14) | 0.4 |
| Bhatt, 2018 (Delhi) *Outcome: Treatment failure as a single outcome* |  |  |  | Values below are adjusted odds ratios |  |
|  | **Age^c^** |  |  |  |  |
|  | Per each year increase in age | Not reported |  | 1.02 (0.96-1.10) | 0.56 |
|  | **Sex** |  |  |  |  |
|  | Female | Not reported |  | Ref |  |
|  | Male | Not reported |  | 2.56 (0.64-12.24) | 0.2 |
|  | **Support duration^c^** |  |  |  |  |
|  | Per each month increase in duration of a support package including counseling, nutritional supplementation, and cash transfer | Not reported |  | 0.93 (0.85-0.998) | 0.051 |
|  | **Initial BMI^c,***^** |  |  |  |  |
|  | Per each unit decrease in BMI | Not reported |  | 1.06 (0.88-1.29) | 0.55 |
|  | **Family size^c^** |  |  |  |  |
|  | Per each increase in member of family | Not reported |  | 0.85 (0.58-1.20) | 0.35 |
| Bhatt, 2018 (Delhi)  *Outcome: Loss to follow-up as a single outcome* |  |  |  | Values below are adjusted odds ratios |  |
|  | **Age^c^** |  |  |  |  |
|  | Per each year increase in age | Not reported |  | 1.07 (1.02-1.13)* | 0.01* |
|  | **Sex** |  |  |  |  |
|  | Female | Not reported |  | Ref |  |
|  | Male | Not reported |  | 6.48 (1.40-40.27)* | 0.03* |
|  | **Support duration^c^** |  |  |  |  |
|  | Per each month increase in duration of a support package including counseling, nutritional supplementation, and cash transfer | Not reported |  | 0.75 (0.60-0.87)* | 0.002* |
|  | **Initial BMI^c,***^** |  |  |  |  |
|  | Per each unit decrease in BMI | Not reported |  | 1.26 (1.004-1.60)* | 0.04* |
|  | **Family size^c^** |  |  |  |  |
|  | Per each increase in member of family | Not reported |  | 1.19 (0.87-1.68) | 0.29 |
| Dela, 2017^a^ (Gujarat) *Outcome: Death, progression to extensively drug-resistant TB, loss to follow-up, and transfer out as a composite outcome* |  | Values below are odds ratios |  |  |  |
|  | **Adverse drug reaction** |  |  |  |  |
|  | Yes | Ref |  |  |  |
|  | No | 3.43 (1.61-7.27)* | 0.0013* |  |  |
| Dela, 2017^a^ (Gujarat) *Outcome: Medication non-adherence as a single outcome* |  | Values below are odds ratios |  |  |  |
|  | **Adverse drug reaction** |  |  |  |  |
|  | Yes | Ref |  |  |  |
|  | No | 5.35 (2.43-11.78)* | <0.0001* |  |  |
| Dole, 2017^b^ (Maharashtra) *Outcome: Death, treatment failure, and loss to follow-up as a composite outcome* |  | Values below are odds ratios |  |  |  |
|  | **Age (years)** |  |  |  |  |
|  | >50 | Ref |  |  |  |
|  | 18-50 | 0.70 (0.28-1.75) | 0.59 |  |  |
|  | **Sex** |  |  |  |  |
|  | Female | Ref |  |  |  |
|  | Male | 1.23 (0.61-2.48) | 0.68 |  |  |
|  | **Residence** |  |  |  |  |
|  | Urban | Ref |  |  |  |
|  | Rural | 2.44 (1.22-4.90)* | 0.02* |  |  |
|  | **X-ray findings** |  |  |  |  |
|  | Moderate | Ref |  |  |  |
|  | Advanced | 3.31 (1.51-7.24)* | 0.003* |  |  |
|  | **Weight (kg)** |  |  |  |  |
|  | 45-70 | Ref |  |  |  |
|  | 26-45 | 1.38 (0.68-2.78) | 0.48 |  |  |
|  | **Test used for diagnosis** |  |  |  |  |
|  | GeneXpert | Ref |  |  |  |
|  | Line probe assay | 2.51 (1.14-5.52)* | 0.03* |  |  |
| Duraisamy, 2014 (Kerala) *Outcome: Death, treatment failure, progression to extensively drug-resistant TB, loss to follow-up, treatment interruption due to adverse drug reaction, and transfer out as a composite outcome* |  | Values below are hazard ratios |  | Values below are adjusted hazard ratios |  |
|  | **Sex** |  |  |  |  |
|  | Female | Ref |  |  |  |
|  | Male | 0.9 (0.6-1.4) |  |  |  |
|  | **Age (years)** |  |  |  |  |
|  | 25-44 | Ref |  |  |  |
|  | 15-24 | 1.0 (0.5-1.9) |  |  |  |
|  | >44 | 1 (0.7-1.5) |  |  |  |
|  | **Living below poverty line** |  |  |  |  |
|  | No | Ref |  |  |  |
|  | Yes | 1.1 (0.7-1.7) |  |  |  |
|  | **Cavitary chest radiograph** |  |  |  |  |
|  | No | Ref |  |  |  |
|  | Yes | 0.7 (0.3-1.9) |  |  |  |
|  | **Number of previous TB episodes** |  |  |  |  |
|  | >=3 | Ref |  |  |  |
|  | 1 to 2 | 0.9 (0.6-1.5) |  |  |  |
|  | **At least 1 adverse drug event** |  |  |  |  |
|  | No | Ref |  |  |  |
|  | Yes | 1 (0.7-1.5) |  |  |  |
|  | **Hospitalization during treatment** |  |  |  |  |
|  | No | Ref |  | Ref |  |
|  | Yes | 1.7 (1.1-2.7)* |  | 1.5 (1.0-2.5)* |  |
|  | **HIV seropositive** |  |  |  |  |
|  | No | Ref |  |  |  |
|  | Yes | 20.3 (0 to infinity) |  |  |  |
|  | **Diabetes** |  |  |  |  |
|  | No | Ref |  |  |  |
|  | Yes | 0.9 (0.6-1.5) |  |  |  |
|  | **Alcohol before treatment** |  |  |  |  |
|  | No | Ref |  |  |  |
|  | Yes | 0.9 (0.6-1.5) |  |  |  |
|  | **Alcohol during treatment** |  |  |  |  |
|  | No | Ref |  | Ref |  |
|  | Yes | 4.9 (1.2-20.3)* |  | 4.3 (1.1-17.6)* |  |
|  | **Tobacco before treatment** |  |  |  |  |
|  | No | Ref |  | Ref |  |
|  | Yes | 0.9 (0.6-1.4) |  | 0.6 (0.2-1.7) |  |
|  | **Tobacco during treatment** |  |  |  |  |
|  | No | Ref |  | Ref |  |
|  | Yes | 1.7 (0.9-3.5) |  | 1.2 (0.3-5.0) |  |
| Duraisamy, 2014 (Kerala) *Outcome: Medication non-adherence (mean number of doses missed in the intensive phase of therapy) as a single outcome* |  | Values below represent the mean number of missed doses (standard deviation) |  | Value below represents the mean difference (95% confidence interval) in missed doses |  |
|  | **Alcohol consumption during treatment** |  |  |  |  |
|  | No | 0.6 (3.3) |  |  |  |
|  | Yes | 7.2 (14.2) |  | 6.6 (3.8-9.3) | <0.0001 |
| Duraisamy, 2014 (Kerala) *Outcome: Medication non-adherence (mean number of doses missed in the continuation phase of therapy) as a single outcome* |  | Values below represent the mean number of missed doses (standard deviation) |  | Value below represents the mean difference (95% confidence interval) in missed doses |  |
|  | **Alcohol consumption during treatment** |  |  |  |  |
|  | No | 4.4 (14.5) |  |  |  |
|  | Yes | 12.5 (28.5) |  | 8.1 (-0.3-16.5) | 0.06 |
| Isaakidis, 2012 (Maharashtra) *Outcome: Treatment failure, LTFU and death as a composite outcome in MDR-TB/HIV coi-infected patients* |  | Values below are odds ratios |  |  |  |
|  | **Occurrence of severe medication adverse event** |  |  |  |  |
|  | No | Ref |  |  |  |
|  | Yes | 1.12 (0.41-3.07) |  |  |  |
| Jain, 2014^a^ (Municipal Corporation Area, Western India) *Outcome: Death, treatment failure, and loss to follow-up as a composite outcome* |  | Values below are odds ratios |  |  |  |
|  | **Sex** |  |  |  |  |
|  | Female | Ref |  |  |  |
|  | Male | 2.27 (1.1-4.7)* | 0.03* |  |  |
|  | **Age (years)** |  |  |  |  |
|  | <=40 | Ref |  |  |  |
|  | >40 | 1.56 (0.63-3.83) | 0.34 |  |  |
|  | **Radiological extent** |  |  |  |  |
|  | No bilateral cavity | Ref |  |  |  |
|  | Bilateral cavity | 2.09 (0.87-5.04) | 0.09 |  |  |
|  | **Radiological improvement** |  |  |  |  |
|  | No | Ref |  |  |  |
|  | Yes | 0.006 (0.0004-0.1)* | 0.0004* |  |  |
|  | **Initial culture colony count** |  |  |  |  |
|  | 1+/scanty | Ref |  |  |  |
|  | 2+/3+ | 0.83 (0.41-1.67) | 0.6 |  |  |
|  | **Culture conversion within 3 months** |  |  |  |  |
|  | No | Ref |  |  |  |
|  | Yes | 0.098 (0.042-0.23)* | <0.0001* |  |  |
|  | **Drug resistance pattern** |  |  |  |  |
|  | <3 | Ref |  |  |  |
|  | >=3 | 0.87 (0.43-1.76) | 0.7 |  |  |
|  | **Concomitant disease** |  |  |  |  |
|  | No | Ref |  |  |  |
|  | Yes | 1.45 (0.4-5.2) | 0.57 |  |  |
|  | **Smoking** |  |  |  |  |
|  | No | Ref |  |  |  |
|  | Yes | 2.51 (1.18-5.38)* | 0.02* |  |  |
|  | **Alcohol** |  |  |  |  |
|  | No | Ref |  |  |  |
|  | Yes | 4.66 (1.64-13.26)* | 0.004* |  |  |
|  | **Tobacco chewing** |  |  |  |  |
|  | No | Ref |  |  |  |
|  | Yes | 1.49 (0.73-3.03) | 0.27 |  |  |
| Janmeja, 2017^b^ (Chandigarh) *Outcome: Death, treatment failure, modification of therapy, and loss to follow-up as a composite outcome* |  | Values below are odds ratios |  | Values below are adjusted odds ratios |  |
|  | **Age^c,***^** |  |  |  |  |
|  | Per each year increase in age | 1.03 (1.0-1.05)* | 0.002* | 1.05 (1.0-1.09)* | 0.01* |
|  | **Gender^***^** |  |  |  |  |
|  | Female | Ref |  |  |  |
|  | Male | 1.26 (0.75-2.12) | 0.38 |  |  |
|  | **Unsuccessful outcome in previous anti-TB therapy^***^** |  |  |  |  |
|  | No | Ref |  |  |  |
|  | Yes | 2.08 (1.17-3.67)* | 0.01* |  |  |
|  | **Number of previous ATT courses >=1^***^** |  |  |  |  |
|  | No | Ref |  |  |  |
|  | Yes | 2.05 (1.15-3.67)* | 0.01* |  |  |
|  | **3 month sputum culture conversion^***^** |  |  |  |  |
|  | Yes | Ref |  |  |  |
|  | No | 1.79 (0.85-3.78) | 0.12 |  |  |
|  | **Treatment adherence^***^** |  |  |  |  |
|  | Yes | Ref |  |  |  |
|  | No | 3.74 (2.04-6.84)* | <0.001* | 4.52 (1.21-16.6)* | 0.02* |
|  | **Adverse drug reaction** |  |  |  |  |
|  | No | Ref |  |  |  |
|  | Yes | 1.79 (0.9-3.56) | 0.98 |  |  |
|  | **BMI^c^** |  |  |  |  |
|  | Per each unit decrease in BMI | 1.15 (1.06-1.24)* | 0.001* |  |  |
|  | **Diabetes** |  |  |  |  |
|  | No | Ref |  |  |  |
|  | Yes | 2.33 (0.98-5.15) | 0.053 |  |  |
|  | **Smoking** |  |  |  |  |
|  | No | Ref |  |  |  |
|  | Yes | 1.25 (0.7-2.23) | 0.46 |  |  |
|  | **Alcoholic** |  |  |  |  |
|  | No | Ref |  |  |  |
|  | Yes | 1.23 (0.66-2.3) | 0.48 |  |  |
|  | **Hemoglobin^c,***^** |  |  |  |  |
|  | Per each unit decrease in hemoglobin | 1.25 (1.09-1.43)* | 0.001* |  |  |
|  | **S. albumin^c,***^** |  |  |  |  |
|  | Per each unit decrease in albumin | 2.26 (1.34-3.80)* | 0.002* | 3.71 (1.22-11.3)* | 0.02* |
| Kandi, 2021^a^ (Telangana) *Outcome: Treatment failure, switch to XDR treatment, LTFU and death as a composite outcome* |  | Values below are odds ratios |  |  |  |
|  | **Gender** |  |  |  |  |
|  | Female | Ref |  |  |  |
|  | Male | 1.99 (1.31-3.02)* | 0.001* |  |  |
|  | **Age** |  |  |  |  |
|  | Younger than 50 |  |  |  |  |
|  | Older than 50 | 1.93 (0.99-3.74) | 0.053 |  |  |
|  | **Treatment initiation** |  |  |  |  |
|  | Less than 1 month | Ref |  |  |  |
|  | More than 1 month | 1.25 (0.67-2.32) | 0.48 |  |  |
|  | **Resistance** |  |  |  |  |
|  | Rifampin resistance | Ref |  |  |  |
|  | Rifampin and isoniazid resistance | 0.98 (0.65-1.48) | 0.98 |  |  |
|  | **Weight** |  |  |  |  |
|  | Healthy weight, overweight, and obese | Ref |  |  |  |
|  | Underweight | 2.00 (1.29-3.08)* | 0.002* |  |  |
|  | **Diabetic status** |  |  |  |  |
|  | Non-diabetic | Ref |  |  |  |
|  | Diabetic | 0.71 (0.33-1.53) | 0.38 |  |  |
|  | **Thyroid** |  |  |  |  |
|  | Euthyroid | Ref |  |  |  |
|  | Hypothyroid | 0.80 (0.50-1.28) | 0.34 |  |  |
|  | **HIV reactivity** |  |  |  |  |
|  | Non-reactive | Ref |  |  |  |
|  | Reactive | 1.34 (0.46-3.90) | 0.59 |  |  |
| Lohiya, 2020 (Delhi) *Outcome: Death, treatment failure, modification of therapy, treatment discontinuation for reasons other than adverse drug reactions, and loss to follow-up as a composite outcome* |  | Values below are relative risk ratios |  | Values below are adjusted relative risk ratios |  |
|  | **Gender** |  |  |  |  |
|  | Female | Ref |  | Ref |  |
|  | Male | 1.2 (0.8-1.8) | 0.2 | 1.3 (0.9-1.9) | 0.13 |
|  | **Age** |  |  |  |  |
|  | Younger than 15 | Ref |  | Ref |  |
|  | 15 and older | 1.6 (0.9-2.7) | 0.08 | 1.6 (0.8-3.0) | 0.14 |
|  | **TB site** |  |  |  |  |
|  | Lymph node | Ref |  |  |  |
|  | Other | 1.2 (0.8-1.7) | 0.4 |  |  |
|  | **Basis of diagnosis** |  |  |  |  |
|  | CBNAAT | Ref |  |  |  |
|  | Other basis | 1.0 (0.6-1.7) | 0.9 |  |  |
|  | **History of previous TB** |  |  |  |  |
|  | No | Ref |  | Ref |  |
|  | Yes | 2.3 (1.0-5.7)* | 0.04* | 2.1 (1.1-4.8)* | 0.03* |
|  | **Adverse reaction** |  |  |  |  |
|  | Yes | Ref |  | Ref |  |
|  | No | 1.3 (0.8-2.1) | 0.2 | 1.4 (0.9-2.2) | 0.15 |
|  | **Weight** |  |  |  |  |
|  | <30 kg | Ref |  | Ref |  |
|  | 31-50 kg | 2.0 (1.1-4.0)* | 0.02* | 1.8 (1.2-3.4)* | 0.02* |
|  | 50 kg or more | 1.8 (0.9-3.6) | 0.09 | 1.6 (0.8-3.0) | 0.1 |
|  | **HIV status** |  |  |  |  |
|  | Negative | Ref |  |  |  |
|  | Positive | 2.0 (0.9-4.5) | 0.3 |  |  |
|  | **Diabetic status** |  |  |  |  |
|  | Non-diabetic | Ref |  | Ref |  |
|  | Diabetic | 1.8 (0.9-3.8) | 0.2 | 1.9 (0.9-3.4) | 0.18 |
|  | **TB center (in Delhi)** |  |  |  |  |
|  | Drug-resistant TB Center 1 | Ref |  | Ref |  |
|  | Drug-resistant TB Center 2 [receives many patients from outside of Delhi] | 1.6 (1.0-2.5) | 0.06 | 1.5 (1.0-2.5)* | 0.05* |
|  | Drug-resistant TB Center 3 | 1.05 (0.6-1.9) | 0.8 | 1.0 (0.7-2.0) | 0.8 |
| Nair, 2016 (Tamil Nadu) *Outcome: Death, treatment failure, switched to extensively drug resistant TB treatment, interrupted treatment due to reasons other than adverse drug reaction, transfer out, and loss to follow-up as a composite outcome* |  | Values below are relative risk ratios |  | Values below are adjusted relative risk ratios |  |
|  | **Gender** |  |  |  |  |
|  | Female | Ref |  | Ref |  |
|  | Male | 1.4 (1.2-1.8)* | <0.001* | 1.4 (1.2-1.6)* | 0.001* |
|  | **Age (years)** |  |  |  |  |
|  | 15-44 | Ref |  | Ref |  |
|  | >=45 | 1.1 (0.9-1.3) | 0.13 | 1.1 (0.9-1.4) | 0.08 |
|  | **Presumptive MDRTB criteria** |  |  |  |  |
|  | Failure | Ref |  |  |  |
|  | Retreatment | 1.2 (0.9-1.4) | 0.1 |  |  |
|  | Smear-positive follow-up | 0.9 (0.6-1.4) | 0.67 |  |  |
|  | Other | 0.6 (0.3-1.4) | 0.15 |  |  |
|  | **Drug Resistance status** |  |  |  |  |
|  | Rifampicin+Isoniazid | Ref |  |  |  |
|  | Rifampicin only | 1.0 (0.9-1.2) | 0.63 |  |  |
|  | **Type of diagnostic test** |  |  |  |  |
|  | Rapid | Ref |  |  |  |
|  | CDST | 1.1 (0.9-1.3) | 0.13 |  |  |
|  | **Time to treatment** |  |  |  |  |
|  | <=14 days | Ref |  | Ref |  |
|  | 15-30 days | 1.1 (0.9-1.5) | 0.28 | 1.1 (0.9-1.4) | 0.36 |
|  | 31+ days | 1.3 (1.0-1.6)* | 0.04* | 1.3 (1.1-1.6)* | 0.04* |
|  | **HIV Status** |  |  |  |  |
|  | Non-reactive | Ref |  |  |  |
|  | Reactive | 1.0 (0.7-1.5) | 0.9 |  |  |
| Parmar, 2018 (7 States) *Outcome: Death treatment failure, and loss to follow-up as a composite outcome* |  | Values below are odds ratios |  |  |  |
|  | **Gender** |  |  |  |  |
|  | Female | Ref |  |  |  |
|  | Male | 1.38 (1.08-1.76)* | 0.01* |  |  |
|  | **Age** |  |  |  |  |
|  | <15 | Ref |  |  |  |
|  | 15-44 | 1.07 (0.27-4.33) | 0.92 |  |  |
|  | 45-64 | 1.06 (0.26-4.36) | 0.94 |  |  |
|  | >64 | 2.58 (0.53-12.65) | 0.24 |  |  |
|  | **Initial registration type** |  |  |  |  |
|  | Relapse | Ref |  |  |  |
|  | Loss to follow-up | 1.26 (0.85-1.86) | 0.25 |  |  |
|  | Treatment after failure | 1.01 (0.77-1.33) | 0.93 |  |  |
|  | New contacts | 1.44 (0.69-3.02) | 0.34 |  |  |
|  | Others | 1.74 (0.85-3.57) | 0.13 |  |  |
|  | **Previous number of treatment episodes^c^** |  |  |  |  |
|  | Per each increase in number of previous treatment episodes | 1.29 (1.09-1.53)* | <0.001* |  |  |
|  | **Retreatment regimen taken twice** |  |  |  |  |
|  | No | Ref |  |  |  |
|  | Yes | 0.8 (0.56-1.15) | 0.23 |  |  |
|  | **Duration of previous episodes^c^** |  |  |  |  |
|  | Per each increase in month of duration of previous episode | 0.98 (0.95-1.0)* | 0.02* |  |  |
|  | **Treatment deal by drug susceptibility testing method** |  |  |  |  |
|  | Phenotypic | Ref |  |  |  |
|  | Genotypic | 1.06 (0.1-11.05) | 0.96 |  |  |
|  | **Ethambutol resistance by Lowenstein-Jensen culture** |  |  |  |  |
|  | Resistance | Ref |  |  |  |
|  | Susceptible | 0.65 (0.48-0.89)* | 0.01* |  |  |
|  | **Streptomycin resistance by Lowenstein-Jensen culture** |  |  |  |  |
|  | Resistance | Ref |  |  |  |
|  | Susceptible | 0.63 (0.39-1.03) | 0.07 |  |  |
|  | **First-line drug resistance** |  |  |  |  |
|  | Rifampin only | Ref |  |  |  |
|  | Isoniazid and rifampin only | 1.67 (0.74-3.77) | 0.22 |  |  |
|  | Isoniazid and rifampin combination | 0.73 (0.27-1.98) | 0.53 |  |  |
|  | Rifampin combination | 2.72 (0.57-12.96) | 0.21 |  |  |
|  | **Treatment adherence (intensive phase)** |  |  |  |  |
|  | <7 missed doses | Ref |  |  |  |
|  | >=7 missed doses | 2.76 (2.03-3.77)* | <0.001* |  |  |
|  | **Treatment adherence (continuation phase)** |  |  |  |  |
|  | <7 missed doses | Ref |  |  |  |
|  | >=7 missed doses | 1.51 (1.15-1.98)* | <0.001* |  |  |
|  | **HIV status** |  |  |  |  |
|  | Negative | Ref |  |  |  |
|  | Positive | 1.08 (0.5-2.34) | 0.84 |  |  |
|  | Unknown | 0.65 (0.26-1.59) | 0.35 |  |  |
|  | **Diabetes** |  |  |  |  |
|  | No | Ref |  |  |  |
|  | Yes | 0.96 (0.63-1.44) | 0.83 |  |  |
|  | **Body mass index** |  |  |  |  |
|  | >=18 | Ref |  |  |  |
|  | <18 | 1.64 (1.28-2.11)* | <0.001* |  |  |
|  | **Cavitation** |  |  |  |  |
|  | No | Ref |  |  |  |
|  | Yes | 1.1 (0.87-1.39) | 0.44 |  |  |
|  | **Weight change at 6 months** |  |  |  |  |
|  | None | Ref |  |  |  |
|  | Any loss | 0.9 (0.59-1.37) | 0.63 |  |  |
|  | Any gain | 1.05 (0.76-1.47) | 0.75 |  |  |
|  | **Weight change at 12 months** |  |  |  |  |
|  | None | Ref |  |  |  |
|  | Any loss | 1.38 (0.81-2.33) | 0.23 |  |  |
|  | Any gain | 0.92 (0.59-1.43) | 0.7 |  |  |
|  | **Source of most recent previous treatment** |  |  |  |  |
|  | Government | Ref |  |  |  |
|  | Private | 0.89 (0.6-1.33) | 0.57 |  |  |
|  | Other | 0.53 (0.25-1.14) | 0.1 |  |  |
| Parmar, 2018 (7 States) *Outcome: Death as a single outcome* |  |  |  | Values below are adjusted odds ratios |  |
|  | **Gender** |  |  |  |  |
|  | Female |  |  | Ref |  |
|  | Male |  |  | 1.01 (0.75-1.36) | 0.94 |
|  | **Age (years)** |  |  |  |  |
|  | <15 |  |  | Ref |  |
|  | 15-44 |  |  | 0.68 (0.16-2.98) | 0.61 |
|  | 45-64 |  |  | 0.84 (0.19-3.78) | 0.82 |
|  | >64 |  |  | 2.29 (0.41-12.79) | 0.35 |
|  | **Initial registration type** |  |  |  |  |
|  | Relapse |  |  | Ref |  |
|  | Loss to follow-up |  |  | 1.33 (0.83-2.13) | 0.24 |
|  | Treatment after failure |  |  | 0.96 (0.67-1.36) | 0.82 |
|  | New contacts |  |  | 1.94 (0.82-4.56) | 0.13 |
|  | Others |  |  | 2.68 (1.2-5.99)* | 0.02* |
|  | **Previous number of treatment episodes^c^** |  |  |  |  |
|  | Per each increase in number of previous treatment episodes |  |  | 1.18 (0.93-1.48) | 0.17 |
|  | **Retreatment regimen taken twice** |  |  |  |  |
|  | No |  |  | Ref |  |
|  | Yes |  |  | 1.04 (0.67-1.61) | 0.88 |
|  | **Duration of previous episode^c^** |  |  |  |  |
|  | Per each increase in month of duration of previous episode |  |  | 0.99 (0.97-1.02) | 0.59 |
|  | **Cavitation** |  |  |  |  |
|  | No |  |  | Ref |  |
|  | Yes |  |  | 1.41 (1.05-1.91)* | 0.02* |
|  | Unknown |  |  | 1.53 (0.78-3.01) | 0.22 |
|  | **Treatment delay by drug susceptibility testing method** |  |  |  |  |
|  | Phenotypic |  |  | Ref |  |
|  | Genotypic |  |  | 1.41 (1.05-1.91)* | 0.02* |
|  | **Ethambutol resistance by Lowenstein-Jensen culture** |  |  |  |  |
|  | Resistance |  |  | Ref |  |
|  | Susceptible |  |  | 1.53 (0.78-3.01) | 0.22 |
|  | **Streptomycin resistance by Lowenstein-Jensen culture** |  |  |  |  |
|  | Resistance |  |  | Ref |  |
|  | Susceptible |  |  | 0.45 (0.24-0.84)* | 0.01* |
|  | Unknown |  |  | 0.6 (0.04-9.84) | 0.72 |
|  | **First-line drug resistance** |  |  |  |  |
|  | Rifampin only |  |  | Ref |  |
|  | Isoniazid and rifampin only |  |  | 1.48 (0.61-3.59) | 0.39 |
|  | Isoniazid and rifampin combination |  |  | 0.53 (0.17-1.71) | 0.29 |
|  | Rifampin combination |  |  | 3.62 (0.64-20.55) | 0.15 |
|  | **Treatment adherence (intensive phase)** |  |  |  |  |
|  | <7 missed doses |  |  | Ref |  |
|  | >=7 missed doses |  |  | 2.13 (1.46-3.12)* | <0.001* |
|  | Unknown |  |  | 0.08 (0.02-0.27)* | <0.001* |
|  | **Treatment adherence (continuation phase)** |  |  |  |  |
|  | <7 missed doses |  |  | Ref |  |
|  | >=7 missed doses |  |  | 0.98 (0.67-1.42) | 0.91 |
|  | Unknown |  |  | 22.06 (8.22-59.21)* | <0.001* |
|  | **HIV Status** |  |  |  |  |
|  | Negative |  |  | Ref |  |
|  | Positive |  |  | 0.76 (0.26-2.26) | 0.63 |
|  | Unknown |  |  | 0.73 (0.22-2.37) | 0.6 |
|  | **Diabetes** |  |  |  |  |
|  | No |  |  | Ref |  |
|  | Yes |  |  | 0.92 (0.53-1.61) | 0.78 |
|  | **Body mass index** |  |  |  |  |
|  | >=18 |  |  | Ref |  |
|  | <18 |  |  | 4.89 (3.4-7.06)* | <0.001* |
|  | **Weight change at 6 months** |  |  |  |  |
|  | No change |  |  | Ref |  |
|  | Any loss |  |  | 0.81 (0.49-1.35) | 0.41 |
|  | Any gain |  |  | 0.92 (0.62-1.37) | 0.68 |
|  | **Weight change at 12 months** |  |  |  |  |
|  | No change |  |  | Ref |  |
|  | Any loss |  |  | 1.34 (0.7-2.55) | 0.38 |
|  | Any gain |  |  | 0.52 (0.3-0.9)* | 0.02* |
|  | **Source of the most recent previous treatment** |  |  |  |  |
|  | Government |  |  | Ref |  |
|  | Private |  |  | 0.86 (0.52-1.43) | 0.55 |
|  | Other |  |  | 0.52 (0.19-1.42) | 0.2 |
| Parmar, 2018 (7 States) *Outcome: Treatment failure as a single outcome* |  |  |  | Values below are adjusted odds ratios |  |
|  | **Gender** |  |  |  |  |
|  | Female |  |  | Ref |  |
|  | Male |  |  | 1.6 (1.1-2.33)* | 0.02* |
|  | **Age (years)** |  |  |  |  |
|  | <15 |  |  | Ref |  |
|  | 15-44 |  |  | 0.67 (0-infinity) | 0.99 |
|  | 45-64 |  |  | 0.55 (0-infinity) | 0.99 |
|  | >64 |  |  | 0.12 (0-infinity) | 0.99 |
|  | **Initial registration type** |  |  |  |  |
|  | Relapse |  |  | Ref |  |
|  | Loss to follow-up |  |  | 1.19 (0.66-2.13) | 0.57 |
|  | Treatment after failure |  |  | 0.95 (0.61-1.47) | 0.82 |
|  | New contacts |  |  | 1.96 (0.67-5.72) | 0.22 |
|  | Others |  |  | 1.62 (0.53-4.94) | 0.39 |
|  | **Previous number of treatment episodes^c^** |  |  |  |  |
|  | Per each increase in number of previous treatment episodes |  |  | 1.04 (0.78-1.39) | 0.78 |
|  | **Retreatment regimen taken twice** |  |  |  |  |
|  | No |  |  | Ref |  |
|  | Yes |  |  | 1.12 (0.64-1.93) | 0.7 |
|  | **Duration of previous episode^c^** |  |  |  |  |
|  | Per each increase in month of duration of previous episode |  |  | 0.99 (0.96-1.03) | 0.75 |
|  | **Cavitation** |  |  |  |  |
|  | No |  |  | Ref |  |
|  | Yes |  |  | 0.89 (0.62-1.26) | 0.5 |
|  | Unknown |  |  | 0.27 (0.06-1.23) | 0.09 |
|  | **Treatment delay by drug susceptibility testing method** |  |  |  |  |
|  | Phenotypic |  |  | Ref |  |
|  | Genotypic |  |  | 0.55 (0.02-13.63) | 0.72 |
|  | **Ethambutol resistance by Lowenstein-Jensen culture** |  |  |  |  |
|  | Resistance |  |  | Ref |  |
|  | Susceptible |  |  | 0.6 (0.37-0.97)* | 0.04* |
|  | **Streptomycin resistance by Lowenstein-Jensen culture** |  |  |  |  |
|  | Resistance |  |  | Ref |  |
|  | Susceptible |  |  | 0.63 (0.31-1.29) | 0.21 |
|  | Unknown |  |  | 4.3 (0.17-107.94) | 0.38 |
|  | **First-line drug resistance** |  |  |  |  |
|  | Rifampin only |  |  | Ref |  |
|  | Isoniazid and rifampin only |  |  | 1.57 (0.46-5.35) | 0.48 |
|  | Isoniazid and rifampin combination |  |  | 0.72 (0.16-3.28) | 0.67 |
|  | Rifampin combination |  |  | 3.46 (0-infinity) | 0.98 |
|  | **Treatment adherence (intensive phase)** |  |  |  |  |
|  | <7 missed doses |  |  | Ref |  |
|  | >=7 missed doses |  |  | 2.29 (1.46-3.59)* | <0.001* |
|  | Unknown |  |  | 0.14 (0.01-1.8) | 0.13 |
|  | **Treatment adherence (continuation phase)** |  |  |  |  |
|  | <7 missed doses |  |  | Ref |  |
|  | >=7 missed doses |  |  | 1.9 (1.29-2.79)* | <0.001* |
|  | Unknown |  |  | 1.89 (0.37-9.64) | 0.44 |
|  | **HIV Status** |  |  |  |  |
|  | Negative |  |  | Ref |  |
|  | Positive |  |  | 1.82 (0.7-4.71) | 0.22 |
|  | Unknown |  |  | 0.38 (0.04-3.31) | 0.38 |
|  | **Diabetes** |  |  |  |  |
|  | No |  |  | Ref |  |
|  | Yes |  |  | 1.31 (0.71-2.41) | 0.39 |
|  | **Body mass index** |  |  |  |  |
|  | >=18 |  |  | Ref |  |
|  | <18 |  |  | 1.82 (1.2-2.76)* | 0.01* |
|  | **Weight change at 6 months** |  |  |  |  |
|  | No change |  |  | Ref |  |
|  | Any loss |  |  | 1.99 (1.04-3.79)* | 0.04* |
|  | Any gain |  |  | 1.59 (0.92-2.74) | 0.1 |
|  | **Weight change at 12 months** |  |  |  |  |
|  | No change |  |  | Ref |  |
|  | Any loss |  |  | 1.12 (0.52-2.41) | 0.77 |
|  | Any gain |  |  | 1.08 (0.56-2.09) | 0.81 |
|  | **Source of the most recent previous treatment** |  |  |  |  |
|  | Government |  |  | Ref |  |
|  | Private |  |  | 1.64 (0.9-2.99) | 0.1 |
|  | Other |  |  | 0.18 (0.04-0.86)* | 0.03* |
| Parmar, 2018 (7 States) *Outcome: Loss to follow-up as a single outcome* |  |  |  | Values below are adjusted odds ratios |  |
|  | **Gender** |  |  |  |  |
|  | Female |  |  | Ref |  |
|  | Male |  |  | 1.91 (1.38-2.66)* | <0.001* |
|  | **Age (years)** |  |  |  |  |
|  | <15 |  |  | Ref |  |
|  | 15-44 |  |  | 2.3 (0.23-23.23) | 0.48 |
|  | 45-64 |  |  | 2.19 (0.21-22.46) | 0.51 |
|  | >64 |  |  | 6.38 (0.53-76.79) | 0.14 |
|  | **Initial registration type** |  |  |  |  |
|  | Relapse |  |  | Ref |  |
|  | Loss to follow-up |  |  | 1.32 (0.81-2.14) | 0.27 |
|  | Treatment after failure |  |  | 0.72 (0.5-1.05) | 0.09 |
|  | New contacts |  |  | 1.2 (0.49-2.95) | 0.69 |
|  | Others |  |  | 0.84 (0.31-2.3) | 0.74 |
|  | **Previous number of treatment episodes^c^** |  |  |  |  |
|  | Per each increase in number of previous treatment episodes |  |  | 0.96 (0.75-1.22) | 0.72 |
|  | **Retreatment regimen taken twice** |  |  |  |  |
|  | No |  |  | Ref |  |
|  | Yes |  |  | 1.02 (0.63-1.64) | 0.94 |
|  | **Duration of previous episode^c^** |  |  |  |  |
|  | Per each increase in month of duration of previous episode |  |  | 1 (0.97-1.03) | 0.89 |
|  | **Cavitation** |  |  |  |  |
|  | No |  |  | Ref |  |
|  | Yes |  |  | 0.98 (0.72-1.34) | 0.91 |
|  | Unknown |  |  | 1.72 (0.88-3.36) | 0.11 |
|  | **Treatment delay by drug susceptibility testing method** |  |  |  |  |
|  | Phenotypic |  |  | Ref |  |
|  | Genotypic |  |  | 1.04 (0.07-15.31) | 0.98 |
|  | **Ethambutol resistance by Lowenstein-Jensen culture** |  |  |  |  |
|  | Resistance |  |  | Ref |  |
|  | Susceptible |  |  | 0.61 (0.41-0.91)* | 0.02* |
|  | **Streptomycin resistance by Lowenstein-Jensen culture** |  |  |  |  |
|  | Resistance |  |  | Ref |  |
|  | Susceptible |  |  | 0.6 (0.32-1.15) | 0.12 |
|  | Unknown |  |  | 2.41 (0.16-35.11) | 0.52 |
|  | **First-line drug resistance** |  |  |  |  |
|  | Rifampin only |  |  | Ref |  |
|  | Isoniazid and rifampin only |  |  | 2.19 (0.86-5.57) | 0.1 |
|  | Isoniazid and rifampin combination |  |  | 0.72 (0.22-2.41) | 0.6 |
|  | Rifampin combination |  |  | 4.88 (0.82-29.03) | 0.08 |
|  | **Treatment adherence (intensive phase)** |  |  |  |  |
|  | <7 missed doses |  |  | Ref |  |
|  | >=7 missed doses |  |  | 4.36 (2.97-6.39)* | <0.001* |
|  | Unknown |  |  | 0.05 (0.01-0.16)* | <0.01* |
|  | **Treatment adherence (continuation phase)** |  |  |  |  |
|  | <7 missed doses |  |  | Ref |  |
|  | >=7 missed doses |  |  | 1.85 (1.27-2.70)* | <0.001* |
|  | Unknown |  |  | 36.69 (13.7-98.4)* | <0.001* |
|  | **MDR-TB patients with baseline second line drug susceptibility testing** |  |  |  |  |
|  | Ofloxacin susceptible |  |  | Ref |  |
|  | Ofloxacin resistant |  |  | 3.19 (1.40-7.28)* | 0.006* |
|  | **HIV Status** |  |  |  |  |
|  | Negative |  |  | Ref |  |
|  | Positive |  |  | 0.55 (0.17-1.77) | 0.32 |
|  | Unknown |  |  | 1.24 (0.42-3.69) | 0.7 |
|  | **Diabetes** |  |  |  |  |
|  | No |  |  | Ref |  |
|  | Yes |  |  | 0.99 (0.56-1.73) | 0.97 |
|  | **Body mass index** |  |  |  |  |
|  | >=18 |  |  | Ref |  |
|  | <18 |  |  | 1.6 (1.12-2.29)* | 0.01* |
|  | **Weight change at 6 months** |  |  |  |  |
|  | No change |  |  | Ref |  |
|  | Any loss |  |  | 0.63 (0.36-1.11) | 0.11 |
|  | Any gain |  |  | 0.95 (0.62-1.47) | 0.83 |
|  | **Weight change at 12 months** |  |  |  |  |
|  | No change |  |  | Ref |  |
|  | Any loss |  |  | 2.27 (0.97-5.34) | 0.06 |
|  | Any gain |  |  | 1.7 (0.81-3.57) | 0.16 |
|  | **Source of the most recent previous treatment** |  |  |  |  |
|  | Government |  |  | Ref |  |
|  | Private |  |  | 1.07 (0.62-1.83) | 0.81 |
|  | Other |  |  | 0.54 (0.19-1.56) | 0.26 |
| Rupani, 2020 (Gujarat) *Outcome: Treatment discontinuation/non-adherence/interruption as a single outcome* |  |  |  | Values below are adjusted odds ratios |  |
|  | **Age (years)^c^** |  |  |  |  |
|  | Per each increase in year |  |  | 1.02 (0.95-1.09) | 0.65 |
|  | **Marital status** |  |  |  |  |
|  | Unmarried |  |  | 1.48 (0.28-7.7) | 0.64 |
|  | Married |  |  | Ref |  |
|  | **Years of schooling^c^** |  |  |  |  |
|  | Per each increase in year of schooling |  |  | 1.09 (0.87-1.35) | 0.45 |
|  | **Per Capita Income^c^** |  |  |  |  |
|  | Per each increase in capita income (unit of change undefined) |  |  | 1.0 (0.99-1.0) | 0.64 |
|  | **Duration of Treatment^c^** |  |  |  |  |
|  | Per each increase in treatment duration (unit of change undefined) |  |  | 1.06 (0.97-1.14) | 0.17 |
|  | **Intensive phase of MDR-TB** |  |  |  |  |
|  | Intensive phase |  |  | 2.31 (0.49-10.8) | 0.29 |
|  | Continuous phase |  |  | Ref |  |
|  | **Adverse drug reaction** |  |  |  |  |
|  | Present |  |  | 1.44 (0.27-7.6) | 0.67 |
|  | Absent |  |  | Ref |  |
| Saha, 2017^a^ (Maharashtra) *Outcome: Death, treatment failure, and loss to follow-up as a composite outcome* |  | Values below are odds ratios |  |  |  |
|  | **Age** |  |  |  |  |
|  | <45 | Ref |  | Ref |  |
|  | >=45 | 6.67 (1.55-28.62)* | 0.01* | 15.3 (1.69-138.99)* | 0.02* |
|  | **Gender** |  |  |  |  |
|  | Female | Ref |  | Ref |  |
|  | Male | 2.86 (0.85-9.63) | 0.1 | 2.09 (0.42-10.36) | 0.37 |
|  | **Past history of TB** |  |  |  |  |
|  | Absent | Ref |  |  |  |
|  | Present | 1.43 (0.44-4.67) | 0.75 |  |  |
|  | **Site of disease** |  |  |  |  |
|  | Extrapulmonary | Ref |  |  |  |
|  | Pulmonary | 2.63 (0.28-24.55) | 0.67 |  |  |
|  | **Comorbidities** |  |  |  |  |
|  | Without comorbidities | Ref |  |  |  |
|  | With comorbidities | 2.86 (0.70-11.65) | 0.21 |  |  |
|  | **Resistance to >=5 drugs** |  |  |  |  |
|  | No | Ref |  | Ref |  |
|  | Yes | 9.51 (2.50-38.18)* | 0.001* | 12.43 (2.04-75.88)* | 0.01* |
| Sharma, 2020 (Delhi) *Outcome: Death, treatment failure, switched to extensively drug-resistant TB therapy, treatment stopped due to adverse drug reaction, and loss to follow-up as a composite outcome* |  |  |  | Values below are adjusted odds ratios |  |
|  | **Age** |  |  |  |  |
|  | Younger than 18 |  |  | Ref |  |
|  | 18-34 |  |  | 1.19 (0.86-1.65)* | <0.001* |
|  | 35 and older |  |  | 2.1 (1.47-3)* | <0.001* |
|  | **Gender** |  |  |  |  |
|  | Female |  |  | Ref |  |
|  | Male |  |  | 1.4 (1.12-1.75)* | 0.002* |
|  | **Pretreatment body mass index** |  |  |  |  |
|  | Normal/overweight  (body mass index >=18.5) |  |  | Ref |  |
|  | Undernourished  (body mass index <18.5) |  |  | 1.88 (1.48-2.38)* | <0.001* |
| Shringarpure, 2015 (Gujarat) *Outcome: Loss to follow-up as a single outcome* |  | Values below are hazard ratios |  | Values below are adjusted hazard ratios |  |
|  | **Age (years)** |  |  |  |  |
|  | <35 | Ref |  | Ref |  |
|  | >35 | 0.99 (0.91-1.08) |  | 0.98 (0.78-1.23) |  |
|  | **Gender** |  |  |  |  |
|  | Female | Ref |  | Ref |  |
|  | Male | 1.14 (0.86-1.50) |  | 1.08 (0.49-1.67) |  |
|  | **Rural residence** |  |  |  |  |
|  | No | Ref |  | Ref |  |
|  | Yes | 0.95 (0.83-1.08) |  | 1.02 (0.76-1.38) |  |
|  | **Living below poverty line** |  |  |  |  |
|  | No | Ref |  | Ref |  |
|  | Yes | 0.97 (0.74-1.27) |  | 0.85 (0.49-1.47) |  |
|  | **Previous TB treatment** |  |  |  |  |
|  | No | Ref |  | Ref |  |
|  | Yes | 0.99 (0.991-0.997)* |  | 0.92 (0.83-1.02) |  |
|  | **Chest X-ray bilateral involvement** |  |  |  |  |
|  | No | Ref |  | Ref |  |
|  | Yes | 1.03 (0.95-1.11) |  | 0.88 (0.57-1.36) |  |
|  | **Chest X-Ray Cavitation** |  |  |  |  |
|  | No | Ref |  | Ref |  |
|  | Yes | 0.99 (0.92-1.08) |  | 1.08 (0.72-1.64) |  |
|  | **Culture Conversion Time >4 months** |  |  |  |  |
|  | No | Ref |  | Ref |  |
|  | Yes | 1.10 (1.06-1.15)* |  | 1.34 (1.21-1.49)* |  |
|  | **No adverse events in IP** |  |  |  |  |
|  | No | Ref |  | Ref |  |
|  | Yes | 0.75 (0.55-1.00) |  | 1.13 (0.61-2.09) |  |
|  | **No adverse events in CP** |  |  |  |  |
|  | No | Ref |  | Ref |  |
|  | Yes | 0.99 (0.65-1.49) |  | 2.63 (0.35-19.95) |  |
|  | **Weight >45 kg** |  |  |  |  |
|  | No | Ref |  | Ref |  |
|  | Yes | 0.89 (0.77-1.04) |  | 1.19 (0.87-1.61) |  |
|  | **Ambulatory initiation of treatment** |  |  |  |  |
|  | No | Ref |  | Ref |  |
|  | Yes | 1.68 (0.62-4.55) |  | 2.63 (1.01-6.86)* |  |
|  | **Different DOT provider in IP and CP** |  |  |  |  |
|  | No | Ref |  | Ref |  |
|  | Yes | 1.15 (1.11-1.19)* |  | 1.27 (1.18-1.38)* |  |
| Studies in multiple populations of TB patients who do not achieve treatment success as a single outcome or part of a composite outcome |  |  |  |  |  |
| Babiarz, 2014 (Bihar) *Population: New and previously treated TB patients (sputum smear positive pulmonary, sputum smear negative pulmonary, and extrapulmonary) as a combined population Outcome: Loss to follow-up as a single outcome (i.e., treatment discontinuation <25 weeks after initiation)* |  | Values below are odds ratios |  | Values below are adjusted odds ratios |  |
|  | **Gender** |  |  |  |  |
|  | Female | Ref |  | Ref |  |
|  | Male | 1.29 (0.85-1.95) |  | 1.32 (0.82-2.14) |  |
|  | **Age^c^** |  |  |  |  |
|  | Per each year increase in age | 0.96 (0.91-1.00) |  | 0.96 (0.91-1.01) |  |
|  | **Religion** |  |  |  |  |
|  | Non-Hindu | Ref |  | Ref |  |
|  | Hindu | 0.83 (0.51-1.35) |  | 0.81 (0.47-1.40) |  |
|  | **Caste/tribe** |  |  |  |  |
|  | Other | Ref |  | Ref |  |
|  | Scheduled caste/tribe/OBC | 0.87 (0.53-1.44) |  | 0.84 (0.47-1.50) |  |
|  | **Number of children in household^c^** |  |  |  |  |
|  | Per each increase in number of children | 0.95 (0.82-1.09) |  | 1.02 (0.86-1.21) |  |
|  | **Education** |  |  |  |  |
|  | Yes | Ref |  | Ref |  |
|  | No | 1.01 (0.96-1.06) |  | 0.99 (0.93-1.06) |  |
|  | **Poor** |  |  |  |  |
|  | No | Ref |  | Ref |  |
|  | Yes | 1.09 (0.68-1.74) |  | 0.97 (0.56-1.66) |  |
|  | **Middle income** |  |  |  |  |
|  | No | Ref |  | Ref |  |
|  | Yes | 0.70 (0.36-1.36) |  | 0.86 (0.48-1.54) |  |
|  | **Household size^c^** |  |  |  |  |
|  | Per each person increase in household | 0.97 (0.88-1.06) |  | 0.96 (0.86-1.07) |  |
|  | **Prior TB treatment episode** |  |  |  |  |
|  | No | Ref |  | Ref |  |
|  | Yes | 6.15 (2.60-14.53)* |  | 4.77 (1.98-11.53)* |  |
|  | **Prior TB and completed prior treatment** |  |  |  |  |
|  | No | Ref |  | Ref |  |
|  | Yes | 0.23 (0.09-0.60)* |  | 0.22 (0.88-0.59) |  |
|  | **Total weeks from symptom onset to treatment initiation^c^** |  |  |  |  |
|  | Per each week increase in symptoms | 1.00 (0.95-1.05) |  | 1.02 (0.97-1.07) |  |
|  | **Number of symptoms at treatment initiation** |  |  |  |  |
|  | >=5 | Ref |  | Ref |  |
|  | 2 or fewer | 1.18 (0.75-1.85) |  | 1.62 (0.96-2.73) |  |
|  | 3 to 4 | 1.38 (0.74-2.56) |  | 1.54 (0.83-2.86) |  |
|  | **Spent money on travel costs to reach care** |  |  |  |  |
|  | No | Ref |  | Ref |  |
|  | Yes | 2.70 (1.07-6.82)* |  | 2.55 (1.03-6.33)* |  |
|  | **Number of providers visited^d^** |  |  |  |  |
|  | Per each increase in number of providers visited | 4.68 (2.64-8.27)* |  | 3.67 (1.94-6.95)* |  |
|  | **Spending on treatment or medication fees** |  |  |  |  |
|  | No | Ref |  | Ref |  |
|  | Yes | 6.83 (2.18-21.41)* |  | 4.6 (1.38-15.40)* |  |
| Bagchi, 2010^a^ (Maharashtra) *Population: New and previously treated sputum smear positive pulmonary TB patients, excluding new patients in the first two months of therapy Outcome: Medication non-adherence as a single outcome (i.e., at least one week's worth of missed TB medication doses in any treatment month)* |  | Values below are odds ratios |  | Values below are adjusted odds ratios |  |
|  | **Sex** |  |  |  |  |
|  | Female | Ref |  |  |  |
|  | Male | 1.9 (1.0-3.6)* | Not reported |  |  |
|  | **Household members** |  |  |  |  |
|  | 0-3 | Ref |  |  |  |
|  | >3 | 0.7 (0.3-1.3) | Not reported |  |  |
|  | **Any history of smoking** |  |  |  |  |
|  | No | Ref |  | Ref |  |
|  | Yes | 2.4 (1.2-5.1) | Not reported | 1.9 (0.8-4.5) | Not reported |
|  | **Any history of tobacco-chewing** |  |  |  |  |
|  | No | Ref |  |  |  |
|  | Yes | 1.3 (0.7-2.3) | Not reported |  |  |
|  | **Any history of alcohol use** |  |  |  |  |
|  | No | Ref |  | Ref |  |
|  | Yes | 4.8 (2.2-10.3)* | Not reported | 3.6 (1.5-8.3)* | Not reported |
|  | **Hid disease from family members** |  |  |  |  |
|  | No | Ref |  | Ref |  |
|  | Yes | 0.5 (0.2-1.7) | Not reported | 1.9 (0.8-4.2) | Not reported |
|  | **Treatment duration perceived as being too long** |  |  |  |  |
|  | No | Ref |  |  |  |
|  | Yes | 1.7 (1.0-2.9)* | Not reported |  |  |
|  | **Treatment discontinued once symptoms resolved** |  |  |  |  |
|  | No | Ref |  |  |  |
|  | Yes | 1.5 (0.8-3.1) | Not reported |  |  |
|  | **Knows about problems with stopping treatment earlly** |  |  |  |  |
|  | Yes | Ref |  |  |  |
|  | No | 1.5 (0.9-2.6) | Not reported |  |  |
|  | **Feels confident about completing treatment** |  |  |  |  |
|  | No | 1 |  | Ref |  |
|  | Yes | 1.8 (0.5-7.0) | Not reported | 2.5 (0.5-11) |  |
|  | **Travel mode to health center** |  |  |  |  |
|  | Walking | Ref |  |  |  |
|  | Other | 0.6 (0.2-1.2) | Not reported |  |  |
|  | **Travel to health center is a problem** |  |  |  |  |
|  | No | Ref |  |  |  |
|  | Yes | 1.8 (0.8-3.9) | Not reported |  |  |
|  | **Has concerns about transportation to the health center** |  |  |  |  |
|  | No | Ref |  |  |  |
|  | Somewhat or very concerned | 1.1 (0.5-2.4) | Not reported |  |  |
|  | **Concerned about distance to the health center** |  |  |  |  |
|  | No | Ref |  |  |  |
|  | Somewhat or very concerned | 1.3 (0.6-2.4) | Not reported |  |  |
|  | **Concerned about the time to reach the health center** |  |  |  |  |
|  | No | Ref |  |  |  |
|  | Somewhat or very concerned | 1.3 (0.7-2.6) | Not reported |  |  |
|  | **Doctor communicated problems related to stopping medication early** |  |  |  |  |
|  | Yes | Ref |  |  |  |
|  | No | 1.3 (0.8-2.2) | Not reported |  |  |
|  | **Where patient gets most TB information** |  |  |  |  |
|  | DOTS center | Ref |  |  |  |
|  | Other sources | 1.2 (0.6-2.2) | Not reported |  |  |
|  | **Missed treatment due to lack of drug availability** |  |  |  |  |
|  | No | Ref |  | Ref |  |
|  | Yes | 5.6 (1.8-15) | Not reported | 5.1 (1.6-16)* | Not reported |
| Balasubramanian, 2004 (Tamil Nadu) *Population: New and previously treated TB patients (smear positive pulmonary, smear negative pulmonary, and extrapulmonary) as a combined population Outcome: Loss to follow-up as a single outcome* |  | Values below are odds ratios |  | Values below are adjusted odds ratios |  |
|  | **Gender** |  |  |  |  |
|  | Female | Not reported |  | Ref |  |
|  | Male | Not reported |  | 2.1 (1.1-3.9)* | 0.02* |
|  | **Employment** |  |  |  |  |
|  | Unemployed | Not reported |  | Ref |  |
|  | Employed | Not reported |  | 1.7 (1.1-2.4)* | <0.01* |
|  | **History of TB** |  |  |  |  |
|  | No previous history of treatment | Not reported |  | Ref |  |
|  | Previous history of treatment | Not reported |  | 3.9 (2.6-5.6)* | <0.001* |
|  | **Alcohol** |  |  |  |  |
|  | No alcohol use | Not reported |  | Ref |  |
|  | Alcoholism | Not reported |  | 2.2 (1.5-3.3)* | <0.001* |
| Banerjee, 2020 (West Bengal) *Population: New and previously treated TB patients (pulmonary and extrapulmonary) as a combined population Outcome: Death, treatment failure, and loss to follow-up as a composite outcome* |  | Values below are odds ratios |  | Values below are adjusted odds ratios |  |
|  | **Sex** |  |  |  |  |
|  | Female | Ref |  | Ref |  |
|  | Male | 2.6 (1.1-6.2)* |  | 3.07 (1.11-8.52)* |  |
|  | **Type of family** |  |  |  |  |
|  | Nuclear | Ref |  | Ref |  |
|  | Joint | 2.8 (1.2-6.7)* |  | 4.71 (1.66-13.39)* |  |
|  | **Educational status** |  |  |  |  |
|  | Literate | Ref |  | Ref |  |
|  | Illiterate | 2.78 (1.14-6.78)* |  | 1.19 (0.36-3.9) |  |
|  | **Type of disease** |  |  |  |  |
|  | Extrapulmonary | Ref |  | Ref |  |
|  | Pulmonary | 3.9 (1.4-11.0)* |  | 1.9 (0.52-6.9) |  |
|  | **Category of disease** |  |  |  |  |
|  | New | Ref |  | Ref |  |
|  | Previously treated | 3.6 (1.5-8.5)* |  | 3.39 (1.23-9.34)* |  |
|  | **Smoking tobacco use** |  |  |  |  |
|  | Never smoker | Ref |  | Ref |  |
|  | Ever smoker | 2.5 (1.2-5.8)* |  | 1.5 (0.3-6.9) |  |
|  | **Perceived discrimination from family, neighborhood residents, or workplace colleagues** |  |  |  |  |
|  | No | Ref |  | Ref |  |
|  | Yes | 2.26 (1.09-5.65)* |  | 2.61 (1.04-7.84)* |  |
| Bhagyalaxmi, 2010^a^ (Gujarat) *Population: New and previously treated TB patients (pulmonary and extrapulmonary) as a combined population Outcome:* *Death, treatment failure, and loss to follow-up as a composite outcome* |  | Values below are odds ratios |  |  |  |
|  | **Type of DOTS supporter** |  |  |  |  |
|  | TB health visitor | Ref |  |  |  |
|  | Non-TB health visitors (e.g., Anganwadi workers, community volunteers, private practitioners) | 0.46 (0.22-0.98)* | 0.045* |  |  |
| Brahmapurkar^a^, 2017 (Chhattisgarh) *Population: New and previously treated TB patients (sputum smear positive pulmonary, sputum smear negative pulmonary, and extrapulmonary) as a combined population Outcome: Death, treatment failure, loss to follow-up, and transferred as a composite outcome* |  | Values below are odds ratios |  |  |  |
|  | **Age** |  |  |  |  |
|  | <40 | Ref |  |  |  |
|  | >40 | 2.4 (1.49-3.86)* | 0.0003* |  |  |
|  | **Sex** |  |  |  |  |
|  | Female | Ref |  |  |  |
|  | Male | 1.53 (0.91-2.58) | 0.11 |  |  |
|  | **Category** |  |  |  |  |
|  | New | Ref |  |  |  |
|  | Previously treated | 2.54 (1.52-4.23)* | 0.0003* |  |  |
|  | **TB case** |  |  |  |  |
|  | New smear positive | Ref |  |  |  |
|  | New smear negative | 0.72 (0.40-1.29) | 0.27 |  |  |
|  | Previously treated | 2.11 (1.16-3.83)* | 0.01* |  |  |
| Cox, 2021 (2 Indian states) *Population: Men with drug-susceptible or presumed drug-susceptible pulmonary TB Outcome: Treatment failure as a single outcome* |  | Values below are odds ratios |  | Values below are adjusted odds ratios |  |
|  | **Ever had a drink of alcohol** |  |  |  |  |
|  | No | Ref |  | Ref |  |
|  | Yes | 2.06 (1.30-3.28)* | 0.002* | 1.60 (0.95-2.68) | 0.08 |
|  | **Unhealthy alcohol use (AUDIT-C >= 4)** |  |  |  |  |
|  | No | Ref |  | Ref |  |
|  | Yes | 1.92 (1.24-2.96)* | 0.003* | 1.36 (0.83-2.22) | 0.25 |
|  | **AUDIT-C continuous^c^** |  |  |  |  |
|  | No | Ref |  | Ref |  |
|  | Yes | 1.09 (1.03-1.16)* | 0.002* | 1.04 (0.98-1.11) | 0.18 |
|  | **Underweight (BMI<18.5) and severe alcohol use (AUDIT-C > 4)** |  |  |  |  |
|  | Not underweight and not severe alcohol use | Not reported |  | Ref |  |
|  | Not underweight but with severe alcohol use | Not reported |  | 0.62 (0.26-1.50) | 0.29 |
|  | Underweight but not severe alcohol use | Not reported |  | 0.77 (0.39-1.50) | 0.44 |
|  | Underweight and severe alcohol use | Not reported |  | 1.68 (0.92-3.07) | 0.09 |
|  | **Severely underweight (BMI<16.5) and severe alcohol use (AUDIT-C > 4)** |  |  |  |  |
|  | Not severely underweight and not severe alcohol use | Not reported |  | Ref |  |
|  | Not severely underweight but with severe alcohol use | Not reported |  | 0.99 (0.54-1.83) | 0.98 |
|  | Severely Underweight but not severe alcohol use | Not reported |  | 0.67 (0.32-1.41) | 0.29 |
|  | Severely Underweight and severe alcohol use | Not reported |  | 1.76 (0.96-3.25) | 0.07 |
| Dandona, 2004 (4 Indian States) *Population: New and previously treated pulmonary TB patients as a combined population  Outcome: Loss to follow-up as a single outcome* |  |  |  | Values below are adjusted odds ratios |  |
|  | **Age (years)** |  |  |  |  |
|  | 16-30 |  |  | Ref |  |
|  | 31-50 |  |  | 1.58 (0.92-1.46) |  |
|  | >50 |  |  | 1.1 (0.85-1.43) |  |
|  | **Sex** |  |  |  |  |
|  | Female |  |  | Ref |  |
|  | Male |  |  | 2.15 (1.76-2.62)* |  |
|  | **Marital status** |  |  |  |  |
|  | Never married |  |  | Ref |  |
|  | Ever married |  |  | 1.69 (1.21-2.37)* |  |
|  | **Literacy** |  |  |  |  |
|  | Literate |  |  | Ref |  |
|  | Illiterate |  |  | 1.17 (0.97-1.42) |  |
|  | **Monthly family income** |  |  |  |  |
|  | >5000 |  |  | Ref |  |
|  | 3001-5000 |  |  | 1.24 (0.75-2.06) |  |
|  | <=3000 |  |  | 1.3 (0.86-1.96) |  |
|  | **Satisfaction with behavior of DOTS provider** |  |  |  |  |
|  | Satisfied |  |  | Ref |  |
|  | Neither satisfied nor dissatisfied |  |  | 3.51 (2.21-5.57)* |  |
|  | Dissatisfied |  |  | 8.68 (4.41-17.09)* |  |
|  | Refused to answer |  |  | 9.03 (3.33-24.52)* |  |
|  | **Distance to DOTS provider from home** |  |  |  |  |
|  | <=10 km |  |  | Ref |  |
|  | >10 km |  |  | 1.15 (0.79-1.67) |  |
|  | **Type of DOTS provider** |  |  |  |  |
|  | Health facility staff |  |  | Ref |  |
|  | Anganwadi worker |  |  | 0.69 (0.56-0.86)* |  |
|  | Community volunteer |  |  | 0.83 (0.58-1.2) |  |
|  | Family member |  |  | 0.63 (0.28-1.39) |  |
|  | Medicine with self |  |  | 0.26 (0.15-0.47)* |  |
|  | Refused to answer |  |  | 0.17 (0.05-0.59)* |  |
|  | **Patient informed that TB is curable** |  |  |  |  |
|  | Yes |  |  | Ref |  |
|  | No |  |  | 1.75 (1.11-2.75)* |  |
|  | Doesn't remember |  |  | 2.29 (1.2-4.26)* |  |
|  | **Patient informed of treatment duration** |  |  |  |  |
|  | Yes |  |  | Ref |  |
|  | No |  |  | 3.11 (2.08-4.66)* |  |
|  | Doesn't remember |  |  | 2.59 (1.66-4.02)* |  |
| Das, 2014^a^ (Nagaland) *Population: New and previously treated TB patients (smear positive pulmonary, smear negative pulmonary, and extrapulmonary) as a combined population Outcome: Death, treatment failure, and loss to follow-up as a composite outcome* |  | Values below are odds ratios |  | Values below are adjusted odds ratios |  |
|  | **Age (years)^c^** |  |  |  |  |
|  | Per each year increase in age | Not reported | Not reported | 1.03 (1.01-1.05)* |  |
|  | **Sex** |  |  |  |  |
|  | Female | Ref |  | Ref |  |
|  | Male | 2.08 (1.13-3.81) | 0.02* | 1.5 (0.78-2.86) |  |
|  | **Residence** |  |  |  |  |
|  | Rural | Ref |  |  |  |
|  | Semi-urban | 1.24 (0.68-2.26) | 0.48 |  |  |
|  | **TB site** |  |  |  |  |
|  | Pulmonary | Ref |  |  |  |
|  | Extrapulmonary | 1.85 (0.82-4.16) | 0.14 |  |  |
|  | **TB treatment regimen** |  |  |  |  |
|  | Category I | Ref |  | Ref |  |
|  | Category II | 2.37 (1.28-4.38)* | 0.006* | 1.81 (0.95-3.47) |  |
| Gopi, 2007 (Tamil Nadu) *Population: New and previously treated pulmonary TB patients (sputum smear positive and sputum smear negative) during the intensive phase of therapy as a combined population  Outcome: Treatment non-adherence as a single outcome* |  | Values below are odds ratios |  | Values below are adjusted odds ratios |  |
|  | **Sex** |  |  |  |  |
|  | Female | Ref |  |  |  |
|  | Male | 1.13 (0.9-1.43) | Not reported |  |  |
|  | **Age (years)** |  |  |  |  |
|  | <=45 | Ref |  |  |  |
|  | >45 | 1.19 (0.96-1.47) | Not reported |  |  |
|  | **Education** |  |  |  |  |
|  | Literate | Ref |  | Ref |  |
|  | Illiterate | 1.47 (1.19-1.83)* | <0.01* | 1.33 (1.07-1.66)* | <0.05* |
|  | **Occupation** |  |  |  |  |
|  | Employed | Ref |  |  |  |
|  | Unemployed | 1.02 (0.82-1.27) | Not reported |  |  |
|  | **Case type** |  |  |  |  |
|  | Previously treated pulmonary TB patients | Ref |  | 2.29 (1.51-3.47) | <0.001* |
|  | New sputum smear positive pulmonary TB patients | 1.86 (1.27-2.75)* | <0.01* | 1.31 (1.05-1.64)* | <0.05* |
|  | New sputum smear-negative pulmonary TB patients | 2.43 (1.66-3.56)* | <0.01* | Ref |  |
|  | **Loss of wages due to directly observed therapy** |  |  |  |  |
|  | Yes | Ref |  |  |  |
|  | No | 1.28 (0.78-2.12) | Not reported |  |  |
|  | **Problem in taking drugs** |  |  |  |  |
|  | No | Ref |  |  |  |
|  | Yes | 1.07 (0.87-1.32) |  |  |  |
|  | **Directly observed therapy interferes with daily activities** |  |  |  |  |
|  | No | Ref |  |  |  |
|  | Yes | 1.49 (1.03-2.16)* | <0.05* |  |  |
|  | **Smoking** |  |  |  |  |
|  | No | Ref |  |  |  |
|  | Yes | 1.16 (0.94-1.43) | Not reported |  |  |
|  | **Alcoholism** |  |  |  |  |
|  | Yes | Ref |  |  |  |
|  | No | 1.06 (0.84-1.33) | Not reported |  |  |
|  | **Difficulty accessing the health facility** |  |  |  |  |
|  | No | Ref |  | Ref |  |
|  | Yes | 2.96 (2.06-4.24)* | <0.01* | 3.02 (2.10-4.34)* | <0.001* |
|  | **Need escort to get to the directly observed therapy center** |  |  |  |  |
|  | Yes | Ref |  |  |  |
|  | No | 1.09 (0.69-1.73) | Not reported |  |  |
|  | **Directly observed therapy center type** |  |  |  |  |
|  | Government | Ref |  | Ref |  |
|  | Non-government | 2.14 (1.73-2.65)* | <0.01* | 2.11 (1.70-2.61)* | <0.001* |
| Huddart, 2021 (Bihar) *Population: New and previously treated TB patients as a combined population Outcome: Death as a single outcome* |  | Values below are unweighted model adjusted hazard ratios^d^ |  | Values below are weighted model adjusted hazard ratios^e^ |  |
|  | **Gender** |  |  |  |  |
|  | Male | Ref |  | Ref |  |
|  | Female | 0.75 (0.54-1.02) |  | 0.71 (0.47-1.05) |  |
|  | **Age** |  |  |  |  |
|  | Per each year increase in age | 1.03 (1.02-1.03)* |  | 1.03 (1.02-1.04)* |  |
|  | **Residence** |  |  |  |  |
|  | Out of Patna | Ref |  | Ref |  |
|  | Rural Patna | 0.90 (0.57-1.33) |  | 0.99 (0.58-1.56) |  |
|  | Urban Patna | 0.72 (0.51-1.08) |  | 0.79 (0.53-1.15) |  |
|  | **Slum residence** |  |  |  |  |
|  | Non-slum | Ref |  | Ref |  |
|  | Slum | 0.81 (0.60-1.07) |  | 0.72 (0.51-1.00) |  |
|  | **Treatment category** |  |  |  |  |
|  | New | Ref |  | Ref |  |
|  | Retreatment | 1.37 (0.86-2.14) |  | 1.34 (0.74-2.26) |  |
|  | **Type of TB** |  |  |  |  |
|  | Pulmonary | Ref |  | Ref |  |
|  | Extrapulmonary | 0.95 (0.68-1.31) |  | 0.84 (0.57-1.19) |  |
| Jaggarajamma, 2007^a^ (Tamil Nadu) *Population: New and previously treated TB patients (sputum smear positive pulmonary, sputum smear negative pulmonary, and extrapulmonary) as a combined population Outcome: Loss to follow-up as a single outcome* |  | Values below are odds ratios |  |  |  |
|  | **Sex** |  |  |  |  |
|  | Female | Ref |  |  |  |
|  | Male | 3.5 (2.12-5.77)* | <0.0001* |  |  |
|  | **Age (years)** |  |  |  |  |
|  | <45 | Ref |  |  |  |
|  | >=45 | 1.31 (0.95-1.81) | 0.1 |  |  |
|  | **Education** |  |  |  |  |
|  | Literate | Ref |  |  |  |
|  | Illiterate | 1.19 (0.83-1.72) | 0.34 |  |  |
|  | **Occupation** |  |  |  |  |
|  | Employed | Ref |  |  |  |
|  | Unemployed | 0.73 (0.49-1.1) | 0.13 |  |  |
|  | **Category of treatment** |  |  |  |  |
|  | Category I (new sputum smear-positive pulmonary TB patients) | Ref |  |  |  |
|  | Category II (previously treated sputum smear-positive pulmonary TB patients) | 2.68 (1.81-3.95)* | <0.0001* |  |  |
|  | Category III (new extrapulmonary and sputum smear-negative pulmonary TB patients) | 0.54 (0.36-0.83)* | 0.004* |  |  |
|  | **Sputum smear status** |  |  |  |  |
|  | Sputum smear-negative pulmonary TB patients | Ref |  |  |  |
|  | Sputum smear-positive pulmonary TB patients | 2.06 (1.46-2.92)* | <0.0001* |  |  |
|  | **Type of disease** |  |  |  |  |
|  | Extrapulmonary | Ref |  |  |  |
|  | Pulmonary | 2.46 (1.11-5.46)* | 0.03* |  |  |
|  | **Alcoholism** |  |  |  |  |
|  | No | Ref |  |  |  |
|  | Yes | 2.63 (1.83-3.77)* | <0.0001* |  |  |
|  | **DOT reported as being convenient by the patient** |  |  |  |  |
|  | Yes | Ref |  |  |  |
|  | No | 2.16 (1.28-3.62)* | 0.004* |  |  |
| Jan Swasthya Sahyog, 2018^a^ (Chhattisgarh) *Population: New and previously treated TB patients (sputum smear positive pulmonary, sputum smear negative pulmonary, and extrapulmonary) as a combined population Outcome:* *Death, treatment failure, loss to follow-up, and transferred out as a composite outcome* |  | Values below are odds ratios |  |  |  |
|  | **Pretreatment sputum status (among patients with sputum samples)** |  |  |  |  |
|  | Positive | Ref |  |  |  |
|  | Negative | 1.04 (0.87-1.23) | 0.69 |  |  |
|  | **AFB Grade (among sputum positive patients)** |  |  |  |  |
|  | 3+ | Ref |  |  |  |
|  | 1+ | 0.89 (0.70-1.13) | 0.35 |  |  |
|  | 2+ | 0.80 (0.63-1.01) | 0.06 |  |  |
| Jan Swasthya Sahyog, 2018 (Chhattisgarh) *Outcome:* *Death, treatment failure, loss to follow-up, and transferred out as a composite outcome* |  | Values below are relative risk ratios |  |  |  |
|  | **Age (years)** |  |  |  |  |
|  | <50 | Ref |  |  |  |
|  | >=50 | 1.71 (1.45-2.02)* | <0.01* |  |  |
|  | **Gender (by treatment site)** |  |  |  |  |
|  | Male, secondary care hospital | Ref |  |  |  |
|  | Male, primary care clinic | 0.56 (0.41-0.74)* | <0.01* |  |  |
|  | Female, secondary care hospital | 0.77 (0.66-0.90)* | <0.01* |  |  |
|  | Female, primary care clinic | 0.59 (0.42-0.84)* | <0.01* |  |  |
|  | **Treatment history** |  |  |  |  |
|  | No prior TB treatment | Ref |  |  |  |
|  | Any prior TB treatment | 1.28 (1.00-1.63)* | 0.05* |  |  |
|  | **Season and distance from treatment** |  |  |  |  |
|  | Winter and <45 minutes | Ref |  |  |  |
|  | Winter and 45 minutes-1.5 hours | 1.01 (0.75-1.37) | 0.93 |  |  |
|  | Winter and 1.5-4 hours | 1.31 (0.96-1.78) | 0.08 |  |  |
|  | Winter and >4 hours | 1.13 (0.83-1.55) | 0.43 |  |  |
|  | Summer and <45 minutes | 1.33 (1.00-1.77)* | 0.05* |  |  |
|  | Summer and 45 minutes-1.5 hours | 1.48 (1.08-2.05)* | 0.02* |  |  |
|  | Summer and 1.5-4 hours | 1.77 (1.24-2.52)* | <0.01* |  |  |
|  | Summer and >4 hours | 2.49 (1.79-3.47)* | <0.01* |  |  |
|  | Monsoon and <45 minutes | 1.21 (0.91-1.61) | 0.19 |  |  |
|  | Monsoon and 45 minutes-1.5 hours | 1.02 (0.75-1.40) | 0.86 |  |  |
|  | Monsoon and 1.5-4 hours | 1.30 (0.93-1.81) | 0.13 |  |  |
|  | Monsoon and >4 hours | 1.35 (0.99-1.85) | 0.06 |  |  |
| Jonnalagada, 2011 (Andhra Pradesh) *Population: New and previously treated TB patients (sputum smear positive pulmonary, sputum smear negative pulmonary, and extrapulmonary) as a combined population Outcome:* *Death, treatment failure, loss to follow-up, and transferred out as a composite outcome* |  | Values below are odds ratios |  |  |  |
|  | **Type and category of TB** |  |  |  |  |
|  | Extrapulmonary TB patients | Ref |  |  |  |
|  | New sputum smear negative pulmonary TB patients | 1.34 (1.06-1.68)* | 0.01* |  |  |
|  | New sputum smear positive pulmonary TB patients | 1.64 (1.35-2.00)* | <0.0001* |  |  |
|  | Previously treated TB patients | 3.14 (2.68-3.69)* | <0.0001* |  |  |
| Joseph, 2011^a^ (Karnataka) *Population: New and previously treated sputum smear positive pulmonary TB patients as a combined population Outcome: Treatment failure and loss to follow-up as a composite outcome* |  | Values below are odds ratios |  |  |  |
|  | **Sex** |  |  |  |  |
|  | Female | Ref |  |  |  |
|  | Male | 1.33 (0.71-2.52) | 0.37 |  |  |
|  | **Location** |  |  |  |  |
|  | Rural | Ref |  |  |  |
|  | Urban | 2.04 (1.25-3.31)* | 0.004* |  |  |
|  | **Treatment category** |  |  |  |  |
|  | Category I (new sputum smear positive pulmonary TB patients) | Ref |  |  |  |
|  | Category II (previously treated sputum smear positive pulmonary TB patients) | 3.81 (2.18-6.65)* | <0.0001* |  |  |
| Karanjekar, 2014^a^ (Maharashtra) *Population: New and previously treated TB patients (sputum smear positive pulmonary, sputum smear negative pulmonary, and extrapulmonary) as a combined population Outcome: Death, loss to follow-up, and transferred out as a composite outcome* |  | Values below are odds ratios |  |  |  |
|  | **Age (years)** |  |  |  |  |
|  | 15-24 | Ref |  |  |  |
|  | 25-34 | 0.92 (0.25-3.36) | 0.89 |  |  |
|  | 35-44 | 1.01 (0.30-3.90) | 0.9 |  |  |
|  | 45-54 | 1.46 (0.37-5.80) | 0.59 |  |  |
|  | 54+ | 1.42 (0.33-6.00) | 0.64 |  |  |
|  | **Sex** |  |  |  |  |
|  | Female | Ref |  |  |  |
|  | Male | 2.54 (0.99-6.48) | 0.05 |  |  |
|  | **Education** |  |  |  |  |
|  | More than high school | Ref |  |  |  |
|  | Up to high school | 0.91 (0.26-3.26) | 0.89 |  |  |
|  | Illiterate | 0.63 (0.16-2.52) | 0.51 |  |  |
|  | **Treatment category** |  |  |  |  |
|  | Category I (new sputum smear-positive pulmonary TB patients) | Ref |  |  |  |
|  | Category II (previously treated sputum smear-positive pulmonary TB patients) | 1.95 (0.64-5.88) | 0.24 |  |  |
|  | Category III (new extrapulmonary and sputum smear-negative pulmonary TB patients) | 1.17 (0.47-2.92) | 0.74 |  |  |
|  | **Type of TB** |  |  |  |  |
|  | Pulmonary | Ref |  |  |  |
|  | Extrapulmonary | 3.86 (1.37-10.89)* | 0.01* |  |  |
| Kumar, 2018 (Madhya Pradesh) *Population: New and previously treated TB patients (sputum smear positive pulmonary, sputum smear negative pulmonary, and extrapulmonary) as a combined population Outcome: Treatment failure, LTFU, transferred out and death as a composite outcome* |  | Values below are odds ratios |  |  |  |
|  | **Sex** |  |  |  |  |
|  | Female | Ref |  |  |  |
|  | Male | 3.86 (2.09-7.12)* | <0.0001* |  |  |
|  | **Age (years)** |  |  |  |  |
|  | 15 to 25 | Ref |  |  |  |
|  | 0 to 14 | 0.11 (0.01-0.86) | 0.04 |  |  |
|  | 26 to 35 | 1.12 (0.55-2.29) | 0.75 |  |  |
|  | 36 to 45 | 1.04 (0.44-2.42) | 0.94 |  |  |
|  | >45 | 3.05 (1.61-5.78)* | 0.0006* |  |  |
|  | **Category of TB** |  |  |  |  |
|  | Category I (new TB patients) | Ref |  |  |  |
|  | Category II (previously treated TB patients) | 2.35 (1.36-4.07)* | 0.002* |  |  |
|  | **Site involvement** |  |  |  |  |
|  | Extrapulmonary TB patients | Ref |  |  |  |
|  | Pulmonary TB patients | 3.66 (1.77 to 7.58)* | 0.0005* |  |  |
| Lata, 2021^a^ (Jammu and Kashmir) *Population: New and previously treated TB patients who completed at least 2 months of TB therapy as a combined population Outcome: Medication non-adherence as a single outcome (i.e., self-reported non-ingestion of at least one medication dose as measured by the Morisky adherence scale)* |  | Values below are odds ratios |  |  |  |
|  | **Gender** |  |  |  |  |
|  | Female | Ref |  |  |  |
|  | Male | 1.25 (0.24-6.47) | 0.79 |  |  |
| Mittal, 2011^a^ (Uttar Pradesh) *Population: New and previously treated TB patients (sputum smear positive pulmonary, sputum smear negative pulmonary, and extrapulmonary) as a combined population Outcome: Death,treatment failure, loss to follow-up, and transferred out as a composite outcome* |  | Values below are odds ratios |  |  |  |
|  | **Treatment category** |  |  |  |  |
|  | Category III (new extrapulmonary and sputum smear-negative pulmonary TB patients) | Ref |  |  |  |
|  | Category I (new sputum smear-positive pulmonary TB patients) | 2.23 (1.49-3.34)* | 0.0001* |  |  |
|  | Category II (previously treated sputum smear-positive pulmonary TB patients) | 5.26 (3.45-8.02)* | <0.0001* |  |  |
|  | **Type of disease** |  |  |  |  |
|  | Extrapulmonary | Ref |  |  |  |
|  | Pulmonary | 2.44 (1.67-3.56)* | <0.0001* |  |  |
|  | **Type of patient based on outcome of previous treatment** |  |  |  |  |
|  | New/transfer-in (i.e., no prior TB history) | Ref |  |  |  |
|  | Treatment after loss to follow-up | 3.48 (2.28-5.33)* | <0.0001* |  |  |
|  | Treatment failure during prior treatment | 2.94 (1.60-5.39)* | 0.0005* |  |  |
|  | Relapse (i.e., previous treatment was completed) | 24.67 (2.94-206.79)* | 0.003* |  |  |
|  | Others | 5.16 (3.31-8.06)* | <0.0001* |  |  |
|  | **Pre-treatment sputum smear status** |  |  |  |  |
|  | Not done | Ref |  |  |  |
|  | Positive smear by sputum microscopy | 2.57 (1.75-3.79)* | <0.0001* |  |  |
|  | Negative smear by sputum microscopy | 2.07 (1.36-3.15)* | 0.0007* |  |  |
| Mittal, 2011^a^ (Uttar Pradesh) *Population: New and previously treated TB patients (sputum smear positive pulmonary, sputum smear negative pulmonary, and extrapulmonary) as a combined population Outcome: Treatment LTFU as a single outcome* |  | Values below are odds ratios |  |  |  |
|  | **Age (years)** |  |  |  |  |
|  | <15 | Ref |  |  |  |
|  | 16-30 | 5.81 (2.46-13.70)* | 0.0001* |  |  |
|  | 31-45 | 5.20 (2.15-12.60)* | 0.0003* |  |  |
|  | >45 | 8.11 (3.24-20.28)* | <0.0001* |  |  |
|  | **Sex** |  |  |  |  |
|  | Female | Ref |  |  |  |
|  | Male | 2.19 (1.44-3.33)* | 0.0002* |  |  |
|  | **Religion** |  |  |  |  |
|  | Hindu | Ref |  |  |  |
|  | Muslim | 0.69 (0.41-1.15) | 0.16 |  |  |
|  | **Occupation** |  |  |  |  |
|  | Laborer | Ref |  |  |  |
|  | Service | 0.90 (0.33-2.41) | 0.83 |  |  |
|  | Business | 1.97 (0.93-4.17) | 1.78 |  |  |
|  | Housewife | 0.51 (0.30-0.86) | 2.51 |  |  |
|  | Unemployed/retired | 1.49 (0.65-3.45) | 0.35 |  |  |
|  | Student | 0.37 (0.19-0.74)* | 0.005* |  |  |
|  | Not defined (child under age of 5) | 0.14 (0.02-1.04) | 0.05 |  |  |
|  | **Type of disease** |  |  |  |  |
|  | Extrapulmonary | Ref |  |  |  |
|  | Pulmonary | 3.28 (1.88-5.74)* | <0.0001* |  |  |
|  | **Type of patients** |  |  |  |  |
|  | New/transfer-in | Ref |  |  |  |
|  | Treatment after default | 4.14 (2.51-6.81)* | <0.0001* |  |  |
|  | Failure | 2.40 (1.11-5.21)* | 0.03* |  |  |
|  | Relapse | 7.81 (1.70-35.77)* | 0.008* |  |  |
|  | Others | 6.42 (3.92-10.51)* | <0.0001* |  |  |
|  | **Treatment category** |  |  |  |  |
|  | Category I (new sputum smear-positive pulmonary TB patients) | Ref |  |  |  |
|  | Category II (previously treated sputum smear-positive pulmonary TB patients) | 2.79 (1.82-4.27)* | <0.0001* |  |  |
|  | Category III (new extrapulmonary and sputum smear-negative pulmonary TB patients) | 0.94 (0.57-1.53) | 0.79 |  |  |
|  | **Pre-treatment sputum status** |  |  |  |  |
|  | Positive | Ref |  |  |  |
|  | Negative | 0.93 (0.62-1.39) | 0.72 |  |  |
| Mukhopadhyay, 2011^a^ (West Bengal) *Population: New and previously treated TB patients (sputum smear positive pulmonary, sputum smear negative pulmonary, and extrapulmonary) as a combined population Outcome:* *Death, treatment failure, loss to follow-up, and transferred out as a composite outcome* |  | Values below are odds ratios |  |  |  |
|  | **Age (years)** |  |  |  |  |
|  | <=19 | Ref |  |  |  |
|  | 20-60 | 4.15 (1.49-11.55)* | 0.007* |  |  |
|  | >60 | 7.38 (2.25-24.15)* | 0.001* |  |  |
|  | **Gender** |  |  |  |  |
|  | Female | Ref |  |  |  |
|  | Male | 2.48 (1.47-4.20)* | 0.0007* |  |  |
|  | **Residence** |  |  |  |  |
|  | Rural | Ref |  |  |  |
|  | Urban | 1.17 (0.78-1.75) | 0.45 |  |  |
|  | **TB classification** |  |  |  |  |
|  | Extrapulmonary | Ref |  |  |  |
|  | New sputum negative | 1.35 (0.49-3.69) | 0.56 |  |  |
|  | **Previous treatment history** |  |  |  |  |
|  | Other (i.e., smear negative or extrapulmonary) | Ref |  |  |  |
|  | Relapse (prior treatment completion) | 2.16 (0.77-6.08) | 0.14 |  |  |
|  | Prior treatment failure | 5.17 (1.24-21.59)* | 0.02* |  |  |
|  | Treatment after loss to follow-up | 3.95 (1.27-12.28)* | 0.02* |  |  |
| Mundra, 2017 (Maharashtra) *Population: New and previously treated TB patients (sputum smear positive pulmonary, sputum smear negative pulmonary, and extrapulmonary) as a combined population Outcome: Death, treatment failure, treatment modification, and loss to follow-up as a composite outcome* |  |  |  | Values below are adjusted hazard ratios |  |
|  | **Age** |  |  |  |  |
|  | 0-29 | Not reported |  | Ref |  |
|  | 30-44 | Not reported |  | 1.55 (0.82-2.90) |  |
|  | 45-59 | Not reported |  | 2.99 (1.58-5.68)* |  |
|  | 60 and older | Not reported |  | 2.43 (1.28-4.61)* |  |
|  | **Gender** |  |  |  |  |
|  | Female | Not reported |  | Ref |  |
|  | Male | Not reported |  | 1.35 (0.82-2.24) |  |
|  | **Residence** |  |  |  |  |
|  | Rural | Not reported |  | Ref |  |
|  | Urban | Not reported |  | 0.99 (0.64-1.54) |  |
|  | **Pulmonary sputum** |  |  |  |  |
|  | Positive | Not reported |  | Ref |  |
|  | Negative | Not reported |  | 0.91 (0.56-1.47) |  |
|  | Extrapulmonary | Not reported |  | 0.33 (0.15-0.75)* |  |
|  | **Category of TB** |  |  |  |  |
|  | Category I | Not reported |  | Ref |  |
|  | Category II | Not reported |  | 1.46 (0.91-2.33) |  |
|  | **HIV** |  |  |  |  |
|  | Negative | Not reported |  | Ref |  |
|  | Positive | Not reported |  | 0.49 (0.15-1.58) |  |
|  | Unknown | Not reported |  | 0.97 (0.39-2.43) |  |
|  | **Diabetic status** |  |  |  |  |
|  | Non-diabetic | Not reported |  | Ref |  |
|  | Diabetic | Not reported |  | 0.35 (0.05-2.56) |  |
|  | Unknown | Not reported |  | 1.06 (0.54-1.89) |  |
| Mundra, 2017 (Maharashtra) *Population: New and previously treated TB patients (sputum smear positive pulmonary, sputum smear negative pulmonary, and extrapulmonary) as a combined population Outcome:* *Treatment failure as a single outcome* |  |  |  | Values below are adjusted hazard ratios |  |
|  | **Age** |  |  |  |  |
|  | 0-29 | Not reported |  | Ref |  |
|  | 30-44 | Not reported |  | 1.59 (0.25-10.05) |  |
|  | 45-59 | Not reported |  | 3.18 (0.35-29.12) |  |
|  | 60 and older | Not reported |  | 2.98 (0.41-21.82) |  |
|  | **Gender** |  |  |  |  |
|  | Female | Not reported |  | Ref |  |
|  | Male | Not reported |  | 1.03 (0.17-6.34) |  |
|  | **Residence** |  |  |  |  |
|  | Rural | Not reported |  | Ref |  |
|  | Urban | Not reported |  | 1.26 (0.31-5.08) |  |
|  | **Pulmonary sputum** |  |  |  |  |
|  | Positive | Not reported |  | Ref |  |
|  | Negative | Not reported |  | 0 |  |
|  | Extrapulmonary | Not reported |  | 0 |  |
|  | **Category of TB** |  |  |  |  |
|  | Category I | Not reported |  | Ref |  |
|  | Category II | Not reported |  | 0.40 (0.07-2.24) |  |
|  | **HIV** |  |  |  |  |
|  | Negative | Not reported |  | Ref |  |
|  | Positive | Not reported |  | 0 |  |
|  | Unknown | Not reported |  | 0 |  |
|  | **Diabetic status** |  |  |  |  |
|  | Non-diabetic | Not reported |  | Ref |  |
|  | Diabetic | Not reported |  | 0 |  |
|  | Unknown | Not reported |  | 4.01 (0.67-23.93) |  |
| Mundra, 2017 (Maharashtra) *Population: New and previously treated TB patients (sputum smear positive pulmonary, sputum smear negative pulmonary, and extrapulmonary) as a combined population Outcome: Loss to follow-up as a single outcome* |  |  |  | Values below are adjusted hazard ratios |  |
|  | **Age** |  |  |  |  |
|  | 0-29 | Not reported |  | Ref |  |
|  | 30-44 | Not reported |  | 2.21 (0.81-6.05) |  |
|  | 45-59 | Not reported |  | 5.18 (1.95-13.79)* |  |
|  | 60 and older | Not reported |  | 3.97 (1.47-10.70)* |  |
|  | **Gender** |  |  |  |  |
|  | Female | Not reported |  | Ref |  |
|  | Male | Not reported |  | 3.03 (1.26-7.31)* |  |
|  | **Residence** |  |  |  |  |
|  | Rural | Not reported |  | Ref |  |
|  | Urban | Not reported |  | 1.08 (0.58-2.03) |  |
|  | **Pulmonary sputum** |  |  |  |  |
|  | Positive | Not reported |  | Ref |  |
|  | Negative | Not reported |  | 1.10 (0.58-2.12) |  |
|  | Extrapulmonary |  |  | 0.44 (0.15-1.33) |  |
|  | **Category of TB** |  |  |  |  |
|  | Category I | Not reported |  | Ref |  |
|  | Category II | Not reported |  | 1.36 (0.70-2.63) |  |
|  | **HIV** |  |  |  |  |
|  | Negative | Not reported |  | Ref |  |
|  | Positive | Not reported |  | 0.60 (0.14-2.55) |  |
|  | Unknown | Not reported |  | 1.78 (0.62-5.10) |  |
|  | **Diabetic status** |  |  |  |  |
|  | Non-diabetic | Not reported |  | Ref |  |
|  | Diabetic | Not reported |  | 0 |  |
|  | Unknown | Not reported |  | 1.01 (0.54-1.89) |  |
| Mundra, 2017 (Maharashtra) *Population: New and previously treated TB patients (sputum smear positive pulmonary, sputum smear negative pulmonary, and extrapulmonary) as a combined population Outcome:* *Treatment modification as a single outcome* |  |  |  | Values below are adjusted hazard ratios |  |
|  | **Age** |  |  |  |  |
|  | 0-29 | Not reported |  | Ref |  |
|  | 30-44 | Not reported |  | 0.11 (0.004-3.34) |  |
|  | 45-59 | Not reported |  | 0 |  |
|  | 60 and older | Not reported |  | 0 |  |
|  | **Gender** |  |  |  |  |
|  | Female | Not reported |  | Ref |  |
|  | Male | Not reported |  | 0.06 (0.002-1.92) |  |
|  | **Residence** |  |  |  |  |
|  | Rural | Not reported |  | Ref |  |
|  | Urban | Not reported |  | 0.62 (0.04-9.54) |  |
|  | **Pulmonary sputum** |  |  |  |  |
|  | Positive | Not reported |  | Ref |  |
|  | Negative | Not reported |  | 0.81 (0.04-15.54) |  |
|  | Extrapulmonary | Not reported |  | 0 |  |
|  | **Category of TB** |  |  |  |  |
|  | Category I | Not reported |  | Ref |  |
|  | Category II | Not reported |  | 23.01 (0.87-609.68) |  |
|  | **HIV** |  |  |  |  |
|  | Negative | Not reported |  | Ref |  |
|  | Positive | Not reported |  | 0 |  |
|  | Unknown | Not reported |  | 0 |  |
|  | **Diabetic status** |  |  |  |  |
|  | Non-diabetic | Not reported |  | Ref |  |
|  | Diabetic | Not reported |  | 0 |  |
|  | Unknown | Not reported |  | 0 |  |
| Mundra, 2017 (Maharashtra) *Population: New and previously treated TB patients (sputum smear positive pulmonary, sputum smear negative pulmonary, and extrapulmonary) as a combined population Outcome: Death as a single outcome* |  |  |  | Values below are adjusted hazard ratios |  |
|  | **Age** |  |  |  |  |
|  | 0-29 | Not reported |  | Ref |  |
|  | 30-44 | Not reported |  | 1.21 (0.44-3.30) |  |
|  | 45-59 | Not reported |  | 1.76 (0.59-5.25) |  |
|  | 60 and older | Not reported |  | 1.74 (0.60-4.99) |  |
|  | **Gender** |  |  |  |  |
|  | Female | Not reported |  | Ref |  |
|  | Male | Not reported |  | 0.74 (0.34-1.60) |  |
|  | **Residence** |  |  |  |  |
|  | Rural | Not reported |  | Ref |  |
|  | Urban | Not reported |  | 0.97 (0.46-2.05) |  |
|  | **Pulmonary sputum** |  |  |  |  |
|  | Positive | Not reported |  | Ref |  |
|  | Negative | Not reported |  | 0.97 (0.43-2.22) |  |
|  | Extrapulmonary | Not reported |  | 0.37 (0.10-1.31) |  |
|  | **Category of TB** |  |  |  |  |
|  | Category I | Not reported |  | Ref |  |
|  | Category II | Not reported |  | 1.63 (0.73-3.66) |  |
|  | **HIV** |  |  |  |  |
|  | Negative | Not reported |  | Ref |  |
|  | Positive | Not reported |  | 0.51 (0.07-3.95) |  |
|  | Unknown | Not reported |  | 0.53 (0.07-3.97) |  |
|  | **Diabetic status** |  |  |  |  |
|  | Non-diabetic | Not reported |  | Ref |  |
|  | Diabetic | Not reported |  | 1.30 (0.16-10.49) |  |
|  | Unknown | Not reported |  | 1.18 (0.53-2.59) |  |
| Mundra, 2018 (Maharashtra) *Population: New and previously treated TB patients (sputum smear positive pulmonary, sputum smear negative pulmonary, and extrapulmonary) as a combined population Outcome: Death, treatment failure, treatment modification, and loss to follow-up as a composite outcome* |  | Values below are odds ratios |  | Values below are adjusted odds ratios |  |
|  | **Age (years)** |  |  |  |  |
|  | 0-29 | Ref | 0.01* |  |  |
|  | 30-44 | 1.63 (0.79-3.37) |  |  |  |
|  | 45-59 | 3.32 (1.57-7.04)* |  |  |  |
|  | >=60 | 2.43 (1.12-5.27)* |  |  |  |
|  | **Sex** |  |  |  |  |
|  | Female | Ref |  |  |  |
|  | Male | 2.16 (1.24-3.77)* | 0.007* |  |  |
|  | **Caste** |  |  |  |  |
|  | General | Ref | 0.91 |  |  |
|  | OBC | 1.17 (0.58-2.35) |  |  |  |
|  | SC/ST/NT | 1.10 (0.56-2.14) |  |  |  |
|  | **Residence** |  |  |  |  |
|  | Urban | Ref |  |  |  |
|  | Rural | 1.24 (0.75-2.07) | 0.4 |  |  |
|  | **Education** |  |  |  |  |
|  | Graduate or above | Ref | 0.01* |  |  |
|  | Less than primary | 4.64 (1.70-12.63)* |  |  |  |
|  | Primary | 4.33 (1.57-11.98)* |  |  |  |
|  | Secondary | 3.58 (1.37-9.36)* |  |  |  |
|  | High School | 2.14 (0.84-5.44) |  |  |  |
|  | **Occupation** |  |  |  |  |
|  | Clerical or professional | Ref | 0.001* |  |  |
|  | Sem-skilled or skilled labor | 3.33 (1.32-8.42)* |  |  |  |
|  | Unskilled labor | 5.00 (1.87-13.36)* |  |  |  |
|  | Unemployed or students | 1.52 (0.60-3.86) |  |  |  |
|  | **Socioeconomic status** |  |  |  |  |
|  | Above the poverty line | Ref |  |  |  |
|  | Below the poverty line | 1.81 (1.07-3.04)* | 0.03* |  |  |
|  | **Type of illness** |  |  |  |  |
|  | New | Ref |  |  |  |
|  | Retreatment | 2.24 (1.29-3.89)* | 0.004* |  |  |
|  | **Site of disease and sputum status** |  |  |  |  |
|  | Extrapulmonary TB patients | Ref | 0.001* | Ref |  |
|  | Pulmonary sputum smear negative TB patients | 6.55 (2.45-17.49)* |  | 3.76 (1.18-11.96)* | 0.03* |
|  | Pulmonary sputum smear positive TB patients | 5.86 (2.11-16.23)* |  | 4.53 (1.36-15.10)* | 0.01* |
|  | **Smear conversion at end of the intensive phase** |  |  |  |  |
|  | Smear converted | Ref |  |  |  |
|  | Smear not converted | 13.33 (5.16-34.42)* | <0.001* |  |  |
|  | **Experienced side effect of medicines** |  |  |  |  |
|  | No | Ref |  |  |  |
|  | Yes | 1.28 (0.64-2.57) | 0.48 |  |  |
|  | **Comorbidities** |  |  |  |  |
|  | Absent | Ref |  |  |  |
|  | Present | 2.90 (1.70-4.93)* | <0.001* |  |  |
|  | **HIV status** |  |  |  |  |
|  | Negative | Ref |  |  |  |
|  | Positive | 0.52 (0.14-1.89) |  |  |  |
|  | Unknown | 1.15 (0.37-3.55) |  |  |  |
|  | **Diabetic status** |  |  |  |  |
|  | Non-diabetic | Ref | 0.42 |  |  |
|  | Diabetic | 1.47 (0.37-5.82) |  |  |  |
|  | Unknown | 0.76 (0.45-1.28) |  |  |  |
|  | **Median delay in days (IQR)^c^** |  |  |  |  |
|  | Per each day increase in the period of delay |  |  |  |  |
|  | In visiting health facility after developing symptoms | 1.00 (0.99-1.01) | 0.71 |  |  |
|  | In diagnosis from initial health facility visit | 0.99 (0.97-1.01) | 0.18 |  |  |
|  | In treatment initiation after diagnosis | 1.02 (0.98-1.07) | 0.25 |  |  |
|  | **Perception of early or late care-seeking** |  |  |  |  |
|  | Early | Ref |  |  |  |
|  | Late | 1.70 (1.01-2.85)* | 0.05* |  |  |
|  | **Ever felt discriminated against due to having TB** |  |  |  |  |
|  | No | Not reported |  | Ref |  |
|  | Yes | Not reported |  | 2.20 (1.08-4.51)* | 0.03* |
|  | **Satisfaction with services at diagnostic facility/DOTS center** |  |  |  |  |
|  | Totally satisfied | Not reported |  | Ref |  |
|  | Good | Not reported |  | 4.08 (1.39-11.97)* | 0.01* |
|  | Average or less | Not reported |  | 3.18 (1.06-9.51)* | 0.04* |
|  | **Addiction** |  |  |  |  |
|  | None | Ref |  |  |  |
|  | Any addiction | 3.00 (1.73-5.20)* | <0.001* |  |  |
|  | Smokeless tobacco | 2.14 (1.27-3.60)* | 0.004* |  |  |
|  | Smoking | 2.27 (1.13-4.59)* | 0.02* |  |  |
|  | Ever smoked | 3.79 (2.17-6.61)* | <0.001* |  |  |
|  | Alcohol | 3.71 (2.11-6.53)* | <0.001* |  |  |
|  | **Missing any dose during treatment** |  |  |  |  |
|  | No | Ref |  |  |  |
|  | Yes | 1.16 (0.69-1.95) |  |  |  |
|  | **Felt cured and the need to stop medicines during treatment** |  |  |  |  |
|  | No | Ref |  |  |  |
|  | Yes | 2.75 (1.35-5.61)* | 0.005* |  |  |
|  | **Family type** |  |  |  |  |
|  | Joint | Ref |  |  |  |
|  | Nuclear | 1.06 (0.60-1.88) | 0.84 |  |  |
|  | **Family problem** |  |  |  |  |
|  | No | Ref |  |  |  |
|  | Yes | 3.04 (1.66-5.59)* | <0.001* |  |  |
|  | **Family support** |  |  |  |  |
|  | Yes | Ref |  |  |  |
|  | No | 7.22 (2.74-19.06)* | <0.001* |  |  |
|  | **Missing work or education during treatment** |  |  |  |  |
|  | No | Ref |  |  |  |
|  | Yes | 1.17 (0.70-1.94) |  |  |  |
|  | **STS ever visited patient** |  |  |  |  |
|  | No | Ref |  |  |  |
|  | Yes | 0.53 (0.32-0.89)* | 0.02* |  |  |
|  | **Indoor air pollution** |  |  |  |  |
|  | Absent | Ref |  | Ref |  |
|  | Present | 5.37 (2.80-10.30)* | <0.001* | 4.06 (1.67-9.89)* | 0.002* |
|  | **Median travel cost in INR (IQR)^c^** |  |  |  |  |
|  | Per each INP increase in cost | Ref |  |  |  |
|  | Cost of travelling to diagnostic center | 1.00 (0.99-1.01) | 0.37 |  |  |
|  | Cost of travelling to DOTS center | 1.00 (0.99-1.02) | 0.87 |  |  |
|  | **Median distance of health facilities from resident in Km (IQR)^c^** |  |  |  |  |
|  | Per each km increase in distance |  |  |  |  |
|  | Nearest government health facility | 0.97 (0.90-1.05) | 0.48 |  |  |
|  | Distance of diagnostic facility | 0.99 (0.97-1.02) | 0.58 |  |  |
|  | Distance of DOTS center | 1.00 (0.97-1.03) | 0.8 |  |  |
|  | **Type of DOTS provider** |  |  |  |  |
|  | Public health facility based | Ref | 0.35 |  |  |
|  | ASHA/community center | 1.03 (0.61-1.75) |  |  |  |
|  | Others | 2.26 (0.74-6.92) |  |  |  |
|  | **Behavior of DOTS provider** |  |  |  |  |
|  | Very good | Ref | 0.03 |  |  |
|  | Good | 3.39 (0.97-11.87) |  |  |  |
|  | Average | 4.12 (1.14-14.83)* |  |  |  |
|  | Bad | 14.67 (1.83-117.68)* |  |  |  |
|  | **Residence of service provider at DMC** |  |  |  |  |
|  | Same village/ward | Ref |  |  |  |
|  | Different village/ward | 1.19 (0.45-3.20) | 0.73 |  |  |
|  | **Counselling before treatment initiation** |  |  |  |  |
|  | Yes | Ref |  |  |  |
|  | No | 1.15 (0.66-1.99) | 0.62 |  |  |
|  | **Residence of regular DOTS provider** |  |  |  |  |
|  | Same village/ward | Ref |  |  |  |
|  | Different village/ward | 0.87 (0.51-1.47) | 0.59 |  |  |
|  | **DOTS provider visited home for giving medicines** |  |  |  |  |
|  | Yes | Ref |  |  |  |
|  | No | 1.25 (0.75-2.08) | 0.4 |  |  |
| Nahar, 2014 (Madhya Pradesh) *Population: TB patients being treated in the government TB program (no further description, but presumed to comprise new and previously treated patients) Outcome:* *Loss to follow-up as a single outcome* |  | Values below are odds ratios |  |  |  |
|  | **Living situation** |  |  |  |  |
|  | Stable living situation | Ref |  |  |  |
|  | Frequent change of residence or homelessness | 2.30 (2.10-2.50)* | 0.03* |  |  |
|  | **Alcoholism** |  |  |  |  |
|  | No | Ref |  |  |  |
|  | Yes | 1.76 (1.40-2.12)***** | 0.04* |  |  |
|  | **Awareness of exact duration of treatment** |  |  |  |  |
|  | Aware | Ref |  |  |  |
|  | Not aware | 1.72 (1.40-2.04)* | 0.04* |  |  |
|  | **Awareness of consequences of cessation of treatment** |  |  |  |  |
|  | Aware | Ref |  |  |  |
|  | Not aware | 1.77 (1.53-2.04)* | 0.04* |  |  |
|  | **Uncertainty about treatment success** |  |  |  |  |
|  | Certain | Ref |  |  |  |
|  | Uncertain | 1.50 (1.24-1.76)* | 0.05* |  |  |
| Nandakumar, 2013 (Kerala) *Population: New and previously treated TB patients (sputum smear positive pulmonary, sputum smear negative pulmonary, and extrapulmonary) as a combined population Outcome:* *Death, treatment failure, loss to follow-up, and transferred out as a composite outcome* |  | Values below are relative risk ratios |  | Values below are adjusted relative risk ratios |  |
|  | **Sex** |  |  |  |  |
|  | Female | Ref |  | Ref |  |
|  | Male | 1.94 (1.57-2.40)* |  | 1.60 (1.28-1.99)* |  |
|  | **Age (years)** |  |  |  |  |
|  | 15-44 | Ref |  | Ref |  |
|  | >45 | 2.06 (1.71-2.50)* |  | 1.72 (1.40-2.10)* |  |
|  | **Site** |  |  |  |  |
|  | Extrapulmonary | Ref |  | Ref |  |
|  | Pulmonary | 1.72 (1.38-2.14)* |  | 1.3 (0.99-1.72) |  |
|  | **Type of case** |  |  |  |  |
|  | New | Ref |  | Ref |  |
|  | Previously treated | 1.64 (1.34-2.01)* |  | 1.43 (1.18-1.75)* |  |
|  | **Sputum smear status** |  |  |  |  |
|  | Negative/unknown smear | Ref |  | Ref |  |
|  | Positive smear | 1.41 (1.19-1.68)* |  | 1.02 (0.86-1.28) |  |
|  | **HIV status** |  |  |  |  |
|  | Negative | Ref |  | Ref |  |
|  | Positive | 1.64 (0.88-3.04) |  | 1.93 (1.06-3.5)* |  |
|  | Unknown | 1.65 (1.37-1.99)* |  | 1.51 (1.23-1.84)* |  |
|  | **Diabetes** |  |  |  |  |
|  | No | Ref |  | Ref |  |
|  | Yes | 1.25 (1.02-1.53)* |  | 0.99 (0.81-1.21) |  |
|  | Unknown | 1.67 (1.33-2.10)* |  | 1.34 (1.05-1.70)* |  |
|  | **Diabetic control** |  |  |  |  |
|  | Yes | Ref |  |  |  |
|  | No | 2 (0.97-4.13) |  |  |  |
|  | Unknown | 2.14 (1.11-4.13)* |  |  |  |
|  | **DOT in IP** |  |  |  |  |
|  | Regular | Ref |  |  |  |
|  | Missed doses | 2.85 (2.32-3.49)* |  |  |  |
| Pardeshi, 2007^a^ (Maharashtra) *Population: New and previously treated sputum smear positive pulmonary TB patients as a combined population  Outcome: Death,treatment failure, and loss to follow-up as a composite outcome* |  | Values below are odds ratios |  |  |  |
|  | **TB classification** |  |  |  |  |
|  | New smear positive | Ref |  |  |  |
|  | Retreatment | 2.71 (2.12-3.45)* | <0.0001* |  |  |
| Pardeshi, 2010^a^ (Maharashtra) *Population: New and previously treated sputum smear positive pulmonary TB patients as a combined population Outcome:* *Death, treatment failure, loss to follow-up, and transferred out as a composite outcome* |  | Values below are odds ratios |  |  |  |
|  | **Follow-up sputum status at the end of intensive phase** |  |  |  |  |
|  | Negative sputum smear (i.e., sputum conversion) | Ref |  |  |  |
|  | Positive sputum smear (i.e., nonconversion of sputum) | 70.29 (26.48-186.62)* | <0.0001* |  |  |
| Patra, 2013 (Delhi) *Population: New and previously treated TB patients (sputum smear positive pulmonary, sputum smear negative pulmonary, and extrapulmonary) as a combined population among individuals >=60 years old Outcome: Death, treatment failure, loss to follow-up, and transferred out as a composite outcome* |  | Values below are odds ratios |  | Values below are adjusted odds ratios |  |
|  | **Age (years)** |  |  |  |  |
|  | 60-64 | Ref |  |  |  |
|  | 65-74 | 1.2 (0.8-1.7) | Not reported |  |  |
|  | 75 and older | 1.4 (0.8-2.7) | Not reported |  |  |
|  | **Sex** |  |  |  |  |
|  | Female | Ref |  | Ref |  |
|  | Male | 1.8 (1.2-2.5)* | Not reported | 1.6 (1.1-2.2)* | 0.02* |
|  | **TB classification** |  |  |  |  |
|  | Extrapulmonary TB patients | Ref |  | Ref |  |
|  | Smear-positive pulmonary TB patients | 2.6 (1.6-4.3)* | Not reported | 2.2 (1.3-3.8)* | 0.002* |
|  | Smear-negative pulmonary TB patients | 1.7 (0.9-3.0) | Not reported | 1.5 (0.8-2.6) |  |
|  | Unknown TB classification | 1.8 (0.5-7.2) | Not reported | 1.8 (0.4-7.0) |  |
|  | **TB patient type** |  |  |  |  |
|  | New | Ref |  |  |  |
|  | Retreatment | 1.5 (1.0-2.1) | Not reported |  |  |
| Pauniikar, 2019 (Maharashtra) *Population: New and previously treated TB patients (sputum smear positive pulmonary, sputum smear negative pulmonary, and extrapulmonary) as a combined population Outcome: Loss to follow-up as a single outcome* |  | Values below are hazard ratios |  |  |  |
|  | **Gender** |  |  |  |  |
|  | Female | Ref |  |  |  |
|  | Male | 9.09 (1.27-60.30)* | 0.04* |  |  |
|  | **HIV status** |  |  |  |  |
|  | Negative | Ref |  |  |  |
|  | Positive | 4.00 (1.33-12.00) | 0.19 |  |  |
|  | **Smoking** |  |  |  |  |
|  | Non-smoker | Ref |  |  |  |
|  | Person who smokes | 3.70 (1.23-11.11)* | 0.02* |  |  |
|  | **Alcohol** |  |  |  |  |
|  | Non-drinker | Ref |  |  |  |
|  | Person who drinks alcohol | 1.67 (0.56-4.99) | 0.36 |  |  |
| Pore, 2020^a^ (Maharashtra) *Population: Presumed drug-susceptible new and previously treated TB patients and multidrug-resistant TB patients (sputum smear positive pulmonary, sputum smear negative pulmonary, and extrapulmonary) as a combined population Outcome: Treatment non-adherence as a single outcome* |  | Values below are odds ratios |  |  |  |
|  | **Age group** |  |  |  |  |
|  | 21-30 | Ref |  |  |  |
|  | 18-20 | 3.41 (0.50-23.36) | 0.21 |  |  |
|  | 31-40 | 1.82 (0.56-5.85) | 0.32 |  |  |
|  | 41-50 | 1.95 (0.53-7.15) | 0.32 |  |  |
|  | 51-60 | 0.85 (0.19-3.84) | 0.84 |  |  |
|  | 61-70 | 3.41 (0.50-23.36) | 0.21 |  |  |
|  | **Sex** |  |  |  |  |
|  | Female | Ref |  |  |  |
|  | Male | 1.63 (0.56-4.77) | 0.37 |  |  |
|  | **Education** |  |  |  |  |
|  | Graduate and above | Ref |  |  |  |
|  | Illiterate | 1.75 (0.47-6.45) | 0.4 |  |  |
|  | School not completed | 0.87 (0.26-2.97) | 0.83 |  |  |
|  | High school completed | 0.42 (0.10-1.68) | 0.22 |  |  |
|  | **Occupation** |  |  |  |  |
|  | Private job | Ref |  |  |  |
|  | Farmer | 0.96 (0.21-4.34) | 0.96 |  |  |
|  | Housewife | 0.89 (0.19-4.24) | 0.88 |  |  |
|  | Laborer | 1.60 (0.38-6.82) | 0.53 |  |  |
|  | Self-employed | 0.44 (0.08-2.38) | 0.34 |  |  |
|  | Service | 0.80 (0.13-4.75) | 0.81 |  |  |
|  | Others | 4.80 (0.38-59.90) | 0.22 |  |  |
|  | **Treatment category** |  |  |  |  |
|  | Category I (new TB patients) | Ref |  |  |  |
|  | Category II (previously treated TB patients) | 52.00 (10.64-254.23)* | <0.0001* |  |  |
|  | Multidrug-resistant TB patients | 13.70 (0.52-358.07) | 0.12 |  |  |
|  | **Disclosure to family about disease** |  |  |  |  |
|  | Yes | Ref |  |  |  |
|  | No | 0.61 (0.11-3.35) | 0.57 |  |  |
| Prudhivi, 2019^b^ (Andhra Pradesh) *Population: New and previously treated pulmonary TB patients (sputum smear positive and sputum smear negative) as a combined population Outcome: Death, treatment failure, loss to follow-up, and transferred out as a composite outcome* |  | Values below are odds ratios |  | Values below are adjusted odds ratios |  |
|  | **Age (years)** |  |  |  |  |
|  | <=50 | Ref |  | Ref |  |
|  | >50 | 1.45 (1.06-1.98)* | <0.001* | 2.13 (0.50-8.98) |  |
|  | **Gender^f^** |  |  |  |  |
|  | Female | Ref |  | Ref |  |
|  | Male | 1.84 (1.28-2.64)* | <0.001* | 1.96 (1.4-2.52)* | <0.001* |
|  | **Residence** |  |  |  |  |
|  | Urban | Ref |  | Ref |  |
|  | Rural | 1.41 (0.93-2.14) |  | 1.52 (0.96-2.38) |  |
|  | **Type of pulmonary TB^f^** |  |  |  |  |
|  | Sputum smear negative | Ref |  | Ref |  |
|  | Sputum smear positive | 0.28 (0.21-0.39)* | <0.001* | 0.35 (0.28-0.42)* | <0.001* |
|  | **TB category** |  |  |  |  |
|  | New TB patients | Ref |  | Ref |  |
|  | Previously treated TB patients | 3.57 (2.56-4.99)* | <0.001* | 2.94 (2.31-3.74)* | <0.001* |
|  | **HIV status^f^** |  |  |  |  |
|  | Negative | Ref |  | Ref |  |
|  | Positive | 2.87 (1.97-4.17)* | <0.001* | 3.01 (2.11-3.91)* | <0.001* |
|  | **Smoking** |  |  |  |  |
|  | No | Ref |  | Ref |  |
|  | Yes | 1.5 (1.12-2.12)* | <0.001* | 1.7 (1.32-2.08)* | <0.001* |
|  | **Alcohol** |  |  |  |  |
|  | No | Ref |  | Ref |  |
|  | Yes | 1.47 (1.07-2.03)* | <0.001* | 1.39 (0.99-1.79)* | <0.001* |
| Ratnesh, 2020 (Uttar Pradesh) *Population: New and previously treated TB patients as a combined population, without further description of the population Outcome: Loss to follow-up as a single outcome* |  | Values below are odds ratios |  |  |  |
|  | **History of treatment interruption** |  |  |  |  |
|  | No | Ref |  |  |  |
|  | Yes | 7.42 (5.34-10.31)* | 0.001* |  |  |
| Shabil, 2019^a^ (Karnataka) *Population: New and previously treated TB patients as a combined population, without further description of the population Outcome: Loss to follow-up as a single outcome* |  | Values below are odds ratios |  |  |  |
|  | **Age** |  |  |  |  |
|  | 18-29 | Ref |  |  |  |
|  | 30-49 | 1.74 (0.17-18.02) | 0.64 |  |  |
|  | 50-59 | 2.77 (0.23-33.88) | 0.43 |  |  |
|  | 60-69 | 9.00 (0.92-88.17) | 0.06 |  |  |
|  | >=70 | 3.00 (0.16-55.72) | 0.46 |  |  |
|  | **Gender** |  |  |  |  |
|  | Female | Ref |  |  |  |
|  | Male | 1.80 (0.50-6.49) | 0.37 |  |  |
|  | **Residence** |  |  |  |  |
|  | Rural | Ref |  |  |  |
|  | Urban | 0.80 (0.22-2.89) | 0.73 |  |  |
|  | **Education** |  |  |  |  |
|  | Primary Education | Ref |  |  |  |
|  | Illiterate | 0.44 (0.08-2.38) | 0.34 |  |  |
|  | Secondary Education | 0.46 (0.11-1.98) | 0.3 |  |  |
|  | Graduate | 0.29 (0.01-5.79) | 0.42 |  |  |
|  | **Association of interruptions with treatment phase** |  |  |  |  |
|  | Late continuation phase | Ref |  |  |  |
|  | Intensive phase | 7.86 (0.28-217.12) | 0.22 |  |  |
|  | Early continuation phase | 8.33 (0.32-215.69) | 0.2 |  |  |
|  | **Smoker** |  |  |  |  |
|  | No | Ref |  |  |  |
|  | Yes | 0.26 (0.01-4.74) | 0.36 |  |  |
|  | **Alcohol use** |  |  |  |  |
|  | No | Ref |  |  |  |
|  | Yes | 8.33 (1.46-47.64)* | 0.02* |  |  |
|  | **Both smoking and alcohol** |  |  |  |  |
|  | No | Ref |  |  |  |
|  | Yes | 1.09 (0.12-9.95) | 0.94 |  |  |
|  | **Family support** |  |  |  |  |
|  | Yes | Ref |  |  |  |
|  | No | 0.50 (1.15-1.70) | 0.27 |  |  |
|  | **Distance** |  |  |  |  |
|  | 0-2 km | Ref |  |  |  |
|  | 2-5 km | 5.33 (0.56-50.82) | 0.15 |  |  |
|  | 5-10 km | 9.41 (1.02-87.20)* | 0.05* |  |  |
|  | >10 km | 12.80 (0.97-168.74) | 0.05 |  |  |
|  | **Problems faced in getting medicine** |  |  |  |  |
|  | None | Ref |  |  |  |
|  | Transport | 2.25 (0.45-11.31) | 0.32 |  |  |
|  | Staying alone | 2.63 (0.41-16.83) | 0.31 |  |  |
|  | Time | 0.56 (0.03-11.31) | 0.7 |  |  |
|  | Workload | 5.25 (0.94-29.44) | 0.06 |  |  |
|  | **Availability of medicines** |  |  |  |  |
|  | Always | Ref |  |  |  |
|  | Sometimes | 4.00 (0.85-18.84) | 0.08 |  |  |
|  | **Satisfaction with DOTS providers attitudes** |  |  |  |  |
|  | Satisfied | Ref |  |  |  |
|  | Dissatisfied | 2.56 (0.72-9.10) | 0.15 |  |  |
| Sharma, 2003^a^ (Delhi) *Population: New and previously treated TB patients as a combined population, without further description of the population Outcome:* *Treatment failure, LTFU, transferred out and death as a composite outcome* |  | Values below are odds ratios |  |  |  |
|  | **Classification** |  |  |  |  |
|  | Extrapulmonary | Ref |  |  |  |
|  | New sputum smear positive pulmonary TB patients | 8.32 (0.43-162.0) | 0.16 |  |  |
|  | New sputum smear negative sputum pulmonary TB patients | 2.16 (0.08-56.71) | 0.64 |  |  |
|  | Previously treated TB patients | 22.14 (0.86-571.32) | 0.06 |  |  |
| Sharma, 2021^a^ (West Bengal) *Population: New and previously treated TB patients (sputum smear positive pulmonary, sputum smear negative pulmonary, and extrapulmonary) as a combined population Outcome: Death, treatment failure, treatment modified to drug-resistant TB therapy, loss to follow-up, and not evaluated as a composite outcome* |  | Values below are relative risk ratios |  | Values below are adjusted relative risk ratios |  |
|  | **Age** |  |  |  |  |
|  | 15-24 | Ref |  | Ref |  |
|  | 0-14 | 1.5 (0.9-2.7) | Not reported | 1.7 (1.0-2.7)* | 0.04* |
|  | 25-34 | 1.2 (0.9-1.8) | Not reported | 1.6 (1.1-2.1)* | 0.008* |
|  | 35-44 | 1.4 (1.0-2.0)* | Not reported | 2.2 (1.5-3.2)* | <0.001* |
|  | 45-54 | 1.4 (1.0-2.0)* | Not reported | 1.7 (1.2-2.5)* | 0.009* |
|  | 55-64 | 1.1 (0.7-1.7) | Not reported | 1.4 (0.9-2.2) | 0.13 |
|  | 65 and older | 1.5 (1.0-2.3)* | Not reported | 2.1 (1.3-3.2)* | 0.002* |
|  | **Sex** |  |  |  |  |
|  | Female | Ref |  |  |  |
|  | Male | 1.0 (0.8-1.3) | Not reported |  |  |
|  | **Type of TB** |  |  |  |  |
|  | Clinically-diagnosed | Ref |  | Ref |  |
|  | Microbiologically confirmed | 1.3 (1.0-1.6)* | Not reported | 1.2 (0.9-8.5) | 0.16 |
|  | **Site of TB** |  |  |  |  |
|  | Pulmonary | Ref |  |  |  |
|  | Extrapulmonary | 1.1 (0.8-1.4) | Not reported |  |  |
|  | **Category of TB** |  |  |  |  |
|  | New | Ref |  | Ref |  |
|  | Previously treated | 1.4 (1.0-2.1)* | Not reported | 1.1 (0.8-1.6) | 0.61 |
|  | **Source of TB medications** |  |  |  |  |
|  | Private pharmacy | Ref |  | Ref |  |
|  | National TB Program (free) | 3.6 (3.0-4.3)* | Not reported | 4.0 (3.1-5.0)* | <0.001* |
|  | **HIV status** |  |  |  |  |
|  | Negative/Unknown | Ref |  |  |  |
|  | Positive | 1.8 (0.3-9.8) | Not reported |  |  |
|  | **Diabetic status** |  |  |  |  |
|  | No/Unknown | Ref |  |  |  |
|  | Diabetic | 1.1 (0.8-1.5) | Not reported |  |  |
|  | **District** |  |  |  |  |
|  | East Medinipur | Ref |  | Ref |  |
|  | Kolkota | 4.8 (1.6-14.6)* | Not reported | 2.8 (0.9-8.5) | 0.07 |
|  | Howrah | 3.7 (1.2-11.5)* | Not reported | 2.5 (0.8-8.3) | 0.13 |
|  | Hooghly | 3.0 (0.9-9.8) | Not reported | 2.0 (0.6-6.6) | 0.25 |
|  | North 24 Parganas | 3.4 (1.1-10.8)* | Not reported | 2.3 (0.7-7.5) | 0.16 |
|  | South 24 Parganas | 4.6 (1.8-17.6)* | Not reported | 3.3 (1.1-10.3)* | 0.04* |
|  | **TB care in the district of residence** |  |  |  |  |
|  | Same district | Ref |  |  |  |
|  | Different district | 1.1 (0.8-1.4) | Not reported |  |  |
|  | **Type of health facility** |  |  |  |  |
|  | Clinic | Ref |  |  |  |
|  | Hospital/Nursing home | 1.5 (0.8-3.0) | Not reported |  |  |
|  | **Education of provider** |  |  |  |  |
|  | MBBS | 1.0 (0.8-1.3) | Not reported |  |  |
| Shivam, 2014^a^ (West Bengal) *Population: New and previously treated TB patients (sputum smear positive pulmonary, sputum smear negative pulmonary, and extrapulmonary) as a combined population Outcome: Death, treatment failure, and loss to follow-up as a composite outcome* |  | Values below are odds ratios |  |  |  |
|  | **Treatment category** |  |  |  |  |
|  | Category I (new) | Ref |  |  |  |
|  | Category II (previously treated) | 2.10 (1.40-3.16)* | 0.0004* |  |  |
| Siddiqui, 2016 (Delhi) *Population: New and previously treated TB patients (sputum smear positive pulmonary, sputum smear negative pulmonary, and extrapulmonary) as a combined population Outcome:* *Death, treatment failure, treatment modified to drug-resistant TB therapy, and loss to follow-up as a composite outcome* |  |  |  | Values below are adjusted odds ratios |  |
|  | **Gender** |  |  |  |  |
|  | Female |  |  | Ref |  |
|  | Male |  |  | 0.31 (0.06-1.76) |  |
|  | **Age (years)^c^** |  |  |  |  |
|  | Per each year increase in age |  |  | 0.96 (0.90-1.02) |  |
|  | **Category of TB** |  |  |  |  |
|  | Cat I |  |  | 0.84 (0.24-2.89) |  |
|  | Cat II |  |  | Ref |  |
|  | **TB history** |  |  |  |  |
|  | No |  |  | Ref |  |
|  | Yes |  |  | 2.59 (0.26-25.82) |  |
|  | **ADR incidence** |  |  |  |  |
|  | No |  |  | Ref |  |
|  | Yes |  |  | 0.64 (0.19-2.21) |  |
|  | **Fever** |  |  |  |  |
|  | No |  |  | Ref |  |
|  | Yes |  |  | 0.81 (0.09-2.16) |  |
|  | **Dyspnea** |  |  |  |  |
|  | No |  |  | Ref |  |
|  | Yes |  |  | 1.97 (0.34-5.87) |  |
|  | **Chest pain** |  |  |  |  |
|  | No |  |  | Ref |  |
|  | Yes |  |  | 1.37 (0.49-6.14) |  |
|  | **Hemoptysis** |  |  |  |  |
|  | No |  |  | Ref |  |
|  | Yes |  |  | 0.81 (0.29-2.28) |  |
|  | **Diabetes** |  |  |  |  |
|  | No |  |  | Ref |  |
|  | Yes |  |  | 0.71 (0.16-3.28) |  |
|  | **BMI^c^** |  |  |  |  |
|  | Per each kg/m^2^ increase in age |  |  | 1.19 (0.97-1.45) |  |
|  | **Weight gain** |  |  |  |  |
|  | No |  |  | Ref |  |
|  | Yes |  |  | 0.71 (0.06-8.52) |  |
|  | **Anorexia** |  |  |  |  |
|  | No |  |  | Ref |  |
|  | Yes |  |  | 0.56 (0.05-3.12) |  |
|  | **Alcohol intake** |  |  |  |  |
|  | No |  |  | Ref |  |
|  | Yes |  |  | 0.67 (0.17-2.56) |  |
|  | **Smoking** |  |  |  |  |
|  | No |  |  | Ref |  |
|  | Yes |  |  | 0.75 (0.24-2.39) |  |
|  | **Chewing tobacco** |  |  |  |  |
|  | No |  |  | Ref |  |
|  | Yes |  |  | 0.78 (0.24-2.54) |  |
| Singh, 2020 (Uttarakhand) *Population: New and previously treated TB patients (sputum smear positive pulmonary, sputum smear negative pulmonary, and extrapulmonary) as a combined population Outcome: Death, treatment failure, treatment regimen modified, loss to follow-up, transferred out and not evaluated as a composite outcome* |  | Values below are relative risk ratios |  | Values below are adjusted relative risk ratios |  |
|  | **Sex** |  |  |  |  |
|  | Female | Ref |  |  |  |
|  | Male | 1.5 (0.8-2.7) | 0.19 | 1.3 (0.7-2.2) | 0.4 |
|  | **Age** |  |  |  |  |
|  | 0-14 | Ref |  |  |  |
|  | 15-44 | 0.9 (0.2-5.5) | 0.9 |  |  |
|  | 45 and older | 1.5 (0.3-9.0) | 0.8 |  |  |
|  | **Category of patient** |  |  |  |  |
|  | Category I (new) | Ref |  | Ref |  |
|  | Category II (previously treated) | 2.0 (1.2-3.6)* | 0.02* | 3.2 (2.1-4.9)* | 0.0001* |
|  | **Site of TB** |  |  |  |  |
|  | Extrapulmonary | Ref |  | Ref |  |
|  | Pulmonary | 9.3 (1.3-35.5)* | 0.003* | 5.6 (0.8-39.8) | 0.08 |
|  | **Bacteriological status** |  |  |  |  |
|  | Clinically confirmed | Ref |  |  |  |
|  | Bacteriologically confirmed | 2.3 (1.2-4.5)* | 0.008* |  |  |
|  | **Type of regimen** |  |  |  |  |
|  | Daily | Ref |  |  |  |
|  | Intermittent | 1.2 (0.7-2.2) | 0.4 |  |  |
|  | **Treatment delays (days)** |  |  |  |  |
|  | 7 or fewer | Ref |  | Ref |  |
|  | More than 7 | 1.7 (0.8-3.9) | 0.2 | 1.5 (0.8-2.0) | 0.2 |
|  | **Type of case finding** |  |  |  |  |
|  | Passive case finding | Ref |  | Ref |  |
|  | Active case finding | 2.5 (1.5-4.2)* | <0.001* | 2.6 (1.7-4.0)* | <0.001* |
| Vasantha, 2008 (Tamil Nadu) *Population: New and previously treated TB patients (sputum smear positive pulmonary, sputum smear negative pulmonary, and extrapulmonary) as a combined population Outcome: Death as a single outcome* |  |  |  | Values below are adjusted hazard ratios |  |
|  | **Gender** |  |  |  |  |
|  | Female |  |  | Ref |  |
|  | Male |  |  | 1.32 (0.73-2.39) | 0.36 |
|  | **Age (years)** |  |  |  |  |
|  | <45 |  |  | Ref |  |
|  | >=45 |  |  | 2.35 (1.56-3.55)* | <0.001* |
|  | **Occupation** |  |  |  |  |
|  | Employed |  |  | Ref |  |
|  | Unemployed |  |  | 1.38 (0.92-2.06) | 0.12 |
|  | **Education** |  |  |  |  |
|  | Literate |  |  | Ref |  |
|  | Illiterate |  |  | 1.28 (0.87-1.88) | 0.22 |
|  | **Treatment category** |  |  |  |  |
|  | Category III |  |  | Ref |  |
|  | Category I |  |  | 1.19 (0.76-1.86) | Not reported |
|  | Category II |  |  | 0.77 (0.39-1.51) | Not reported |
|  | **Previous treatment** |  |  |  |  |
|  | New patient |  |  | Ref |  |
|  | Retreatment (previously treated) |  |  | 1.62 (1.10-2.37)* | <0.05* |
|  | **Bodyweight** |  |  |  |  |
|  | >=35 kg |  |  | Ref |  |
|  | <35 kg |  |  | 3.71 (2.43-5.65)* | <0.001* |
|  | **Alcoholism** |  |  |  |  |
|  | No alcoholism |  |  | Ref |  |
|  | Alcoholism |  |  | 2.02 (1.36-2.99)* | <0.005* |
|  | **Smoking** |  |  |  |  |
|  | No smoking |  |  | Ref |  |
|  | Smoking |  |  | 0.73 (0.44-1.23) | 0.24 |
|  | **Type of DOT provider** |  |  |  |  |
|  | Friends, relatives, self, and others |  |  | Ref |  |
|  | Government DOT |  |  | 1.31 (0.72-2.38) | 0.38 |
|  | Community DOT |  |  | 0.99 (0.54-1.81) | 0.97 |
|  | **Supervision under IP** |  |  |  |  |
|  | Always |  |  | Ref |  |
|  | Never |  |  | 1.17 (0.77-1.78) | 0.47 |
| Vashishtha, 2013^a^ (Delhi) *Population: New and previously treated TB patients (sputum smear positive pulmonary, sputum smear negative pulmonary, and extrapulmonary) as a combined population among people with and without HIV Outcome: Death, treatment failure, treatment regimen modified, and loss to follow-up as a composite outcome* |  | Values below are odds ratios |  |  |  |
|  | **HIV status** |  |  |  |  |
|  | HIV-negative | Ref |  |  |  |
|  | HIV-positive | 4.25 (2.02-8.96)* | 0.0001* |  |  |
| Vasudevan, 2014^a^ (Puducherry) *Population: New and previously treated TB patients (sputum smear positive pulmonary, sputum smear negative pulmonary, and extrapulmonary) as a combined population Outcome:* *Loss to follow-up as a single outcome* |  | Values below are odds ratios |  |  |  |
|  | **Treatment category** |  |  |  |  |
|  | Category I (new) | Ref |  |  |  |
|  | Category II (previously treated) | 2.03 (1.13-3.65)* | 0.02* |  |  |
| Viswanathan, 2014^a^ (Tamil Nadu) *Population: New and previously treated TB patients (sputum smear positive pulmonary, sputum smear negative pulmonary, and extrapulmonary) as a combined population Outcome: Death, treatment failure, and loss to follow-up as a composite outcome* |  | Values below are odds ratios |  |  |  |
|  | **Diabetic status** |  |  |  |  |
|  | Non-diabetic | Ref |  |  |  |
|  | Diabetic | 2.59 (0.84-8.01) | 0.1 |  |  |
| Washington, 2020 (Karnataka and Telangana) *Population: Presumed drug-susceptible new and previously treated TB patients and multidrug-resistant TB patients (sputum smear positive pulmonary, sputum smear negative pulmonary, and extrapulmonary) as a combined population Outcome:* *Death, treatment failure, and loss to follow-up as a composite outcome* |  |  |  | Values below are adjusted odds ratios |  |
|  | **Gender** |  |  |  |  |
|  | Female |  |  | Ref |  |
|  | Male |  |  | 1.69 (1.24-2.30)* | <0.001* |
|  | **Age** |  |  |  |  |
|  | Below 60 |  |  | Ref |  |
|  | 60 and above |  |  | 1.12 (0.79-1.60) | 0.53 |
|  | **State** |  |  |  |  |
|  | Telangana |  |  | Ref |  |
|  | Karnataka |  |  | 2.46 (1.79-3.39)* | <0.001* |
|  | **Religion** |  |  |  |  |
|  | Muslim |  |  | Ref |  |
|  | Hindu |  |  | 1.10 (0.82-1.47) | 0.54 |
|  | Other religion |  |  | 1.40 (0.74-2.6) | 0.3 |
|  | **Marital status** |  |  |  |  |
|  | Single |  |  | Ref |  |
|  | Married |  |  | 0.95 (0.68-1.34) | 0.78 |
|  | Marriage dissolved |  |  | 1.22 (0.66-2.24) | 0.52 |
|  | Marriage not known |  |  | 3.23 (0.70-14.96) | 0.13 |
|  | **Living alone** |  |  |  |  |
|  | No |  |  | Ref |  |
|  | Yes |  |  | 0.40 (0.38-1.42) | 0.47 |
|  | **Education** |  |  |  |  |
|  | 10th standard or more |  |  | Ref |  |
|  | Less than 5th standard |  |  | 2.74 (1.71-4.41)* | <0.001***** |
|  | 5th to 10th standard |  |  | 2.41 (1.51-3.87)* | <0.001* |
|  | **TB site** |  |  |  |  |
|  | Extrapulmonary |  |  | Ref |  |
|  | Pulmonary |  |  | 1.37 (0.96-1.95) | 0.08 |
|  | **Previously treated for TB** |  |  |  |  |
|  | No |  |  | Ref |  |
|  | Yes |  |  | 1.58 (1.18-2.11)* | 0.003* |
|  | **Drug resistant-TB** |  |  |  |  |
|  | No |  |  | Ref |  |
|  | Yes |  |  | 2.33 (1.41-3.87)* | 0.001* |
|  | **Drink alcohol** |  |  |  |  |
|  | No |  |  | Ref |  |
|  | Yes |  |  | 1.38 (1.01-1.88)* | 0.04* |
|  | **HIV** |  |  |  |  |
|  | No |  |  | Ref |  |
|  | Yes |  |  | 2.61 (1.41-4.82)* | 0.002* |
|  | **Diabetes** |  |  |  |  |
|  | No |  |  | Ref |  |
|  | Yes |  |  | 0.70 (0.37-1.32) | 0.27 |
|  | **Initial weight** |  |  |  |  |
|  | Greater than median |  |  | Ref |  |
|  | Less than median |  |  | 1.89 (1.43-2.50)* | <0.001* |
|  | Unknown |  |  | 1.70 (1.22-2.37)* | 0.002* |
| Washington, 2020  (Karnataka and Telangana) *Population: Presumed drug-susceptible new and previously treated TB patients and multidrug-resistant TB patients (sputum smear positive pulmonary, sputum smear negative pulmonary, and extrapulmonary) as a combined population Outcome:* *Death as a single outcome* |  |  |  | Values below are adjusted odds ratios |  |
|  | **Gender** |  |  |  |  |
|  | Female |  |  | Ref |  |
|  | Male |  |  | 0.95 (0.61-1.48) | 0.83 |
|  | **Age** |  |  |  |  |
|  | Below 60 |  |  | Ref |  |
|  | 60 and above |  |  | 2.15 (1.37-3.37)* | 0.001* |
|  | **State** |  |  |  |  |
|  | Telangana |  |  | Ref |  |
|  | Karnataka |  |  | 1.54 (1.01-2.35)* | 0.05* |
|  | **Religion** |  |  |  |  |
|  | Muslim |  |  | Ref |  |
|  | Hindu |  |  | 0.94 (0.62-1.45) | 0.79 |
|  | Other religion |  |  | 1.71 (0.75-3.91) | 0.21 |
|  | **Marital status** |  |  |  |  |
|  | Single |  |  | Ref |  |
|  | Married |  |  | 1.09 (0.62-1.93) | 0.76 |
|  | Marriage dissolved |  |  | 0.88 (0.35-2.23) | 0.8 |
|  | **Living alone** |  |  |  |  |
|  | No |  |  | Ref |  |
|  | Yes |  |  | 0.68 (0.24-1.89) | 0.46 |
|  | **Education** |  |  |  |  |
|  | 10th standard or more |  |  | Ref |  |
|  | Less than 5th standard |  |  | 5.38 (2.10-13.83)* | <0.001***** |
|  | 5th to 10th standard |  |  | 3.99 (1.55-10.31)* | <0.004* |
|  | **TB site** |  |  |  |  |
|  | Extrapulmonary |  |  | Ref |  |
|  | Pulmonary |  |  | 1.09 (0.66-1.80) | 0.74 |
|  | **Previously treated for TB** |  |  |  |  |
|  | No |  |  | Ref |  |
|  | Yes |  |  | 1.65 (1.08-2.51)* | 0.02* |
|  | **Drug-resistant TB** |  |  |  |  |
|  | No |  |  | Ref |  |
|  | Yes |  |  | 1.83 (0.86-3.89) | 0.12 |
|  | **Drink alcohol** |  |  |  |  |
|  | No |  |  | Ref |  |
|  | Yes |  |  | 2.09 (1.35-3.25)* | 0.001* |
|  | **HIV** |  |  |  |  |
|  | No |  |  | Ref |  |
|  | Yes |  |  | 4.75 (2.29-9.86)* | <0.001* |
|  | **Diabetes** |  |  |  |  |
|  | No |  |  | Ref |  |
|  | Yes |  |  | 0.74 (0.29-1.86) | 0.52 |
|  | **Initial weight** |  |  |  |  |
|  | Greater than median |  |  | Ref |  |
|  | Less than median |  |  | 1.98 (1.30-3.00)* | 0.001* |
|  | Unknown |  |  | 1.96 (1.22-3.15)* | 0.005* |
| Studies in pediatric TB patients who do not achieve treatment success as a single outcome or part of a composite outcome |  |  |  |  |  |
| Dhakulkar, 2021 (Maharashtra) *Population: Children ages 0-19 with drug-resistant TB Outcome: Death, treatment failure, and loss to follow-up as a composite outcome* |  | Values below are odds ratios |  | Values below are adjusted odds ratios |  |
|  | **Age** |  |  |  |  |
|  | 0-9 | Ref |  | Ref |  |
|  | 10—19 | 10.0 (1.3-77.1)* |  | 4.4 (0.4-42.9) |  |
|  | **Sex** |  |  |  |  |
|  | Female | Ref |  | Ref |  |
|  | Male | 1.4 (0.8-2.3) |  | 1.2 (0.6-2.4) |  |
|  | **TB site** |  |  |  |  |
|  | Extrapulmonary | Ref |  | Ref |  |
|  | Pulmonary | 3.5 (1.8-6.8)* |  | 1.9 (0.8-4.4) |  |
|  | **TB resistant patterns** |  |  |  |  |
|  | Multidrug-resistant TB | Ref |  | Ref |  |
|  | Pre-extensively drug-resistant TB | 1.9 (1.1-3.4)* |  | 1.7 (0.8-3.4) |  |
|  | Extensively drug-resistant TB | 7.5 (3.2-17.3)* |  | 4.3 (1.3-13.8)* |  |
|  | **Previous TB episodes** |  |  |  |  |
|  | Absent | Ref |  | Ref |  |
|  | Present | 2.0 (1.2-3.4)* |  | 1.4 (0.7-2.9) |  |
|  | **Nutritional status** |  |  |  |  |
|  | Normal | Ref |  | Ref |  |
|  | Undernourished | 3.8 (2.1-6.9)* |  | 2.5 (1.3-4.8)* |  |
| Raizada, 2018^a^ (4 Indian States) *Population: Children ages 0-14 with drug-susceptible TB Outcome: Death as a single outcome versus treatment completion (with exclusion of patients still on treatment, with treatment failure, or with loss to follow-up from the analysis)* |  | Values below are odds ratios |  |  |  |
|  | **Age (years)** |  |  |  |  |
|  | 10 to 14 | Ref |  |  |  |
|  | 5 to 9 | 1.59 (0.86-2.94) | 0.14 |  |  |
|  | 0 to 4 | 3.31 (1.92-5.72)* | <0.0001* |  |  |
|  | **Sex** |  |  |  |  |
|  | Female | Ref |  |  |  |
|  | Male | 1.36 (0.84-2.19) | 0.21 |  |  |
| Raizada, 2018^a^ (4 Indian States) *Population: Children ages 0-14 with drug-susceptible TB Outcome: Death, treatment failure, and loss to follow-up as a composite outcome versus treatment completion (with exclusion of patients still on treatment from the analysis)* |  | Values below are odds ratios |  |  |  |
|  | **Age (years)** |  |  |  |  |
|  | 10 to 14 | Ref |  |  |  |
|  | 5 to 9 | 1.56 (0.88-2.75) | 0.12 |  |  |
|  | 0 to 4 | 2.76 (1.63-4.67)* | 0.0002* |  |  |
|  | **Sex** |  |  |  |  |
|  | Female | Ref |  |  |  |
|  | Male | 1.17 (0.74-1.86) | 0.49 |  |  |
| Sadana, 2020^a^ (Punjab) *Population: Children 0-14 years with new or previously treated TB Outcome:* *Death, treatment modified, and loss to follow-up as a composite outcome* |  | Values below are odds ratios |  |  |  |
|  | **Age** |  |  |  |  |
|  | 0 to 5 | Ref |  |  |  |
|  | 6 to 10 | 7.0 (0.32-152.96) | 0.22 |  |  |
|  | 11 to 14 | 0.82 (0.03-21.58) | 0.9 |  |  |
|  | **Sex** |  |  |  |  |
|  | Male | Ref |  |  |  |
|  | Female | 0.45 (0.06-3.45) | 0.44 |  |  |
|  | **Area of residence** |  |  |  |  |
|  | Urban | Ref |  |  |  |
|  | Rural | 1.14 (0.11-11.81) | 0.91 |  |  |
|  | **Diagnosis** |  |  |  |  |
|  | Ziehl-Neelson sputum microscopy | Ref |  |  |  |
|  | Chest X-Ray | 2.30 (0.13-40.55) | 0.57 |  |  |
|  | Others | 2.19 (0.18-25.96) | 0.53 |  |  |
|  | Cartridge-based nucleic acid amplification testing | 1.74 (0.06-49.90) | 0.75 |  |  |
|  | **Site** |  |  |  |  |
|  | Pulmonary | Ref |  |  |  |
|  | Extrapulmonary | 3.21 (0.32-32.74) | 0.32 |  |  |
|  | **Type of case** |  |  |  |  |
|  | Retreatment (previously treated) | Ref |  |  |  |
|  | New | 0.93 (0.04-19.55) | 0.96 |  |  |
|  | **Contact history** |  |  |  |  |
|  | Absent | Ref |  |  |  |
|  | Present | 33.5 (1.69-665.30)* | 0.02* |  |  |
| Satyanarayana, 2010^a^ (Delhi) *Population: Children 0-14 years with new or previously treated TB Outcome: Death, treatment failure, lost to follow-up, and transferred out as a composite outcome* |  | Values below are odds ratios |  |  |  |
|  | **Sex** |  |  |  |  |
|  | Female | Ref |  |  |  |
|  | Male | 0.70 (0.39-1.26) | 0.23 |  |  |
|  | **Age (years)** |  |  |  |  |
|  | <5 | Ref |  |  |  |
|  | 5 to 10 | 1.19 (0.46-3.07) | 0.72 |  |  |
|  | 11 to 15 | 1.59 (0.65-3.89) | 0.31 |  |  |
|  | **TB classification** |  |  |  |  |
|  | Extrapulmonary | Ref |  |  |  |
|  | Pulmonary | 2.08 (1.20-3.61)* | 0.009* |  |  |
|  | **Type of TB** |  |  |  |  |
|  | New extrapulmonary | Ref |  |  |  |
|  | New smear negative | 1.85 (0.89-3.86) | 0.1 |  |  |
|  | New smear positive | 2.78 (1.33-5.83)* | 0.007* |  |  |
|  | Previously treated "other" (i.e., smear negative or extrapulmonary) | 3.23(1.16-9.00)* | 0.02* |  |  |
|  | Previously treated smear positive | 5.82 (1.57-21.60)* | 0.008* |  |  |
|  | **Extrapulmonary TB site** |  |  |  |  |
|  | Peripheral lymph node | Ref |  |  |  |
|  | Other sites | 1.27 (0.51-3.21) | 0.61 |  |  |
|  | **Treatment category** |  |  |  |  |
|  | Category I  (mostly new smear positive pulmonary TB patients) | Ref |  |  |  |
|  | Category II  (previously treated TB patients) | 2.40 (1.07-5.37)* | 0.03* |  |  |
|  | Category III (mostly new smear negative and extrapulmonary TB patients who were not seriously ill) | 0.69 (0.31-1.50) | 0.35 |  |  |
|  | **RNTCP pre-treatment weight bands** |  |  |  |  |
|  | >17-25 kg | Ref |  |  |  |
|  | <=10 kg | 1.17 (0.42-3.27) | 0.76 |  |  |
|  | >10-17 kg | 0.95 (0.42-2.17) | 0.9 |  |  |
|  | >25-30 kg | 1.38 (0.60-3.18) | 0.45 |  |  |
|  | >30 kg | 0.74 (0.33-1.64) | 0.46 |  |  |
|  | **DOT center type** |  |  |  |  |
|  | Government health facility | Ref |  |  |  |
|  | Other DOT provider | 0.87 (0.45-1.65) | 0.67 |  |  |
| Studies in HIV-TB coinfected patients who do not achieve treatment success as a single outcome or part of a composite outcome |  |  |  |  |  |
| Sharma, 2014 (Delhi) *Outcome: Death, treatment failure, and loss to follow-up as a composite outcome* |  | Values below are odds ratios |  | Values below are adjusted odds ratios |  |
|  | **Age (years)** |  |  |  |  |
|  | >40 | Ref |  |  |  |
|  | <=40 | 1.17 (0.67-2.03) | 0.57 |  |  |
|  | **Gender** |  |  |  |  |
|  | Female | Ref |  |  |  |
|  | Male | 1.97 (1.02-3.8)* | 0.04* | 1.87 (0.96-3.67) | 0.06 |
|  | **Classification** |  |  |  |  |
|  | Extrapulmonary | Ref |  |  |  |
|  | Pulmonary | 1.16 (0.73-1.84) | 0.51 |  |  |
|  | **Sputum smear** |  |  |  |  |
|  | Smear negative | Ref |  |  |  |
|  | Smear positive | 1.41 (0.67-2.96) | 0.35 |  |  |
|  | **CD4 count** |  |  |  |  |
|  | >200 | Ref |  |  |  |
|  | <=200 | 2.61 (1.20-5.66)* | 0.01* | 2.32 (1.06-5.09)* | 0.03* |
|  | **Patient type** |  |  |  |  |
|  | New | Ref |  |  |  |
|  | Previously treated | 3.33 (1.42-7.81)* | 0.004* | 2.91 (1.22-6.89)* | 0.02* |
|  | **ATT type** |  |  |  |  |
|  | Daily therapy | Ref |  |  |  |
|  | DOTS | 0.99 (0.53-1.85) | 0.98 |  |  |
|  | **ATT side effects** |  |  |  |  |
|  | No | Ref |  |  |  |
|  | Yes | 1.52 (0.90-2.57) | 0.11 |  |  |
|  | **ART at diagnosis of TB** |  |  |  |  |
|  | On ART | No |  |  |  |
|  | ART naïve | 2.62 (0.90-7.58) | 0.06 | 2.42 (0.82-7.07) | 0.1 |
| Shastri, 2013^a^ (Karnataka) *Outcome: Death, treatment failure, and loss to follow-up as a composite outcome* |  | Values below are odds ratios |  |  |  |
|  | **ART** |  |  |  |  |
|  | On ART | Ref |  |  |  |
|  | Not on ART | 2.83 (2.43-3.29)* | <0.0001* |  |  |
|  | **HIV co-infection** |  |  |  |  |
|  | TB only | Ref |  |  |  |
|  | HIV/TB co-infection | 1.16 (1.08-1.25)* | <0.0001* |  |  |
| Vijay, 2011 (Karnataka) *Outcome: Death, treatment failure, and loss to follow-up as a composite outcome* |  | Values below are odds ratios |  | Values below are adjusted odds ratios |  |
|  | **Disease Classification** |  |  |  |  |
|  | Pulmonary | 2.46 (1.35-4.53) | 0.002* | 1.96 (1.02-3.77)* | 0.04* |
|  | Extra-pulmonary | Ref |  | Ref |  |
|  | **Sputum smear** |  |  |  |  |
|  | Smear positive | 1.77 (0.80-3.94) | 0.12 |  |  |
|  | Smear negative | Ref |  |  |  |
|  | **Type of patient** |  |  |  |  |
|  | Previously treated patient | 4.04 (1.96-8.35) | <0.001* | 4.78 (2.12-10.76)* | <0.001* |
|  | New patient | Ref |  | Ref |  |
|  | **Regularity of treatment** |  |  |  |  |
|  | Irregular | 1.63 (0.86-3.09) | 0.107 |  |  |
|  | Regular | Ref |  |  |  |
|  | **Baseline CD4 count/mm3** |  |  |  |  |
|  | <=350 | 0.80 (0.26-3.29) | 0.83 |  |  |
|  | >350 | Ref |  |  |  |
|  | **ART initiation** |  |  |  |  |
|  | Not on ART | 9.12 (4.48-18.88)* | <0.001* | 4.9 (1.85-12.96)* | 0.001* |
|  | On ART | Ref |  | Ref |  |
|  | **CPT provision** |  |  |  |  |
|  | Not initiated | 6.65 (3.53-12.61)* | <0.001* | 2.19 (0.90-5.32) | 0.08 |
|  | Initiated | Ref |  | Ref |  |
| Vijay, 2011 (Karnataka) *Outcome:* *Death as a single outcome* |  | Values below are odds ratios |  | Values below are adjusted odds ratios |  |
|  | **Disease Classification** |  |  |  |  |
|  | Pulmonary | 2.21 (1.26-3.88)* | 0.002* | 1.82 (1.00-1.33)* | 0.050* |
|  | Extra-pulmonary | Ref |  | Ref |  |
|  | **Sputum smear** |  |  |  |  |
|  | Smear positive | 1.53 (0.72-3.26) | 0.23 |  |  |
|  | Smear negative | Ref |  |  |  |
|  | **Type of patient** |  |  |  |  |
|  | Previously treated patient | 2.05 (1.01-4.18)* | 0.03* | 1.94 (0.89-4.21) | 0.09 |
|  | New patient | Ref |  | Ref |  |
|  | **Regularity of treatment** |  |  |  |  |
|  | Irregular | 2.07 (1.14-3.78)* | 0.01* |  |  |
|  | Regular | Ref |  |  |  |
|  | **Baseline CD4 count/mm3** |  |  |  |  |
|  | <=200 | 0.59 (0.26-1.35) | 0.17 |  |  |
|  | >200 | Ref |  |  |  |
|  | **ART initiation** |  |  |  |  |
|  | Not on ART | 7.75 (4.12-14.71)* | <0.001* | 2.80 (1.15-6.81)* | 0.023* |
|  | On ART | Ref |  | Ref |  |
|  | **Cotrimoxazole prophylactic therapy provision** |  |  |  |  |
|  | Not initiated | 7.76 (4.22-14.35)* | <0.001* | 3.46 (1.47-8.14)* | 0.004* |
|  | Initiated | Ref |  | Ref |  |

SAT, Self-administered Treatment; DOTS, Directly Observed Treatment Shortcourse; TB, tuberculosis.

*Indicates statistical significance

^**^The treatment category of retreatment "other" was used by the TB program to refer to previously treated patients with smear-negative or extrapulmonary TB.

^***^The reference group, and therefore the effect estimate and confidence interval, was "flipped" to facilitate comparability of the reference group with other studies

^a^Unadjusted odds ratios and/or p-values were estimated by the systematic review team from the raw data, as these were not provided in the original study.

^b^Study reported successful treatment as the outcome, so effect estimates (odds ratio or relative risk and 95% confidence interval) were flipped to show effect estimates for the outcome of not achieving treatment success.

^c^Variable was included in the analysis as a continuous variable, we have specified the unit of change in the variable associated with the effect estimate.

^d^This is also an adjusted model but represents analysis on an unweighted cohort (i.e., only individuals who responded to a survey)

^e^This is an adjusted model that represents analysis on a weighted cohort (i.e., includes individual who responded to a survey

^f^Reference group was switched for comparability across studies, resulting in flipping of the effect estimate

### References

[to be inserted after review]
