## Supplemental Appendix 5 for "Barriers to engagement by people with active tuberculosis in the care cascade in India: a systematic review of two decades of quantitative research"

**Supplementary Appendix 5**

**Supplement to:**

Barriers to engagement by people with active tuberculosis in the care cascade in India: a systematic review of two decades of quantitative research

Tulip A. Jhaveri, Disha Jhaveri, Amith Galivanche, Dominic Voehler, Maya Lubeck-Schricker, Mei Chung, Pruthu Thekkur, Vineet Chadha, Ruvandhi Nathavitharana, Ajay M.V. Kumar, Hemant Deepak Shewade, Katherine Powers, Kenneth H. Mayer, Jessica E. Haberer, Paul Bain, Madhukar Pai, Srinath Satyanarayana, Ramnath Subbaraman

**Correspondence:**

Ramnath Subbaraman, MD, MSc, FACP

Tufts University School of Medicine

Department of Public Health and Community Medicine

136 Harrison Ave., MV237

Boston, MA 02130, USA

**Methods and findings for the systematic review of barriers to achieving recurrence-free survival after completion of tuberculosis treatment (Gap 5)**

### Methods

##### Objectives

The objective of this systematic review was to understand why some individuals who complete tuberculosis (TB) treatment do not subsequently achieve TB recurrence-free survival (Gap 5 in the care cascade). We included patients who underwent TB treatment either in India’s National TB Elimination Programme (NTEP) or at private sector facilities. We included studies that evaluated patient outcomes starting from TB treatment initiation or treatment completion, as long as patients were followed (and outcomes captured) into the initial few months or years of the post-treatment period.

Unfavorable outcomes in this analysis comprised post-treatment TB recurrence, post-treatment mortality, or both. As such, we break down our reporting of Gap 5 findings as follows:

(a) *Studies that evaluated post-treatment TB recurrence (with or without evaluating post-treatment mortality)*: These studies followed patients starting either from treatment initiation or treatment completion and captured information on post-treatment TB recurrence alone or as part of a composite outcome (e.g., along with unfavorable outcomes during treatment or post-treatment mortality).
(b) *Studies that only evaluated post-treatment mortality (without evaluating post-treatment TB recurrence)*: These studies followed patients starting either from treatment initiation or treatment completion and captured information on post-treatment TB mortality alone or as part of a composite outcome (e.g., along with unfavorable outcomes during treatment), but *did not* evaluate patients for TB recurrence.

We extracted data on factors associated with post-treatment recurrence and/or death from studies comparing individuals who had or had not achieved favorable post-treatment outcomes. For these studies, we extracted effect estimates for independent variables (i.e., exposures or predictors) associated with post-treatment recurrence and/or death, or related composite outcomes. Effect estimates included odds ratios, relative risk ratios, hazard ratios, or beta-coefficients, depending on the approach to analysis in each study. While we also initially aimed to extract reasons reported by patients or their family members for post-treatment TB recurrence or mortality (e.g., as captured in structured surveys), none of the identified studies reported such findings.

##### Search strategy

Three separate searches were conducted to identify articles. The first search was conducted as part of a previously published study quantifying gaps in India’s TB care cascade [1]. We used articles identified for that review that evaluated Gap 5 in the TB care cascade but that also reported factors and reasons associated with post-treatment TB recurrence or mortality. For that review, a medical librarian searched PubMed, Embase, and Web of Science for studies published between January 1, 2000 and October 9, 2015, without language restrictions, using search terms and related variants for “tuberculosis”, “India”, and “recurrence” (Table A). In addition, we carried out electronic searches of key Indian journals that were not indexed in the above databases for that entire time window: the Indian Journal of Tuberculosis, Lung India, the Indian Journal of Chest and Allied Sciences, the India Journal of Public Health, and the Indian Journal of Community Medicine. Additional studies were identified by searching the reference lists of the primary studies and relevant review articles. We screened all identified studies from this previous review for potential inclusion in our current review; however, different data were extracted from studies that met inclusion criteria.

To update our review, we conducted second refresher search using the same search terms for October 10, 2015 to October 1, 2019. We did not repeat hand searches of the Indian journals listed above, because all of these journals had been indexed in PubMed prior to the time period of this more recent search. Due to the extensive time required to extract data from the articles identified for this systematic review, we performed a third refresher search using the same search terms for October 2, 2019 to May 17, 2021. Finally, additional studies were identified by looking through the reference lists of the included primary studies and relevant review articles that were identified by the searches and by outreach to experts in the field, including co-authors of this review.

###### Table A. Search strategy to identify studies evaluating recurrence-free survival after completion of tuberculosis treatment (Gap 5)

| Terms for tuberculosis: | “tuberculosis”[Mesh] OR *Mycobacterium tuberculosis*[tiab] OR TB[tiab] OR MDRTB[tiab] OR XDRTB[tiab] |
| --- | --- |
| Terms for India: | “India”[Mesh] OR India[tiab] OR India[ad] OR Indian[tiab] OR Indians[tiab] |
| Terms for recurrence: | recurrence[Mesh] OR relapse[tiab] OR recrudescence[tiab] OR reinfection[tiab] OR follow-up[tiab] OR failure[tiab] |

##### Inclusion and exclusion criteria

We applied the following criteria for inclusion and exclusion of studies for this systematic review.

*Inclusion criteria* included the following:

(1) Studies that followed patients starting either from TB treatment initiation or treatment completion to evaluate outcomes of TB recurrence and/or mortality in the immediate few months to years after completing TB treatment.
(2) Studies also had to have assessed reasons why patient may have experienced post-treatment TB recurrence or mortality. These studies could have compared characteristics of those who did or did not experience these unfavorable post-treatment outcomes (e.g., using regression analyses) or conducted structured interviews with patients who experienced these outcomes to understand contributing reasons.

*Exclusion criteria* included the following:

(1) Studies that described only the proportions of TB patients who experienced TB recurrence or died after completing treatment without describing reasons why these outcomes occurred.
(2) Any studies of TB recurrence during a clinical trial as these studies tend to be testing out new interventions or medications that may not be standard of care within India’s NTEP, and these study scenarios are designed to retain patients and therefore may not represent care under routine programmatic conditions.
(3) Studies with data collected prior to the year 2000, as India’s Revised National TB Control Programme (now called the NTEP) did not achieve nationwide coverage until the early 2000s.
(4) Studies only containing qualitative data evaluating reasons for unfavorable post-treatment outcomes. Findings from studies containing qualitative data will be reported in a separate paper.

##### Study selection

Each citation identified by the search was independently assessed by at least two reviewers (among TJ, DJ, AG, DV, MLS and KP) for their eligibility at the title and abstract evaluation stage and again subsequently at the full text evaluation stage (Fig A). Disagreements between the two reviewers were resolved by discussion or, if necessary, through consultation of a third reviewer (RS). Independent selection of articles at the title and abstract and full text stages was conducted using Covidence software (Veritas Health Innovations, Melbourne, Australia); however, quality assessment and extraction of study findings was conducted using an Excel spreadsheet.

###### Fig A. PRISMA flowchart: study selection for the systematic review of barriers to achieving recurrence-free survival after completion of tuberculosis treatment (Gap 5)

**Search 1: January 1, 2000 to October 9, 2015**

**Search 3: October 2, 2019 to May 17, 2021**

**Search 2: October 10, 2015 to October 1, 2019**

Reports identified from search of PubMed, Embase, and Web of Science and hand search of relevant journals (n=4,125), of which 1,578 were duplicates.

Duplicates removed before screening

(n = 63)

Records identified from search of PubMed, Embase, and Web of Science (n = 468)

Duplicate records removed before screening (n = 2)

Records identified from search of PubMed, Embase, and Web of Science (n = 849)

**Identification**

Records excluded after title and abstract screen

(n = 367)

Records excluded after title and abstract screen

(n = 832)

Records that underwent screening of title and abstract

(n = 405)

Records that underwent screening of title and abstract

(n = 847)

Total studies included in the systematic review (n = 16)

- Unique (n=10)
- Multiple gaps (n=6)

Reports shortlisted after removal of duplicates (n=2,547) of which 2,517 were excluded after title and abstract review.

Records not retrieved (n = 0)

Records for which full text articles were retrieved (n = 34)

Reports not retrieved

(n = 0)

Records for which full text articles were retrieved (n = 15)

**Screening**

Reports eligible for full text review (n=30), of which 20 were excluded.

Reports excluded after full text review

(n = 30), for the following reasons:

Wrong outcomes evaluated (n = 26)

Wrong study design

(n = 2)

Wrong setting (n = 2)

Reports assessed for eligibility via full text review (n = 34)

Reports excluded after full text review (n = 13), for the following reasons:

Wrong outcomes evaluated (n = 9)

Wrong patient population (n = 2)

Abstract (n = 2)

Reports assessed for eligibility via full text review (n = 15)

New studies included in the systematic review from the 2015 to 2019 search (n = 2)

Studies included in previous care cascade study with relevant data on reasons for not seeking care (n = 10)

New studies included in the systematic review from the 2019 to 2021 search (n = 4)

**Included**

Studies identified by outreach to experts (n = 0)

Studies identified by reviewing references of other articles (n = 0)

##### Quality assessment of quantitative studies

In our previous systematic review, we developed quality criteria relevant to studies focused on identifying patients who experienced post-treatment TB recurrence or mortality (Table B), because there were no existing standardized guidelines for assessing the quality of these types of studies. Although we extracted different data from these studies in the current systematic review, we used the same quality criteria for studies that included relevant quantitative information, because these criteria are still relevant for assessing the overall quality of these types of observational studies.

We classified the population-based sampling strategy based on whether it was comprehensive, random, or convenience. Studies using convenience sampling were excluded from analysis. We considered studies that evaluated patient outcomes for more than 100 patients or at multiple TB treatment centers to be higher in quality than single-center studies, since they may better represent outcomes in a local area. Prospective cohort studies and studies using active surveillance to identify TB recurrence were rated as being higher in quality than retrospective cohort studies and those employing passive surveillance to identify cases of TB recurrence. Studies that screened all symptomatic patients with cartridge-based nucleic acid amplification testing (CBNAAT) or mycobacterial culture to diagnose TB recurrence were rated as being higher in quality than studies that diagnosed recurrence using only sputum microscopy or clinical diagnosis alone, because studies using CBNAAT or mycobacterial culture have greater sensitivity for detecting TB recurrence.

###### Table B. Criteria for assessing the quality of studies evaluating recurrence-free survival after completion of tuberculosis treatment (Gap 5)

| **Criterion** | **Quality level** |
| --- | --- |
| **Sampling strategy** | |
| Random or comprehensive sampling of patients | High |
| Random or comprehensive sampling of patients but <50% of cohort retained in follow-up | Low (exclude from analysis) |
| Convenience sampling | Low (exclude from analysis) |
| **Sample size and distribution** | |
| >1 treatment center and 100+ patients | High |
| Single treatment center study with 100+ patients | Medium |
| <100 patients | Low |
| **Cohort design and surveillance for recurrence** | |
| Active surveillance (prospective) | High |
| Passive surveillance (generally retrospective) | Medium to Low |
| **Method for diagnosing recurrence** | |
| Sputum smear, mycobacterial culture, or CBNAAT performed uniformly during one or multiple post-treatment follow-up visits | High |
| Sputum smear, mycobacterial culture, or CBNAAT only performed if patient returns to the clinic or is found to have persistent symptoms | Medium |
| Clinical diagnosis alone without microbiological tests or approach for diagnosing recurrence not reported | Low |

##### Data extraction and analysis

Two reviewers (among TJ, DJ, AG, DV, MLS, and KP) independently extracted data from each included study into a structured data extraction form. Disagreements were resolved by discussion or, if necessary, by consulting a third reviewer (RS). From each study, we extracted information on the study design, location, setting (i.e., urban versus rural), sample size, and variables of interest (Table C). For odds ratios, relative risk ratios, hazard ratios, beta-coefficients, and proportions, we also extracted information on 95% confidence intervals (95% CIs) where available; if 95% CIs were not reported, we calculated these from the data provided, if possible.

For some variables, we also changed the reference group as needed for consistency of reporting across studies. For example, because most studies compared men to the reference group of women, we “flipped” effect estimates and confidence intervals for studies that presented men as the reference group. This allowed us to consistently present women as the reference group for findings regarding gender.

We reported all unadjusted and adjusted effect estimates, regardless of statistical significance, organized by study (Table D). For the main manuscript and Forest plots, we restricted ourselves to presenting statistically significant adjusted effect estimates from multivariable analyses, as these may represent more meaningful associations from higher-quality analyses. After extracting this subset of findings, we organized findings into categories using our framework of demand- and supply-side factors (main manuscript, Table 1). To visualize quantitative findings, we generated Forest plots of effect estimates (odds ratios, risk ratios, hazard ratios, beta-coefficients, and proportions) using Stata version 16.1 (College Station, TX, USA). We did not conduct a meta-analysis of data, because we extracted findings regarding a diverse set of variables from every study.

###### Table C. Characteristics of the included studies that evaluated barriers to achieving recurrence-free survival after completion of tuberculosis treatment—specifically outcomes of post-treatment tuberculosis recurrence or mortality (Gap 5)

| **Citation** (year) | **Location** | **Urban, rural, both, or unknown** | **Type of population** | **Single or multiple designated microscopy centers (DMCs)** | **Sample size**  (Overall sample studies to evaluate these outcomes) | **Time period of evaluation** (timepoint of enrollment and follow-up period) | **Post-treatment surveillance and methodology for assessing recurrence** | **Type of outcomes assessed** | **Type of findings included in the study** (sample size for each type of analysis) |
| --- | --- | --- | --- | --- | --- | --- | --- | --- | --- |
| Studies with TB recurrence as an outcome or part of a composite outcome |  |  |  |  |  |  |  |  |  |
| Cox (2021) [2] | Maharashtra and Tamil Nadu | Both | Adult men with drug-susceptible TB in public sector treatment | Multiple | 751 | TB treatment initiation until up to 18 months after treatment completion | Active surveillance for recurrence with follow-up visits every 6 months after treatment completion with possible microbiological evaluation* | (1) Post-treatment recurrence; (2) composite outcome of treatment failure, on-treatment and post-treatment mortality, and post-treatment recurrence | Incidence rate ratio regression (N=751) |
| Dandekar (2014) [3] | Maharashtra | Urban | Patients of all ages (including children) with presumed drug-susceptible TB in public sector treatment | Multiple | 304 | TB treatment initiation until up to 24 months after treatment completion | Passive surveillance,* though microbiological evaluation was conducted for patients with persistent symptoms at a follow-up visit 24 months after treatment completion* | Treatment failure, on-treatment and post-treatment mortality, and post-treatment recurrence as a composite outcome. | Logistic regression (N=304) |
| Gupte (2019) [4] | Maharashtra and Tamil Nadu | Both | Adults with drug-susceptible TB in public sector treatment | Multiple | 455 | TB treatment initiation until up to 18 months after treatment completion | Active surveillance for recurrence with follow-up visits every 6 months after treatment completion with possible microbiological evaluation* | (1) Post-treatment recurrence; (2) composite outcome of treatment failure, on-treatment and post-treatment mortality, and post-treatment recurrence | Incident rate ratio regression (N=455) |
| Huddart (2021) [5] | Bihar | Both | Adults with presumed drug-susceptible TB treated in the private sector with support of a private provider interface agency (PPIA) | Multiple | 4000 | TB treatment initiation until up to a maximum of 5.5 years of follow-up time | Passive surveillance,* with outcome determined by a single follow-up phone survey and recurrence identified by self-reported reenrollment in TB treatment* | Post-treatment recurrence | Fine and Gray sub-distributional hazard regression model (N=4000; for 2240 the outcome was observed and for 1760 the outcome was unobserved and adjusted for using inverse probability selection weighting) |
| Lisha (2012) [6] | Kerala | Urban | Adults and adolescents (>14 years of age) with presumed drug-susceptible smear-positive pulmonary TB treated at a public sector tertiary care hospital | Single* | 224 | Tuberculosis treatment initiation until 5 years of follow-up | Passive surveillance,* with outcome determined by a single follow-up visit 5 years after treatment initiation that involved uniform sputum smear microscopy but mycobacterial culture only in select cases | Post-treatment recurrence | Logistic regression analyses (N=224) |
| Mahishale (2015) [7] | Karnataka | Both | Adults and adolescents (>15 years of age) with presumed drug-susceptible pulmonary TB treated in a public sector tertiary care hospital | Single* | 2350 | Tuberculosis treatment initiation until 2 years of follow-up | Active surveillance (prospective cohort study) but unclear how frequently follow-up was conducted and whether efforts were made to diagnosed recurrence outside of routine evaluations for recurrent symptoms* | Post-treatment recurrence | Cox proportional hazard regression model (N=2350) |
| Mave (2021) [8] | Maharashtra | Urban | Adults with drug-susceptible pulmonary TB treated in two public sector tertiary care hospitals | Multiple | 799 | Tuberculosis treatment initiation until 18 months of follow-up | Active surveillance for recurrence with follow-up visits every 6 months after treatment completion, but unclear whether microbiological evaluation was performed uniformly or for symptomatic patients during post-treatment follow-up* | Post-treatment recurrence | Logistic regression (N=799) |
| Ramachandran (2020) [9] | Tamil Nadu and Maharashtra | Urban | Adults with newly diagnosed pulmonary TB on Cat I ATT for ≥2 weeks under directly observed treatment | Multiple | 404 | Tuberculosis treatment completion with subsequent follow-up at ≥4 months | Active surveillance for recurrence with follow-up visit at ≥4 months after treatment completion with mycobacterial culture or sputum microscopy or symptom assessment of all participants performed with samples | (1) Post-treatment recurrence; (2) composite outcome of treatment failure, on-treatment and post-treatment mortality, and post-treatment recurrence | Poisson Regression Analysis (N=404) |
| Thomas (2005) [10] | Tamil Nadu | Rural | Adult patients with new smear-positive pulmonary TB, presumed to be drug-susceptible, who achieved cure | Multiple | 534 | Tuberculosis treatment completion (with confirmed cure) with 18 months of subsequent follow-up | Active surveillance for recurrence with follow-up visits every 6 months after treatment completion with mycobacterial culture of all participants performed with samples from each visit | Post-treatment recurrence | Logistic regression (N=503) |
| Tripathy (2011) [11] | Maharashtra | Urban | Adult patients with presumed drug-susceptible TB with and without HIV | Single* | 283 (121 with HIV and 162 without HIV) | Treatment initiation with 30 months of subsequent follow-up | Active surveillance for recurrence with follow-up every 3 months after treatment completion with uniform annual chest X-ray for all participants and sputum microscopy conducted for new symptoms* | Post-treatment recurrence | Logistic regression (N=283)^a^ |
| Vashishtha (2013) [12] | Delhi National Capital Region | Urban | Adult patients with presumed drug-susceptible TB with newly diagnosed HIV and without HIV at a large tertiary care hospital | Single* | 305 (150 with HIV and 155 without HIV) | Treatment initiation until 24 months after treatment completion | Active surveillance for recurrence with follow-up every 3 months after treatment completion with microbiological tests if appropriate for symptoms* | Post-treatment recurrence | Logistic regression (N=211)^a^ |
| Velayutham (2018) [13] | Tamil Nadu, Karnataka, Delhi, Maharashtra, Madhya Pradesh, and Kerala | Both | Adult patients with presumed drug-susceptible TB who achieved cure (confirmed with mycobacterial culture) at the end of TB treatment | Multiple | 1565 | Treatment initiation until 12 months after treatment completion | Active surveillance for recurrence with follow-up visits at 3 months, 6 months, and 12 months after treatment completion with sputum collected from all participants for microscopy and mycobacterial culture | Post-treatment recurrence | Relative risk regression (N=1108) |
| Studies with post-treatment mortality as an outcome (without evaluation of recurrence) |  |  |  |  |  |  |  |  |  |
| Cox (2021) [2] | Maharashtra and Tamil Nadu | Both | Adult men with drug-susceptible TB in public sector treatment | Multiple | 751 | TB treatment initiation until up to 18 months after treatment completion | Active post-treatment surveillance to assess for mortality | On-treatment and post-treatment mortality | Incidence rate ratio regression (N=751) |
| Gupte (2019) [4] | Maharashtra and Tamil Nadu | Both | Adult men with drug-susceptible TB in public sector treatment | Multiple | 455 | TB treatment initiation until up to 18 months after treatment completion | Active post-treatment surveillance to assess for mortality | On-treatment and post-treatment mortality | Incident rate ratio regression (N=455) |
| Huddart (2021) [5] | Bihar | Both | Adults with presumed drug-susceptible TB treated in the private sector with support of a private provider interface agency (PPIA) | Multiple | 4000 | TB treatment initiation until up to a maximum of 5.5 years of follow-up time | Passive surveillance* with outcome determined by a single follow-up phone survey | Post-treatment mortality | Cox proportional hazards regression model (N=4000; for 2240 the outcome was observed and for 1760 the outcome was unobserved and adjusted for using inverse probability selection weighting) |
| Kolappan (2006) [14] | Tamil Nadu | Urban | Adults and adolescents (age >14) with any form of TB (smear-positive pulmonary, smear-negative pulmonary, extrapulmonary) including new and retreatment patients started on treatment for presumed drug-susceptible TB | Multiple | 2674 | Treatment initiation until a minimum of 20 months of follow-up | Approach to surveillance was unclear.* At least one home visit was conducted with the patient or their family; however, the approach to subsequent follow-up to determine outcomes over time was not reported. | On-treatment and post-treatment mortality | Cox proportional hazards regression model (N=2674) |
| Kolappan (2008) [15] | Tamil Nadu | Rural | Adults and adolescents (age >14) with any form of TB (smear-positive pulmonary, smear-negative pulmonary, extrapulmonary) including new and retreatment patients started on treatment for presumed drug-susceptible TB | Multiple | 3388 | Treatment initiation with an average follow-up duration of 40 months | Approach to surveillance was unclear.* At least one home visit was conducted with the patient or their family; however, the approach to subsequent follow-up to determine outcomes over time was not reported. | On-treatment and post-treatment mortality | Cox proportional hazards regression model (N=3388) |
| Lisha (2012) [6] | Kerala | Urban | Adults and adolescents (>14 years of age) with presumed drug-susceptible smear-positive pulmonary TB treated at a public sector tertiary care hospital | Single* | 224 | Tuberculosis treatment initiation until 5 years of follow-up | Passive surveillance,* with outcome determined by a single attempt at outreach or follow-up visit 5 years after treatment initiation | Post-treatment mortality | Logistic regression analyses (N=224) |
| Mave (2021) [8] | Maharashtra | Urban | Adults with drug-susceptible pulmonary TB treated in two public sector tertiary care hospitals | Multiple | 799 | Tuberculosis treatment initiation until 18 months of follow-up | Active surveillance for mortality with follow-up visits every 6 months after treatment completion | (1) post-treatment mortality alone; and (2) composite outcome of on-treatment and post-treatment mortality (we do not report findings for the composite outcome as significant associations were driven by on-treatment mortality, which is reported in the Gap 4 analysis). | Logistic regression (N=799) |
| Sadacharam (2007) [16] | Tamil Nadu | Rural | General | Multiple | 988 | Tuberculosis treatment initiation until 24 to 36 months of follow-up | Passive surveillance,* with a single visit conducted around 24 to 36 months to determine the outcome | Post-treatment mortality | Logistic regression (N=988) |
| Sharma (2019) [17] | Gujarat | Urban | Adult patients treated for new, presumed drug-susceptible TB | Multiple | 365 | Tuberculosis cure until 12 to 36 months after completion of treatment | Passive surveillance,* with a single visit conducted between 12 to 36 months after treatment completion to determine the outcome | Post-treatment mortality | Logistic regression (N=191)^a^ |
| Tripathy (2011) [11] | Maharashtra | Urban | Adult patients with presumed drug-susceptible TB with and without HIV | Single* | 283 (121 with HIV and 162 without HIV) | Treatment initiation with 30 months of subsequent follow-up | Active surveillance for recurrence with follow-up every 3 months after treatment completion to determine post-treatment mortality | On-treatment and post-treatment mortality | Logistic regression (N=283)^a^ |
| Vashishtha (2013) [12] | Delhi National Capital Region | Urban | Adult patients with presumed drug-susceptible TB with newly diagnosed HIV and without HIV at a large tertiary care hospital | Single* | 305 (150 with HIV and 155 without HIV) | Treatment initiation until 24 months after treatment completion | Active surveillance for recurrence with follow-up every 3 months after treatment completion to determine post-treatment mortality | Post-treatment mortality | Logistic regression (N=211)^a^ |

TB, tuberculosis.

*Medium or low quality for this indicator

^a^Unadjusted odds ratios and/or p-values were estimated by the systematic review team from the raw data, as these were not provided in the original study.

###### Table D. Factors associated with patients not achieving recurrence-free survival after completion of tuberculosis treatment-- specifically outcomes of post-treatment tuberculosis recurrence or mortality (Gap 5)

| Study and specific outcome | Exposure/Independent variable | Unadjusted Effect Estimate (95% Confidence Interval) | P-value | Adjusted Effect Estimate (95% Confidence Interval) | P-value |
| --- | --- | --- | --- | --- | --- |
| Studies with TB recurrence as an outcome or part of a composite outcome |  |  |  |  |  |
| Cox, 2021 (Maharashtra and Tamil Nadu)  *Outcome: TB recurrence as a single outcome up to 18 months after treatment completion* [2] |  | Values below are incidence rate ratios |  | Values below are adjusted incidence rate ratios |  |
|  | **Ever had a drink of alcohol** |  |  |  |  |
|  | No | Ref |  | Ref |  |
|  | Yes | 1.11 (0.65-1.87) | 0.71 | 1.53 (0.86-2.73) | 0.15 |
|  | **Unhealthy alcohol use** (AUDIT-C >=4)^1^ |  |  |  |  |
|  | No | Ref |  | Ref |  |
|  | Yes | 1.39 (0.83-2.34) | 0.21 | 1.68 (0.94-3.00) | 0.08 |
|  | **AUDIT-C**  (continuous measure) |  |  |  |  |
|  | No | Ref |  | Ref |  |
|  | Yes | 1.05 (0.98-1.13) | 0.13 | 1.09 (1.01-1.17)* | 0.03* |
|  | **BMI and alcohol use** |  |  |  |  |
|  | Not underweight (BMI >=18.5) and not severe alcohol use (AUDIT-C <4) | Not reported | Not reported | Ref |  |
|  | Not underweight (BMI >=18.5) but with severe alcohol use (AUDIT-C >=4) | Not reported | Not reported | 1.98 (0.80-4.94) | 0.14 |
|  | Underweight (BMI <18.5) but not severe alcohol use (AUDIT-C <4) | Not reported | Not reported | 1.49 (0.68-3.26) | 0.32 |
|  | Underweight (BMI <18.5) and severe alcohol use (BMI >=4) | Not reported | Not reported | 2.25 (1.02-4.96)* | 0.04* |
|  | Not severely underweight (BMI >=16.5) and not severe alcohol use (AUDIT-C <4) | Not reported | Not reported | Ref |  |
|  | Not severely underweight (BMI >=16.5) but with severe alcohol use (AUDIT-C >=4) | Not reported | Not reported | 1.76 (0.87-3.56) | 0.12 |
|  | Severely Underweight (BMI <16.5) but not severe alcohol use (AUDIT-C <4) | Not reported | Not reported | 1.51 (0.69-3.29) | 0.31 |
|  | Severely Underweight (BMI <16.5) and severe alcohol use (AUDIT-C >=4) | Not reported | Not reported | 2.15 (0.96-4.79) | 0.06 |
| Cox, 2021 (Maharashtra and Tamil Nadu)  *Outcome: Composite outcome of treatment failure, TB recurrence, or death up to 18 months after treatment completion* [2] |  | Values below are incidence rate ratios |  | Values below are adjusted incidence rate ratios |  |
|  | **Ever had a drink of alcohol** |  |  |  |  |
|  | No | Ref |  | Ref |  |
|  | Yes | 1.87 (1.30-2.69)* | <0.001 | 1.62 (1.14-2.31)* | 0.01 |
|  | **Unhealthy alcohol use** (AUDIT-C >=4) |  |  |  |  |
|  | No | Ref |  | Ref |  |
|  | Yes | 1.99 (1.40-2.81)* | <0.001 | 1.47 (1.05-2.06)* | 0.03 |
|  | **AUDIT-C**  (continuous measure) |  |  |  |  |
|  | No | Ref |  | Ref |  |
|  | Yes | 1.10 (1.05-1.15)* | <0.001 | 1.06 (1.01-1.10)* | 0.01 |
|  | **BMI and alcohol use** |  |  |  |  |
|  | Not underweight (BMI >=18.5) and not severe alcohol use (AUDIT-C <4) | Not reported | Not reported | Ref |  |
|  | Not underweight (BMI >=18.5) but with severe alcohol use (AUDIT-C >=4) | Not reported | Not reported | 1.12 (0.64-1.96) | 0.69 |
|  | Underweight (BMI <18.5) but not severe alcohol use (AUDIT-C <4) | Not reported | Not reported | 1.18 (0.75-1.86) | 0.48 |
|  | Underweight (BMI <18.5) and severe alcohol use (BMI >=4) | Not reported | Not reported | 2.22 (1.44-.344)* | <0.001 |
|  | Not severely underweight (BMI >=16.5) and not severe alcohol use (AUDIT-C <4) | Not reported | Not reported | Ref |  |
|  | Not severely underweight (BMI >=16.5) but with severe alcohol use (AUDIT-C >=4) | Not reported | Not reported | 1.20 (0.78-1.84) | 0.40 |
|  | Severely Underweight (BMI <16.5) but not severe alcohol use (AUDIT-C <4) | Not reported | Not reported | 1.20 (0.74-1.97) | 0.46 |
|  | Severely Underweight (BMI <16.5) and severe alcohol use (AUDIT-C >=4) | Not reported | Not reported | 2.78 (1.78-4.34)* | <0.001 |
| Dandekar, 2014 (Maharashtra)  *Outcome: Composite outcome of treatment failure, on-treatment and post-treatment mortality, and post-treatment recurrence 24 months after treatment completion* [3] |  | Values below are odds ratios |  |  |  |
|  | **Gender** |  |  |  |  |
|  | Female | Ref |  |  |  |
|  | Male | 2.39 (1.21-4.76)* | 0.01* |  |  |
|  | **Age (Years)** |  |  |  |  |
|  | <35 | Ref |  |  |  |
|  | >35 | 7.18 (3.50-14.72)* | <0.0001* |  |  |
|  | **Religion** |  |  |  |  |
|  | Other than Hindu | Ref |  |  |  |
|  | Hindu | 1.15 (0.63-2.11) | 0.65 |  |  |
|  | **Occupation** |  |  |  |  |
|  | Employed | Ref |  |  |  |
|  | Unemployed | 1.45 (0.78-2.69) | 0.23 |  |  |
|  | **Education** |  |  |  |  |
|  | Literate | Ref |  |  |  |
|  | Illiterate | 2.97 (1.60-5.51)* | 0.0006* |  |  |
|  | **Type of TB lesion** |  |  |  |  |
|  | Extrapulmonary | Ref |  |  |  |
|  | Pulmonary | 1.69 (0.82-3.47) | 0.15 |  |  |
|  | **Type of TB case** |  |  |  |  |
|  | New case | Ref |  |  |  |
|  | Retreatment | 1.29 (0.63-2.64) | 0.49 |  |  |
|  | **Comorbidities** |  |  |  |  |
|  | Without comorbidities | Ref |  |  |  |
|  | With comorbidities including HIV, liver disease, diabetes, and hypertension | 4.31 (2.01-9.24)* | 0.0002* |  |  |
|  | **Type of family** |  |  |  |  |
|  | Nuclear | Ref |  |  |  |
|  | Other | 1.43 (0.72-2.84) | 0.3 |  |  |
| Gupte, 2019 (Tamil Nadu and Maharashtra)  *Outcome: TB recurrence as a single outcome up to 18 months after treatment completion* [4] |  | Values below are incident rate ratios |  | Values below are adjusted incident rate ratios |  |
|  | **Enrollment SGRQ score** (per every 4-point increase) |  |  |  |  |
|  |  | 0.98 (0.89-1.08) | 0.81 | 1.02 (0.92-1.13) | 0.63 |
|  | **6-month SGRQ score** (per every 4-point increase) |  |  |  |  |
|  |  | 1.03 (0.88-1.21) | 0.66 | 1.02 (0.88-1.19) | 0.73 |
|  | **Time-updated SGRQ score** (per every 4-point increase) |  |  |  |  |
|  |  | 1.17 (1.06-1.29)* | 0.002* | 1.15 (1.04-1.27)* | 0.004* |
| Gupte, 2019 (Tamil Nadu and Maharashtra)  *Outcome: Composite outcome of treatment failure, on-treatment and post-treatment mortality, and post-treatment recurrence up to 18 months after treatment completion* [4] |  | Values below are incident rate ratios |  | Values below are adjusted incident rate ratios |  |
|  | **Enrollment SGRQ score** (per every 4-point increase) |  |  |  |  |
|  |  | 1.01 (0.96-1.06) | 0.6 | 1.01 (0.96-1.07) | 0.56 |
|  | **Time-updated SGRQ score** (per every 4-point increase) |  |  |  |  |
|  |  | 1.11 (1.04-1.18)* | 0.001* | 1.11 (1.03-1.19)* | 0.002* |
| Huddart, 2021 (Bihar)  *Outcome: TB recurrence as a single outcome up to a maximum of 5.5 years after treatment initiation* [5] |  | Values below are unweighted hazard ratios |  | Values below are weighted hazard ratios |  |
|  | **Gender** |  |  |  |  |
|  | Male | Ref |  | Ref |  |
|  | Female | 1.02 (0.63-1.59) | Not reported | 1.03 (0.61-1.67) | Not reported |
|  | **Age (per year)** | 0.99 (0.98-1.01) | Not reported | 1.00 (0.99-1.01) | Not reported |
|  | **Residence** |  |  |  |  |
|  | Out of Patna | Ref |  | Ref |  |
|  | Rural Patna | 1.97 (0.93-3.83) | Not reported | 1.82 (0.86-3.81) | Not reported |
|  | Urban Patna | 0.96 (0.62-2.02) | Not reported | 0.91 (0.55-1.57) | Not reported |
|  | **Slum residence** |  |  |  |  |
|  | Non-slum |  |  |  |  |
|  | Slum | 0.71 (0.45-1.10) | Not reported | 0.68 (0.40-1.14) | Not reported |
|  | **Treatment category** |  |  |  |  |
|  | New |  |  |  |  |
|  | Retreatment | 1.41 (0.61-2.54) | Not reported | 1.27 (0.54-2.57) | Not reported |
|  | **Type of TB** |  |  |  |  |
|  | Pulmonary |  |  |  |  |
|  | Extrapulmonary | 0.96 (0.56-1.68) | Not reported | 1.01 (0.58-1.63) | Not reported |
|  | **Adherence** |  |  |  |  |
|  | Good adherence |  |  |  |  |
|  | <1month adherence | 0.86 (0.33-1.63) | Not reported | 0.71 (0.23-1.42) | Not reported |
|  | Poor adherence for 80% of doses | 1.14 (0.72-1.89) | Not reported | 1.07 (0.64-1.80) | Not reported |
|  | **Months of treatment** | 1.05 (0.97-1.09) | Not reported | 1.05 (0.97-1.10) | Not reported |
| Lisha, 2012 (Kerala)  *Outcome:* *TB recurrence as a single outcome up to 5 years after treatment completion* [6] |  | Note logistic regression was used but odds ratios were not reported | Values below are p-values on univariate analysis |  | Values below are p-values on multivariate analysis |
|  | **Gender** |  |  |  |  |
|  | Female | Ref |  |  |  |
|  | Male | Not reported | 0.38 |  |  |
|  | **Age (Years)** |  |  |  |  |
|  | <=45 | Ref |  |  |  |
|  | >45 | Not reported | 0.94 |  |  |
|  | **Socioeconomic Status** |  |  |  |  |
|  | High socioeconomic status | Ref |  |  |  |
|  | Low socioeconomic status | Not reported | 0.75 |  |  |
|  | **Current symptomatic** |  |  |  |  |
|  | No | Ref |  | Ref |  |
|  | Yes | Not reported | 0.04* | Not reported | 0.031* |
|  | **Fever at initial presentation** |  |  |  |  |
|  | No | Ref |  | Ref |  |
|  | Yes | Not reported | 0.007* | Not reported | 0.79 |
|  | **Diabetes mellitus** |  |  |  |  |
|  | No | Ref |  |  |  |
|  | Yes | Not reported | 0.99 |  |  |
|  | **COPD** |  |  |  |  |
|  | No | Ref |  |  |  |
|  | Yes | Not reported | 0.15 |  |  |
|  | **Addictions** |  |  |  |  |
|  | No | Ref |  |  |  |
|  | Yes | Not reported | <0.001* |  |  |
|  | **Smoking** |  |  |  |  |
|  | No | Ref |  |  |  |
|  | Yes | 1.60 (0.40-6.35) | 0.5 |  |  |
| Mahishale, 2015 (Karnataka)  *Outcome: TB recurrence as a single outcome up to 2 years after treatment initiation* [7] |  | Values below are odds ratios |  | Values below are adjusted odds ratios |  |
|  | **Smoking status and treatment success** |  |  |  |  |
|  | Never smokers | Ref |  | Ref |  |
|  | Ex-smokers | Not reported | Not reported | 1.43 (1.06-1.61)* | <0.001* |
|  | Current smokers | Not reported | Not reported | 1.68 (1.46-1.98)* | <0.001* |
| Mave, 2021 (Maharashtra)  *Outcome: TB recurrence as a single outcome with 18-month follow-up after treatment initiation* [8] |  | Values below are hazard ratios |  | Values below are adjusted hazard ratios |  |
|  | **Diabetes** |  |  |  |  |
|  | Patients with TB only | Ref |  | Ref |  |
|  | Patients with TB and diabetes mellitus | 0.62 (0.30-1.27) | 0.19 | 0.73 (0.31-1.70) | 0.46 |
|  | **Metformin usage among TB patients with diabetes** |  |  |  |  |
|  | No |  |  | Ref |  |
|  | Yes |  |  | 0.18 (0.04-0.89)* | Not reported |
| Mave, 2021 (Maharashtra)  *Outcome: Composite outcome of treatment failure, TB recurrence, and all-cause mortality with 18-month follow-up after treatment initiation* [8] |  | Values are relative risk ratios |  | Values are adjusted relative risk ratios |  |
|  | **Gender** |  |  |  |  |
|  | Female | Ref |  | Ref |  |
|  | Male | 1.86 (1.29-2.67) | 0.001* | 1.16 (0.74-1.80) | 0.52 |
|  | **Age** |  |  |  |  |
|  | <25 | Ref |  | Ref |  |
|  | 25-40 | 1.16 (0.78-1.72) | 0.45 | 0.94 (0.61-1.44) | 0.77 |
|  | >40 | 1.37 (0.93-2.01) | 0.11 | 1.30 (0.82-2.05) | 0.26 |
|  | **Household income** |  |  |  |  |
|  | >10,000 | Ref |  | Ref |  |
|  | <10,000 | 1.68 (1.18-2.39) | 0.004* | 1.39 (0.96-2.01) | 0.08 |
|  | **Anemia** |  |  |  |  |
|  | No | Ref |  |  |  |
|  | Yes | 0.77 (0.48-1.23) | 0.27 | Not included |  |
|  | **Diabetes** |  |  |  |  |
|  | Patients with TB only | Ref |  | Ref |  |
|  | Patients with TB and diabetes mellitus | 1.01 (0.71-1.42) | >0.95 | 1.13 (0.75-1.70) | 0.56 |
|  | **HBA1c (per every one-unit increase)** | 0.94 (0.87-1.01) | 0.10 | 0.96 (0.88-1.04) | 0.31 |
|  | **Smoking** |  |  |  |  |
|  | Non-smokers | Ref |  | Ref |  |
|  | Smokers | 1.63 (1.14-2.31) | 0.007* | 1.07 (0.73-1.58) | 0.73 |
|  | **Alcohol use** |  |  |  |  |
|  | No | Ref |  | Ref |  |
|  | Yes | 2.42 (1.78-3.29) | <0.001* | 1.97 (1.34-2.89) | <0.001* |
|  | **Smear grade** |  |  |  |  |
|  | Negative | Ref |  | Ref |  |
|  | 1+ | 1.36 (0.90-2.05) | 0.14 | 1.18 (0.77-1.80) | 0.45 |
|  | 2+ | 1.57 (1.00-2.47) | 0.05 | 1.28 (0.80-2.04) | 0.30 |
|  | 3+ | 1.62 (1.00-2.63) | 0.05 | 1.42 (0.86-2.34) | 0.17 |
|  | **Body mass index** |  |  |  |  |
|  | Normal | Ref |  | Ref |  |
|  | Underweight | 1.77 (1.22-2.57) | 0.003* | 1.60 (1.07-2.39) | 0.02* |
|  | Overweight/Obese | 1.13 (0.48-2.68) | 0.78 | 0.96 (0.37-2.48) | 0.93 |
|  | **Cavity on X-ray** |  |  |  |  |
|  | Absent | Ref |  |  |  |
|  | Present | 1.38 (0.98-1.96) | 0.07 | Not included |  |
|  | **TB dosing** |  |  |  |  |
|  | Alternate days | Ref |  | Ref |  |
|  | Daily | 0.64 (0.42-0.99) | 0.07 | 0.68 (0.43-1.08) | 0.11 |
| Ramachandran, 2020 (Tamil Nadu and Maharashtra)  *Outcome: TB recurrence as a single outcome at ≥4 months after treatment completion* [9] |  |  |  |  |  |
|  |  |  |  | Values below are adjusted incident rate ratios |  |
|  | **Rifampicin^a^** |  |  |  |  |
|  | 1 unit decrease in drug concentration |  |  | 1.02 (0.91-1.14) | 0.75 |
|  | **Isoniazid^a^** |  |  |  |  |
|  | 1 unit decrease in drug concentration |  |  | 1.05 (0.94-1.17) | 0.37 |
|  | **Pyrazinamide^a^** |  |  |  |  |
|  | 1 unit decrease in drug concentration |  |  | 1.05 (1.01-1.11)* | 0.05* |
| Ramachandran, 2020 (Tamil Nadu and Maharashtra) *Outcome: Composite outcome of treatment failure, on-treatment and post-treatment mortality, and post-treatment recurrence* [9] |  |  |  | Values below are adjusted incident rate ratios |  |
|  | **Rifampicin^a^** |  |  |  |  |
|  | 1 unit decrease in drug concentration |  |  | 1.21 (1.01-1.47)* | 0.05* |
|  | **Isoniazid^a^** |  |  |  |  |
|  | 1 unit decrease in drug concentration |  |  | 1.03 (0.92-1.16) | 0.59 |
|  | **Pyrazinamide^a^** |  |  |  |  |
|  | 1 unit decrease in drug concentration |  |  | 1.27 (0.88-1.83) | 0.21 |
| Thomas, 2005 (Tamil Nadu)  *Outcome: TB recurrence as a single outcome up to 18 months after treatment completion* [10] |  | Values below are odds ratios |  | Values below are adjusted odds ratios |  |
|  | **Age** |  |  |  |  |
|  | <45 | Ref |  |  |  |
|  | >=45 | 1.5 (0.8-2.6) | 0.2 |  |  |
|  | **Gender** |  |  |  |  |
|  | Female | Ref |  |  |  |
|  | Male | 1.8 (0.8-3.9) | 0.1 |  |  |
|  | **Initial weight** |  |  |  |  |
|  | >=42 kg | Ref |  |  |  |
|  | <42 kg | 1.3 (0.7-2.3) | 0.4 |  |  |
|  | **Education** |  |  |  |  |
|  | Illiterate | Ref |  |  |  |
|  | Literate | 1.2 (0.6-2.1) | 0.7 |  |  |
|  | **Occupation** |  |  |  |  |
|  | Employed | Ref |  |  |  |
|  | Unemployed | 1.2 (0.7-2.3) | 0.5 |  |  |
|  | **Smear conversion at 2 months** |  |  |  |  |
|  | Yes | Ref |  |  |  |
|  | No | 1.1 (0.5-2.2) | 0.9 |  |  |
|  | **Initial smear grading** |  |  |  |  |
|  | Scanty/1+ | Ref |  |  |  |
|  | 2+/3+ | 1.0 (0.6-1.8) | 0.9 |  |  |
|  | **Drug sensitivity profile at 0 months** |  |  |  |  |
|  | Sensitive | Ref |  | Ref |  |
|  | Resistant to H and/or HR | 3.6 (1.5-8.5)* | <0.01* | 4.8 (2.0-11.6)* |  |
|  | **Drug regularity** |  |  |  |  |
|  | Regular | Ref |  | Ref |  |
|  | Irregular | 2.6 (1.5-4.7)* | <0.001* | 2.5 (1.4-4.6)* |  |
|  | **Smoking** |  |  |  |  |
|  | No | Ref |  | Ref |  |
|  | Yes | 2.8 (1.5-5.2)* | <0.001* | 3.1 (1.6-6.0)* |  |
|  | **Drinking (alcoholism)** |  |  |  |  |
|  | No | Ref |  |  |  |
|  | Yes | 2.3 (1.3-4.1)* | <0.01* |  |  |
| Tripathy, 2011^b^ (Maharashtra)  *Outcome: TB recurrence as a single outcome at 30 months after treatment initiation* [11] |  | Values below are odds ratios |  |  |  |
|  | **HIV seropositivity**^b^ |  |  |  |  |
|  | HIV Seronegative | Ref |  |  |  |
|  | HIV Seropositive | 0.74 (0.15-3.69) | 0.71 |  |  |
| Tripathy, 2011^b^ (Maharashtra)  *Outcome: Composite outcome of TB recurrence, on-treatment and post-treatment mortality, transfer out, and loss to follow-up at 30 months after treatment initiation* [11] |  | Values below are odds ratios |  |  |  |
|  | **HIV seropositivity** |  |  |  |  |
|  | HIV Seronegative | Ref |  |  |  |
|  | HIV Seropositive | 5.33 (3.17-8.98)* | <0.0001* |  |  |
| Vashishtha, 2013^b^ (Delhi)  *Outcome: TB recurrence as a single outcome at 24 months after treatment initiation* [12] |  | Values below are odds ratios |  |  |  |
|  | **HIV**^c^ |  |  |  |  |
|  | HIV-negative | Ref |  |  |  |
|  | HIV-positive | 4.82 (0.90-25.92) | 0.07 |  |  |
| Velayutham, 2018 (6 Indian States)  *Outcome: TB recurrence as a single outcome at 12 months after treatment completion* [13] |  | Values below are relative risk ratios |  | Values below are adjusted relative risk ratios |  |
|  | **Gender** |  |  |  |  |
|  | Female | Ref |  | Ref |  |
|  | Male | 2.23 (1.47-3.40)* | <0.001* | 2.43 (1.29-4.58)* | 0.006* |
|  | **Age** |  |  |  |  |
|  | 18-24 | Ref |  |  |  |
|  | 25-34 | 1.23 (0.75-2.02) | 0.39 |  |  |
|  | 35-44 | 1.31 (0.79-2.19) | 0.28 |  |  |
|  | 45-54 | 1.13 (0.68-1.88) | 0.61 |  |  |
|  | 55-64 | 0.79 (0.43-1.45) | 0.45 |  |  |
|  | >65 | 0.75 (0.35-1.61) | 0.47 |  |  |
|  | **Baseline sputum smear grade** |  |  |  |  |
|  | Scanty/1+ | Ref |  |  |  |
|  | 2+/3+ | 1.21 (0.88-1.66) | 0.23 |  |  |
|  | **Baseline sputum culture grade** |  |  |  |  |
|  | Cols/1+ | Ref |  |  |  |
|  | 2+/3+ | 1.05 (0.76-1.46) | 0.73 |  |  |
|  | **Baseline drug susceptibility test** |  |  |  |  |
|  | Sensitive | Ref |  |  |  |
|  | Resistant | 0.92 (0.48-1.76) | 0.81 |  |  |
|  | **Duration of Intensive Phase** |  |  |  |  |
|  | 2 months | Ref |  |  |  |
|  | 3 months | 0.73 (0.42-1.27) | 0.28 |  |  |
|  | **Duration of anti-TB treatment** |  |  |  |  |
|  | 24-28 weeks | Ref |  |  |  |
|  | >28 weeks | 1.85 (0.25-13.49) | 0.54 |  |  |
|  | **Weight gain from baseline to end of treatment in kg** |  |  |  |  |
|  | None | 1.02 (0.63-1.68) | 0.9 |  |  |
|  | 0-2 | 1.15 (0.75-1.75) | 0.53 |  |  |
|  | 2.01-6.0 | Ref |  |  |  |
|  | 6.01-10.0 | 0.96 (0.62-1.49) | 0.85 |  |  |
|  | 10.01-14.0 | 0.76 (0.35-1.65) | 0.48 |  |  |
|  | >14 | 0.61 (0.19-1.94) | 0.4 |  |  |
|  | **Missed doses in intensive phase of treatment** |  |  |  |  |
|  | None | Ref |  | Ref |  |
|  | 1 to 6 | 1.42 (0.91-2.22) | 0.12 | 0.77 (0.26-2.27) | 0.64 |
|  | 7 to 12 | 1.50 (0.55-4.05) | 0.43 | 0.99 (0.59-1.67) | 0.99 |
|  | >12 | 2.72 (1.00-7.36)* | 0.05* | 1.22 (0.73-2.05) | 0.44 |
|  | **Respiratory symptom at end of treatment** |  |  |  |  |
|  | Any symptom | Ref |  | Ref |  |
|  | No | 2.33 (0.95-5.7) | 0.06 | 2.43 (0.97-6.07) | 0.06 |
|  | **BMI** |  |  |  |  |
|  | <16 | 1.73 (1.13-2.66)* | 0.01* | 1.22 (0.73-2.05) | 0.44 |
|  | 16-18.4 | 1.42 (0.91-2.21) | 0.12 | 0.99 (0.59-1.67) | 0.99 |
|  | 18.5-22.9 | Ref |  | Ref |  |
|  | >23 | 0.99 (0.47-2.07) | 0.97 | 0.77 (0.26-2.27) | 0.64 |
|  | **Diabetes Mellitus** |  |  |  |  |
|  | Nondiabetic | Ref |  | Ref |  |
|  | Diabetic on anti-diabetic treatment | 0.65 (0.37-1.12) | 0.12 | 1.02 (0.48-2.20) | 0.94 |
|  | Diabetic not on anti-diabetic treatment | 0.45 (0.20-1.02) | 0.06 | 0.34 (0.12-1.12) | 0.07 |
|  | **HIV status** |  |  |  |  |
|  | Nonreactive | Ref |  |  |  |
|  | Reactive, on ART | 1.49 (0.55-4.01) | 0.43 |  |  |
|  | Reactive, not on ART | Indeterminate |  |  |  |
|  | **Taking alcohol during treatment/follow-up** |  |  |  |  |
|  | No | Ref |  | Ref |  |
|  | Yes | 1.46 (1.07-2.00)* | 0.02* | 1.06 (0.66-1.71) | 0.81 |
|  | **Smoking during treatment/follow-up** |  |  |  |  |
|  | No | Ref |  | Ref |  |
|  | Yes | 1.53 (1.12-2.10)* | 0.007* | 1.13 (0.70-1.84) | 0.61 |
| Studies with post-treatment mortality as an outcome (without evaluation of recurrence) |  |  |  |  |  |
| Cox, 2021 (Maharashtra and Tamil Nadu)  *Outcome: On-treatment and post-treatment all-cause mortality up to 18 months after treatment completion* [2] |  | Values below are incidence rate ratios |  | Values below are adjusted incidence rate ratios |  |
|  | **Ever had a drink of alcohol** |  |  |  |  |
|  | No | Ref |  | Ref |  |
|  | Yes | 2.83 (1.55-5.15) | 0.001* | 2.37 (1.24-4.55) | 0.01* |
|  | **Unhealthy alcohol use** (AUDIT-C >=4) |  |  |  |  |
|  | No | Ref |  | Ref |  |
|  | Yes | 2.88 (1.69-4.91) | <0.001* | 1.90 (1.08-3.34) | 0.03* |
|  | **AUDIT-C**  (continuous measure) |  |  |  |  |
|  | No | Ref |  | Ref |  |
|  | Yes | 1.14 (1.06-1.22) | <0.001* | 1.07 (1.00-1.14) | 0.05 |
|  | **BMI and alcohol use** |  |  |  |  |
|  | Not underweight (BMI >=18.5) and not severe alcohol use (AUDIT-C <4) | Not reported | Not reported | Ref |  |
|  | Not underweight (BMI >=18.5) but with severe alcohol use (AUDIT-C >=4) | Not reported | Not reported | 2.04 (0.77-5.42) | 0.15 |
|  | Underweight (BMI <18.5) but not severe alcohol use (AUDIT-C <4) | Not reported | Not reported | 2.15 (0.88-5.24) | 0.09 |
|  | Underweight (BMI <18.5) and severe alcohol use (BMI >=4) | Not reported | Not reported | 5.48 (2.19-13.72) | <0.001* |
|  | Not severely underweight (BMI >=16.5) and not severe alcohol use (AUDIT-C <4) | Not reported | Not reported | Ref |  |
|  | Not severely underweight (BMI >=16.5) but with severe alcohol use (AUDIT-C >=4) | Not reported | Not reported | 2.00 (0.93-4.34) | 0.08 |
|  | Severely Underweight (BMI <16.5) but not severe alcohol use (AUDIT-C <4) | Not reported | Not reported | 3.11 (1.28-7.57) | 0.01* |
|  | Severely Underweight (BMI <16.5) and severe alcohol use (AUDIT-C >=4) | Not reported | Not reported | 8.58 (3.73-19.70) | <0.001* |
| Gupte, 2019 (Tamil Nadu and Maharashtra)  *Outcome: On-treatment and post-treatment mortality up to 18 months after treatment completion* [4] |  | Values below are incident rate ratios |  | Values below are adjusted incident rate ratios |  |
|  | **Enrollment SGRQ^4^ scores** |  |  |  |  |
|  |  | 1.12 (1.01-1.24)* | 0.03* | 1.14 (1.02-1.28)* | 0.01* |
|  | **Time-updated SGRQ scores** |  |  |  |  |
|  |  | 1.12 (0.99-1.27) | 0.07 | 1.18 (1.04-1.33)* | 0.008* |
| Huddart, 2021 (Bihar)  *Outcome: Post-treatment case fatality as a single outcome up to a maximum of 5.5 years after treatment initiation* [5] |  | Values below are unweighted hazard ratios |  | Values below are weighted hazard ratios |  |
|  | **Gender** |  |  |  |  |
|  | Male | Ref |  | Ref |  |
|  | Female | 0.96 (0.49-1.74) | Not reported | 0.96 (0.52-1.79) | Not reported |
|  | **Age (per year)** | 1.06 (1.05-1.07)* | Not reported | 1.06 (1.05-1.08)* | Not reported |
|  | **Residence** |  |  |  |  |
|  | Out of Patna | Ref |  | Ref |  |
|  | Rural Patna | 0.39 (0.10-0.86)* | Not reported | 0.39 (0.11-0.88)* | Not reported |
|  | Urban Patna | 0.60 (0.33-1.10) | Not reported | 0.61 (0.32-1.14) | Not reported |
|  | **Slum residence** |  |  |  |  |
|  | Non-slum | Ref |  | Ref |  |
|  | Slum | 1.45 (0.82-2.73) | Not reported | 1.29 (0.69-2.50) | Not reported |
|  | **Treatment category** |  |  |  |  |
|  | New | Ref |  | Ref |  |
|  | Retreatment | 1.38 (0.39-3.07) | Not reported | 1.70 (0.51-3.68) | Not reported |
|  | **Type of TB** |  |  |  |  |
|  | Pulmonary | Ref |  | Ref |  |
|  | Extrapulmonary | 0.85 (0.35-1.61) | Not reported | 0.97 (0.44-1.88) | Not reported |
|  | **Months of treatment** | 1.00 (0.92-1.07) | Not reported | 0.98 (0.90-1.05) | Not reported |
| Kolappan, 2006 (Tamil Nadu)  *Outcome: On-treatment and post-treatment mortality up to 20 months from treatment initiation* [14] |  | Values below are hazard ratios |  | Values below are adjusted hazard ratios |  |
|  | **Age (Years)** |  |  |  |  |
|  | 15-44 | Ref |  | Ref |  |
|  | 45-59 | 1.8 | Not reported | 1.6 (1.0-2.6) | 0.061 |
|  | >=60 | 3 | Not reported | 3.0 (1.7-5.3)* | <0.001* |
|  | **Treatment outcome** |  |  |  |  |
|  | Cured | Ref |  | Ref |  |
|  | Loss to follow-up | 3.4 | Not reported | 3.3 (1.9-5.5)* | <0.001* |
|  | Failed | 6.8 | Not reported | 7.7 (3.4-17.5)* | <0.001* |
|  | Treatment completed | 0.5 | Not reported | 0.6 (0.3-1.0)* | 0.048* |
|  | **Behavioral factors** |  |  |  |  |
|  | Non-smoker and non-alcoholic | Ref |  | Ref |  |
|  | Smoker | 1.4 | Not reported | 1.1 (0.5-2.3) | 0.86 |
|  | Alcoholic | 1.8 | Not reported | 1.3 (0.5-3.4) | 0.58 |
|  | Smoker and alcoholic | 3.8 | Not reported | 2.9 (1.8-4.7)* | <0.001* |
| Kolappan, 2008 (Tamil Nadu)  *Outcome: On-treatment and post-treatment mortality for an average duration of 40 months of follow-up* [15] |  | Values below are hazard ratios |  | Values below are adjusted hazard ratios |  |
|  | **Age (Years)** |  |  |  |  |
|  | 15-44 | Ref |  | Ref |  |
|  | 45-59 | 2 |  | 1.9 (1.6-2.3)* | <0.001* |
|  | >=60 | 3 |  | 3.1 (2.6-3.8)* | <0.001* |
|  | **Treatment category** |  |  |  |  |
|  | Category 1 | Ref |  | Ref |  |
|  | Category 2 | 2.2 |  | 1.8 (1.3-2.3)* | <0.001* |
|  | Category 3 | 1 |  | 2.0 (1.4-2.7)* | <0.001* |
|  | **Treatment completion** |  |  |  |  |
|  | Yes | Ref |  | Ref |  |
|  | No | 4.8 |  | 6.4 (5.4-7.4)* | <0.001* |
|  | **Initial smear** |  |  |  |  |
|  | Negative | Ref |  | Ref |  |
|  | Scanty/positive | 0.7 |  | 0.4 (0.2-0.9)* | 0.032* |
|  | 1+ | 0.9 |  | 0.5 (0.3-0.6)* | <0.001* |
|  | 2+ | 1.1 |  | 0.6 (0.4-0.8)* | 0.002* |
|  | 3+ | 1.5 |  | 0.8 (0.6-1.0) | 0.08 |
|  | **Behavioral factors** |  |  |  |  |
|  | Non-smoker and non-alcoholic | Ref |  | Ref |  |
|  | Smoker | 1.7 |  | 1.0 (0.8-1.3) | 0.9 |
|  | Alcoholic | 1.2 |  | 0.8 (0.6-1.2) | 0.35 |
|  | Smoker and alcoholic | 1.9 |  | 1.2 (1.0-1.4)* | 0.03* |
| Lisha, 2012 (Kerala)  *Outcome:* *Post-treatment mortality as a single outcome up to 5 years after treatment completion* [6] |  | Values below are unadjusted odds ratio | Values below are p-values on univariate analysis |  | Values below are p-values on multivariate analysis |
|  | **Gender** |  |  |  |  |
|  | Female | Ref |  |  |  |
|  | Male | 2.64 (0.33-21.01) | 0.36 |  |  |
|  | **Age (Years)** |  |  |  |  |
|  | <=45 | Ref |  |  |  |
|  | >45 | Not reported | 0.11 |  |  |
|  | **Socioeconomic Status** |  |  |  |  |
|  | High socioeconomic status | Ref |  |  |  |
|  | Low socioeconomic status | Not reported | 0.63 |  |  |
|  | **Initial sputum smear** |  |  |  |  |
|  | No | Ref |  | Ref |  |
|  | >2+ | Not reported | 0.02* | Not reported | 0.06 |
|  | **Initial dyspnea** |  |  |  |  |
|  | No | Ref |  | Ref |  |
|  | Yes | Not reported | 0.05* | Not reported | 0.82 |
|  | **Relapse** |  |  |  |  |
|  | No | Ref |  |  |  |
|  | Yes | Not reported | 0.59 |  |  |
|  | **Weight loss at initial presentation** |  |  |  |  |
|  | No | Ref |  | Ref |  |
|  | Yes | Not reported | 0.04* | Not reported | 0.1 |
|  | **Diabetes mellitus** |  |  |  |  |
|  | No | Ref |  |  |  |
|  | Yes | Not reported | 0.86 |  |  |
|  | **Smoking** |  |  |  |  |
|  | No | Ref |  | Ref |  |
|  | Yes | Not reported | 0.02* | Not reported | 0.011* |
|  | **Smoking behavior** |  |  |  |  |
|  | No | Ref |  |  |  |
|  | Yes | Not reported | 0.001* |  |  |
| Mave, 2021 (Maharashtra)  *Outcome: Post-treatment mortality as a single outcome with 18-month follow-up after treatment initiation* [8] |  | Values below are hazard ratios |  | Values below are adjusted hazard ratios |  |
|  | **Diabetes** |  |  |  |  |
|  | TB only | Ref |  | Ref |  |
|  | TB-DM | 0.54 (0.22-1.28) | 0.16 | 0.58 (0.22-1.51) | 0.27 |
|  | New DM | 0.64 (0.15-2.69) | 0.55 | 0.42 (0.10-1.6) | 0.25 |
|  | Known DM | 0.50 (0.18-1.41) | 0.19 | 0.72 (0.23-2.22) | 0.57 |
|  | DM on metformin | 0.31 (0.07-1.29) | 0.11 | 0.47 (0.10-2.17) | 0.33 |
|  | DM – no metformin | 0.84 (0.30-2.39) | 0.75 | 0.65 (0.22-1.96) | 0.45 |
| Sadacharam, 2007 (Tamil Nadu)  *Outcome: Post-treatment mortality as a single outcome (follow-up for 2-3 years from treatment initiation)* [16] |  | Values below are odds ratios |  | Values below are adjusted odds ratios |  |
|  | **Age (Years)** |  |  |  |  |
|  | <=40 years | Ref |  | Ref |  |
|  | >40 | 3.2 (2.2-4.8)* | Not reported | 2.2 (1.3-3.7)* | Not reported |
|  | **Gender** |  |  |  |  |
|  | Female | Ref |  | Ref |  |
|  | Male | 2.7 (1.6-4.7)* | Not reported | 2.8 (1.4-5.8)* | Not reported |
|  | **Weight on admission** |  |  |  |  |
|  | >40 kg | Ref |  | Ref |  |
|  | <=40 kg | 2.1 (1.4-3.2)* | Not reported | 2.8 (1.7-4.6)* | Not reported |
|  | **Education** |  |  |  |  |
|  | Illiterate | Ref |  |  |  |
|  | Literate | 1.5 (1.0-2.2)* | Not reported |  |  |
|  | **Occupation** |  |  |  |  |
|  | Employed | Ref |  | Ref |  |
|  | Unemployed | 1.5 (1.0-2.2)* | Not reported | 2.0 (1.2-3.3)* | Not reported |
|  | **Sensitive on admission** |  |  |  |  |
|  | Sensitive | Ref |  |  |  |
|  | Resistant | 2.8 (1.7-4.4)* | Not reported |  |  |
|  | **Type of Patients** |  |  |  |  |
|  | New | Ref |  |  |  |
|  | Old | 2.6 (1.7-3.8)* | Not reported |  |  |
|  | **Outcome** |  |  |  |  |
|  | Success | Ref |  | Ref |  |
|  | Loss to follow-up/Failure | 10.9 (7.2-16.4)* | Not reported | 10.5 (6.4-17.1)* | Not reported |
|  | **Smoking** |  |  |  |  |
|  | No | Ref |  |  |  |
|  | Yes | 2.5 (1.7-3.7)* | Not reported |  |  |
|  | **Drinking** |  |  |  |  |
|  | No | Ref |  |  |  |
|  | Yes | 1.7 (1.2-2.5)* | Not reported |  |  |
|  | **First Action** |  |  |  |  |
|  | Private/Others | Ref |  |  |  |
|  | Government | 1.5 (1.0-2.3)* | Not reported |  |  |
| Sharma, 2019^b^ (Gujarat)  *Outcome: Post-treatment mortality as a single outcome at 1.5-5 years after treatment completion* [17] |  | Values below are odds ratios |  |  |  |
|  | **Gender** |  |  |  |  |
|  | Female | Ref |  |  |  |
|  | Male | 1.79 (0.79-4.03) | 0.16 |  |  |
|  | **Age** |  |  |  |  |
|  | 18-25 | Ref |  |  |  |
|  | 26-45 | 4.64 (1.32-16.22)* | 0.02* |  |  |
|  | 46-60 | 4.58 (1.10-19.10)* | 0.04* |  |  |
|  | >61 | 2.62 (0.49-14.02) | 0.26 |  |  |
| Tripathy, 2011^b^ (Maharashtra)  *Outcome: Post-treatment mortality as a single outcome at 30 months after treatment initiation* [11] |  | Values below are odds ratios |  |  |  |
|  | **HIV seropositivity**^c^ |  |  |  |  |
|  | HIV Seronegative | Ref |  |  |  |
|  | HIV Seropositive | 21.02 (7.08-62.41)* | <0.0001* |  |  |
| Vashishtha, 2013^b^ (Delhi)  *Outcome: Post-treatment mortality as a single outcome at 24 months after treatment initiation* [12] |  | Values below are odds ratios |  |  |  |
|  | **HIV**^d^ |  |  |  |  |
|  | HIV-negative | Ref |  |  |  |
|  | HIV-positive | 15.85 (1.95-129.09)* | 0.0098* |  |  |

AUDIT, Alcohol Use Disoder Identification Test; BMI, body mass index; SGRQ, Saint George’s Respiratory Questionnaire (a measure of respiratory symptom severity); TB, tuberculosis.

*Indicates statistical significance

^a^Variable was included in the analysis as a continuous variable, we have specified the unit of change in the variable associated with the effect estimate.

^b^Unadjusted odds ratios and/or p-values were estimated by the systematic review team from the raw data, as these were not provided in the original study.

^c^Estimated from findings in Table III of the study. Among patients with HIV, 49 had recurrence-free survival, 2 experienced recurrence, and 32 had post-treatment death. Among patients without HIV, 127 had recurrence-free survival, 7 experienced recurrence, and 4 had post-treatment death. Risk of recurrence was estimated excluding patients who died after treatment completion, while risk of death was estimated including those who experienced recurrence.

^d^Estimated by integrating findings from Figure 1, Table 5, and Table 8 of the study. We identified patients with and without HIV who achieved treatment success, excluding patients who experienced death, loss to follow-up, treatment failure, or modification of therapy. We then excluded those who did not complete the 24 months follow-up period or who were lost to follow-up post-treatment. Among those who completed follow-up, we estimated that, among patients with HIV, 41 had recurrence-free survival, 5 experienced relapse, and 9 died. Among patients without HIV, 79 had recurrence-free survival, 2 experienced relapse, and 1 died. Risk of recurrence was estimated excluding patients who died after treatment completion, while risk of death was estimated including those who experienced recurrence.

### References

1.	Subbaraman R, Nathavitharana RR, Satyanarayana S, Pai M, Thomas BE, Chadha VK, et al. The Tuberculosis Cascade of Care in India’s Public Sector: A Systematic Review and Meta-analysis. PLoS Med. 2016;13: e1002149. doi:10.1371/journal.pmed.1002149

2.	Cox SR, Gupte AN, Thomas B, Gaikwad S, Mave V, Padmapriyadarsini C, et al. Unhealthy alcohol use independently associated with unfavorable TB treatment outcomes among Indian men. Int J Tuberc Lung Dis. 2021;25: 182–190. doi:10.5588/ijtld.20.0778

3.	Dandekar R, Dixit J, Srinivasan D. The fate of tuberculosis cases after two years of DOTS chemotherapy in Aurangabad city, Maharashtra. National Journal of Community Medicine. 2014;5: 174–178.

4.	Gupte AN, Selvaraju S, Paradkar M, Danasekaran K, Shivakumar SVBY, Thiruvengadam K, et al. Respiratory health status is associated with treatment outcomes in pulmonary tuberculosis. Int J Tuberc Lung Dis. 2019;23: 450–457. doi:10.5588/ijtld.18.0551

5.	Huddart S, Singh M, Jha N, Benedetti A, Pai M. Case fatality and recurrent tuberculosis among patients managed in the private sector: A cohort study in Patna, India. PLOS ONE. 2021;16: e0249225. doi:10.1371/journal.pone.0249225

6.	Lisha PV, James PT, Ravindran C. Morbidity and mortality at five years after initiating Category I treatment among patients with new sputum smear positive pulmonary tuberculosis. Indian J Tuberc. 2012;59: 83–91.

7.	Mahishale V, Patil B, Lolly M, Eti A, Khan S. Prevalence of Smoking and Its Impact on Treatment Outcomes in Newly Diagnosed Pulmonary Tuberculosis Patients: A Hospital-Based Prospective Study. Chonnam Med J. 2015;51: 86–90. doi:10.4068/cmj.2015.51.2.86

8.	Mave V, Gaikwad S, Barthwal M, Chandanwale A, Lokhande R, Kadam D, et al. Diabetes Mellitus and Tuberculosis Treatment Outcomes in Pune, India. Open Forum Infectious Diseases. 2021;8. doi:10.1093/ofid/ofab097

9.	Ramachandran G, Chandrasekaran P, Gaikwad S, Agibothu Kupparam HK, Thiruvengadam K, Gupte N, et al. Subtherapeutic Rifampicin Concentration Is Associated With Unfavorable Tuberculosis Treatment Outcomes. Clin Infect Dis. 2020;70: 1463–1470. doi:10.1093/cid/ciz380

10.	Thomas A, Gopi PG, Santha T, Chandrasekaran V, Subramani R, Selvakumar N, et al. Predictors of relapse among pulmonary tuberculosis patients treated in a DOTS programme in South India. Int J Tuberc Lung Dis. 2005;9: 556–561.

11.	Tripathy S, Anand A, Inamdar V, Manoj MM, Khillare KM, Datye AS, et al. Clinical response of newly diagnosed HIV seropositive & seronegative pulmonary tuberculosis patients with the RNTCP Short Course regimen in Pune, India. Indian J Med Res. 2011;133: 521–528.

12.	Vashishtha R, Mohan K, Singh B, Devarapu SK, Sreenivas V, Ranjan S, et al. Efficacy and safety of thrice weekly DOTS in tuberculosis patients with and without HIV co-infection: an observational study. BMC Infect Dis. 2013;13: 468. doi:10.1186/1471-2334-13-468

13.	Velayutham B, Chadha VK, Singla N, Narang P, Gangadhar Rao V, Nair S, et al. Recurrence of tuberculosis among newly diagnosed sputum positive pulmonary tuberculosis patients treated under the Revised National Tuberculosis Control Programme, India: A multi-centric prospective study. PLoS One. 2018;13: e0200150. doi:10.1371/journal.pone.0200150

14.	Kolappan C, Subramani R, Karunakaran K, Narayanan PR. Mortality of tuberculosis patients in Chennai, India. Bull World Health Organ. 2006;84: 555–560. doi:10.2471/blt.05.022087

15.	Kolappan C, Subramani R, Kumaraswami V, Santha T, Narayanan PR. Excess mortality and risk factors for mortality among a cohort of TB patients from rural south India. Int J Tuberc Lung Dis. 2008;12: 81–86.

16.	Sadacharam K, Gopi PG, Chandrasekaran V, Eusuff SI, Subramani R, Santha T, et al. Status of smear-positive TB patients at 2-3 years after initiation of treatment under a DOTS programme. Indian J Tuberc. 2007;54: 199–203.

17.	Sharma R, Prajapati S, Patel P, Patel B, Gajjar S, Bapat N. An Outcome-Based Follow-up Study of Cured Category I Pulmonary Tuberculosis Adult Cases from Various Tuberculosis Units under Revised National Tuberculosis Control Program from a Western Indian City. Indian J Community Med. 2019;44: 48–52. doi:10.4103/ijcm.IJCM_310_18
